# Personalized Tuberculosis Treatment Recommendation System (PTTRS) version 1: A Precision-Medicine Based Application for Recommending Personalized Treatment to Tuberculosis Patients

**DOI:** 10.1101/2025.06.23.25330035

**Authors:** Ananya Anurag Anand, Rajat Kumar Mondal, Baishali Sarkar, Sintu Kumar Samanta

## Abstract

Tuberculosis is one of the leading causes of death in underdeveloped and developing countries. The limited access to detailed drug susceptibility testing, lack of knowledge of second-line anti-TB drugs in underdeveloped nations, insufficient adherence to drug dosages, and comorbidities challenge the management of drug-resistant TB. Further, the number of deaths due to TB is increasing with time due to reasons such as high levels of resistance mutation in TB strains, lack of on-time delivery of treatment, and lack of personalized medicine. Thus, we have developed a web-based application for aiding in the personalized treatment of TB patients based on their medical condition. The application takes geographical location, age, sex, medical and travel history, AMR report and associated medical conditions (AMCs) of the patient as input, and thereafter, outputs the list of drugs which are safe for the patient. The application also helps in knowing the possible side-effects of various drug combinations administered for TB. Additionally, the application also outputs the side-effects of combination of drugs for TB and for any AMC that the patient is suffering with. PTTRS can be accessed at https://pttrs-bblserver.streamlit.app/. Our polypharmacy side effect predictor can be used for any other disease as well. It can be accessed at https://psep-bblserver.streamlit.app/.

## **1.** Introduction

Tuberculosis (TB) is a contagious air-borne lung disease caused by the bacteria *Mycobacterium tuberculosis*, which shows varied drug resistance in different patients (**Tobin and Tristram, 2024**). It spreads via airborne respiratory droplets (coughs or sneezes) and saliva (kissing or shared drinks), making it a highly transmissible disease. TB is generally characterized by a persistent cough that lasts more than three weeks and usually brings up phlegm, which may be bloody **(Guglielmetti et al., 2025)**. It may also present other clinical manifestations, such as weight loss, night sweats, high fever, tiredness, fatigue, loss of appetite, and swellings that persist even after a few weeks. It may disseminate to organs other than lungs in later stages of the disease.

In 2019, 10 million people were estimated to have developed active tuberculosis (TB), out of which 1·2 million people died **(Chakaya et al., 2021)**. In 2019, 3% of the fresh TB cases worldwide were estimated to be multidrug-resistant (MDR). Additionally, 18% of individuals who had been previously treated had MDR TB. Thus, TB stands tall as one of the world’s most fatal infections affecting millions.

Until the breakout of the COVID-19 pandemic, TB was the first such disease which could cause death due to a single infectious agent (https://www.who.int/publications/i/item/9789240093461). Launched in 2015, the WHO End TB Strategy offers a comprehensive framework for lowering the incidence, mortality rate, and economic impact of TB, particularly DR-TB, by 2030 (https://www.who.int/publications/i/item/WHO-HTM-TB-2015.19). However, progress has been slower than anticipated, and by 2021, the 2020 targets had not yet been met (https://www.who.int/teams/global-tuberculosis-programme/tb-reports/global-tuberculosis-report-2022). This is partially explained by the COVID-19 epidemic, which has severely impacted the availability of TB detection, treatment, and care services in numerous nations. According to estimates, the COVID-19 pandemic reversed years of decline in the incidence and mortality of TB, particularly RR-TB illness, globally for the first time in decades **(Agins et al., 2019; Dheda et al., 2022).** As a result, MDR *Mycobacterium tuberculosis* has been included in the 2024 BPPL list (bacterial priority pathogen list) of WHO.

MDR-TB (Multidrug-resistant Tuberculosis) occurs when a patient becomes resistant to both rifampin and isoniazid, the first-line anti-TB drugs **(Khan and Das, 2020)**. This severe problem is quite common in TB patients. As a result, various drug combinations are suggested by doctors to the patients (Polypharmacy) **(Guglielmetti et al., 2025)**. While in most cases the drugs remain ineffective due to the mutations occurring in the TB strains that make them resistant to these drugs, another reason for TB treatment failure is the side-effects of polypharmacy depending on the severity of the case and the background of the patient, which includes age, sex, medical history, family history, travel history and most importantly any other medical condition the patient is associated with **(Chakaya et al., 2021; Correia et al., 2022)**.

The combination of multiple medications can lead to drug-drug interactions, wherein the activity of one medication is altered, either positively or negatively, when combined with another (**Dehghan et al., 2021; Masumshah et al., 2021).** Identifying and understanding these interactions is crucial for ensuring patient safety and optimal treatment outcomes **(Zitnik et al., 2018).** Therefore, it becomes important for us to overcome this problem to reduce the severity of the morbidity and mortality rates involved in TB. One of the approaches by which we can cure TB faster in affected patients is by giving personalized medicine.

The current treatment for the constantly evolving MDR-TB bacteria includes antibiotics such as rifampin and isoniazid, which can be supplemented with one or more drugs, most preferably ciprofloxacin and amikacin (**Ananthanarayan, R., & Jayaram Paniker, C. K., 2010**). The treatment of tuberculosis patients has more or less followed a one-dimensional approach during the last 50 years (**Lange et al., 2020**). This standardization ignores individual differences in susceptibility to infection. The efficacy of the treatment is lowered in cases where the patient is suffering from other medical conditions as well, such as AIDS, thyroid, and rheumatoid arthritis among others. TB is responsible for causing approximately 40% of deaths related to HIV and AIDS (**Lelisho et al., 2022; Gupta et al., 2015)**. People with HIV are at greater risk of acquiring MDR tuberculosis than people who are HIV-negative (**Müller et al., 2018; WHO, 2020**). Also, treatment outcomes in people with HIV and MDR tuberculosis are worse than among HIV-negative patients with MDR tuberculosis. Another example of an associated medical condition is diabetes **(Krishna and Jacob, 2000; Tornheim and Dooley, 2017)**. A patient left with untreated latent TB who also has diabetes is more prone to developing TB disease than one without diabetes (https://www.cdc.gov/tb/topic/basics/tb-and-diabetes.html). Shockingly, there exists 2-4-fold higher risk of active TB in patients of diabetes mellitus. Additionally, according to a previous report, nearly 30% of TB patients are likely to develop diabetes. This shows that there is a need to consider the associated medical conditions (AMCs) as well. For a world that is so diverse, the diversity of patients comes in automatically, and, this is due to the genetic differences between individuals, and the environment in which they live (**Khan and Das, 2020**). Thus, the need for personalized medicine rises up **(Verboven et al., 2022).**

However, due to lack of such personalized strategies, effective treatment is delivered to the patient at quite a later stage of the disease, consequently leading to the death of the patient.

To solve the above problem, we have designed an application that will help doctors suggest treatment to tuberculosis patients in a personalized manner. The application requires the user to input details like patient age, gender, geographical location, travel history, number of tuberculosis cases in family or neighborhood, medical history, associated medical conditions (AMCs), and current medication being taken. Patient information will then be collected and saved. However, all details remain confidential. Here, we have considered five important AMCs with respect to TB, which include diabetes, thyroid, asthama, HIV/AIDS and rhematoid arthritis (**Correia et al., 2022**). Out of these, diabetes and HIV have been regarded as comorbidities, in case of TB, by WHO (https://www.who.int/southeastasia/activities/co-morbidities-tb). Based on what the user (most probably a clinician or a researcher) inputs regarding the patient, the application will first identify the probable strains the patient could be infected with, depending on one’s current location and travel history. Anti-microbial resistance (AMR) report, if available, will be taken into consideration. The drugs to which the patient is already resistant will be eliminated. Thereafter, based on the AMCs and the drugs being consumed for the AMCs, the application will be able to suggest the combinations of drugs suitable for intake and their side effects. The application will also help in extending knowledge about the incompatibility of certain drugs in patients with particular medical conditions. For example, Isoniazid and pyrazinamide must be avoided as treatment options in TB patients suffering from rheumatoid arthritis. Another helpful feature of this web-based application is that the application allows the doctor or the clinician to register on it and log in with the same details every time. This is done so that the clinician or physician can save the patient’s information at the time of consultation and can refer to it the next time the patient comes. This will aid in keeping track of the patient’s record and the progress of his treatment. Thus, in all aspects, this application is an effort to help in the timely recovery of TB patients of varied types.

Further, although few models to predict drug-drug side effects, like NNPS and decagon model, exist, these lack the ability to uptake user input and lack any graphical user interface (GUI) **(Masumshah et al., 2021; Zitnik et al., 2018)**. Additionally, our model is specialized for predicting drug drug side-effects especially in case of TB patients by considering the overall status and well-being of the patient. We have, thus, created a Personalized Tuberculosis Treatment Recommendation System (PTTRS) comprising: i) a data collection and integration component configured to aggregate patient data, drug information, and treatment recommendations from diverse sources; ii) machine learning models employing algorithms such as XGBoost and Isolation Forest to analyze patient data and generate personalized treatment recommendations; iii) a graphical user interface (GUI) developed using front-end technologies for inputting patient data, viewing treatment recommendations, and managing patient health information; iv) a web-based application enabling remote access to the PTTRS from any device with an internet connection; v) a database management system utilizing relational database management systems (RDBMS) databases to securely store and manage patient data, drug information, and treatment recommendations; vi) collaboration mechanisms with the health science industry to access pharmacogenomics data and other pertinent information essential for TB treatment; vii) security measures including encryption, access controls, and compliance measures to safeguard patient data and ensure system integrity.

The system allow the individuals to create unique usernames and passwords to access the PTTRS, ensuring multi-user management across different geographical locations. Further, it allows the user to beautifully visualize mono drug and drug-drug side effects as well, allowing users to select specific TB drugs and visualize their side effects individually or in combination. Finally, the system comprises a patient database for doctors to monitor patient-related data, track treatment outcomes, and make adjustments to treatment plans as necessary.

## **2.** Materials and method

### 2.1. Overall Strategy

We created a very simple strategy before executing the work. First, about the data, we collected data about the TB strains (worldwide) and locations (worldwide), TB drugs, AMCs that were well observed with TB patients and its drugs, drugs that can’t be given in a particular AMC if the patient has TB, mono drug and polydrug side effects. We only use the poly-drug side effect data to build ML models. We used 2 ML models, a generalized which can predict any side effect of a drug combination and a specific one that can predict only one side effect of a drug combination. The consensus of these two predictions will be displayed to the user. Based on the models a web application was developed to take the canonical SMILES/InChI of 2 drugs & predict the side effects. Moreover, another web application was developed where we incorporated all the information for interaction and the same model for the prediction of TB/AMC polypharmacy side effects and a database to maintain TB patient records for doctors. We hope the overall description helps to understand the whole methodology briefly.

### **2.1.** Data acquisition

#### 2.1.1. Predominant TB strains (worldwide) and locations (worldwide)

All the information about predominant TB strains (worldwide) which are found in different locations (worldwide) was curated from literature and websites. The curated data (with data source) is available in the supplementary file S1.

#### **2.1.1.** TB drugs and AMC drugs

The TB and AMC drugs including their name, SMILES, and InChI were collected from DrugBank (https://go.drugbank.com/), PubChem (https://pubchem.ncbi.nlm.nih.gov/), and ChEMBL (https://www.ebi.ac.uk/chembl/) databases. Only name was collected if the drug didn’t contain a discrete structure (i.e., SMILES or InChI) on PubChem (https://pubchem.ncbi.nlm.nih.gov/) or ChEMBL (https://www.ebi.ac.uk/chembl/) or DrugBank (https://go.drugbank.com/). In Table 1 the TB drug and AMC drug statistics are shown.

**Table 1.**
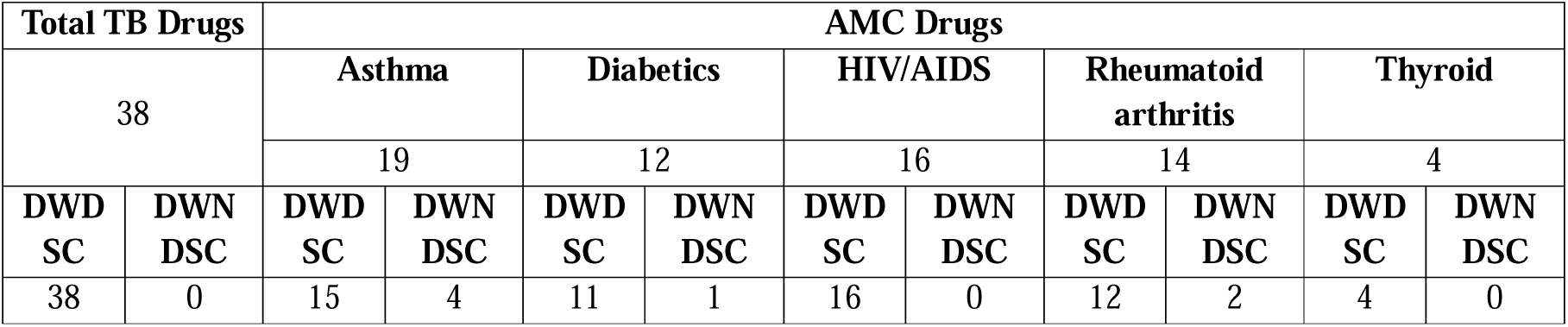
Table of TB drugs and AMC drugs that are currently enlisted within PTTRS.

The table contains the counts of all TB and AMC drugs that are provided within PTTRS. As we mentioned earlier in the case of a few drugs SMILES or InChI was not found in PubChem, ChEMBL, and DrugBank, hence in the table we also show the drugs with district structure count (DWDSC) and drugs with no district structure count (DWNDSC).

#### **2.1.2.** Mono-drug side effects

The mono-drug side effects are collected from the SIDER database (**Kuhn et al., 2016**). For a few drugs, the mono drug side effects information was not available in the SIDER database (http://sideeffects.embl.de/), in that case, we used PubMed literature (https://pubmed.ncbi.nlm.nih.gov/), MedlinePlus (https://medlineplus.gov/), Mayo Clinic (https://www.mayoclinic.org/), Drugs.com (https://www.drugs.com/), etc. to collect the mono-drug side effect information.

#### **2.1.3.** Poly-drug side effects

The poly-drug side effects data was obtained from the TWOSIDES database (**Martinez et al., 1970**). The dataset contains 645 unique drugs (in terms of PubChem compound/substance IDs, which were used later to collect their respective SMILES by using PubChem Identifier Exchange Service (https://pubchem.ncbi.nlm.nih.gov/docs/identifier-exchange-service)), 63,473 unique drug pairs, and 1,317 unique side effects. The dataset contains a total of 4,649,441 (4.64 M) drug pairs because one drug combination can show at least 1 side effect(s). This dataset was considered as a raw dataset for further processing.

#### **2.2.** Choice of algorithm

As we discussed earlier 2 types of models i.e., one generalized and one specialized. For individual model training 2 different types of algorithms were chosen.

#### **2.2.1.** For generalized model

To make a generalized model XGBoost algorithm (**Chen, T. & Guestrin, C., 2016**) was chosen for several reasons. First and foremost, its exceptional performance and speed, bolstered by optimization and parallelization, set it apart from traditional gradient-boosting implementations. Moreover, XGBoost offers built-in regularization techniques like L1 and L2 regularization, crucial for preventing overfitting and enhancing generalization. Its flexibility shines through in supporting various objective functions and evaluation criteria, making it adaptable to diverse data types and tasks. Additionally, XGBoost employs tree pruning to simplify models, handles missing values seamlessly, and provides insights into feature importance for enhanced interpretability. Its effectiveness in handling unbalanced datasets, thanks to weighted classes, robustness to noise, specialized objective functions, evaluation metrics, and ensemble learning, further cements its superiority.

#### **2.2.2.** For specific models

For making specific models we used single class classification or anomaly detection techniques. Here we considered Isolation Forest (**Liu et al., 2008**) (a bagging technique of the ensemble learning method) for several reasons. Isolation Forest is well-suited for single-class classification due to its efficiency in isolating anomalies, which aligns with the task of identifying instances belonging to a particular class. Its scalability enables the handling of large datasets with numerous features, while its robustness to irrelevant features ensures reliable performance. The interpretability of anomaly scores aids in understanding the degree of "belongingness" to the positive class, making it valuable for classification tasks. With readily available implementations in popular machine learning libraries, Isolation Forest stands out as a practical choice for single-class classification **(Li et al., 2021).**

### **2.3.** Raw data set optimization

#### 2.3.1. For generalized model training

As we mentioned earlier, we have obtained the poly-drug side effects dataset from the TWOSIDES database (**Martinez et al., 1970**), consisting of 4.64 M data points (i.e., drug pairs) and 1,317 classes (i.e., side effects). The dataset is utilized for both the NNPS and Decagon methods. Within this dataset, class distribution varies significantly, with the minimum number of data points in a class being 1 (Class 1099) and the maximum being 28,568 (Class 119). Our computational resources impose constraints, limiting the maximum number of data points usable, at a particular time quantum, for training to 1.54 M. To optimize the dataset, initially, we considered randomly selecting one-third of all data points. However, this approach does not ensure consideration for all classes. To address this, we devised a strategy wherein 1,176 data points are randomly selected from each class, totaling 1.54 million data points overall. Nevertheless, challenges arise when a class has fewer than 1,176 data points. To mitigate this, we established conditional rules. Classes with 1,176 or more data points have their samples randomly selected based on their index and then sorted by the index in ascending order. Classes with between 588 and 1,176 data points are retained as is, while classes with fewer than 588 data points have their data points duplicated until reaching 588. This conditional approach ensures a degree of balance in the dataset while minimizing the introduction of biases from excessive duplication. Consequently, the final raw dataset is not perfectly balanced but maintains a reasonable degree of balance and minimizes duplicate instances.

#### 4.2.2. For specialized model training

In the case of specialized model training, classes with 1,176 or more data points are retained as is, while classes with fewer than 588 data points have their data points duplicated until reaching 588. Since one specialized model was trained at a time, the data points and resource usage were at the limit. By training the specialized models we are able to train specific models one by one based on all the data points of a respective class (e.g., model 0 is trained based on all the data points of class 0, model 1 is trained based on all the data points of class 1, and so on).

### 4.4. Feature Engineering

#### 4.4.1. For generalized model training

Physicochemical properties and the side effect matrix was combinedly used as features to train the generalized model. However, before training the model, the features were modified logically. First, we take a drug pair that contains 2 drugs i.e., drug 1 and drug 2, now by using the RDKit python package (version 2023.09.05) (https://www.rdkit.org/docs/GettingStartedInPython.html) we computed all the physicochemical properties (there were 200 physicochemical properties which can be computed by RDKit). RDKit returns all the physicochemical properties in terms of some float value (SFV) for a compound, which can be either negative or positive. After computing those 200 physicochemical properties for the individual drug in a drug pair, we took their absolute of all physicochemical properties values, so now we have the positive value only. Then, the values (i.e., 200 values) of respected physicochemical properties of Drug 1 were subtracted from the values (i.e., 200 values) of respected physicochemical properties of Drug 2. After subtracting the value again we take the absolute value. Below the strategy is represented as a formula.

ppd_x_ = absolute (absolute (pp_d1i_) – absolute (pp_d2j_)) … [formula 1]

Where ppd = minimum physicochemical properties differences between 2 drugs; x = 1 to 200 absolute physicochemical property difference; i = 1 to 200 physicochemical properties; d1 = drug1; j = 1 to 200 physicochemical properties; d2 = drug2.

This strategy was specifically used because after training the model when it will be public, we don’t know in which particular manner the user gives input (as SMILES or InChI) to that model via user interface (UI). Suppose during model training there were d1 and d2, but the user gives input d2 as first and d1 as second because the user should not bother about the drug sequence during model training. So, if we train the model by considering the minimum absolute physicochemical properties difference feature, then the model can predict the side effects for any kind of drug combination in this world, only the condition is both drugs of the drug combination must have a valid SMILES or InChI.

After this, the side effect matrix (SEM) was incorporated with the physicochemical properties difference values The SEM was built by using the dummy encoding scheme (https://www.analyticsvidhya.com/blog/2020/08/types-of-categorical-data-encoding/) technique to avoid the dummy variable trap (https://medium.com/data-science-365/what-is-the-dummy-variable-trap-and-how-to-avoid-it-aeb227c2cd92) issue during model training. Hence the SEM contains 1316 columns instead of 1317 columns (a total of 1317 side effects were there in the dataset). The SEM was incorporated because a single drug pair can show different side effects, so if we train the generalized model with the only minimum absolute physicochemical properties difference then the model will be confused. Let us explain a little bit. Suppose, drug 1 and drug 2 show side effect x, side effect y, and side effect z. So, without SEM there is no uniqueness to the feature. By adding the SEM with the minimum absolute physicochemical property difference of a drug combination, we are able to maintain the feature uniqueness. In Table 2, a glimpse of the feature engineering of the dataset for generalized model training is shown.

**Table 2.** Representation of features that were used to train the generalized model and specialized models.

|  |  |  | Dataset for generalized model training |  |  |  |  |  |  |  |  |
| --- | --- | --- | --- | --- | --- | --- | --- | --- | --- | --- | --- |
| Drug pair |  | Side effect | y | x1 | x2 | ... |  |  |  |  | x1516 |
|  |  |  | Label<br>d SE | d1_d2_abs<br>_diff_p1 | d1_d2_abs<br>_diff_p2 | ... | d1_d2_<br>abs_dif<br>f_p200 | SE1 | SE2 | ... | SE1316 |
| Drug 1 | Drug 2 | SE 1 | 0 | SFV | SFV | ... | SFV | 1 | 0 | ... | 0 |
| Drug 1 | Drug 3 | SE 3 | 2 | SFV | SFV | ... | SFV | 0 | 0 | ... | 0 |
| Drug 5 | Drug 6 | SE 10 | 9 | SFV | SFV | ... | SFV | 0 | 0 | ... | 0 |
| ⋮ | ⋮ | ⋮ | ⋮ | ⋮ | ⋮ | ⋮ | ⋮ | ⋮ | ⋮ | ⋮ | ⋮ |
| Drug 643 | Drug 644 | SE 1316 | 1315 | SFV | SFV | ... | SFV | 0 | 0 | ... | 1 |
| Drug 643 | Drug 645 | SE 1317 | 1316 | SFV | SFV | ... | SFV | 0 | 0 | ... | 0 |
| Drug 644 | Drug 645 | SE 2 | 1 | SFV | SFV | ... | SFV | 0 | 1 | ... | 0 |
|  |  |  | Dataset for specialized models training |  |  |  |  |  |  |  |  |

Upon the completion of feature engineering the features i.e., independent variables were stored in a sparse matrix (from scipy python package (version 1.12.0) (https://scipy.org/)) and the dependent variable was stored in a numpy array (from numpy python package (version 1.26.0) (https://numpy.org/)). Those data structures were exclusively used to train the model faster.

#### 4.4.2. For specialized models training

To train the specialized models (i.e., individual models for individual classes), we only consider the minimum absolute physicochemical properties difference values. Since individual models for individual classes, there is no support required with the minimum absolute physicochemical properties difference values. In Table 2, a glimpse of the feature engineering of the dataset for specialized model training is shown. After completion of the feature engineering, individual class-wise independent variables were stored in a sparse matrix and the dependent variable was stored in a numpy array.

The table contain a visual representation of the features that were used to train the model. In the table it is observed that what are the independent variables (x1…x200 for specialized models and x1…x1316 for generalized model) and dependent variable (y). d1_d2_abs_diff_p1 represents the minimum absolute physicochemical properties difference value of physicochemical property 1 for 2 drugs in a drug pair, d1_d2_abs_diff_p2 represents the minimum absolute physicochemical properties difference value of physicochemical property 2 for 2 drugs in a drug pair, and so on. SFV represents some floating value of minimum absolute physicochemical properties difference. SE1 represents side effect 1, SE2 represents side effect 2, and so on.

### 4.5. Dataset split

The dataset was split into 3 parts (for the generalized model and specialized models) i.e., the training set (80%), test set (10%), and validation set (10%). The sklearn python package (version 1.4) (https://scikit-learn.org/stable/) was used for this purpose.

### 4.6. System architecture and model training

We used the high-performance computing (HPC) facility of the central computing facility (CCF) of our institute (https://ccf.iiita.ac.in/). Only one node from core160 (https://ccf.iiita.ac.in/hpc.html) was used with 100% CPU utilization. The node architecture is shown in Table 3.

**Table 3.**
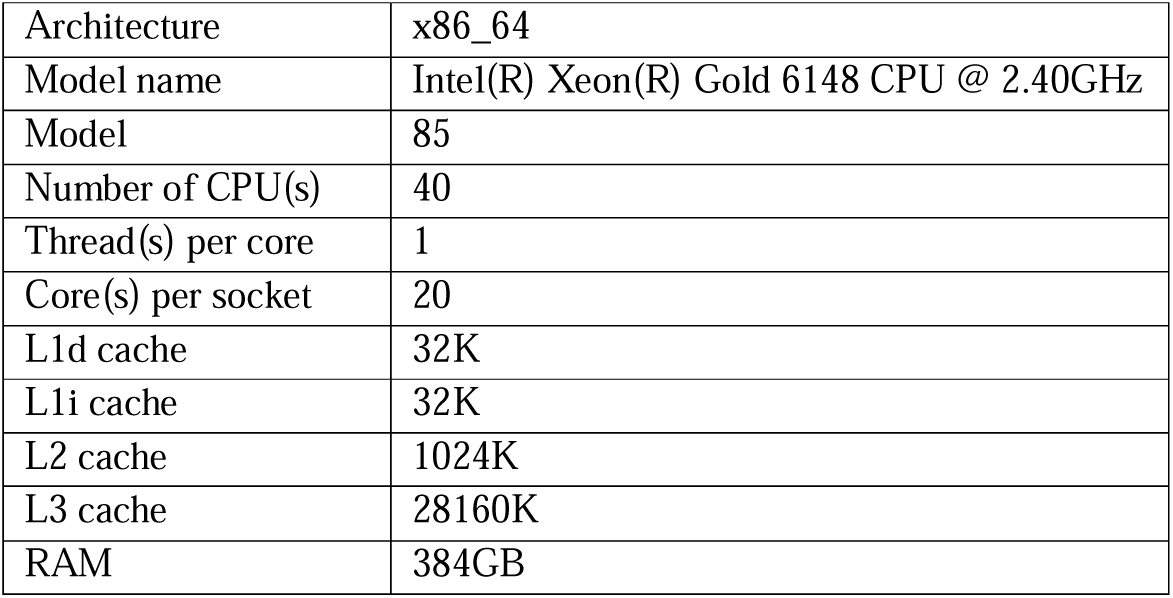
The table shows a node architecture of core160 of HPC of CCF of our institute.

|  |  |
| --- | --- |
| Architecture | x86_64 |
| Model name | Intel(R) Xeon(R) Gold 6148 CPU @ 2.40GHz |
| Model | 85 |
| Number of CPU(s) | 40 |
| Thread(s) per core | 1 |
| Core(s) per socket | 20 |
| L1d cache | 32K |
| L1i cache | 32K |
| L2 cache | 1024K |
| L3 cache | 28160K |
| RAM | 384GB |

The generalized model was trained based on the XGBoost algorithm (implemented in xgboost python package (version 2.0.3) (https://xgboost.readthedocs.io/en/stable/python/python_intro.html)) with 100% CPU utilization and took 1235.09 seconds (20.5848 minutes). The specialized model training information is discussed in the letter section.

### 4.7. Model validation

#### 4.7.1. Generalized model

For model validation we considered the parameters such as training score, testing score, validation score, accuracy, F1 score, precision, recall, Matthews correlation coefficient (MCC), true positive (TP), false positive (FP), false negative (FN), true negative (TN), sensitivity, and specificity, area under the receiver operating characteristic curve (AUROC) (micro average), AUROC (macro average), area under the precision-recall curve (AUPRC) (micro average), AUROC (macro average). In Table 4 all the values are shown.

**Table 4.** Validation parameters of the generalized model.

| Training score | Test score | Validation score | Accuracy | F1 Score | Precision | Recall | MC C | TP | FP | FN | TN | Sensitivity | Specificity | AUROC (micro average) | AUROC (macro average) | AUPRC (micro average) | AUPRC (macro average) |
| --- | --- | --- | --- | --- | --- | --- | --- | --- | --- | --- | --- | --- | --- | --- | --- | --- | --- |
| 1.00 | 1.00 | 1.00 | 1.00 | 1.00 | 0.99 | 1.00 | 1.00 | 112154 | 29 | 29 | 146062237 | 0.99 | 0.99 | 1.00 | 0.99 | 1.00 | 0.98 |

We got the above values of validation parameters without any hyperparameter tuning. Hence, further hyperparameter tuning is not done. The trend of AUROC and AUPRC of Table 4 is shown in Figure 1. After that, we did 10-fold cross-validation by using GridSearchCV. The result of the cross-validation is shown in Table 5.

**Figure 1.**
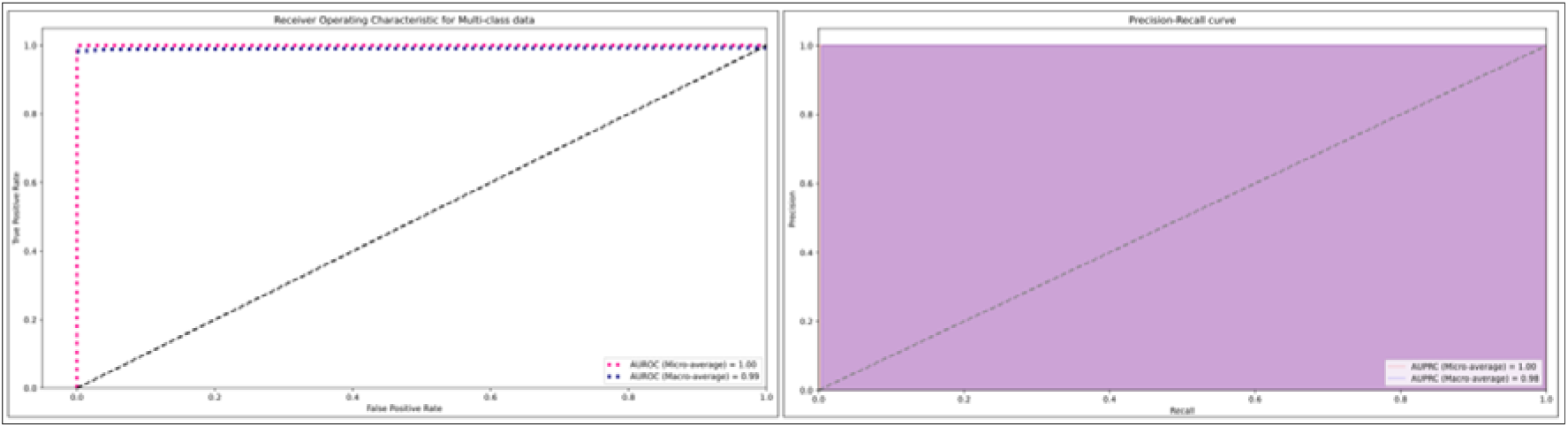
Trend of AUROC (micro & macro) and AUPRC (micro & macro).

**Table 5.**
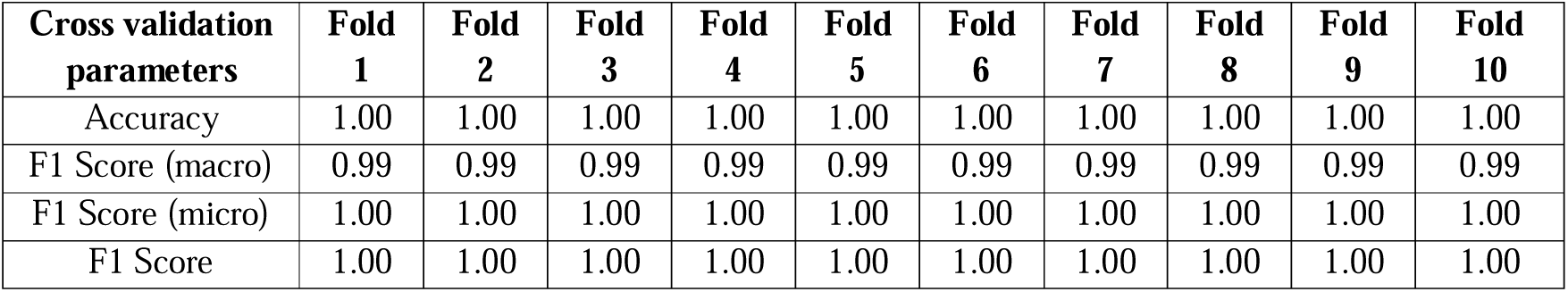
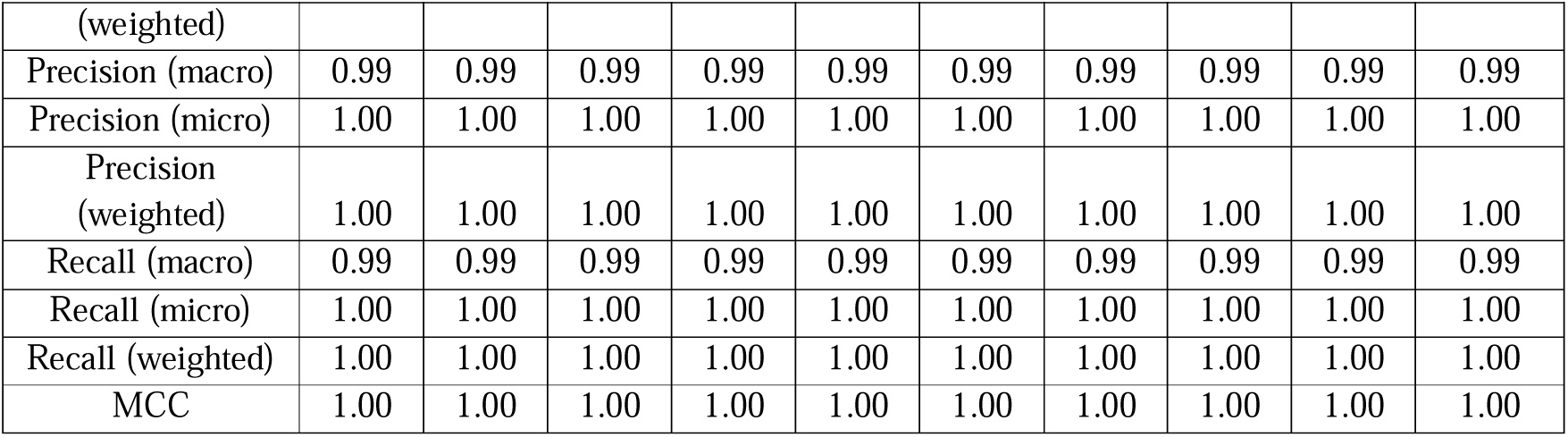
10-fold cross validation of result of the generalized model.

From Table 4, Table 5, Figure 1, it can be easily interpreted that the generalized model is highly robust to determine the 1317 side effects of any drug combinations accurately. In Table 5 we show all the parameters, which are applicable and can be calculated by GridSearchCV (https://scikit-learn.org/stable/modules/generated/sklearn.model_selection.GridSearchCV.html) for multiclass classification. By using the default parameters of the XGBoost algorithm and our feature engineering, we are able to achieve a good accuracy score as well as other validation parameters, hence further hyperparameter tuning is not done.

#### 4.7.2. Specialized models

As we mentioned earlier, we used the Isolation Forest algorithm (implemented in sklearn python package (version 1.4) (https://scikit-learn.org/stable/)) for train the specialized models one by one. For validation of each model, we considered those parameters that are applicable for single class classification such as accuracy, validation score, TP, FP, TN, FN, recall, precision, F1 score, mean square error (MSE), AUROC, and AUPRC. All the validation parameters along with a few important terms (like training time) of the first 20 individual models is shown in Table 6 and the information for all models is shown in supplementary file S2. The average value of all these parameters of all models is shown in Table 7.

**Table 6.**
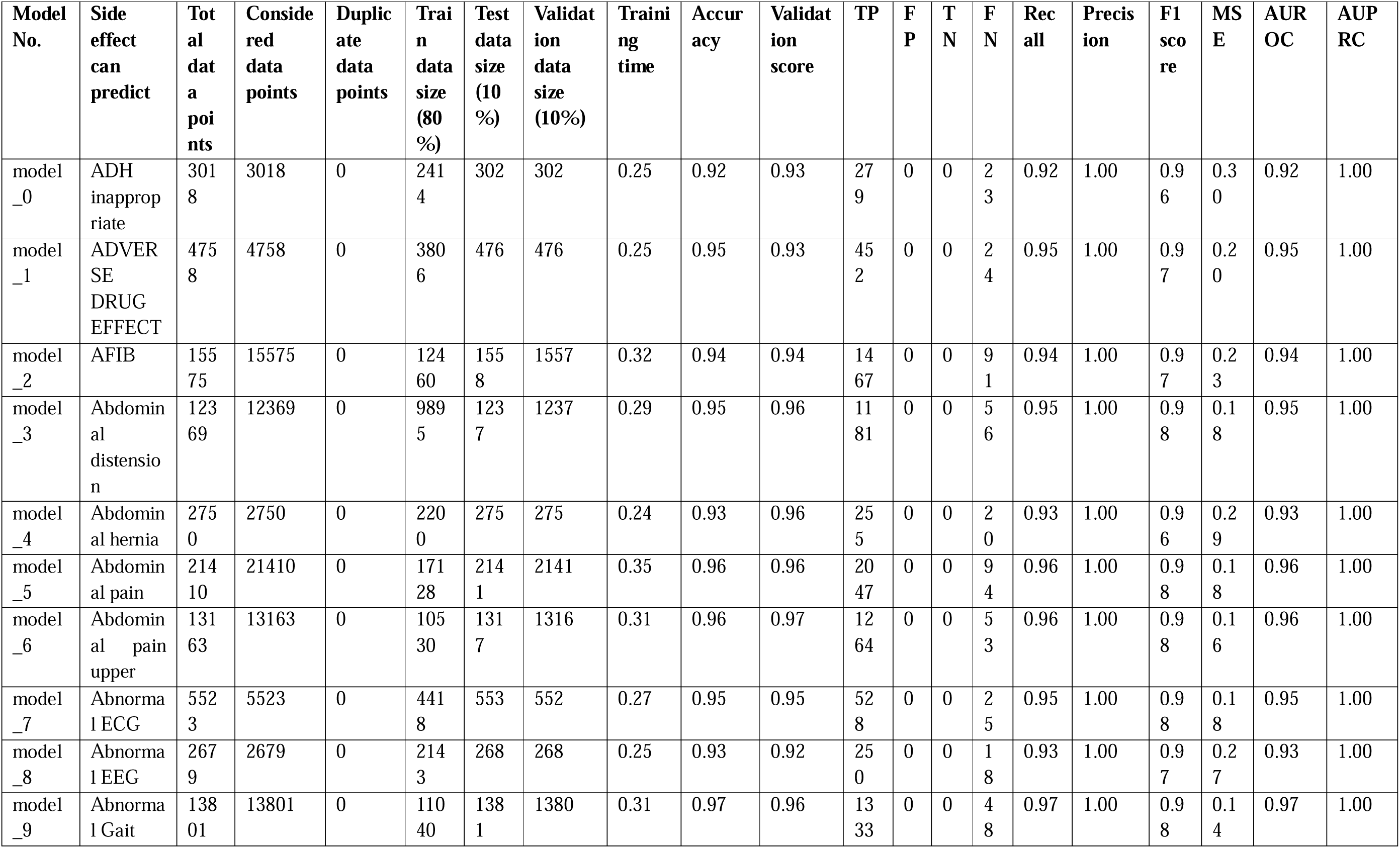
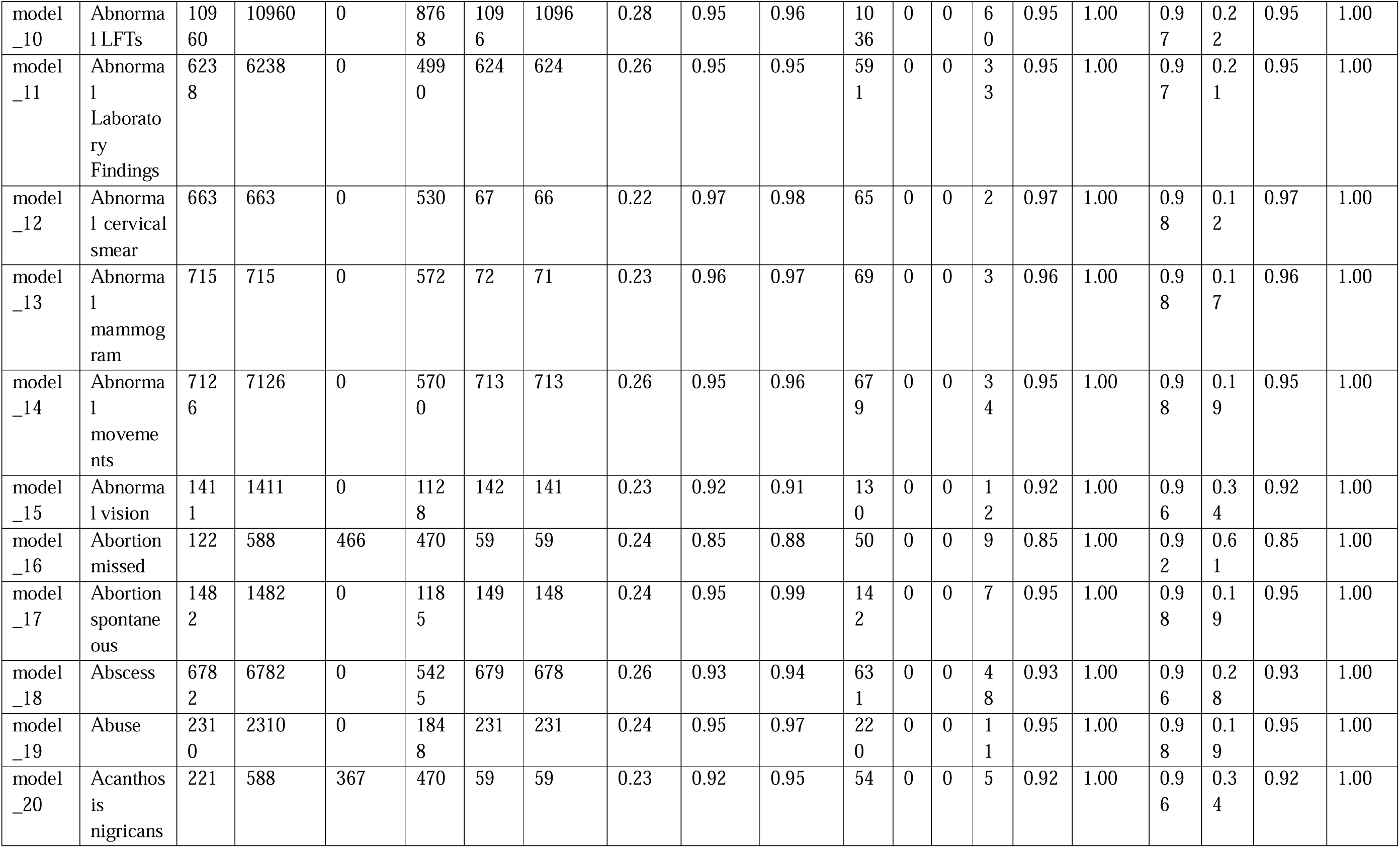
Table of all parameters of first 20 individual models.

**Table 7.**
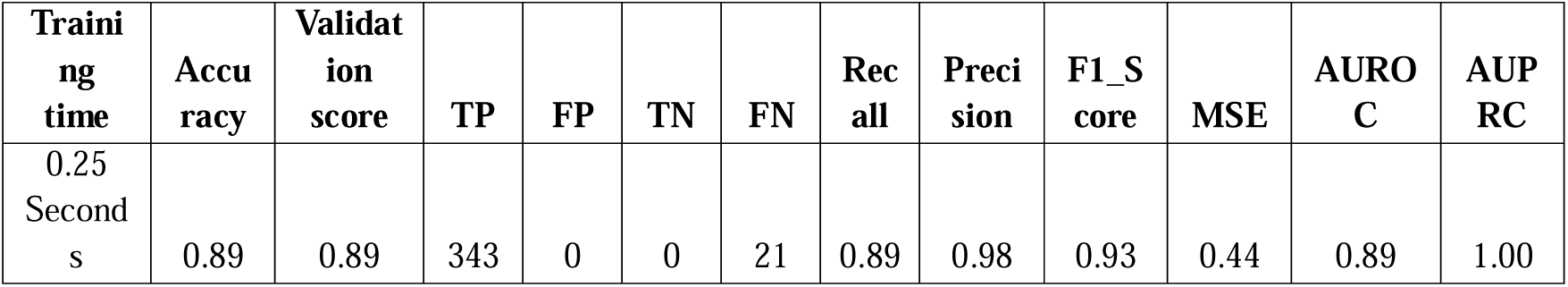
Average values of validation parameters of 1317 models.

| Training time | Accuracy | Validation score | TP | FP | TN | FN | Recall | Precision | F1_Score | MSE | AUROC | AUPRC |
| --- | --- | --- | --- | --- | --- | --- | --- | --- | --- | --- | --- | --- |
| 0.25 Seconds | 0.89 | 0.89 | 343 | 0 | 0 | 21 | 0.89 | 0.98 | 0.93 | 0.44 | 0.89 | 1.00 |

In the above table there are 3 terms i.e., ‘Total data points’, ‘Considered data points’, and ‘Duplicate data points’ present. Let us discuss one by one. ‘Total data points’ indicate all the data points that were present in the dataset; ‘Considered data points’ are those data points that were considered to train the model. ‘Duplicate data points’ indicates how many data points are duplicated out of ‘Considered data points’. ‘Duplicates data points’ are only found if ‘Total data points’ are less than 588. For example, in the case of model 20 ‘Total data points’ is 221 which is less than 588 so to increase the data set up to 588, 367 data points are duplicated from 221 data points which is shown in ‘Duplicate data points’ column. This strategy was followed because few classes contain very few data points. Though here we train the model based on a single class classification algorithm, based on a very low amount of data points it becomes very difficult to split and train the model. Hence, that strategy was followed.

### 4.8. Store the models

After training the generalized model and specialized models, all the models were stored in binary joblib file format by using joblib python package (version 1.4) (https://joblib.readthedocs.io/en/stable/). Those joblib files were used inside the application for prediction purpose.

### 4.9. Application development

We developed 2 apps, one can only predict the 1317 side effects for any drug combination (contains very simple UI, no login required) named ‘polypharmacy side effect predictor’ and one for TB specific (contain interactive UI, TB drug recommender, patient data management, login required) named ‘PTTRS’.

#### 4.9.1. Backend development

All the information including TB strain and location details, TB drugs details, AMC and it’s associated drug details, well-known cross-reactivity information between tb and AMC drugs were stored inside the MySQL server (Relational Database Management System (RDBMS)) (version 5.7.36) of phpMyAdmin (version 5.1.1) with HTTPS server. All the models were stored in a GitHub repository.

#### 4.9.2. Frontend development and app deployment

Streamlit (version 1.32.0) (https://streamlit.io/) python package (along with numpy, pandas (version 2.2.1) (https://pandas.pydata.org/), SQLAlchemy (version 2.0.28) (https://www.sqlalchemy.org/) [for data display & managements]; mysql.connector (version 8.3.0) (https://pypi.org/project/mysql-connector-python/) [for frontend backend connection management]; pyvis (version 0.1.3.1) (https://pyvis.readthedocs.io/en/latest/), network (version 3.2.1) (https://networkx.org/), community (version 1.0.0b.1) (https://pypi.org/project/community/), python-louvain (version 0.16) (https://pypi.org/project/python-louvain/) [for side effects visualization]; RDKit (version 2023.09.6) (https://www.rdkit.org/) [for feature extraction]; joblib (version 1.4) (https://joblib.readthedocs.io/en/stable/), sklearn (version 1.4) (https://scikit-learn.org/stable/), & xgboost (version 2.0.3) (https://xgboost.readthedocs.io/en/stable/python/python_intro.html) [for drug-drug side effect prediction]; reportlab (version 4.1.0) (https://pypi.org/project/reportlab/) [for patient report generation]) was used to develop the frontend of both app, once the apps were tested and ready, the apps were deployed to the streamlit community cloud (https://streamlit.io/cloud) to make it public.

### 4.10. App interface

As a result of this project 2 app UI developed one named ‘PTTRS’ and another ‘Polypharmacy Side Effect Predictor’. The UI of 2 app discussed below.

#### 4.10.1. PTTRS interface

To use PTTRS users have to create their login username and password by the registration system of PTTRS. This login step of PTTRS is essential for multiuser management at a time from different locations of the globe. Once a user logged in, the user can find the following. Moreover, the overall workflow of PTTRS is shown in Figure 2 & some of screenshots of PTTRS is shown in Figure 3.

**Figure 2.**
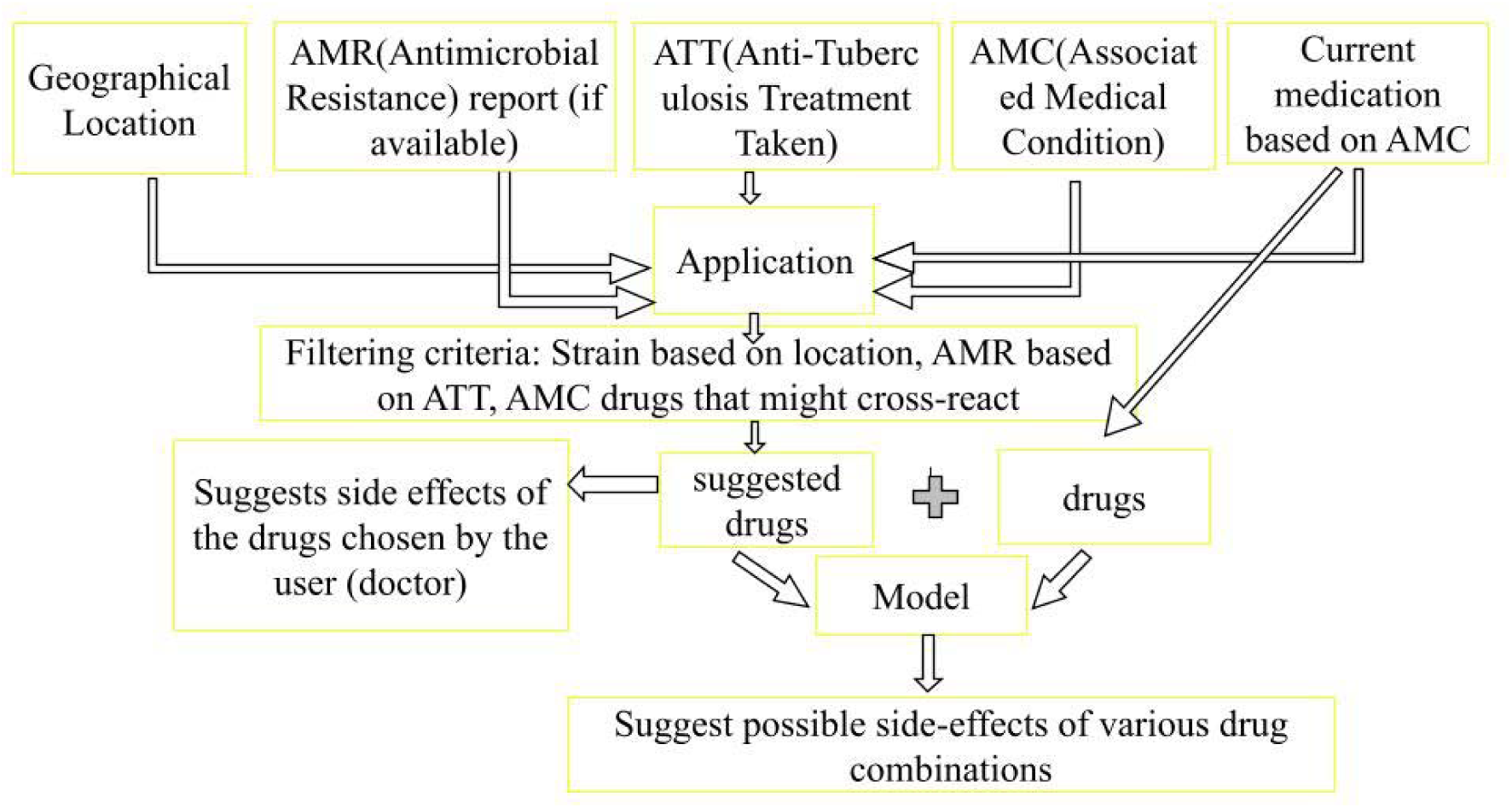
Diagrammatic representation of the overall workflow of PTTRS.

**Figure 3.**
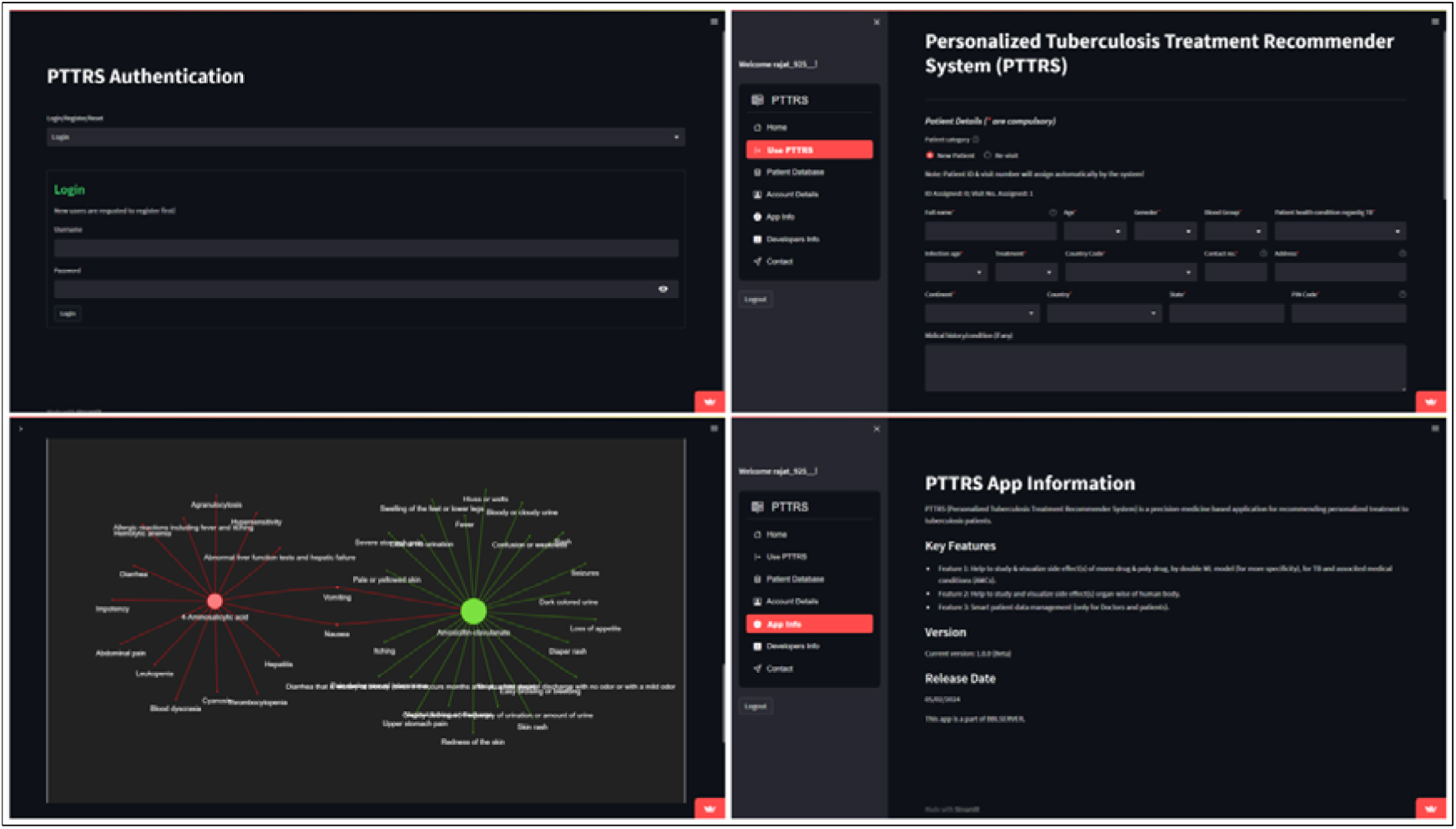
Some screenshots PTTRS app interface.

##### 4.10.1.1. Home

Here only a welcome message is shown.

##### 4.10.1.2. Use PTTRS

This is the main part of the application and is divided into 2 categories, which are the following.

##### 4.10.1.2.1. For doctors

If a new patient comes then the patient details like name, age, gender, health condition regarding TB, infection age, treatment status, etc. have to be filled. Then if any location, strain, and antimicrobial resistance strain details are available with the patient that has to be put. After that if any associated medical condition also can be put. Now all the available TB drugs and AMC (which are selected) will be displayed. From those, at least one drug can be selected to read and visualize the side effects. More than one drug needs to be selected to predict and visualize the drug-drug side effects (not applicable for peptide drugs). The prediction result also can be studied and visualized organ-wise of the human body. This facility helps doctors to provide a good drug combination to the patient so that the patient suffers from as less as polypharmacy side effects as possible. After all this, the user can select the next visit date (if required) or save the data to get the report in PDF format.

If the same patient revisits the doctor next time the doctor can track all the previous records about the patient by patient ID (every time assigned for a new patient by system), and change the drugs for the patient (by studying and visualizing the side effects) to improve TB conditions. By the patient ID the doctor can keep track of how much time a patient requires to cure of TB, how severe the patient had, what are the drugs by which the patient cured, etc.

##### 4.10.1.2.2. For others (scientists/researchers/scholars/students)

For other users only the patient-related facility is not provided. All other things remain the same.

##### 4.10.1.3. Patient database

Again, this facility is only for doctors to keep track of their patient’s related data like health condition, last visiting date, etc.; other than doctors its usage is strictly restricted.

##### 4.10.1.4. Account details

Here a user can find all information like name, gender, address, mail ID, affiliation, etc. regarding his/her account.

##### 4.10.1.5. App info

Here is all information like app version, release date, etc. available.

##### 4.10.1.6. Developers’ info

Here developers’ information is available. User may contact any of the developer if they want.

##### 4.10.1.7. Contact

Here all the contact details along with a contact form is provided, user can also reach to us via this.

#### 4.10.2. Polypharmacy side effect predictor

In this UI a user only needs to enter canonical SMILES/InChI of a drug pair, in respective fields, and hit the ‘predict side effect button’. Within a few minutes, all the side effects results will appear with probability. A few screenshots of this interface are shown in Figure 4.

**Figure 4.**
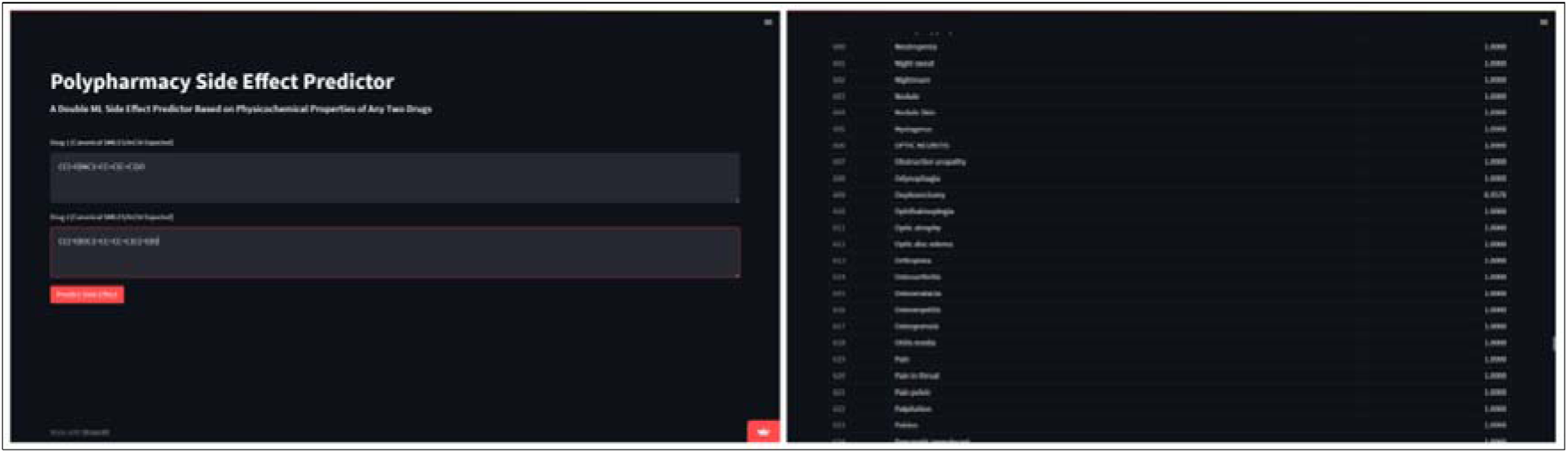
Screenshots of polypharmacy side effect predictor interface.

## **3.** Result & discussion

Through this project we developed 2 applications, first one can predict 1317 side effects for any drug pair in the globe, and the second one is an app for recommended personalized recommendation along with patients’ TB health data management system by applying the core part (i.e., side effect prediction models) of the first app. The first app is going to be the world’s first graphics user interface (GUI) based app for predict side effects of a drug pair. The second app is also going to be the world’s first GUI based app for TB drug recommendation along with data management for doctors. By the comprehensive development and analysis of our project, we have achieved significant milestones in the field of drug side effect prediction and tuberculosis (TB) management. This section discusses the outcomes and implications of our work.

### **3.1.** Model comparison

For model comparison, we only considered our generalized model, since all the existing models in this field are generalized.

In Table 8, the best performance is marked in bold font. In this case only NNPS & Decagon were considered because the comparison parameters information about other models is not available.

**Table 8.** Comparison of our generalized ML model with NNPS & Decagon model for polypharmacy side effect prediction.

| Model name | Can predict | Accuracy | F1 Score | Precision | Recall | MC C | TP | FP | FN | TN | Can take external input | GUI available |
| --- | --- | --- | --- | --- | --- | --- | --- | --- | --- | --- | --- | --- |
| Our model | **1317 side effects** | **1.00** | **1.00** | **0.99** | **1.00** | **1.00** | 112,154 | **29** | **29** | **146,062,237** | **Yes** | **Yes** |
| NNPS | 964 side effects | 0.93 | 0.94 | 0.90 | 0.98 | 0.87 | 105,153 | 12,013 | 2608 | 95,955 | No | No |
| Decagon | 964 side effects | 0.83 | 0.85 | 0.77 | 0.95 | 0.69 | **208,963** | 54,855 | 9880 | 163,988 | No | No |

In Table 9, best performance is marked in bold font.

**Table 9.** Comparison of the micro average of AUROC & AUPRC of our generalized ML model with all existing model.

| Model | AUROC | AUPRC |
| --- | --- | --- |
| Our model | **1.00** | **1.00** |
| NNPS | 0.97 | 0.95 |
| Decagon | 0.87 | 0.83 |
| Concatenated drug features | 0.79 | 0.76 |
| DeepWalk | 0.76 | 0.74 |
| DEDICOM | 0.71 | 0.64 |
| RESCAL | 0.69 | 0.61 |

In Table 10 best performance is denoted in bold font. In this comparison, NNPS and our model both show the same best efficiency in terms of AUROC and AUPRC for predicting dangerous side effects, than Decagon.

**Table 10.** Comparison of results of dangerous side effects in our model, NNPS and Decagon on AUROC and AUPRC.

| Polypharmacy side effects | AUROC |  |  | AUPRC |  |  | Evidence |
| --- | --- | --- | --- | --- | --- | --- | --- |
|  | Our model | NNPS | Decagon | Our model | NNPS | Decagon |  |
| Sarcoma | **1.00** | **1.00** | 0.79 | **1.00** | **1.00** | 0.79 |  |
| Carcinoma of the cervix | **1.00** | **1.00** | 0.88 | **1.00** | **1.00** | 0.81 |  |
| Malignant hypertension | **1.00** | **1.00** | 0.91 | **1.00** | **1.00** | 0.86 |  |
| Epidural hematoma | **1.00** | **1.00** | 0.94 | **1.00** | **1.00** | 0.91 |  |
| Oophorectomy | **1.00** | **1.00** | 0.92 | **1.00** | **1.00** | 0.91 |  |

**Table 10.** Pros and cons of Polypharmacy side effect predictor.

| Pros | Cons |
| --- | --- |
| GUI-based, interactive and compatible with any device | Polypharmacy side effect prediction may take little longer time, since the app is hosted on shared cloud |
| Helps any user to study poly polypharmacy side effects of mono as well as poly TB/AMC drugs to make smooth decisions to provide a drug pair to patient | High-speed network connection is required to use the app |

**Table 11.**
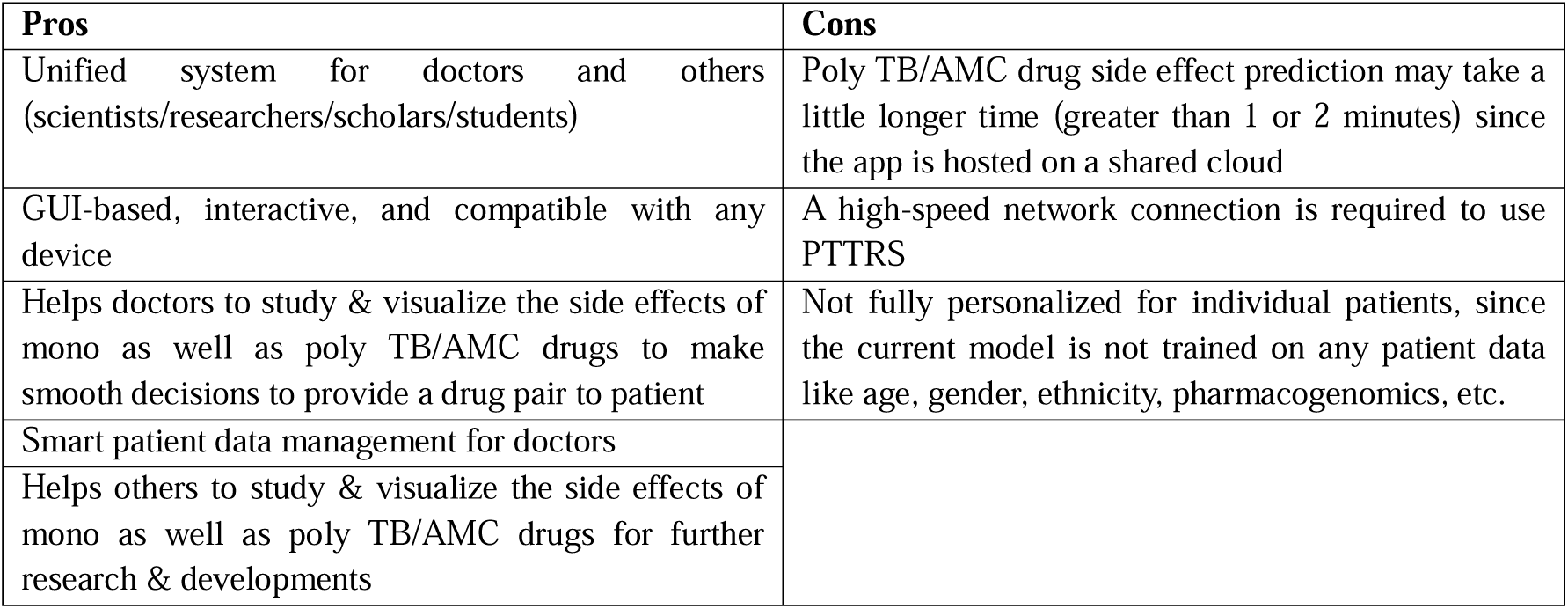
Pros & cons of PTTRS.

| Pros | Cons |
| --- | --- |
| Unified system for doctors and others (scientists/researchers/scholars/students) | Poly TB/AMC drug side effect prediction may take a little longer time (greater than 1 or 2 minutes) since the app is hosted on a shared cloud |
| GUI-based, interactive, and compatible with any device | A high-speed network connection is required to use PTTRS |
| Helps doctors to study & visualize the side effects of mono as well as poly TB/AMC drugs to make smooth decisions to provide a drug pair to patient | Not fully personalized for individual patients, since the current model is not trained on any patient data like age, gender, ethnicity, pharmacogenomics, etc. |
| Smart patient data management for doctors |  |
| Helps others to study & visualize the side effects of mono as well as poly TB/AMC drugs for further research & developments |  |

Our generalized model has demonstrated superior performance compared to existing models such as NNPS and Decagon for polypharmacy side effect prediction. The comparison, as shown in Tables 8 and 9, highlights the exceptional results in terms of accuracy, TP, FP, TN, FN, F1 score, precision, recall, MCC, AUROC, and AUPRC achieved by our model. Notably, our model can predict 1317 side effects with exceptional accuracy, making it a robust tool for healthcare professionals and researchers. We are not able to compare our specialized models but they are also robust, based on all of their validation parameters (first 20 models shown in Table 6, all are shown in supplementary file S2).

### **3.2.** Pros and cons of the apps

There are some pros and cons that we observed in case of our both app, which are discussed below in Tables 10 & 11.

Both applications offer several advantages, including GUI-based, interactive interfaces compatible with any device. However, polypharmacy side effect prediction may take longer due to shared cloud hosting, and a high-speed network connection is required for optimal usage. Despite these limitations, our applications provide valuable tools for healthcare professionals and researchers to enhance patient care and advance scientific knowledge in the field of drug side effect prediction and TB management.

Our project represents a significant contribution to the field of healthcare technology, offering innovative solutions for drug-drug side effect prediction and TB management. By leveraging machine learning algorithms and developing user-friendly applications, we have provided valuable tools to empower healthcare professionals and researchers in improving patient care and advancing scientific knowledge. We are committed to continuing our research and development efforts to address evolving challenges in healthcare management and contribute to the global fight against infectious diseases like tuberculosis.

### **3.3.** Scalability

At present, we developed the app for TB only, but it is also possible to create a single robust app, by using the same technology, for all the dangerous disease management where drug combination is given to patients. By this, we are not only providing a helping hand to all doctors to give the best drug combination to the patient (so that the patient experiences as less as side effects as possible) but also, we are creating a global patient health care data management environment, by which doctors as well as we can keep track of the health of all patient.

## **4.** Conclusion and future perspectives

Overall, this web-based application is an effort towards precision medicine which will hopefully aid in: (i) identifying side-effects of different combinations of drugs, (ii) identifying incompatibility of certain drugs in particular patients with certain medical conditions, and (iii) pave way for geared-up research as it will act like a comparatively reliable guide. This will help in timely recovery of the patient. By reducing the duration of treatment, it will also reduce the transmission of bacteria in the community. This will also lead to a reduction in the cost of treatment. Thus, it will help reduce the economic burden as a whole.

However, this opens up gates for building geared-up models for precision medicine, by incorporating other factors, such as ethnicity, geographical location, age, and gender of the patient. These will help in further enhancing the confidence level of the output. Last but not least, we know that the current approach i.e., prediction of side effects by double ML model (a generalized that can predict all side effects at a time, and individual specialized models that predict only one side effect at a time) is quite time-consuming. We are in the process of finding a better HPC facility with no restrictions like our CCF. So that, we can train our generalized model on all data points. Once we can do this, we shall modify the prediction mechanism of ‘Polypharmacy side effects predictor’, and ‘PTTRS’ and then users will be able to predict side effects for multiple drug combinations within less than a minute. Hopefully, these will be incorporated in the next version of PTTRS. Moreover, we also try to get pharmacogenomics data of patients by collaborating with the health science industry, so that we can train the model also on those data. By this, in the future, we can predict side effects individually by for any patient for any drug combination that is given for any disease, in addition to recommending drugs after a more strict screening.

## **5.** App public URL

Polypharmacy side effect predictor: https://psep-bblserver.streamlit.app/ PTTRS: https://pttrs-bblserver.streamlit.app/

## Data Availability

All data produced in the present study are available upon reasonable request to the authors.

https://pttrs-bblserver.streamlit.app/

https://psep-bblserver.streamlit.app/

## Acknowledgements

Ananya Anurag Anand is thankful to MoE-GoI for her PhD fellowship. Rajat Kumar Mondal and Baishali Sarkar are thankful to MoE-GoI for their MTech fellowship. All the authors are extremely thankful to Dr. Pramit Ghosh, Senior Scientist E, Indian Council of Medical Research (ICMR), for providing his valuable insights. Ananya Anurag Anand and Sintu Kumar Samanta are thankful to NewGen IEDC Centre at IIIT Allahabad (supported by EDII & NSTEDB, DST), for supporting and funding the project, and for collaboration opportunities. All the authors are extremely thankful to IIIT-A for providing the infrastructural facility i.e. Central Computing Facility.

## **6.** Author contributions

A.A.A.: Conceptualization, Investigation, Data mining and curation, Data Analysis, Methodology, Writing-original draft, review and editing; R.K.M.: Data mining and curation, Data Analysis, Methodology, Writing-original draft; B.S.: Data mining and curation, Data Analysis, Methodology; S.K.S.: Conceptualization, Formal Analysis, Supervision, Investigation, Resources, Writing-review & editing.

## **9.** Data availability

Data shall be made available on request.

