## Supplementary material for "Personalized Tuberculosis Treatment Recommendation System (PTTRS) version 1: A Precision-Medicine Based Application for Recommending Personalized Treatment to Tuberculosis Patients": S2 Supplementary Data

**All specialised model training & validation information**

| Model No. | Side effect can predict | Total data points | Considered data points | Duplicate data points | Train data size (80%) | Test data size (10%) | Validation data size | Training time | Accuracy | Validation score | TP | FP | TN | FN | Recall | Precision | F1 score | MSE | AUROC | AUPRC |
|---|---|---|---|---|---|---|---|---|---|---|---|---|---|---|---|---|---|---|---|---|
| model_0 | ADH inappropriate | 3018 | 3018 | 0 | 2414 | 302 | 302 | 0.25 | 0.92 | 0.93 | 279 | 0 | 0 | 23 | 0.92 | 1.00 | 0.96 | 0.3 | 0.92 | 1.00 |
| model_1 | ADVERSE DRUG EFFECT | 4758 | 4758 | 0 | 3806 | 476 | 476 | 0.25 | 0.95 | 0.93 | 452 | 0 | 0 | 24 | 0.95 | 1.00 | 0.97 | 0.2 | 0.95 | 1.00 |
| model_2 | AFIB | 15575 | 15575 | 0 | 12460 | 1558 | 1557 | 0.32 | 0.94 | 0.94 | 1467 | 0 | 0 | 91 | 0.94 | 1.00 | 0.97 | 0.23 | 0.94 | 1.00 |
| model_3 | Abdominal distension | 12369 | 12369 | 0 | 9895 | 1237 | 1237 | 0.29 | 0.95 | 0.96 | 1181 | 0 | 0 | 56 | 0.95 | 1.00 | 0.98 | 0.18 | 0.95 | 1.00 |
| model_4 | Abdominal hernia | 2750 | 2750 | 0 | 2200 | 275 | 275 | 0.24 | 0.93 | 0.96 | 255 | 0 | 0 | 20 | 0.93 | 1.00 | 0.96 | 0.29 | 0.93 | 1.00 |
| model_5 | Abdominal pain | 21410 | 21410 | 0 | 17128 | 2141 | 2141 | 0.35 | 0.96 | 0.96 | 2047 | 0 | 0 | 94 | 0.96 | 1.00 | 0.98 | 0.18 | 0.96 | 1.00 |
| model_6 | Abdominal pain upper | 13163 | 13163 | 0 | 10530 | 1317 | 1316 | 0.31 | 0.96 | 0.97 | 1264 | 0 | 0 | 53 | 0.96 | 1.00 | 0.98 | 0.16 | 0.96 | 1.00 |
| model_7 | Abnormal ECG | 5523 | 5523 | 0 | 4418 | 553 | 552 | 0.27 | 0.95 | 0.95 | 528 | 0 | 0 | 25 | 0.95 | 1.00 | 0.98 | 0.18 | 0.95 | 1.00 |
| model_8 | Abnormal EEG | 2679 | 2679 | 0 | 2143 | 268 | 268 | 0.25 | 0.93 | 0.92 | 250 | 0 | 0 | 18 | 0.93 | 1.00 | 0.97 | 0.27 | 0.93 | 1.00 |
| model_9 | Abnormal Gait | 13801 | 13801 | 0 | 11040 | 1381 | 1380 | 0.31 | 0.97 | 0.96 | 1333 | 0 | 0 | 48 | 0.97 | 1.00 | 0.98 | 0.14 | 0.97 | 1.00 |
| model_10 | Abnormal LFTs | 10960 | 10960 | 0 | 8768 | 1096 | 1096 | 0.28 | 0.95 | 0.96 | 1036 | 0 | 0 | 60 | 0.95 | 1.00 | 0.97 | 0.22 | 0.95 | 1.00 |
| model_11 | Abnormal Laboratory Findings | 6238 | 6238 | 0 | 4990 | 624 | 624 | 0.26 | 0.95 | 0.95 | 591 | 0 | 0 | 33 | 0.95 | 1.00 | 0.97 | 0.21 | 0.95 | 1.00 |
| model_12 | Abnormal cervical smear | 663 | 663 | 0 | 530 | 67 | 66 | 0.22 | 0.97 | 0.98 | 65 | 0 | 0 | 2 | 0.97 | 1.00 | 0.98 | 0.12 | 0.97 | 1.00 |
| model_13 | Abnormal mammogram | 715 | 715 | 0 | 572 | 72 | 71 | 0.23 | 0.96 | 0.97 | 69 | 0 | 0 | 3 | 0.96 | 1.00 | 0.98 | 0.17 | 0.96 | 1.00 |
| model_14 | Abnormal movements | 7126 | 7126 | 0 | 5700 | 713 | 713 | 0.26 | 0.95 | 0.96 | 679 | 0 | 0 | 34 | 0.95 | 1.00 | 0.98 | 0.19 | 0.95 | 1.00 |
| model_15 | Abnormal vision | 1411 | 1411 | 0 | 1128 | 142 | 141 | 0.23 | 0.92 | 0.91 | 130 | 0 | 0 | 12 | 0.92 | 1.00 | 0.96 | 0.34 | 0.92 | 1.00 |
| model_16 | Abortion missed | 122 | 588 | 466 | 470 | 59 | 59 | 0.24 | 0.85 | 0.88 | 50 | 0 | 0 | 9 | 0.85 | 1.00 | 0.92 | 0.61 | 0.85 | 1.00 |
| model_17 | Abortion spontaneous | 1482 | 1482 | 0 | 1185 | 149 | 148 | 0.24 | 0.95 | 0.99 | 142 | 0 | 0 | 7 | 0.95 | 1.00 | 0.98 | 0.19 | 0.95 | 1.00 |
| model_18 | Abscess | 6782 | 6782 | 0 | 5425 | 679 | 678 | 0.26 | 0.93 | 0.94 | 631 | 0 | 0 | 48 | 0.93 | 1.00 | 0.96 | 0.28 | 0.93 | 1.00 |
| model_19 | Abuse | 2310 | 2310 | 0 | 1848 | 231 | 231 | 0.24 | 0.95 | 0.97 | 220 | 0 | 0 | 11 | 0.95 | 1.00 | 0.98 | 0.19 | 0.95 | 1.00 |
| model_20 | Acanthosis nigricans | 221 | 588 | 367 | 470 | 59 | 59 | 0.23 | 0.92 | 0.95 | 54 | 0 | 0 | 5 | 0.92 | 1.00 | 0.96 | 0.34 | 0.92 | 1.00 |
| model_21 | Aching joints | 18061 | 18061 | 0 | 14448 | 1807 | 1806 | 0.32 | 0.95 | 0.95 | 1711 | 0 | 0 | 96 | 0.95 | 1.00 | 0.97 | 0.21 | 0.95 | 1.00 |
| model_22 | Aching muscles | 12590 | 12590 | 0 | 10072 | 1259 | 1259 | 0.29 | 0.96 | 0.96 | 1212 | 0 | 0 | 47 | 0.96 | 1.00 | 0.98 | 0.15 | 0.96 | 1.00 |
| model_23 | Acid reflux | 14122 | 14122 | 0 | 11297 | 1413 | 1412 | 0.32 | 0.96 | 0.97 | 1359 | 0 | 0 | 54 | 0.96 | 1.00 | 0.98 | 0.15 | 0.96 | 1.00 |
| model_24 | Acidosis | 8621 | 8621 | 0 | 6896 | 863 | 862 | 0.27 | 0.95 | 0.95 | 816 | 0 | 0 | 47 | 0.95 | 1.00 | 0.97 | 0.22 | 0.95 | 1.00 |
| model_25 | Acne | 3495 | 3495 | 0 | 2796 | 350 | 349 | 0.24 | 0.96 | 0.96 | 337 | 0 | 0 | 13 | 0.96 | 1.00 | 0.98 | 0.15 | 0.96 | 1.00 |
| model_26 | Acne rosacea | 1940 | 1940 | 0 | 1552 | 194 | 194 | 0.23 | 0.96 | 0.97 | 187 | 0 | 0 | 7 | 0.96 | 1.00 | 0.98 | 0.14 | 0.96 | 1.00 |
| model_27 | Acquired immune deficiency syndrome | 448 | 588 | 140 | 470 | 59 | 59 | 0.23 | 0.92 | 0.88 | 54 | 0 | 0 | 5 | 0.92 | 1.00 | 0.96 | 0.34 | 0.92 | 1.00 |
| model_28 | Acromegaly | 14 | 588 | 574 | 470 | 59 | 59 | 0.25 | 0 | 0 | 0 | 0 | 0 | 59 | 0 | 0.00 | 0 | 4 | 0 | 1.00 |
| model_29 | Actinic keratosis | 1792 | 1792 | 0 | 1433 | 180 | 179 | 0.23 | 0.95 | 0.97 | 171 | 0 | 0 | 9 | 0.95 | 1.00 | 0.97 | 0.2 | 0.95 | 1.00 |
| model_30 | Acute Respiratory Distress Syndrome | 7161 | 7161 | 0 | 5728 | 717 | 716 | 0.28 | 0.94 | 0.93 | 675 | 0 | 0 | 42 | 0.94 | 1.00 | 0.97 | 0.23 | 0.94 | 1.00 |
| model_31 | Acute brain syndrome | 7111 | 7111 | 0 | 5688 | 712 | 711 | 0.26 | 0.94 | 0.95 | 669 | 0 | 0 | 43 | 0.94 | 1.00 | 0.97 | 0.24 | 0.94 | 1.00 |
| model_32 | Acute bronchitis | 5343 | 5343 | 0 | 4274 | 535 | 534 | 0.26 | 0.94 | 0.97 | 502 | 0 | 0 | 33 | 0.94 | 1.00 | 0.97 | 0.25 | 0.94 | 1.00 |
| model_33 | Acute febrile neutrophilic dermatosis | 360 | 588 | 228 | 470 | 59 | 59 | 0.23 | 0.97 | 0.92 | 57 | 0 | 0 | 2 | 0.97 | 1.00 | 0.98 | 0.14 | 0.97 | 1.00 |
| model_34 | Acute glomerulonephritis | 80 | 588 | 508 | 470 | 59 | 59 | 0.24 | 0.63 | 0.56 | 37 | 0 | 0 | 22 | 0.63 | 1.00 | 0.77 | 1.49 | 0.63 | 1.00 |
| model_35 | Acute intermittent porphyria | 71 | 588 | 517 | 470 | 59 | 59 | 0.25 | 0.75 | 0.81 | 44 | 0 | 0 | 15 | 0.75 | 1.00 | 0.85 | 1.02 | 0.75 | 1.00 |
| model_36 | Acute kidney failure | 17945 | 17945 | 0 | 14356 | 1795 | 1794 | 0.32 | 0.94 | 0.95 | 1686 | 0 | 0 | 109 | 0.94 | 1.00 | 0.97 | 0.24 | 0.94 | 1.00 |
| model_37 | Acute myeloblastic leukemia | 2576 | 2576 | 0 | 2060 | 258 | 258 | 0.24 | 0.87 | 0.92 | 225 | 0 | 0 | 33 | 0.87 | 1.00 | 0.93 | 0.51 | 0.87 | 1.00 |
| model_38 | Acute otitis media | 466 | 588 | 122 | 470 | 59 | 59 | 0.21 | 0.9 | 0.9 | 53 | 0 | 0 | 6 | 0.9 | 1.00 | 0.95 | 0.41 | 0.9 | 1.00 |
| model_39 | Acute pancreatitis | 6128 | 6128 | 0 | 4902 | 613 | 613 | 0.26 | 0.94 | 0.95 | 578 | 0 | 0 | 35 | 0.94 | 1.00 | 0.97 | 0.23 | 0.94 | 1.00 |
| model_40 | Acute psychosis | 890 | 890 | 0 | 712 | 89 | 89 | 0.23 | 0.96 | 0.96 | 85 | 0 | 0 | 4 | 0.96 | 1.00 | 0.98 | 0.18 | 0.96 | 1.00 |
| model_41 | Acute sinusitis | 2984 | 2984 | 0 | 2387 | 299 | 298 | 0.24 | 0.96 | 0.96 | 286 | 0 | 0 | 13 | 0.96 | 1.00 | 0.98 | 0.17 | 0.96 | 1.00 |
| model_42 | Adenitis | 1655 | 1655 | 0 | 1324 | 166 | 165 | 0.23 | 0.95 | 0.95 | 158 | 0 | 0 | 8 | 0.95 | 1.00 | 0.98 | 0.19 | 0.95 | 1.00 |
| model_43 | Adenocarcinoma | 7474 | 7474 | 0 | 5979 | 748 | 747 | 0.27 | 0.95 | 0.94 | 714 | 0 | 0 | 34 | 0.95 | 1.00 | 0.98 | 0.18 | 0.95 | 1.00 |

| | | | | | | | | | | | | | | | | | | | | |
|---|---|---|---|---|---|---|---|---|---|---|---|---|---|---|---|---|---|---|---|---|
| model_44 | Adenoid hypertrophy | 100 | 588 | 488 | 470 | 59 | 59 | 0.24 | 0.46 | 0.47 | 27 | 0 | 0 | 32 | 0.46 | 1.00 | 0.63 | 2.17 | 0.46 | 1.00 |
| model_45 | Adenoma | 2018 | 2018 | 0 | 1614 | 202 | 202 | 0.23 | 0.95 | 0.97 | 191 | 0 | 0 | 11 | 0.95 | 1.00 | 0.97 | 0.22 | 0.95 | 1.00 |
| model_46 | Adenomyosis | 316 | 588 | 272 | 470 | 59 | 59 | 0.23 | 0.92 | 0.92 | 54 | 0 | 0 | 5 | 0.92 | 1.00 | 0.96 | 0.34 | 0.92 | 1.00 |
| model_47 | Adenopathy | 9159 | 9159 | 0 | 7327 | 916 | 916 | 0.26 | 0.94 | 0.94 | 863 | 0 | 0 | 53 | 0.94 | 1.00 | 0.97 | 0.23 | 0.94 | 1.00 |
| model_48 | Adjustment disorder | 972 | 972 | 0 | 777 | 98 | 97 | 0.23 | 0.98 | 0.9 | 96 | 0 | 0 | 2 | 0.98 | 1.00 | 0.99 | 0.08 | 0.98 | 1.00 |
| model_49 | Adrenal carcinoma | 68 | 588 | 520 | 470 | 59 | 59 | 0.26 | 0.73 | 0.69 | 43 | 0 | 0 | 16 | 0.73 | 1.00 | 0.84 | 1.08 | 0.73 | 1.00 |
| model_50 | Adrenal insufficiency | 3080 | 3080 | 0 | 2464 | 308 | 308 | 0.24 | 0.96 | 0.94 | 297 | 0 | 0 | 11 | 0.96 | 1.00 | 0.98 | 0.14 | 0.96 | 1.00 |
| model_51 | Adynamic ileus | 8164 | 8164 | 0 | 6531 | 817 | 816 | 0.27 | 0.94 | 0.94 | 766 | 0 | 0 | 51 | 0.94 | 1.00 | 0.97 | 0.25 | 0.94 | 1.00 |
| model_52 | Agitated | 14192 | 14192 | 0 | 11353 | 1420 | 1419 | 0.31 | 0.95 | 0.95 | 1347 | 0 | 0 | 73 | 0.95 | 1.00 | 0.97 | 0.21 | 0.95 | 1.00 |
| model_53 | Agnosia | 430 | 588 | 158 | 470 | 59 | 59 | 0.22 | 0.92 | 0.88 | 54 | 0 | 0 | 5 | 0.92 | 1.00 | 0.96 | 0.34 | 0.92 | 1.00 |
| model_54 | Agoraphobia | 1153 | 1153 | 0 | 922 | 116 | 115 | 0.23 | 0.95 | 0.97 | 110 | 0 | 0 | 6 | 0.95 | 1.00 | 0.97 | 0.21 | 0.95 | 1.00 |
| model_55 | Agranulocytoses | 4279 | 4279 | 0 | 3423 | 428 | 428 | 0.25 | 0.92 | 0.92 | 395 | 0 | 0 | 33 | 0.92 | 1.00 | 0.96 | 0.31 | 0.92 | 1.00 |
| model_56 | Albuminuria | 139 | 588 | 449 | 470 | 59 | 59 | 0.24 | 0.92 | 0.85 | 54 | 0 | 0 | 5 | 0.92 | 1.00 | 0.96 | 0.34 | 0.92 | 1.00 |
| model_57 | Alcohol abuse | 219 | 588 | 369 | 470 | 59 | 59 | 0.25 | 0.83 | 0.95 | 49 | 0 | 0 | 10 | 0.83 | 1.00 | 0.91 | 0.68 | 0.83 | 1.00 |
| model_58 | Alcohol consumption | 2868 | 2868 | 0 | 2294 | 287 | 287 | 0.24 | 0.96 | 0.93 | 275 | 0 | 0 | 12 | 0.96 | 1.00 | 0.98 | 0.17 | 0.96 | 1.00 |
| model_59 | Alcoholic cirrhosis | 92 | 588 | 496 | 470 | 59 | 59 | 0.24 | 0.73 | 0.83 | 43 | 0 | 0 | 16 | 0.73 | 1.00 | 0.84 | 1.08 | 0.73 | 1.00 |
| model_60 | Alcoholic intoxication | 1487 | 1487 | 0 | 1189 | 149 | 149 | 0.23 | 0.97 | 0.88 | 144 | 0 | 0 | 5 | 0.97 | 1.00 | 0.98 | 0.13 | 0.97 | 1.00 |
| model_61 | Alkalosis | 356 | 588 | 232 | 470 | 59 | 59 | 0.23 | 0.88 | 0.95 | 52 | 0 | 0 | 7 | 0.88 | 1.00 | 0.94 | 0.47 | 0.88 | 1.00 |
| model_62 | Allergic alveolitis | 1147 | 1147 | 0 | 917 | 115 | 115 | 0.22 | 0.9 | 0.95 | 103 | 0 | 0 | 12 | 0.9 | 1.00 | 0.94 | 0.42 | 0.9 | 1.00 |
| model_63 | Allergic bronchitis | 13 | 588 | 575 | 470 | 59 | 59 | 0.23 | 0 | 0 | 0 | 0 | 0 | 59 | 0 | 0.00 | 0 | 4 | 0 | 1.00 |
| model_64 | Allergic dermatitis | 12906 | 12906 | 0 | 10324 | 1291 | 1291 | 0.3 | 0.95 | 0.94 | 1230 | 0 | 0 | 61 | 0.95 | 1.00 | 0.98 | 0.19 | 0.95 | 1.00 |
| model_65 | Allergic rhinitis | 4695 | 4695 | 0 | 3756 | 470 | 469 | 0.26 | 0.97 | 0.96 | 454 | 0 | 0 | 16 | 0.97 | 1.00 | 0.98 | 0.14 | 0.97 | 1.00 |
| model_66 | Allergic vasculitis | 2052 | 2052 | 0 | 1641 | 206 | 205 | 0.24 | 0.97 | 0.91 | 199 | 0 | 0 | 7 | 0.97 | 1.00 | 0.98 | 0.14 | 0.97 | 1.00 |
| model_67 | Allergies | 11218 | 11218 | 0 | 8974 | 1122 | 1122 | 0.29 | 0.96 | 0.97 | 1079 | 0 | 0 | 43 | 0.96 | 1.00 | 0.98 | 0.15 | 0.96 | 1.00 |
| model_68 | Alopecia | 8438 | 8438 | 0 | 6750 | 844 | 844 | 0.27 | 0.97 | 0.97 | 819 | 0 | 0 | 25 | 0.97 | 1.00 | 0.98 | 0.12 | 0.97 | 1.00 |
| model_69 | Als | 792 | 792 | 0 | 633 | 80 | 79 | 0.23 | 0.95 | 0.91 | 76 | 0 | 0 | 4 | 0.95 | 1.00 | 0.97 | 0.2 | 0.95 | 1.00 |
| model_70 | Altered Bowel Function | 1681 | 1681 | 0 | 1344 | 169 | 168 | 0.24 | 0.92 | 0.92 | 155 | 0 | 0 | 14 | 0.92 | 1.00 | 0.96 | 0.33 | 0.92 | 1.00 |
| model_71 | Alveolar proteinosis | 150 | 588 | 438 | 470 | 59 | 59 | 0.25 | 0.78 | 0.85 | 46 | 0 | 0 | 13 | 0.78 | 1.00 | 0.88 | 0.88 | 0.78 | 1.00 |
| model_72 | Amaurosis fugax | 361 | 588 | 227 | 470 | 59 | 59 | 0.23 | 0.92 | 0.95 | 54 | 0 | 0 | 5 | 0.92 | 1.00 | 0.96 | 0.34 | 0.92 | 1.00 |
| model_73 | Amblyopia | 355 | 588 | 233 | 470 | 59 | 59 | 0.23 | 0.97 | 0.95 | 57 | 0 | 0 | 2 | 0.97 | 1.00 | 0.98 | 0.14 | 0.97 | 1.00 |
| model_74 | Amenorrhea | 1756 | 1756 | 0 | 1404 | 176 | 176 | 0.23 | 0.98 | 0.94 | 172 | 0 | 0 | 4 | 0.98 | 1.00 | 0.99 | 0.09 | 0.98 | 1.00 |
| model_75 | Amentia | 4155 | 4155 | 0 | 3324 | 416 | 415 | 0.24 | 0.97 | 0.95 | 402 | 0 | 0 | 14 | 0.97 | 1.00 | 0.98 | 0.13 | 0.97 | 1.00 |
| model_76 | Amnesia | 14099 | 14099 | 0 | 11279 | 1410 | 1410 | 0.3 | 0.96 | 0.96 | 1358 | 0 | 0 | 52 | 0.96 | 1.00 | 0.98 | 0.15 | 0.96 | 1.00 |
| model_77 | Amyloidosis | 833 | 833 | 0 | 666 | 84 | 83 | 0.23 | 0.92 | 0.96 | 77 | 0 | 0 | 7 | 0.92 | 1.00 | 0.96 | 0.33 | 0.92 | 1.00 |
| model_78 | Amyotrophy | 3557 | 3557 | 0 | 2845 | 356 | 356 | 0.24 | 0.94 | 0.95 | 335 | 0 | 0 | 21 | 0.94 | 1.00 | 0.97 | 0.24 | 0.94 | 1.00 |
| model_79 | Anaemia | 27006 | 27006 | 0 | 21604 | 2701 | 2701 | 0.4 | 0.94 | 0.94 | 2552 | 0 | 0 | 149 | 0.94 | 1.00 | 0.97 | 0.22 | 0.94 | 1.00 |
| model_80 | Anaemia hypochromic | 510 | 588 | 78 | 470 | 59 | 59 | 0.25 | 0.92 | 0.92 | 54 | 0 | 0 | 5 | 0.92 | 1.00 | 0.96 | 0.34 | 0.92 | 1.00 |
| model_81 | Anal fissure | 2013 | 2013 | 0 | 1610 | 202 | 201 | 0.22 | 0.95 | 0.96 | 192 | 0 | 0 | 10 | 0.95 | 1.00 | 0.97 | 0.2 | 0.95 | 1.00 |
| model_82 | Anal fistula | 912 | 912 | 0 | 729 | 92 | 91 | 0.24 | 0.98 | 0.99 | 90 | 0 | 0 | 2 | 0.98 | 1.00 | 0.99 | 0.09 | 0.98 | 1.00 |
| model_83 | Anal pruritus | 239 | 588 | 349 | 470 | 59 | 59 | 0.25 | 0.93 | 0.92 | 55 | 0 | 0 | 4 | 0.93 | 1.00 | 0.96 | 0.27 | 0.93 | 1.00 |
| model_84 | Anaphylactic reaction | 5893 | 5893 | 0 | 4714 | 590 | 589 | 0.25 | 0.95 | 0.94 | 560 | 0 | 0 | 30 | 0.95 | 1.00 | 0.97 | 0.2 | 0.95 | 1.00 |
| model_85 | Anemia aplastic | 1670 | 1670 | 0 | 1336 | 167 | 167 | 0.24 | 0.96 | 0.93 | 160 | 0 | 0 | 7 | 0.96 | 1.00 | 0.98 | 0.17 | 0.96 | 1.00 |
| model_86 | Aneurysm | 2439 | 2439 | 0 | 1951 | 244 | 244 | 0.24 | 0.96 | 0.95 | 235 | 0 | 0 | 9 | 0.96 | 1.00 | 0.98 | 0.15 | 0.96 | 1.00 |
| model_87 | Angiitis | 5599 | 5599 | 0 | 4479 | 560 | 560 | 0.25 | 0.94 | 0.93 | 524 | 0 | 0 | 36 | 0.94 | 1.00 | 0.97 | 0.26 | 0.94 | 1.00 |
| model_88 | Angina | 9889 | 9889 | 0 | 7911 | 989 | 989 | 0.28 | 0.95 | 0.95 | 943 | 0 | 0 | 46 | 0.95 | 1.00 | 0.98 | 0.19 | 0.95 | 1.00 |
| model_89 | Angioedema | 4849 | 4849 | 0 | 3879 | 485 | 485 | 0.26 | 0.95 | 0.96 | 462 | 0 | 0 | 23 | 0.95 | 1.00 | 0.98 | 0.19 | 0.95 | 1.00 |
| model_90 | Angiopathy | 5197 | 5197 | 0 | 4157 | 520 | 520 | 0.24 | 0.94 | 0.93 | 487 | 0 | 0 | 33 | 0.94 | 1.00 | 0.97 | 0.25 | 0.94 | 1.00 |
| model_91 | Angle closure glaucoma | 470 | 588 | 118 | 470 | 59 | 59 | 0.22 | 0.95 | 0.92 | 56 | 0 | 0 | 3 | 0.95 | 1.00 | 0.97 | 0.2 | 0.95 | 1.00 |
| model_92 | Animal bite | 826 | 826 | 0 | 660 | 83 | 83 | 0.23 | 0.96 | 0.99 | 80 | 0 | 0 | 3 | 0.96 | 1.00 | 0.98 | 0.14 | 0.96 | 1.00 |
| model_93 | Anisocoria | 992 | 992 | 0 | 793 | 100 | 99 | 0.23 | 0.95 | 0.91 | 95 | 0 | 0 | 5 | 0.95 | 1.00 | 0.97 | 0.2 | 0.95 | 1.00 |

| | | | | | | | | | | | | | | | | | | | | |
|---|---|---|---|---|---|---|---|---|---|---|---|---|---|---|---|---|---|---|---|---|
| model_94 | Ankle fracture | 3331 | 3331 | 0 | 2664 | 334 | 333 | 0.25 | 0.97 | 0.95 | 324 | 0 | 0 | 10 | 0.97 | 1.00 | 0.98 | 0.12 | 0.97 | 1.00 |
| model_95 | Ankylosing spondylitis | 439 | 588 | 149 | 470 | 59 | 59 | 0.22 | 0.97 | 0.9 | 57 | 0 | 0 | 2 | 0.97 | 1.00 | 0.98 | 0.14 | 0.97 | 1.00 |
| model_96 | Anogenital warts | 327 | 588 | 261 | 470 | 59 | 59 | 0.22 | 0.95 | 0.9 | 56 | 0 | 0 | 3 | 0.95 | 1.00 | 0.97 | 0.2 | 0.95 | 1.00 |
| model_97 | Anorexia | 18177 | 18177 | 0 | 14541 | 1818 | 1818 | 0.32 | 0.95 | 0.95 | 1734 | 0 | 0 | 84 | 0.95 | 1.00 | 0.98 | 0.18 | 0.95 | 1.00 |
| model_98 | Anosmia | 2461 | 2461 | 0 | 1968 | 247 | 246 | 0.24 | 0.96 | 0.97 | 237 | 0 | 0 | 10 | 0.96 | 1.00 | 0.98 | 0.16 | 0.96 | 1.00 |
| model_99 | Anovulatory | 12 | 588 | 576 | 470 | 59 | 59 | 0.24 | 0 | 0 | 0 | 0 | 0 | 59 | 0 | 0.00 | 0 | 4 | 0 | 1.00 |
| model_100 | Anthrax | 9 | 588 | 579 | 470 | 59 | 59 | 0.25 | 0 | 0 | 0 | 0 | 0 | 59 | 0 | 0.00 | 0 | 4 | 0 | 1.00 |
| model_101 | Antinuclear antibody positive | 3111 | 3111 | 0 | 2488 | 312 | 311 | 0.25 | 0.96 | 0.97 | 299 | 0 | 0 | 13 | 0.96 | 1.00 | 0.98 | 0.17 | 0.96 | 1.00 |
| model_102 | Anxiety | 18980 | 18980 | 0 | 15184 | 1898 | 1898 | 0.32 | 0.96 | 0.96 | 1818 | 0 | 0 | 80 | 0.96 | 1.00 | 0.98 | 0.17 | 0.96 | 1.00 |
| model_103 | Aortic aneurysm | 3101 | 3101 | 0 | 2480 | 311 | 310 | 0.25 | 0.94 | 0.95 | 292 | 0 | 0 | 19 | 0.94 | 1.00 | 0.97 | 0.24 | 0.94 | 1.00 |
| model_104 | Aortic regurgitation | 5599 | 5599 | 0 | 4479 | 560 | 560 | 0.26 | 0.96 | 0.95 | 540 | 0 | 0 | 20 | 0.96 | 1.00 | 0.98 | 0.14 | 0.96 | 1.00 |
| model_105 | Aortic stenosis | 3833 | 3833 | 0 | 3066 | 384 | 383 | 0.25 | 0.96 | 0.96 | 370 | 0 | 0 | 14 | 0.96 | 1.00 | 0.98 | 0.15 | 0.96 | 1.00 |
| model_106 | Aphasia | 5643 | 5643 | 0 | 4514 | 565 | 564 | 0.27 | 0.94 | 0.94 | 531 | 0 | 0 | 34 | 0.94 | 1.00 | 0.97 | 0.24 | 0.94 | 1.00 |
| model_107 | Aphonia | 1613 | 1613 | 0 | 1290 | 162 | 161 | 0.23 | 0.98 | 0.96 | 159 | 0 | 0 | 3 | 0.98 | 1.00 | 0.99 | 0.07 | 0.98 | 1.00 |
| model_108 | Aphthous stomatitis | 2598 | 2598 | 0 | 2078 | 260 | 260 | 0.24 | 0.97 | 0.95 | 253 | 0 | 0 | 7 | 0.97 | 1.00 | 0.99 | 0.11 | 0.97 | 1.00 |
| model_109 | Aplasia pure red cell | 2183 | 2183 | 0 | 1746 | 219 | 218 | 0.24 | 0.94 | 0.91 | 205 | 0 | 0 | 14 | 0.94 | 1.00 | 0.97 | 0.26 | 0.94 | 1.00 |
| model_110 | Apnea | 4030 | 4030 | 0 | 3224 | 403 | 403 | 0.25 | 0.95 | 0.95 | 381 | 0 | 0 | 22 | 0.95 | 1.00 | 0.97 | 0.22 | 0.95 | 1.00 |
| model_111 | Apoplexy | 15235 | 15235 | 0 | 12188 | 1524 | 1523 | 0.31 | 0.96 | 0.97 | 1461 | 0 | 0 | 63 | 0.96 | 1.00 | 0.98 | 0.17 | 0.96 | 1.00 |
| model_112 | Appendectomy | 1190 | 1190 | 0 | 952 | 119 | 119 | 0.22 | 0.96 | 0.92 | 114 | 0 | 0 | 5 | 0.96 | 1.00 | 0.98 | 0.17 | 0.96 | 1.00 |
| model_113 | Appendicitis | 1805 | 1805 | 0 | 1444 | 181 | 180 | 0.23 | 0.96 | 0.92 | 174 | 0 | 0 | 7 | 0.96 | 1.00 | 0.98 | 0.15 | 0.96 | 1.00 |
| model_114 | Apraxia | 912 | 912 | 0 | 729 | 92 | 91 | 0.24 | 0.95 | 0.93 | 87 | 0 | 0 | 5 | 0.95 | 1.00 | 0.97 | 0.22 | 0.95 | 1.00 |
| model_115 | Aptyalism | 9525 | 9525 | 0 | 7620 | 953 | 952 | 0.28 | 0.97 | 0.97 | 928 | 0 | 0 | 25 | 0.97 | 1.00 | 0.99 | 0.1 | 0.97 | 1.00 |
| model_116 | Arachnoiditis | 816 | 816 | 0 | 652 | 82 | 82 | 0.23 | 0.99 | 0.93 | 81 | 0 | 0 | 1 | 0.99 | 1.00 | 0.99 | 0.05 | 0.99 | 1.00 |
| model_117 | Arrhythmia | 12409 | 12409 | 0 | 9927 | 1241 | 1241 | 0.29 | 0.96 | 0.96 | 1195 | 0 | 0 | 46 | 0.96 | 1.00 | 0.98 | 0.15 | 0.96 | 1.00 |
| model_118 | Arterial insufficiency | 337 | 588 | 251 | 470 | 59 | 59 | 0.24 | 0.88 | 0.95 | 52 | 0 | 0 | 7 | 0.88 | 1.00 | 0.94 | 0.47 | 0.88 | 1.00 |
| model_119 | Arterial pressure NOS decreased | 28568 | 28568 | 0 | 22854 | 2857 | 2857 | 0.41 | 0.95 | 0.95 | 2711 | 0 | 0 | 146 | 0.95 | 1.00 | 0.97 | 0.2 | 0.95 | 1.00 |
| model_120 | Arterial pressure NOS increased | 13302 | 13302 | 0 | 10641 | 1331 | 1330 | 0.3 | 0.96 | 0.96 | 1282 | 0 | 0 | 49 | 0.96 | 1.00 | 0.98 | 0.15 | 0.96 | 1.00 |
| model_121 | Arteriosclerosis | 7225 | 7225 | 0 | 5780 | 723 | 722 | 0.27 | 0.96 | 0.97 | 694 | 0 | 0 | 29 | 0.96 | 1.00 | 0.98 | 0.16 | 0.96 | 1.00 |
| model_122 | Arteriosclerotic heart disease | 13482 | 13482 | 0 | 10785 | 1349 | 1348 | 0.3 | 0.97 | 0.97 | 1302 | 0 | 0 | 47 | 0.97 | 1.00 | 0.98 | 0.14 | 0.97 | 1.00 |
| model_123 | Arteritis | 379 | 588 | 209 | 470 | 59 | 59 | 0.23 | 0.98 | 0.95 | 58 | 0 | 0 | 1 | 0.98 | 1.00 | 0.99 | 0.07 | 0.98 | 1.00 |
| model_124 | Arthritis | 9600 | 9600 | 0 | 7680 | 960 | 960 | 0.28 | 0.96 | 0.97 | 925 | 0 | 0 | 35 | 0.96 | 1.00 | 0.98 | 0.15 | 0.96 | 1.00 |
| model_125 | Arthritis bacterial | 1465 | 1465 | 0 | 1172 | 147 | 146 | 0.24 | 0.97 | 0.92 | 142 | 0 | 0 | 5 | 0.97 | 1.00 | 0.98 | 0.14 | 0.97 | 1.00 |
| model_126 | Arthritis infective | 1751 | 1751 | 0 | 1400 | 176 | 175 | 0.22 | 0.93 | 0.95 | 164 | 0 | 0 | 12 | 0.93 | 1.00 | 0.96 | 0.27 | 0.93 | 1.00 |
| model_127 | Arthritis rheumatoid | 5785 | 5785 | 0 | 4628 | 579 | 578 | 0.25 | 0.98 | 0.97 | 566 | 0 | 0 | 13 | 0.98 | 1.00 | 0.99 | 0.09 | 0.98 | 1.00 |
| model_128 | Arthropathy | 7766 | 7766 | 0 | 6212 | 777 | 777 | 0.28 | 0.96 | 0.97 | 746 | 0 | 0 | 31 | 0.96 | 1.00 | 0.98 | 0.16 | 0.96 | 1.00 |
| model_129 | Ascites | 8979 | 8979 | 0 | 7183 | 898 | 898 | 0.28 | 0.93 | 0.95 | 835 | 0 | 0 | 63 | 0.93 | 1.00 | 0.96 | 0.28 | 0.93 | 1.00 |
| model_130 | Aseptic meningitis | 1105 | 1105 | 0 | 884 | 111 | 110 | 0.22 | 0.97 | 0.95 | 108 | 0 | 0 | 3 | 0.97 | 1.00 | 0.99 | 0.11 | 0.97 | 1.00 |
| model_131 | Aseptic necrosis bone | 9279 | 9279 | 0 | 7423 | 928 | 928 | 0.28 | 0.96 | 0.95 | 890 | 0 | 0 | 38 | 0.96 | 1.00 | 0.98 | 0.16 | 0.96 | 1.00 |
| model_132 | Aspartate Aminotransferase Increase | 13117 | 13117 | 0 | 10493 | 1312 | 1312 | 0.3 | 0.94 | 0.93 | 1233 | 0 | 0 | 79 | 0.94 | 1.00 | 0.97 | 0.24 | 0.94 | 1.00 |
| model_133 | Aspergillosis | 2572 | 2572 | 0 | 2057 | 258 | 257 | 0.23 | 0.9 | 0.94 | 233 | 0 | 0 | 25 | 0.9 | 1.00 | 0.95 | 0.39 | 0.9 | 1.00 |
| model_134 | Aspiration pneumonia | 7411 | 7411 | 0 | 5928 | 742 | 741 | 0.27 | 0.95 | 0.94 | 703 | 0 | 0 | 39 | 0.95 | 1.00 | 0.97 | 0.21 | 0.95 | 1.00 |
| model_135 | Asthenia | 23515 | 23515 | 0 | 18812 | 2352 | 2351 | 0.37 | 0.96 | 0.96 | 2249 | 0 | 0 | 103 | 0.96 | 1.00 | 0.98 | 0.18 | 0.96 | 1.00 |
| model_136 | Asthenopia | 661 | 661 | 0 | 528 | 67 | 66 | 0.21 | 0.96 | 0.95 | 64 | 0 | 0 | 3 | 0.96 | 1.00 | 0.98 | 0.18 | 0.96 | 1.00 |
| model_137 | Asthma | 11142 | 11142 | 0 | 8913 | 1115 | 1114 | 0.28 | 0.97 | 0.96 | 1076 | 0 | 0 | 39 | 0.97 | 1.00 | 0.98 | 0.14 | 0.97 | 1.00 |
| model_138 | Astrocytoma | 113 | 588 | 475 | 470 | 59 | 59 | 0.25 | 0.76 | 0.8 | 45 | 0 | 0 | 14 | 0.76 | 1.00 | 0.87 | 0.95 | 0.76 | 1.00 |
| model_139 | Asystole | 12501 | 12501 | 0 | 10000 | 1251 | 1250 | 0.32 | 0.93 | 0.94 | 1165 | 0 | 0 | 86 | 0.93 | 1.00 | 0.96 | 0.27 | 0.93 | 1.00 |
| model_140 | Ataxia | 3675 | 3675 | 0 | 2940 | 368 | 367 | 0.25 | 0.94 | 0.95 | 346 | 0 | 0 | 22 | 0.94 | 1.00 | 0.97 | 0.24 | 0.94 | 1.00 |
| model_141 | Atelectasis | 12062 | 12062 | 0 | 9649 | 1207 | 1206 | 0.31 | 0.96 | 0.94 | 1153 | 0 | 0 | 54 | 0.96 | 1.00 | 0.98 | 0.18 | 0.96 | 1.00 |
| model_142 | Atherosclerosis | 3370 | 3370 | 0 | 2696 | 337 | 337 | 0.24 | 0.96 | 0.95 | 324 | 0 | 0 | 13 | 0.96 | 1.00 | 0.98 | 0.15 | 0.96 | 1.00 |
| model_143 | Atrial ectopic beats | 3586 | 3586 | 0 | 2868 | 359 | 359 | 0.24 | 0.96 | 0.96 | 345 | 0 | 0 | 14 | 0.96 | 1.00 | 0.98 | 0.16 | 0.96 | 1.00 |

| | | | | | | | | | | | | | | | | | | | | |
|---|---|---|---|---|---|---|---|---|---|---|---|---|---|---|---|---|---|---|---|---|
| model_144 | Atrial flutter | 4269 | 4269 | 0 | 3415 | 427 | 427 | 0.24 | 0.93 | 0.91 | 398 | 0 | 0 | 29 | 0.93 | 1.00 | 0.96 | 0.27 | 0.93 | 1.00 |
| model_145 | Atrial septal defect | 1387 | 1387 | 0 | 1109 | 139 | 139 | 0.23 | 0.96 | 0.96 | 133 | 0 | 0 | 6 | 0.96 | 1.00 | 0.98 | 0.17 | 0.96 | 1.00 |
| model_146 | Atrioventricular block | 3257 | 3257 | 0 | 2605 | 326 | 326 | 0.24 | 0.93 | 0.95 | 303 | 0 | 0 | 23 | 0.93 | 1.00 | 0.96 | 0.28 | 0.93 | 1.00 |
| model_147 | Atrioventricular block complete | 3087 | 3087 | 0 | 2469 | 309 | 309 | 0.26 | 0.93 | 0.9 | 287 | 0 | 0 | 22 | 0.93 | 1.00 | 0.96 | 0.28 | 0.93 | 1.00 |
| model_148 | Atrioventricular block first degree | 4288 | 4288 | 0 | 3430 | 429 | 429 | 0.24 | 0.97 | 0.96 | 417 | 0 | 0 | 12 | 0.97 | 1.00 | 0.99 | 0.11 | 0.97 | 1.00 |
| model_149 | Atrioventricular block second degree | 1726 | 1726 | 0 | 1380 | 173 | 173 | 0.24 | 0.91 | 0.92 | 157 | 0 | 0 | 16 | 0.91 | 1.00 | 0.95 | 0.37 | 0.91 | 1.00 |
| model_150 | Atrophy of skin | 725 | 725 | 0 | 580 | 73 | 72 | 0.23 | 0.92 | 0.99 | 67 | 0 | 0 | 6 | 0.92 | 1.00 | 0.96 | 0.33 | 0.92 | 1.00 |
| model_151 | Attempted suicide | 5879 | 5879 | 0 | 4703 | 588 | 588 | 0.25 | 0.96 | 0.97 | 565 | 0 | 0 | 23 | 0.96 | 1.00 | 0.98 | 0.16 | 0.96 | 1.00 |
| model_152 | Atypical mycobacterial infection | 510 | 588 | 78 | 470 | 59 | 59 | 0.22 | 0.9 | 0.93 | 53 | 0 | 0 | 6 | 0.9 | 1.00 | 0.95 | 0.41 | 0.9 | 1.00 |
| model_153 | Autonomic instability | 1487 | 1487 | 0 | 1189 | 149 | 149 | 0.24 | 0.95 | 0.93 | 141 | 0 | 0 | 8 | 0.95 | 1.00 | 0.97 | 0.21 | 0.95 | 1.00 |
| model_154 | Autonomic neuropathy | 750 | 750 | 0 | 600 | 75 | 75 | 0.23 | 0.95 | 0.95 | 71 | 0 | 0 | 4 | 0.95 | 1.00 | 0.97 | 0.21 | 0.95 | 1.00 |
| model_155 | Avascular necrosis | 498 | 588 | 90 | 470 | 59 | 59 | 0.23 | 0.88 | 0.88 | 52 | 0 | 0 | 7 | 0.88 | 1.00 | 0.94 | 0.47 | 0.88 | 1.00 |
| model_156 | Azotaemia | 3924 | 3924 | 0 | 3139 | 393 | 392 | 0.24 | 0.95 | 0.93 | 372 | 0 | 0 | 21 | 0.95 | 1.00 | 0.97 | 0.21 | 0.95 | 1.00 |
| model_157 | Back Ache | 19930 | 19930 | 0 | 15944 | 1993 | 1993 | 0.33 | 0.95 | 0.95 | 1899 | 0 | 0 | 94 | 0.95 | 1.00 | 0.98 | 0.19 | 0.95 | 1.00 |
| model_158 | Back injury | 3662 | 3662 | 0 | 2929 | 367 | 366 | 0.25 | 0.95 | 0.95 | 350 | 0 | 0 | 17 | 0.95 | 1.00 | 0.98 | 0.19 | 0.95 | 1.00 |
| model_159 | Bacteraemia | 6816 | 6816 | 0 | 5452 | 682 | 682 | 0.26 | 0.93 | 0.93 | 635 | 0 | 0 | 47 | 0.93 | 1.00 | 0.96 | 0.28 | 0.93 | 1.00 |
| model_160 | Bacterial conjunctivitis | 97 | 588 | 491 | 470 | 59 | 59 | 0.24 | 0.86 | 0.85 | 51 | 0 | 0 | 8 | 0.86 | 1.00 | 0.93 | 0.54 | 0.86 | 1.00 |
| model_161 | Bacterial endocarditis | 310 | 588 | 278 | 470 | 59 | 59 | 0.23 | 0.9 | 0.88 | 53 | 0 | 0 | 6 | 0.9 | 1.00 | 0.95 | 0.41 | 0.9 | 1.00 |
| model_162 | Bacterial infection | 8928 | 8928 | 0 | 7142 | 893 | 893 | 0.27 | 0.94 | 0.95 | 843 | 0 | 0 | 50 | 0.94 | 1.00 | 0.97 | 0.22 | 0.94 | 1.00 |
| model_163 | Bacterial pneumonia | 3323 | 3323 | 0 | 2658 | 333 | 332 | 0.24 | 0.96 | 0.95 | 319 | 0 | 0 | 14 | 0.96 | 1.00 | 0.98 | 0.17 | 0.96 | 1.00 |
| model_164 | Bacterial vaginitis | 526 | 588 | 62 | 470 | 59 | 59 | 0.23 | 0.86 | 0.9 | 51 | 0 | 0 | 8 | 0.86 | 1.00 | 0.93 | 0.54 | 0.86 | 1.00 |
| model_165 | Bad breath | 1476 | 1476 | 0 | 1180 | 148 | 148 | 0.23 | 0.98 | 0.94 | 145 | 0 | 0 | 3 | 0.98 | 1.00 | 0.99 | 0.08 | 0.98 | 1.00 |
| model_166 | Balance disorder | 9907 | 9907 | 0 | 7925 | 991 | 991 | 0.29 | 0.96 | 0.96 | 949 | 0 | 0 | 42 | 0.96 | 1.00 | 0.98 | 0.17 | 0.96 | 1.00 |
| model_167 | Balanitis | 516 | 588 | 72 | 470 | 59 | 59 | 0.22 | 0.92 | 0.92 | 54 | 0 | 0 | 5 | 0.92 | 1.00 | 0.96 | 0.34 | 0.92 | 1.00 |
| model_168 | Basal cell carcinoma | 2889 | 2889 | 0 | 2311 | 289 | 289 | 0.25 | 0.92 | 0.94 | 267 | 0 | 0 | 22 | 0.92 | 1.00 | 0.96 | 0.3 | 0.92 | 1.00 |
| model_169 | Belching | 3470 | 3470 | 0 | 2776 | 347 | 347 | 0.24 | 0.97 | 0.95 | 337 | 0 | 0 | 10 | 0.97 | 1.00 | 0.99 | 0.12 | 0.97 | 1.00 |
| model_170 | Benign Neoplasm | 339 | 588 | 249 | 470 | 59 | 59 | 0.24 | 0.93 | 0.95 | 55 | 0 | 0 | 4 | 0.93 | 1.00 | 0.96 | 0.27 | 0.93 | 1.00 |
| model_171 | Benign intracranial hypertension | 634 | 634 | 0 | 507 | 64 | 63 | 0.21 | 0.98 | 0.98 | 63 | 0 | 0 | 1 | 0.98 | 1.00 | 0.99 | 0.06 | 0.98 | 1.00 |
| model_172 | Benign prostatic hyperplasia | 4727 | 4727 | 0 | 3781 | 473 | 473 | 0.25 | 0.98 | 0.96 | 462 | 0 | 0 | 11 | 0.98 | 1.00 | 0.99 | 0.09 | 0.98 | 1.00 |
| model_173 | Bicuspid Aortic Valve | 155 | 588 | 433 | 470 | 59 | 59 | 0.23 | 0.76 | 0.83 | 45 | 0 | 0 | 14 | 0.76 | 1.00 | 0.87 | 0.95 | 0.76 | 1.00 |
| model_174 | Biliary cirrhosis | 261 | 588 | 327 | 470 | 59 | 59 | 0.22 | 0.93 | 0.93 | 55 | 0 | 0 | 4 | 0.93 | 1.00 | 0.96 | 0.27 | 0.93 | 1.00 |
| model_175 | Biliary tract disorder | 2054 | 2054 | 0 | 1643 | 206 | 205 | 0.29 | 0.93 | 0.95 | 192 | 0 | 0 | 14 | 0.93 | 1.00 | 0.96 | 0.27 | 0.93 | 1.00 |
| model_176 | Bilirubinaemia | 7441 | 7441 | 0 | 5952 | 745 | 744 | 0.26 | 0.94 | 0.96 | 699 | 0 | 0 | 46 | 0.94 | 1.00 | 0.97 | 0.25 | 0.94 | 1.00 |
| model_177 | Bipolar disorder | 5199 | 5199 | 0 | 4159 | 520 | 520 | 0.25 | 0.97 | 0.97 | 505 | 0 | 0 | 15 | 0.97 | 1.00 | 0.99 | 0.12 | 0.97 | 1.00 |
| model_178 | Birth defect | 2862 | 2862 | 0 | 2289 | 287 | 286 | 0.25 | 0.96 | 0.94 | 276 | 0 | 0 | 11 | 0.96 | 1.00 | 0.98 | 0.15 | 0.96 | 1.00 |
| model_179 | Black stools | 5842 | 5842 | 0 | 4673 | 585 | 584 | 0.27 | 0.93 | 0.94 | 543 | 0 | 0 | 42 | 0.93 | 1.00 | 0.96 | 0.29 | 0.93 | 1.00 |
| model_180 | Bladder Neoplasm | 390 | 588 | 198 | 470 | 59 | 59 | 0.22 | 0.92 | 0.93 | 54 | 0 | 0 | 5 | 0.92 | 1.00 | 0.96 | 0.34 | 0.92 | 1.00 |
| model_181 | Bladder atony | 198 | 588 | 390 | 470 | 59 | 59 | 0.23 | 0.88 | 0.88 | 52 | 0 | 0 | 7 | 0.88 | 1.00 | 0.94 | 0.47 | 0.88 | 1.00 |
| model_182 | Bladder cancer | 1612 | 1612 | 0 | 1289 | 162 | 161 | 0.23 | 0.94 | 0.95 | 153 | 0 | 0 | 9 | 0.94 | 1.00 | 0.97 | 0.22 | 0.94 | 1.00 |
| model_183 | Bladder diverticulum | 27 | 588 | 561 | 470 | 59 | 59 | 0.24 | 0.27 | 0.25 | 16 | 0 | 0 | 43 | 0.27 | 1.00 | 0.43 | 2.92 | 0.27 | 1.00 |
| model_184 | Bladder inflammation | 11028 | 11028 | 0 | 8822 | 1103 | 1103 | 0.29 | 0.94 | 0.95 | 1035 | 0 | 0 | 68 | 0.94 | 1.00 | 0.97 | 0.25 | 0.94 | 1.00 |
| model_185 | Bladder retention | 7710 | 7710 | 0 | 6168 | 771 | 771 | 0.28 | 0.95 | 0.95 | 729 | 0 | 0 | 42 | 0.95 | 1.00 | 0.97 | 0.22 | 0.95 | 1.00 |
| model_186 | Bleb | 7119 | 7119 | 0 | 5695 | 712 | 712 | 0.26 | 0.96 | 0.94 | 680 | 0 | 0 | 32 | 0.96 | 1.00 | 0.98 | 0.18 | 0.96 | 1.00 |
| model_187 | Bleeding | 14143 | 14143 | 0 | 11314 | 1415 | 1414 | 0.31 | 0.94 | 0.95 | 1332 | 0 | 0 | 83 | 0.94 | 1.00 | 0.97 | 0.23 | 0.94 | 1.00 |
| model_188 | Bleeding Vaginal | 3355 | 3355 | 0 | 2684 | 336 | 335 | 0.24 | 0.96 | 0.95 | 322 | 0 | 0 | 14 | 0.96 | 1.00 | 0.98 | 0.17 | 0.96 | 1.00 |
| model_189 | Bleeding gums | 3764 | 3764 | 0 | 3011 | 377 | 376 | 0.25 | 0.95 | 0.97 | 357 | 0 | 0 | 20 | 0.95 | 1.00 | 0.97 | 0.21 | 0.95 | 1.00 |
| model_190 | Bleeding hemorrhoids | 2145 | 2145 | 0 | 1716 | 215 | 214 | 0.23 | 0.94 | 0.94 | 202 | 0 | 0 | 13 | 0.94 | 1.00 | 0.97 | 0.24 | 0.94 | 1.00 |
| model_191 | Blepharitis | 1176 | 1176 | 0 | 940 | 118 | 118 | 0.24 | 0.94 | 0.97 | 111 | 0 | 0 | 7 | 0.94 | 1.00 | 0.97 | 0.24 | 0.94 | 1.00 |
| model_192 | Blepharoptosis | 2062 | 2062 | 0 | 1649 | 207 | 206 | 0.23 | 0.93 | 0.92 | 193 | 0 | 0 | 14 | 0.93 | 1.00 | 0.97 | 0.27 | 0.93 | 1.00 |
| model_193 | Blepharospasm | 5855 | 5855 | 0 | 4684 | 586 | 585 | 0.27 | 0.95 | 0.94 | 559 | 0 | 0 | 27 | 0.95 | 1.00 | 0.98 | 0.18 | 0.95 | 1.00 |

| | | | | | | | | | | | | | | | | | | | | |
|---|---|---|---|---|---|---|---|---|---|---|---|---|---|---|---|---|---|---|---|---|
| model_194 | Blindness | 6237 | 6237 | 0 | 4989 | 624 | 624 | 0.25 | 0.96 | 0.94 | 596 | 0 | 0 | 28 | 0.96 | 1.00 | 0.98 | 0.18 | 0.96 | 1.00 |
| model_195 | Blood Calcium Increased | 6050 | 6050 | 0 | 4840 | 605 | 605 | 0.26 | 0.95 | 0.96 | 576 | 0 | 0 | 29 | 0.95 | 1.00 | 0.98 | 0.19 | 0.95 | 1.00 |
| model_196 | Blood calcium decreased | 10633 | 10633 | 0 | 8506 | 1064 | 1063 | 0.29 | 0.95 | 0.95 | 1012 | 0 | 0 | 52 | 0.95 | 1.00 | 0.97 | 0.2 | 0.95 | 1.00 |
| model_197 | Blood disorder | 3265 | 3265 | 0 | 2612 | 327 | 326 | 0.24 | 0.94 | 0.95 | 309 | 0 | 0 | 18 | 0.94 | 1.00 | 0.97 | 0.22 | 0.94 | 1.00 |
| model_198 | Blood in urine | 9201 | 9201 | 0 | 7360 | 921 | 920 | 0.3 | 0.96 | 0.95 | 883 | 0 | 0 | 38 | 0.96 | 1.00 | 0.98 | 0.17 | 0.96 | 1.00 |
| model_199 | Blood pressure abnormal | 2510 | 2510 | 0 | 2008 | 251 | 251 | 0.25 | 0.92 | 0.95 | 230 | 0 | 0 | 21 | 0.92 | 1.00 | 0.96 | 0.33 | 0.92 | 1.00 |
| model_200 | Blood sodium decreased | 14665 | 14665 | 0 | 11732 | 1467 | 1466 | 0.32 | 0.96 | 0.95 | 1401 | 0 | 0 | 66 | 0.96 | 1.00 | 0.98 | 0.18 | 0.96 | 1.00 |
| model_201 | Blurred vision | 13404 | 13404 | 0 | 10723 | 1341 | 1340 | 0.32 | 0.96 | 0.96 | 1283 | 0 | 0 | 58 | 0.96 | 1.00 | 0.98 | 0.17 | 0.96 | 1.00 |
| model_202 | Body temperature increased | 21806 | 21806 | 0 | 17444 | 2181 | 2181 | 0.38 | 0.95 | 0.95 | 2069 | 0 | 0 | 112 | 0.95 | 1.00 | 0.97 | 0.21 | 0.95 | 1.00 |
| model_203 | Body tinea | 318 | 588 | 270 | 470 | 59 | 59 | 0.26 | 0.93 | 0.97 | 55 | 0 | 0 | 4 | 0.93 | 1.00 | 0.96 | 0.27 | 0.93 | 1.00 |
| model_204 | Bone Fracture Spontaneous | 2385 | 2385 | 0 | 1908 | 239 | 238 | 0.25 | 0.93 | 0.93 | 222 | 0 | 0 | 17 | 0.93 | 1.00 | 0.96 | 0.28 | 0.93 | 1.00 |
| model_205 | Bone fracture | 2970 | 2970 | 0 | 2376 | 297 | 297 | 0.25 | 0.97 | 0.98 | 289 | 0 | 0 | 8 | 0.97 | 1.00 | 0.99 | 0.11 | 0.97 | 1.00 |
| model_206 | Bone inflammation | 1461 | 1461 | 0 | 1168 | 147 | 146 | 0.23 | 0.95 | 0.96 | 139 | 0 | 0 | 8 | 0.95 | 1.00 | 0.97 | 0.22 | 0.95 | 1.00 |
| model_207 | Bone marrow failure | 12525 | 12525 | 0 | 10020 | 1253 | 1252 | 0.31 | 0.94 | 0.93 | 1180 | 0 | 0 | 73 | 0.94 | 1.00 | 0.97 | 0.23 | 0.94 | 1.00 |
| model_208 | Bone marrow fibrosis | 298 | 588 | 290 | 470 | 59 | 59 | 0.23 | 0.93 | 0.85 | 55 | 0 | 0 | 4 | 0.93 | 1.00 | 0.96 | 0.27 | 0.93 | 1.00 |
| model_209 | Bone marrow transplant | 2125 | 2125 | 0 | 1700 | 213 | 212 | 0.24 | 0.91 | 0.91 | 193 | 0 | 0 | 20 | 0.91 | 1.00 | 0.95 | 0.38 | 0.91 | 1.00 |
| model_210 | Bone neoplasm | 427 | 588 | 161 | 470 | 59 | 59 | 0.23 | 0.93 | 0.95 | 55 | 0 | 0 | 4 | 0.93 | 1.00 | 0.96 | 0.27 | 0.93 | 1.00 |
| model_211 | Bone pain | 8398 | 8398 | 0 | 6718 | 840 | 840 | 0.28 | 0.95 | 0.96 | 796 | 0 | 0 | 44 | 0.95 | 1.00 | 0.97 | 0.21 | 0.95 | 1.00 |
| model_212 | Bone sarcoma | 273 | 588 | 315 | 470 | 59 | 59 | 0.22 | 0.9 | 0.92 | 53 | 0 | 0 | 6 | 0.9 | 1.00 | 0.95 | 0.41 | 0.9 | 1.00 |
| model_213 | Bone spur | 4031 | 4031 | 0 | 3224 | 404 | 403 | 0.24 | 0.95 | 0.96 | 385 | 0 | 0 | 19 | 0.95 | 1.00 | 0.98 | 0.19 | 0.95 | 1.00 |
| model_214 | Borderline personality disorder | 540 | 588 | 48 | 470 | 59 | 59 | 0.23 | 0.93 | 0.97 | 55 | 0 | 0 | 4 | 0.93 | 1.00 | 0.96 | 0.27 | 0.93 | 1.00 |
| model_215 | Brachial neuritis | 138 | 588 | 450 | 470 | 59 | 59 | 0.23 | 0.75 | 0.66 | 44 | 0 | 0 | 15 | 0.75 | 1.00 | 0.85 | 1.02 | 0.75 | 1.00 |
| model_216 | Brachial plexus injury | 89 | 588 | 499 | 470 | 59 | 59 | 0.26 | 0.85 | 0.75 | 50 | 0 | 0 | 9 | 0.85 | 1.00 | 0.92 | 0.61 | 0.85 | 1.00 |
| model_217 | Bradyarrhythmia | 1066 | 1066 | 0 | 852 | 107 | 107 | 0.22 | 0.97 | 0.97 | 104 | 0 | 0 | 3 | 0.97 | 1.00 | 0.99 | 0.11 | 0.97 | 1.00 |
| model_218 | Bradycardia | 12141 | 12141 | 0 | 9712 | 1215 | 1214 | 0.3 | 0.95 | 0.95 | 1153 | 0 | 0 | 62 | 0.95 | 1.00 | 0.97 | 0.2 | 0.95 | 1.00 |
| model_219 | Brain abscess | 821 | 821 | 0 | 656 | 83 | 82 | 0.23 | 0.9 | 0.9 | 75 | 0 | 0 | 8 | 0.9 | 1.00 | 0.95 | 0.39 | 0.9 | 1.00 |
| model_220 | Brain concussion | 1607 | 1607 | 0 | 1285 | 161 | 161 | 0.22 | 0.97 | 0.96 | 156 | 0 | 0 | 5 | 0.97 | 1.00 | 0.98 | 0.12 | 0.97 | 1.00 |
| model_221 | Brain neoplasm | 1468 | 1468 | 0 | 1174 | 147 | 147 | 0.23 | 0.89 | 0.97 | 131 | 0 | 0 | 16 | 0.89 | 1.00 | 0.94 | 0.44 | 0.89 | 1.00 |
| model_222 | Breakthrough bleeding | 2686 | 2686 | 0 | 2148 | 269 | 269 | 0.25 | 0.96 | 0.93 | 258 | 0 | 0 | 11 | 0.96 | 1.00 | 0.98 | 0.16 | 0.96 | 1.00 |
| model_223 | Breast Lump | 2977 | 2977 | 0 | 2381 | 298 | 298 | 0.24 | 0.95 | 0.97 | 283 | 0 | 0 | 15 | 0.95 | 1.00 | 0.97 | 0.2 | 0.95 | 1.00 |
| model_224 | Breast abscess | 141 | 588 | 447 | 470 | 59 | 59 | 0.24 | 0.98 | 0.88 | 58 | 0 | 0 | 1 | 0.98 | 1.00 | 0.99 | 0.07 | 0.98 | 1.00 |
| model_225 | Breast cancer | 4736 | 4736 | 0 | 3788 | 474 | 474 | 0.26 | 0.95 | 0.96 | 449 | 0 | 0 | 25 | 0.95 | 1.00 | 0.97 | 0.21 | 0.95 | 1.00 |
| model_226 | Breast cyst | 1013 | 1013 | 0 | 810 | 102 | 101 | 0.22 | 0.93 | 0.98 | 95 | 0 | 0 | 7 | 0.93 | 1.00 | 0.96 | 0.27 | 0.93 | 1.00 |
| model_227 | Breast discharge | 574 | 588 | 14 | 470 | 59 | 59 | 0.23 | 0.97 | 0.92 | 57 | 0 | 0 | 2 | 0.97 | 1.00 | 0.98 | 0.14 | 0.97 | 1.00 |
| model_228 | Breast disorder | 1021 | 1021 | 0 | 816 | 103 | 102 | 0.23 | 0.91 | 0.95 | 94 | 0 | 0 | 9 | 0.91 | 1.00 | 0.95 | 0.35 | 0.91 | 1.00 |
| model_229 | Breast dysplasia | 1454 | 1454 | 0 | 1163 | 146 | 145 | 0.24 | 0.96 | 0.94 | 140 | 0 | 0 | 6 | 0.96 | 1.00 | 0.98 | 0.16 | 0.96 | 1.00 |
| model_230 | Breast enlargement | 987 | 987 | 0 | 789 | 99 | 99 | 0.22 | 0.94 | 0.97 | 93 | 0 | 0 | 6 | 0.94 | 1.00 | 0.97 | 0.24 | 0.94 | 1.00 |
| model_231 | Breast inflammation | 862 | 862 | 0 | 689 | 87 | 86 | 0.23 | 0.95 | 0.93 | 83 | 0 | 0 | 4 | 0.95 | 1.00 | 0.98 | 0.18 | 0.95 | 1.00 |
| model_232 | Breast malformation | 14 | 588 | 574 | 470 | 59 | 59 | 0.25 | 0.02 | 0.03 | 1 | 0 | 0 | 58 | 0.02 | 1.00 | 0.03 | 3.93 | 0.02 | 1.00 |
| model_233 | Breast neoplasm | 366 | 588 | 222 | 470 | 59 | 59 | 0.23 | 0.95 | 0.98 | 56 | 0 | 0 | 3 | 0.95 | 1.00 | 0.97 | 0.2 | 0.95 | 1.00 |
| model_234 | Breast pain | 2804 | 2804 | 0 | 2243 | 281 | 280 | 0.24 | 0.98 | 0.97 | 274 | 0 | 0 | 7 | 0.98 | 1.00 | 0.99 | 0.1 | 0.98 | 1.00 |
| model_235 | Breast swelling | 646 | 646 | 0 | 516 | 65 | 65 | 0.22 | 0.97 | 0.97 | 63 | 0 | 0 | 2 | 0.97 | 1.00 | 0.98 | 0.12 | 0.97 | 1.00 |
| model_236 | Breast tenderness | 1178 | 1178 | 0 | 942 | 118 | 118 | 0.22 | 0.92 | 0.94 | 108 | 0 | 0 | 10 | 0.92 | 1.00 | 0.96 | 0.34 | 0.92 | 1.00 |
| model_237 | Bronchial spasm | 5743 | 5743 | 0 | 4594 | 575 | 574 | 0.26 | 0.96 | 0.95 | 554 | 0 | 0 | 21 | 0.96 | 1.00 | 0.98 | 0.15 | 0.96 | 1.00 |
| model_238 | Bronchiectasis | 1979 | 1979 | 0 | 1583 | 198 | 198 | 0.23 | 0.95 | 0.96 | 189 | 0 | 0 | 9 | 0.95 | 1.00 | 0.98 | 0.18 | 0.95 | 1.00 |
| model_239 | Bronchiolitis | 636 | 636 | 0 | 508 | 64 | 64 | 0.22 | 0.97 | 0.97 | 62 | 0 | 0 | 2 | 0.97 | 1.00 | 0.98 | 0.13 | 0.97 | 1.00 |
| model_240 | Bronchitis | 13602 | 13602 | 0 | 10881 | 1361 | 1360 | 0.3 | 0.96 | 0.96 | 1311 | 0 | 0 | 50 | 0.96 | 1.00 | 0.98 | 0.15 | 0.96 | 1.00 |
| model_241 | Bruise | 10902 | 10902 | 0 | 8721 | 1091 | 1090 | 0.29 | 0.95 | 0.96 | 1040 | 0 | 0 | 51 | 0.95 | 1.00 | 0.98 | 0.19 | 0.95 | 1.00 |
| model_242 | Bruxism | 10666 | 10666 | 0 | 8532 | 1067 | 1067 | 0.28 | 0.96 | 0.97 | 1021 | 0 | 0 | 46 | 0.96 | 1.00 | 0.98 | 0.17 | 0.96 | 1.00 |
| model_243 | Bulging | 10677 | 10677 | 0 | 8541 | 1068 | 1068 | 0.29 | 0.95 | 0.96 | 1017 | 0 | 0 | 51 | 0.95 | 1.00 | 0.98 | 0.19 | 0.95 | 1.00 |

| | | | | | | | | | | | | | | | | | | | | |
|---|---|---|---|---|---|---|---|---|---|---|---|---|---|---|---|---|---|---|---|---|
| model_244 | Bulimia | 381 | 588 | 207 | 470 | 59 | 59 | 0.23 | 0.92 | 0.95 | 54 | 0 | 0 | 5 | 0.92 | 1.00 | 0.96 | 0.34 | 0.92 | 1.00 |
| model_245 | Bundle branch block | 1507 | 1507 | 0 | 1205 | 151 | 151 | 0.23 | 0.96 | 0.91 | 145 | 0 | 0 | 6 | 0.96 | 1.00 | 0.98 | 0.16 | 0.96 | 1.00 |
| model_246 | Bundle branch block left | 4414 | 4414 | 0 | 3531 | 442 | 441 | 0.25 | 0.95 | 0.97 | 420 | 0 | 0 | 22 | 0.95 | 1.00 | 0.97 | 0.2 | 0.95 | 1.00 |
| model_247 | Bundle branch block right | 5558 | 5558 | 0 | 4446 | 556 | 556 | 0.26 | 0.96 | 0.94 | 531 | 0 | 0 | 25 | 0.96 | 1.00 | 0.98 | 0.18 | 0.96 | 1.00 |
| model_248 | Bunion | 1030 | 1030 | 0 | 824 | 103 | 103 | 0.22 | 0.97 | 0.93 | 100 | 0 | 0 | 3 | 0.97 | 1.00 | 0.99 | 0.12 | 0.97 | 1.00 |
| model_249 | Burning sensation | 8154 | 8154 | 0 | 6523 | 816 | 815 | 0.27 | 0.95 | 0.97 | 778 | 0 | 0 | 38 | 0.95 | 1.00 | 0.98 | 0.19 | 0.95 | 1.00 |
| model_250 | Burns Second Degree | 766 | 766 | 0 | 612 | 77 | 77 | 0.23 | 0.9 | 0.86 | 69 | 0 | 0 | 8 | 0.9 | 1.00 | 0.95 | 0.42 | 0.9 | 1.00 |
| model_251 | Bursitis | 6091 | 6091 | 0 | 4872 | 610 | 609 | 0.26 | 0.97 | 0.98 | 594 | 0 | 0 | 16 | 0.97 | 1.00 | 0.99 | 0.1 | 0.97 | 1.00 |
| model_252 | CMV infection | 3303 | 3303 | 0 | 2642 | 331 | 330 | 0.24 | 0.94 | 0.94 | 310 | 0 | 0 | 21 | 0.94 | 1.00 | 0.97 | 0.25 | 0.94 | 1.00 |
| model_253 | CYSTO | 36 | 588 | 552 | 470 | 59 | 59 | 0.26 | 0.25 | 0.19 | 15 | 0 | 0 | 44 | 0.25 | 1.00 | 0.41 | 2.98 | 0.25 | 1.00 |
| model_254 | Cachectic | 2848 | 2848 | 0 | 2278 | 285 | 285 | 0.25 | 0.93 | 0.95 | 265 | 0 | 0 | 20 | 0.93 | 1.00 | 0.96 | 0.28 | 0.93 | 1.00 |
| model_255 | Caesarean Section | 1587 | 1587 | 0 | 1269 | 159 | 159 | 0.23 | 0.92 | 0.93 | 147 | 0 | 0 | 12 | 0.92 | 1.00 | 0.96 | 0.3 | 0.92 | 1.00 |
| model_256 | Calculus ureteric | 1172 | 1172 | 0 | 937 | 118 | 117 | 0.23 | 0.92 | 0.93 | 109 | 0 | 0 | 9 | 0.92 | 1.00 | 0.96 | 0.31 | 0.92 | 1.00 |
| model_257 | Cancer | 3321 | 3321 | 0 | 2656 | 333 | 332 | 0.25 | 0.98 | 0.97 | 325 | 0 | 0 | 8 | 0.98 | 1.00 | 0.99 | 0.1 | 0.98 | 1.00 |
| model_258 | Candida Infection | 9808 | 9808 | 0 | 7846 | 981 | 981 | 0.27 | 0.94 | 0.94 | 922 | 0 | 0 | 59 | 0.94 | 1.00 | 0.97 | 0.24 | 0.94 | 1.00 |
| model_259 | Candidiasis of mouth | 6998 | 6998 | 0 | 5598 | 700 | 700 | 0.27 | 0.95 | 0.95 | 667 | 0 | 0 | 33 | 0.95 | 1.00 | 0.98 | 0.19 | 0.95 | 1.00 |
| model_260 | Candidiasis of vagina | 1553 | 1553 | 0 | 1242 | 156 | 155 | 0.23 | 0.92 | 0.95 | 144 | 0 | 0 | 12 | 0.92 | 1.00 | 0.96 | 0.31 | 0.92 | 1.00 |
| model_261 | Carbuncle | 509 | 588 | 79 | 470 | 59 | 59 | 0.22 | 0.92 | 0.95 | 54 | 0 | 0 | 5 | 0.92 | 1.00 | 0.96 | 0.34 | 0.92 | 1.00 |
| model_262 | Carcinoid syndrome | 2511 | 2511 | 0 | 2008 | 252 | 251 | 0.24 | 0.9 | 0.91 | 228 | 0 | 0 | 24 | 0.9 | 1.00 | 0.95 | 0.38 | 0.9 | 1.00 |
| model_263 | Carcinoid tumour | 183 | 588 | 405 | 470 | 59 | 59 | 0.25 | 0.95 | 0.86 | 56 | 0 | 0 | 3 | 0.95 | 1.00 | 0.97 | 0.2 | 0.95 | 1.00 |
| model_264 | Carcinoma | 333 | 588 | 255 | 470 | 59 | 59 | 0.23 | 0.86 | 0.95 | 51 | 0 | 0 | 8 | 0.86 | 1.00 | 0.93 | 0.54 | 0.86 | 1.00 |
| model_265 | Carcinoma of Prostate | 3403 | 3403 | 0 | 2722 | 341 | 340 | 0.24 | 0.93 | 0.95 | 318 | 0 | 0 | 23 | 0.93 | 1.00 | 0.97 | 0.27 | 0.93 | 1.00 |
| model_266 | Carcinoma of the cervix | 656 | 656 | 0 | 524 | 66 | 66 | 0.22 | 0.95 | 0.94 | 63 | 0 | 0 | 3 | 0.95 | 1.00 | 0.98 | 0.18 | 0.95 | 1.00 |
| model_267 | Carcinoma of the colon | 1939 | 1939 | 0 | 1551 | 194 | 194 | 0.24 | 0.97 | 0.96 | 189 | 0 | 0 | 5 | 0.97 | 1.00 | 0.99 | 0.1 | 0.97 | 1.00 |
| model_268 | Cardiac decompensation | 16555 | 16555 | 0 | 13244 | 1656 | 1655 | 0.31 | 0.96 | 0.96 | 1596 | 0 | 0 | 60 | 0.96 | 1.00 | 0.98 | 0.14 | 0.96 | 1.00 |
| model_269 | Cardiac disease | 11412 | 11412 | 0 | 9129 | 1142 | 1141 | 0.28 | 0.95 | 0.95 | 1088 | 0 | 0 | 54 | 0.95 | 1.00 | 0.98 | 0.19 | 0.95 | 1.00 |
| model_270 | Cardiac enlargement | 10979 | 10979 | 0 | 8783 | 1098 | 1098 | 0.29 | 0.95 | 0.95 | 1039 | 0 | 0 | 59 | 0.95 | 1.00 | 0.97 | 0.21 | 0.95 | 1.00 |
| model_271 | Cardiac failure | 9377 | 9377 | 0 | 7501 | 938 | 938 | 0.28 | 0.94 | 0.94 | 886 | 0 | 0 | 52 | 0.94 | 1.00 | 0.97 | 0.22 | 0.94 | 1.00 |
| model_272 | Cardiac ischemia | 9133 | 9133 | 0 | 7306 | 914 | 913 | 0.27 | 0.95 | 0.95 | 869 | 0 | 0 | 45 | 0.95 | 1.00 | 0.97 | 0.2 | 0.95 | 1.00 |
| model_273 | Cardiac murmur | 6410 | 6410 | 0 | 5128 | 641 | 641 | 0.26 | 0.96 | 0.97 | 617 | 0 | 0 | 24 | 0.96 | 1.00 | 0.98 | 0.15 | 0.96 | 1.00 |
| model_274 | Cardiac tamponade | 1697 | 1697 | 0 | 1357 | 170 | 170 | 0.24 | 0.93 | 0.92 | 158 | 0 | 0 | 12 | 0.93 | 1.00 | 0.96 | 0.28 | 0.93 | 1.00 |
| model_275 | Cardiac valvulopathy | 4812 | 4812 | 0 | 3849 | 482 | 481 | 0.24 | 0.94 | 0.94 | 452 | 0 | 0 | 30 | 0.94 | 1.00 | 0.97 | 0.25 | 0.94 | 1.00 |
| model_276 | Cardiomyopathy | 7893 | 7893 | 0 | 6314 | 790 | 789 | 0.26 | 0.95 | 0.94 | 752 | 0 | 0 | 38 | 0.95 | 1.00 | 0.98 | 0.19 | 0.95 | 1.00 |
| model_277 | Cardiovascular collapse | 8681 | 8681 | 0 | 6944 | 869 | 868 | 0.27 | 0.93 | 0.94 | 805 | 0 | 0 | 64 | 0.93 | 1.00 | 0.96 | 0.29 | 0.93 | 1.00 |
| model_278 | Carotid artery stenosis | 5424 | 5424 | 0 | 4339 | 543 | 542 | 0.25 | 0.96 | 0.96 | 520 | 0 | 0 | 23 | 0.96 | 1.00 | 0.98 | 0.17 | 0.96 | 1.00 |
| model_279 | Carotid bruit | 1670 | 1670 | 0 | 1336 | 167 | 167 | 0.22 | 0.95 | 0.96 | 158 | 0 | 0 | 9 | 0.95 | 1.00 | 0.97 | 0.22 | 0.95 | 1.00 |
| model_280 | Carpal tunnel | 6215 | 6215 | 0 | 4972 | 622 | 621 | 0.26 | 0.95 | 0.96 | 592 | 0 | 0 | 30 | 0.95 | 1.00 | 0.98 | 0.19 | 0.95 | 1.00 |
| model_281 | Cataplexy | 107 | 588 | 481 | 470 | 59 | 59 | 0.24 | 0.86 | 0.8 | 51 | 0 | 0 | 8 | 0.86 | 1.00 | 0.93 | 0.54 | 0.86 | 1.00 |
| model_282 | Cataract | 7425 | 7425 | 0 | 5940 | 743 | 742 | 0.27 | 0.94 | 0.96 | 699 | 0 | 0 | 44 | 0.94 | 1.00 | 0.97 | 0.24 | 0.94 | 1.00 |
| model_283 | Causalgia | 925 | 925 | 0 | 740 | 93 | 92 | 0.23 | 1 | 0.97 | 93 | 0 | 0 | 0 | 1 | 1.00 | 1 | 0 | 1 | 1.00 |
| model_284 | Cavernous sinus thrombosis | 86 | 588 | 502 | 470 | 59 | 59 | 0.26 | 0.61 | 0.73 | 36 | 0 | 0 | 23 | 0.61 | 1.00 | 0.76 | 1.56 | 0.61 | 1.00 |
| model_285 | Cellulitis | 14377 | 14377 | 0 | 11501 | 1438 | 1438 | 0.32 | 0.95 | 0.95 | 1369 | 0 | 0 | 69 | 0.95 | 1.00 | 0.98 | 0.19 | 0.95 | 1.00 |
| model_286 | Cerebellar ataxia | 242 | 588 | 346 | 470 | 59 | 59 | 0.25 | 0.97 | 0.9 | 57 | 0 | 0 | 2 | 0.97 | 1.00 | 0.98 | 0.14 | 0.97 | 1.00 |
| model_287 | Cerebral artery embolism | 1236 | 1236 | 0 | 988 | 124 | 124 | 0.23 | 0.87 | 0.84 | 108 | 0 | 0 | 16 | 0.87 | 1.00 | 0.93 | 0.52 | 0.87 | 1.00 |
| model_288 | Cerebral atrophy | 4775 | 4775 | 0 | 3820 | 478 | 477 | 0.26 | 0.96 | 0.95 | 458 | 0 | 0 | 20 | 0.96 | 1.00 | 0.98 | 0.17 | 0.96 | 1.00 |
| model_289 | Cerebral haemorrhage | 5143 | 5143 | 0 | 4114 | 515 | 514 | 0.26 | 0.94 | 0.92 | 486 | 0 | 0 | 29 | 0.94 | 1.00 | 0.97 | 0.23 | 0.94 | 1.00 |
| model_290 | Cerebral infarct | 8223 | 8223 | 0 | 6578 | 823 | 822 | 0.26 | 0.94 | 0.93 | 776 | 0 | 0 | 47 | 0.94 | 1.00 | 0.97 | 0.23 | 0.94 | 1.00 |
| model_291 | Cerebral ischaemia | 4555 | 4555 | 0 | 3644 | 456 | 455 | 0.25 | 0.95 | 0.94 | 432 | 0 | 0 | 24 | 0.95 | 1.00 | 0.97 | 0.21 | 0.95 | 1.00 |
| model_292 | Cerebral palsy | 413 | 588 | 175 | 470 | 59 | 59 | 0.23 | 0.95 | 0.93 | 56 | 0 | 0 | 3 | 0.95 | 1.00 | 0.97 | 0.2 | 0.95 | 1.00 |
| model_293 | Cerebral thrombosis | 593 | 593 | 0 | 474 | 60 | 59 | 0.22 | 0.97 | 0.97 | 58 | 0 | 0 | 2 | 0.97 | 1.00 | 0.98 | 0.13 | 0.97 | 1.00 |

| | | | | | | | | | | | | | | | | | | | |
|---|---|---|---|---|---|---|---|---|---|---|---|---|---|---|---|---|---|---|---|
| model_294 | Cerebral vascular disorder | 2375 | 2375 | 0 | 1900 | 238 | 237 | 0.23 | 0.97 | 0.95 | 231 | 0 | 0 | 7 | 0.97 | 1.00 | 0.99 | 0.12 | 0.97 | 1.00 |
| model_295 | Cerumen impaction | 923 | 923 | 0 | 738 | 93 | 92 | 0.23 | 0.96 | 0.98 | 89 | 0 | 0 | 4 | 0.96 | 1.00 | 0.98 | 0.17 | 0.96 | 1.00 |
| model_296 | Cervical dysplasia | 821 | 821 | 0 | 656 | 83 | 82 | 0.22 | 0.93 | 0.95 | 77 | 0 | 0 | 6 | 0.93 | 1.00 | 0.96 | 0.29 | 0.93 | 1.00 |
| model_297 | Cervical polyp | 311 | 588 | 277 | 470 | 59 | 59 | 0.24 | 0.93 | 0.97 | 55 | 0 | 0 | 4 | 0.93 | 1.00 | 0.96 | 0.27 | 0.93 | 1.00 |
| model_298 | Cervical stenosis | 6 | 588 | 582 | 470 | 59 | 59 | 0.22 | 0 | 0 | 0 | 0 | 0 | 59 | 0 | 0.00 | 0 | 4 | 0 | 1.00 |
| model_299 | Cervical vertebral fracture | 557 | 588 | 31 | 470 | 59 | 59 | 0.23 | 0.95 | 0.98 | 56 | 0 | 0 | 3 | 0.95 | 1.00 | 0.97 | 0.2 | 0.95 | 1.00 |
| model_300 | Cervicalgia | 10213 | 10213 | 0 | 8170 | 1022 | 1021 | 0.28 | 0.96 | 0.96 | 980 | 0 | 0 | 42 | 0.96 | 1.00 | 0.98 | 0.16 | 0.96 | 1.00 |
| model_301 | Cervicitis | 918 | 918 | 0 | 734 | 92 | 92 | 0.23 | 0.99 | 0.97 | 91 | 0 | 0 | 1 | 0.99 | 1.00 | 0.99 | 0.04 | 0.99 | 1.00 |
| model_302 | Chalazion | 128 | 588 | 460 | 470 | 59 | 59 | 0.24 | 0.85 | 0.88 | 50 | 0 | 0 | 9 | 0.85 | 1.00 | 0.92 | 0.61 | 0.85 | 1.00 |
| model_303 | Cheilosis | 1523 | 1523 | 0 | 1218 | 153 | 152 | 0.24 | 0.92 | 0.96 | 140 | 0 | 0 | 13 | 0.92 | 1.00 | 0.96 | 0.34 | 0.92 | 1.00 |
| model_304 | Chest infection | 4067 | 4067 | 0 | 3253 | 407 | 407 | 0.25 | 0.96 | 0.95 | 390 | 0 | 0 | 17 | 0.96 | 1.00 | 0.98 | 0.17 | 0.96 | 1.00 |
| model_305 | Chest pain | 21013 | 21013 | 0 | 16810 | 2102 | 2101 | 0.35 | 0.96 | 0.96 | 2023 | 0 | 0 | 79 | 0.96 | 1.00 | 0.98 | 0.15 | 0.96 | 1.00 |
| model_306 | Chest wall pain | 2508 | 2508 | 0 | 2006 | 251 | 251 | 0.25 | 0.99 | 0.95 | 248 | 0 | 0 | 3 | 0.99 | 1.00 | 0.99 | 0.05 | 0.99 | 1.00 |
| model_307 | Chicken pox | 516 | 588 | 72 | 470 | 59 | 59 | 0.23 | 0.92 | 0.88 | 54 | 0 | 0 | 5 | 0.92 | 1.00 | 0.96 | 0.34 | 0.92 | 1.00 |
| model_308 | Chill | 13749 | 13749 | 0 | 10999 | 1375 | 1375 | 0.31 | 0.94 | 0.94 | 1296 | 0 | 0 | 79 | 0.94 | 1.00 | 0.97 | 0.23 | 0.94 | 1.00 |
| model_309 | Chloasma | 6 | 588 | 582 | 470 | 59 | 59 | 0.22 | 0 | 0 | 0 | 0 | 0 | 59 | 0 | 0.00 | 0 | 4 | 0 | 1.00 |
| model_310 | Choking | 2645 | 2645 | 0 | 2116 | 265 | 264 | 0.26 | 0.98 | 0.96 | 259 | 0 | 0 | 6 | 0.98 | 1.00 | 0.99 | 0.09 | 0.98 | 1.00 |
| model_311 | Cholangitis | 1828 | 1828 | 0 | 1462 | 183 | 183 | 0.23 | 0.96 | 0.93 | 175 | 0 | 0 | 8 | 0.96 | 1.00 | 0.98 | 0.17 | 0.96 | 1.00 |
| model_312 | Cholangitis sclerosing | 326 | 588 | 262 | 470 | 59 | 59 | 0.24 | 0.93 | 0.88 | 55 | 0 | 0 | 4 | 0.93 | 1.00 | 0.96 | 0.27 | 0.93 | 1.00 |
| model_313 | Cholecystectomies | 3887 | 3887 | 0 | 3109 | 389 | 389 | 0.27 | 0.95 | 0.97 | 368 | 0 | 0 | 21 | 0.95 | 1.00 | 0.97 | 0.22 | 0.95 | 1.00 |
| model_314 | Cholecystitis | 6573 | 6573 | 0 | 5258 | 658 | 657 | 0.25 | 0.96 | 0.96 | 633 | 0 | 0 | 25 | 0.96 | 1.00 | 0.98 | 0.15 | 0.96 | 1.00 |
| model_315 | Cholecystitis acute | 2076 | 2076 | 0 | 1660 | 208 | 208 | 0.24 | 0.96 | 0.97 | 200 | 0 | 0 | 8 | 0.96 | 1.00 | 0.98 | 0.15 | 0.96 | 1.00 |
| model_316 | Cholecystitis chronic | 2311 | 2311 | 0 | 1848 | 232 | 231 | 0.25 | 0.95 | 0.95 | 220 | 0 | 0 | 12 | 0.95 | 1.00 | 0.97 | 0.21 | 0.95 | 1.00 |
| model_317 | Cholelithiasis | 11027 | 11027 | 0 | 8821 | 1103 | 1103 | 0.29 | 0.96 | 0.96 | 1059 | 0 | 0 | 44 | 0.96 | 1.00 | 0.98 | 0.16 | 0.96 | 1.00 |
| model_318 | Chondrocalcinosis pyrophosphate | 401 | 588 | 187 | 470 | 59 | 59 | 0.23 | 0.85 | 0.93 | 50 | 0 | 0 | 9 | 0.85 | 1.00 | 0.92 | 0.61 | 0.85 | 1.00 |
| model_319 | Chondrodystrophy | 36 | 588 | 552 | 470 | 59 | 59 | 0.26 | 0.2 | 0.32 | 12 | 0 | 0 | 47 | 0.2 | 1.00 | 0.34 | 3.19 | 0.2 | 1.00 |
| model_320 | Choreoathetosis | 810 | 810 | 0 | 648 | 81 | 81 | 0.22 | 0.9 | 0.93 | 73 | 0 | 0 | 8 | 0.9 | 1.00 | 0.95 | 0.4 | 0.9 | 1.00 |
| model_321 | Choriocarcinoma | 18 | 588 | 570 | 470 | 59 | 59 | 0.24 | 0.07 | 0.05 | 4 | 0 | 0 | 55 | 0.07 | 1.00 | 0.13 | 3.73 | 0.07 | 1.00 |
| model_322 | Chorioretinitis | 25 | 588 | 563 | 470 | 59 | 59 | 0.26 | 0.34 | 0.37 | 20 | 0 | 0 | 39 | 0.34 | 1.00 | 0.51 | 2.64 | 0.34 | 1.00 |
| model_323 | Chronic Kidney Disease | 9143 | 9143 | 0 | 7314 | 915 | 914 | 0.29 | 0.96 | 0.94 | 881 | 0 | 0 | 34 | 0.96 | 1.00 | 0.98 | 0.15 | 0.96 | 1.00 |
| model_324 | Chronic active hepatitis | 294 | 588 | 294 | 470 | 59 | 59 | 0.25 | 0.93 | 0.97 | 55 | 0 | 0 | 4 | 0.93 | 1.00 | 0.96 | 0.27 | 0.93 | 1.00 |
| model_325 | Chronic hepatitis | 1145 | 1145 | 0 | 916 | 115 | 114 | 0.24 | 0.93 | 0.93 | 107 | 0 | 0 | 8 | 0.93 | 1.00 | 0.96 | 0.28 | 0.93 | 1.00 |
| model_326 | Chronic obstructive airway disease | 11605 | 11605 | 0 | 9284 | 1161 | 1160 | 0.28 | 0.95 | 0.96 | 1104 | 0 | 0 | 57 | 0.95 | 1.00 | 0.97 | 0.2 | 0.95 | 1.00 |
| model_327 | Chronic sinusitis | 2692 | 2692 | 0 | 2153 | 270 | 269 | 0.25 | 0.96 | 0.96 | 260 | 0 | 0 | 10 | 0.96 | 1.00 | 0.98 | 0.15 | 0.96 | 1.00 |
| model_328 | Chronic ulcerative colitis | 1984 | 1984 | 0 | 1587 | 199 | 198 | 0.24 | 0.95 | 0.94 | 189 | 0 | 0 | 10 | 0.95 | 1.00 | 0.97 | 0.2 | 0.95 | 1.00 |
| model_329 | Cirrhosis | 5337 | 5337 | 0 | 4269 | 534 | 534 | 0.27 | 0.96 | 0.95 | 510 | 0 | 0 | 24 | 0.96 | 1.00 | 0.98 | 0.18 | 0.96 | 1.00 |
| model_330 | Cleft lip | 96 | 588 | 492 | 470 | 59 | 59 | 0.25 | 0.88 | 0.86 | 52 | 0 | 0 | 7 | 0.88 | 1.00 | 0.94 | 0.47 | 0.88 | 1.00 |
| model_331 | Cleft palate | 167 | 588 | 421 | 470 | 59 | 59 | 0.24 | 0.9 | 0.9 | 53 | 0 | 0 | 6 | 0.9 | 1.00 | 0.95 | 0.41 | 0.9 | 1.00 |
| model_332 | Clotting | 6520 | 6520 | 0 | 5216 | 652 | 652 | 0.26 | 0.95 | 0.93 | 617 | 0 | 0 | 35 | 0.95 | 1.00 | 0.97 | 0.21 | 0.95 | 1.00 |
| model_333 | Cluster headache | 603 | 603 | 0 | 482 | 61 | 60 | 0.24 | 0.98 | 0.98 | 60 | 0 | 0 | 1 | 0.98 | 1.00 | 0.99 | 0.07 | 0.98 | 1.00 |
| model_334 | Coated tongue | 1006 | 1006 | 0 | 804 | 101 | 101 | 0.24 | 0.91 | 0.94 | 92 | 0 | 0 | 9 | 0.91 | 1.00 | 0.95 | 0.36 | 0.91 | 1.00 |
| model_335 | Coccidioidomycosis | 381 | 588 | 207 | 470 | 59 | 59 | 0.24 | 0.9 | 0.88 | 53 | 0 | 0 | 6 | 0.9 | 1.00 | 0.95 | 0.41 | 0.9 | 1.00 |
| model_336 | Coccydynia | 508 | 588 | 80 | 470 | 59 | 59 | 0.23 | 0.95 | 0.93 | 56 | 0 | 0 | 3 | 0.95 | 1.00 | 0.97 | 0.2 | 0.95 | 1.00 |
| model_337 | Coeliac disease | 1353 | 1353 | 0 | 1082 | 136 | 135 | 0.24 | 0.96 | 0.93 | 130 | 0 | 0 | 6 | 0.96 | 1.00 | 0.98 | 0.18 | 0.96 | 1.00 |
| model_338 | Cognitive disorder | 6771 | 6771 | 0 | 5416 | 678 | 677 | 0.26 | 0.94 | 0.96 | 639 | 0 | 0 | 39 | 0.94 | 1.00 | 0.97 | 0.23 | 0.94 | 1.00 |
| model_339 | Cold extremities | 3163 | 3163 | 0 | 2530 | 317 | 316 | 0.25 | 0.97 | 0.96 | 307 | 0 | 0 | 10 | 0.97 | 1.00 | 0.98 | 0.13 | 0.97 | 1.00 |
| model_340 | Colitis | 6052 | 6052 | 0 | 4841 | 606 | 605 | 0.27 | 0.96 | 0.93 | 583 | 0 | 0 | 23 | 0.96 | 1.00 | 0.98 | 0.15 | 0.96 | 1.00 |
| model_341 | Colitis ischemic | 3528 | 3528 | 0 | 2822 | 353 | 353 | 0.25 | 0.94 | 0.96 | 332 | 0 | 0 | 21 | 0.94 | 1.00 | 0.97 | 0.24 | 0.94 | 1.00 |
| model_342 | Colitis pseudomembranous | 1287 | 1287 | 0 | 1029 | 129 | 129 | 0.23 | 0.91 | 0.95 | 118 | 0 | 0 | 11 | 0.91 | 1.00 | 0.96 | 0.34 | 0.91 | 1.00 |
| model_343 | Collagen disease | 504 | 588 | 84 | 470 | 59 | 59 | 0.24 | 0.95 | 0.97 | 56 | 0 | 0 | 3 | 0.95 | 1.00 | 0.97 | 0.2 | 0.95 | 1.00 |

| | | | | | | | | | | | | | | | | | | | | |
|---|---|---|---|---|---|---|---|---|---|---|---|---|---|---|---|---|---|---|---|---|
| model_344 | Collagenvascular disease | 224 | 588 | 364 | 470 | 59 | 59 | 0.24 | 0.83 | 0.85 | 49 | 0 | 0 | 10 | 0.83 | 1.00 | 0.91 | 0.68 | 0.83 | 1.00 |
| model_345 | Colon Spastic | 4738 | 4738 | 0 | 3790 | 474 | 474 | 0.25 | 0.96 | 0.96 | 453 | 0 | 0 | 21 | 0.96 | 1.00 | 0.98 | 0.18 | 0.96 | 1.00 |
| model_346 | Colon neoplasm | 555 | 588 | 33 | 470 | 59 | 59 | 0.23 | 0.9 | 0.98 | 53 | 0 | 0 | 6 | 0.9 | 1.00 | 0.95 | 0.41 | 0.9 | 1.00 |
| model_347 | Colon polypectomy | 381 | 588 | 207 | 470 | 59 | 59 | 0.23 | 0.93 | 0.92 | 55 | 0 | 0 | 4 | 0.93 | 1.00 | 0.96 | 0.27 | 0.93 | 1.00 |
| model_348 | Colonic obstruction | 736 | 736 | 0 | 588 | 74 | 74 | 0.23 | 0.92 | 0.92 | 68 | 0 | 0 | 6 | 0.92 | 1.00 | 0.96 | 0.32 | 0.92 | 1.00 |
| model_349 | Colonic polyp | 4698 | 4698 | 0 | 3758 | 470 | 470 | 0.25 | 0.97 | 0.96 | 455 | 0 | 0 | 15 | 0.97 | 1.00 | 0.98 | 0.13 | 0.97 | 1.00 |
| model_350 | Color blindness | 457 | 588 | 131 | 470 | 59 | 59 | 0.23 | 0.93 | 0.88 | 55 | 0 | 0 | 4 | 0.93 | 1.00 | 0.96 | 0.27 | 0.93 | 1.00 |
| model_351 | Colostomy | 776 | 776 | 0 | 620 | 78 | 78 | 0.23 | 0.96 | 0.99 | 75 | 0 | 0 | 3 | 0.96 | 1.00 | 0.98 | 0.15 | 0.96 | 1.00 |
| model_352 | Colpitis | 2415 | 2415 | 0 | 1932 | 242 | 241 | 0.24 | 0.95 | 0.96 | 231 | 0 | 0 | 11 | 0.95 | 1.00 | 0.98 | 0.18 | 0.95 | 1.00 |
| model_353 | Coma | 9215 | 9215 | 0 | 7372 | 922 | 921 | 0.28 | 0.94 | 0.94 | 863 | 0 | 0 | 59 | 0.94 | 1.00 | 0.97 | 0.26 | 0.94 | 1.00 |
| model_354 | Confusion | 19365 | 19365 | 0 | 15492 | 1937 | 1936 | 0.34 | 0.94 | 0.95 | 1828 | 0 | 0 | 109 | 0.94 | 1.00 | 0.97 | 0.23 | 0.94 | 1.00 |
| model_355 | Congenital heart disease | 912 | 912 | 0 | 729 | 92 | 91 | 0.23 | 0.92 | 0.96 | 85 | 0 | 0 | 7 | 0.92 | 1.00 | 0.96 | 0.3 | 0.92 | 1.00 |
| model_356 | Conjunctivitis | 8170 | 8170 | 0 | 6536 | 817 | 817 | 0.28 | 0.95 | 0.96 | 778 | 0 | 0 | 39 | 0.95 | 1.00 | 0.98 | 0.19 | 0.95 | 1.00 |
| model_357 | Conjunctivitis viral | 166 | 588 | 422 | 470 | 59 | 59 | 0.24 | 0.93 | 0.9 | 55 | 0 | 0 | 4 | 0.93 | 1.00 | 0.96 | 0.27 | 0.93 | 1.00 |
| model_358 | Connective tissue disease | 978 | 978 | 0 | 782 | 98 | 98 | 0.24 | 0.93 | 0.94 | 91 | 0 | 0 | 7 | 0.93 | 1.00 | 0.96 | 0.29 | 0.93 | 1.00 |
| model_359 | Constipated | 17639 | 17639 | 0 | 14111 | 1764 | 1764 | 0.32 | 0.96 | 0.95 | 1695 | 0 | 0 | 69 | 0.96 | 1.00 | 0.98 | 0.16 | 0.96 | 1.00 |
| model_360 | Constrictive pericarditis | 339 | 588 | 249 | 470 | 59 | 59 | 0.23 | 0.98 | 1 | 58 | 0 | 0 | 1 | 0.98 | 1.00 | 0.99 | 0.07 | 0.98 | 1.00 |
| model_361 | Contact dermatitis | 2062 | 2062 | 0 | 1649 | 207 | 206 | 0.25 | 0.95 | 0.95 | 196 | 0 | 0 | 11 | 0.95 | 1.00 | 0.97 | 0.21 | 0.95 | 1.00 |
| model_362 | Contracture | 1669 | 1669 | 0 | 1335 | 167 | 167 | 0.23 | 0.93 | 0.91 | 156 | 0 | 0 | 11 | 0.93 | 1.00 | 0.97 | 0.26 | 0.93 | 1.00 |
| model_363 | Conversion disorder | 1181 | 1181 | 0 | 944 | 119 | 118 | 0.24 | 0.98 | 0.9 | 117 | 0 | 0 | 2 | 0.98 | 1.00 | 0.99 | 0.07 | 0.98 | 1.00 |
| model_364 | Convulsion | 13688 | 13688 | 0 | 10950 | 1369 | 1369 | 0.3 | 0.95 | 0.95 | 1301 | 0 | 0 | 68 | 0.95 | 1.00 | 0.97 | 0.2 | 0.95 | 1.00 |
| model_365 | Convulsions Grand Mal | 5447 | 5447 | 0 | 4357 | 545 | 545 | 0.26 | 0.94 | 0.96 | 515 | 0 | 0 | 30 | 0.94 | 1.00 | 0.97 | 0.22 | 0.94 | 1.00 |
| model_366 | Cor pulmonale | 1521 | 1521 | 0 | 1216 | 153 | 152 | 0.24 | 0.93 | 0.95 | 143 | 0 | 0 | 10 | 0.93 | 1.00 | 0.97 | 0.26 | 0.93 | 1.00 |
| model_367 | Corneal abrasion | 687 | 687 | 0 | 549 | 69 | 69 | 0.23 | 0.91 | 0.96 | 63 | 0 | 0 | 6 | 0.91 | 1.00 | 0.95 | 0.35 | 0.91 | 1.00 |
| model_368 | Corneal clouding | 200 | 588 | 388 | 470 | 59 | 59 | 0.24 | 0.83 | 0.9 | 49 | 0 | 0 | 10 | 0.83 | 1.00 | 0.91 | 0.68 | 0.83 | 1.00 |
| model_369 | Corneal degeneration | 11 | 588 | 577 | 470 | 59 | 59 | 0.23 | 0 | 0 | 0 | 0 | 0 | 59 | 0 | 0.00 | 0 | 4 | 0 | 1.00 |
| model_370 | Corneal disorder | 634 | 634 | 0 | 507 | 64 | 63 | 0.23 | 0.94 | 0.94 | 60 | 0 | 0 | 4 | 0.94 | 1.00 | 0.97 | 0.25 | 0.94 | 1.00 |
| model_371 | Corneal dystrophy | 410 | 588 | 178 | 470 | 59 | 59 | 0.23 | 0.88 | 0.88 | 52 | 0 | 0 | 7 | 0.88 | 1.00 | 0.94 | 0.47 | 0.88 | 1.00 |
| model_372 | Corneal ulcer | 814 | 814 | 0 | 651 | 82 | 81 | 0.24 | 0.96 | 0.98 | 79 | 0 | 0 | 3 | 0.96 | 1.00 | 0.98 | 0.15 | 0.96 | 1.00 |
| model_373 | Coronary angioplasty | 653 | 653 | 0 | 522 | 66 | 65 | 0.23 | 0.95 | 0.92 | 63 | 0 | 0 | 3 | 0.95 | 1.00 | 0.98 | 0.18 | 0.95 | 1.00 |
| model_374 | Coronary artery bypass graft | 1327 | 1327 | 0 | 1061 | 133 | 133 | 0.23 | 0.96 | 0.98 | 128 | 0 | 0 | 5 | 0.96 | 1.00 | 0.98 | 0.15 | 0.96 | 1.00 |
| model_375 | Corticosteroid therapy | 7 | 588 | 581 | 470 | 59 | 59 | 0.23 | 0 | 0 | 0 | 0 | 0 | 59 | 0 | 0.00 | 0 | 4 | 0 | 1.00 |
| model_376 | Costochondritis | 1703 | 1703 | 0 | 1362 | 171 | 170 | 0.25 | 0.95 | 0.94 | 162 | 0 | 0 | 9 | 0.95 | 1.00 | 0.97 | 0.21 | 0.95 | 1.00 |
| model_377 | Cough | 17648 | 17648 | 0 | 14118 | 1765 | 1765 | 0.32 | 0.96 | 0.96 | 1696 | 0 | 0 | 69 | 0.96 | 1.00 | 0.98 | 0.16 | 0.96 | 1.00 |
| model_378 | Coughing blood | 7692 | 7692 | 0 | 6153 | 770 | 769 | 0.27 | 0.93 | 0.93 | 719 | 0 | 0 | 51 | 0.93 | 1.00 | 0.97 | 0.26 | 0.93 | 1.00 |
| model_379 | Cramp | 3858 | 3858 | 0 | 3086 | 386 | 386 | 0.25 | 0.97 | 0.95 | 373 | 0 | 0 | 13 | 0.97 | 1.00 | 0.98 | 0.13 | 0.97 | 1.00 |
| model_380 | Cryptococcosis | 1056 | 1056 | 0 | 844 | 106 | 106 | 0.23 | 0.91 | 0.9 | 96 | 0 | 0 | 10 | 0.91 | 1.00 | 0.95 | 0.38 | 0.91 | 1.00 |
| model_381 | Cryptorchidism | 72 | 588 | 516 | 470 | 59 | 59 | 0.26 | 0.75 | 0.71 | 44 | 0 | 0 | 15 | 0.75 | 1.00 | 0.85 | 1.02 | 0.75 | 1.00 |
| model_382 | Cryptosporidiosis | 57 | 588 | 531 | 470 | 59 | 59 | 0.26 | 0.44 | 0.63 | 26 | 0 | 0 | 33 | 0.44 | 1.00 | 0.61 | 2.24 | 0.44 | 1.00 |
| model_383 | Cutaneous candidiasis | 617 | 617 | 0 | 493 | 62 | 62 | 0.23 | 0.9 | 0.94 | 56 | 0 | 0 | 6 | 0.9 | 1.00 | 0.95 | 0.39 | 0.9 | 1.00 |
| model_384 | Cutaneous mycosis | 937 | 937 | 0 | 749 | 94 | 94 | 0.22 | 0.9 | 0.88 | 85 | 0 | 0 | 9 | 0.9 | 1.00 | 0.95 | 0.38 | 0.9 | 1.00 |
| model_385 | Cutaneous ulcer | 6735 | 6735 | 0 | 5388 | 674 | 673 | 0.27 | 0.95 | 0.95 | 640 | 0 | 0 | 34 | 0.95 | 1.00 | 0.97 | 0.2 | 0.95 | 1.00 |
| model_386 | Cyst | 5076 | 5076 | 0 | 4060 | 508 | 508 | 0.25 | 0.97 | 0.95 | 493 | 0 | 0 | 15 | 0.97 | 1.00 | 0.99 | 0.12 | 0.97 | 1.00 |
| model_387 | Cystic Fibrosis | 112 | 588 | 476 | 470 | 59 | 59 | 0.25 | 0.83 | 0.88 | 49 | 0 | 0 | 10 | 0.83 | 1.00 | 0.91 | 0.68 | 0.83 | 1.00 |
| model_388 | Cystic acne | 318 | 588 | 270 | 470 | 59 | 59 | 0.24 | 0.75 | 0.9 | 44 | 0 | 0 | 15 | 0.75 | 1.00 | 0.85 | 1.02 | 0.75 | 1.00 |
| model_389 | Cystitis Interstitial | 1085 | 1085 | 0 | 868 | 109 | 108 | 0.23 | 0.93 | 0.95 | 101 | 0 | 0 | 8 | 0.93 | 1.00 | 0.96 | 0.29 | 0.93 | 1.00 |
| model_390 | Dacrocystitis | 32 | 588 | 556 | 470 | 59 | 59 | 0.28 | 0.03 | 0.15 | 2 | 0 | 0 | 57 | 0.03 | 1.00 | 0.07 | 3.86 | 0.03 | 1.00 |
| model_391 | Dandruff | 207 | 588 | 381 | 470 | 59 | 59 | 0.23 | 0.88 | 0.92 | 52 | 0 | 0 | 7 | 0.88 | 1.00 | 0.94 | 0.47 | 0.88 | 1.00 |
| model_392 | Deafness | 5165 | 5165 | 0 | 4132 | 517 | 516 | 0.25 | 0.96 | 0.96 | 497 | 0 | 0 | 20 | 0.96 | 1.00 | 0.98 | 0.15 | 0.96 | 1.00 |
| model_393 | Deafness neurosensory | 1371 | 1371 | 0 | 1096 | 138 | 137 | 0.22 | 0.97 | 0.98 | 134 | 0 | 0 | 4 | 0.97 | 1.00 | 0.99 | 0.12 | 0.97 | 1.00 |

| | | | | | | | | | | | | | | | | | | | | |
|---|---|---|---|---|---|---|---|---|---|---|---|---|---|---|---|---|---|---|---|---|
| model_394 | Decreased Libido | 3664 | 3664 | 0 | 2931 | 367 | 366 | 0.25 | 0.96 | 0.97 | 351 | 0 | 0 | 16 | 0.96 | 1.00 | 0.98 | 0.17 | 0.96 | 1.00 |
| model_395 | Decreased body temperature | 5255 | 5255 | 0 | 4204 | 526 | 525 | 0.26 | 0.93 | 0.94 | 490 | 0 | 0 | 36 | 0.93 | 1.00 | 0.96 | 0.27 | 0.93 | 1.00 |
| model_396 | Decreased hearing | 4335 | 4335 | 0 | 3468 | 434 | 433 | 0.25 | 0.95 | 0.96 | 412 | 0 | 0 | 22 | 0.95 | 1.00 | 0.97 | 0.2 | 0.95 | 1.00 |
| model_397 | Decreased lacrimation | 108 | 588 | 480 | 470 | 59 | 59 | 0.25 | 0.85 | 0.88 | 50 | 0 | 0 | 9 | 0.85 | 1.00 | 0.92 | 0.61 | 0.85 | 1.00 |
| model_398 | Decubitus ulcer | 4875 | 4875 | 0 | 3900 | 488 | 487 | 0.25 | 0.93 | 0.94 | 455 | 0 | 0 | 33 | 0.93 | 1.00 | 0.97 | 0.27 | 0.93 | 1.00 |
| model_399 | Deep vein thromboses | 11940 | 11940 | 0 | 9552 | 1194 | 1194 | 0.3 | 0.95 | 0.95 | 1130 | 0 | 0 | 64 | 0.95 | 1.00 | 0.97 | 0.21 | 0.95 | 1.00 |
| model_400 | Defaecation urgency | 729 | 729 | 0 | 583 | 73 | 73 | 0.22 | 0.96 | 0.93 | 70 | 0 | 0 | 3 | 0.96 | 1.00 | 0.98 | 0.16 | 0.96 | 1.00 |
| model_401 | Deglutition disorder | 14387 | 14387 | 0 | 11509 | 1439 | 1439 | 0.3 | 0.96 | 0.96 | 1377 | 0 | 0 | 62 | 0.96 | 1.00 | 0.98 | 0.17 | 0.96 | 1.00 |
| model_402 | Dehydration | 19100 | 19100 | 0 | 15280 | 1910 | 1910 | 0.34 | 0.95 | 0.95 | 1824 | 0 | 0 | 86 | 0.95 | 1.00 | 0.98 | 0.18 | 0.95 | 1.00 |
| model_403 | Delirium tremens | 421 | 588 | 167 | 470 | 59 | 59 | 0.22 | 0.93 | 0.85 | 55 | 0 | 0 | 4 | 0.93 | 1.00 | 0.96 | 0.27 | 0.93 | 1.00 |
| model_404 | Dental abscess | 4450 | 4450 | 0 | 3560 | 445 | 445 | 0.25 | 0.96 | 0.97 | 426 | 0 | 0 | 19 | 0.96 | 1.00 | 0.98 | 0.17 | 0.96 | 1.00 |
| model_405 | Dental caries | 5268 | 5268 | 0 | 4214 | 527 | 527 | 0.26 | 0.95 | 0.97 | 502 | 0 | 0 | 25 | 0.95 | 1.00 | 0.98 | 0.19 | 0.95 | 1.00 |
| model_406 | Dental pain | 5059 | 5059 | 0 | 4047 | 506 | 506 | 0.27 | 0.97 | 0.97 | 491 | 0 | 0 | 15 | 0.97 | 1.00 | 0.98 | 0.12 | 0.97 | 1.00 |
| model_407 | Depigmentation | 232 | 588 | 356 | 470 | 59 | 59 | 0.25 | 0.83 | 0.93 | 49 | 0 | 0 | 10 | 0.83 | 1.00 | 0.91 | 0.68 | 0.83 | 1.00 |
| model_408 | Dermatitides | 4215 | 4215 | 0 | 3372 | 422 | 421 | 0.25 | 0.93 | 0.96 | 394 | 0 | 0 | 28 | 0.93 | 1.00 | 0.97 | 0.27 | 0.93 | 1.00 |
| model_409 | Dermatitis diaper | 86 | 588 | 502 | 470 | 59 | 59 | 0.28 | 0.73 | 0.78 | 43 | 0 | 0 | 16 | 0.73 | 1.00 | 0.84 | 1.08 | 0.73 | 1.00 |
| model_410 | Dermatitis exfoliative | 2546 | 2546 | 0 | 2036 | 255 | 255 | 0.25 | 0.95 | 0.91 | 242 | 0 | 0 | 13 | 0.95 | 1.00 | 0.97 | 0.2 | 0.95 | 1.00 |
| model_411 | Dermatitis medicamentosa | 3737 | 3737 | 0 | 2989 | 374 | 374 | 0.25 | 0.93 | 0.95 | 347 | 0 | 0 | 27 | 0.93 | 1.00 | 0.96 | 0.29 | 0.93 | 1.00 |
| model_412 | Dermatitis seborrheic | 1145 | 1145 | 0 | 916 | 115 | 114 | 0.24 | 0.95 | 0.96 | 109 | 0 | 0 | 6 | 0.95 | 1.00 | 0.97 | 0.21 | 0.95 | 1.00 |
| model_413 | Dermatomyositis | 788 | 788 | 0 | 630 | 79 | 79 | 0.23 | 0.99 | 0.94 | 78 | 0 | 0 | 1 | 0.99 | 1.00 | 0.99 | 0.05 | 0.99 | 1.00 |
| model_414 | Dermatophytosis | 143 | 588 | 445 | 470 | 59 | 59 | 0.25 | 0.71 | 0.81 | 42 | 0 | 0 | 17 | 0.71 | 1.00 | 0.83 | 1.15 | 0.71 | 1.00 |
| model_415 | Dermoid cyst | 269 | 588 | 319 | 470 | 59 | 59 | 0.23 | 0.8 | 0.9 | 47 | 0 | 0 | 12 | 0.8 | 1.00 | 0.89 | 0.81 | 0.8 | 1.00 |
| model_416 | Detached retina | 2110 | 2110 | 0 | 1688 | 211 | 211 | 0.25 | 0.94 | 0.96 | 199 | 0 | 0 | 12 | 0.94 | 1.00 | 0.97 | 0.23 | 0.94 | 1.00 |
| model_417 | Diabetes | 12718 | 12718 | 0 | 10174 | 1272 | 1272 | 0.3 | 0.98 | 0.97 | 1246 | 0 | 0 | 26 | 0.98 | 1.00 | 0.99 | 0.08 | 0.98 | 1.00 |
| model_418 | Diabetes insipidus | 1236 | 1236 | 0 | 988 | 124 | 124 | 0.23 | 0.94 | 0.96 | 116 | 0 | 0 | 8 | 0.94 | 1.00 | 0.97 | 0.26 | 0.94 | 1.00 |
| model_419 | Diabetic Nephropathy | 1264 | 1264 | 0 | 1011 | 127 | 126 | 0.24 | 0.94 | 0.94 | 120 | 0 | 0 | 7 | 0.94 | 1.00 | 0.97 | 0.22 | 0.94 | 1.00 |
| model_420 | Diabetic Retinopathy | 2982 | 2982 | 0 | 2385 | 299 | 298 | 0.25 | 0.95 | 0.96 | 285 | 0 | 0 | 14 | 0.95 | 1.00 | 0.98 | 0.19 | 0.95 | 1.00 |
| model_421 | Diabetic acidosis | 3523 | 3523 | 0 | 2818 | 353 | 352 | 0.25 | 0.96 | 0.96 | 339 | 0 | 0 | 14 | 0.96 | 1.00 | 0.98 | 0.16 | 0.96 | 1.00 |
| model_422 | Diabetic neuropathy | 5168 | 5168 | 0 | 4134 | 517 | 517 | 0.25 | 0.95 | 0.95 | 490 | 0 | 0 | 27 | 0.95 | 1.00 | 0.97 | 0.21 | 0.95 | 1.00 |
| model_423 | Diaphragmatic hernia | 858 | 858 | 0 | 686 | 86 | 86 | 0.23 | 0.93 | 0.94 | 80 | 0 | 0 | 6 | 0.93 | 1.00 | 0.96 | 0.28 | 0.93 | 1.00 |
| model_424 | Diarrhea | 23848 | 23848 | 0 | 19078 | 2385 | 2385 | 0.36 | 0.95 | 0.95 | 2259 | 0 | 0 | 126 | 0.95 | 1.00 | 0.97 | 0.21 | 0.95 | 1.00 |
| model_425 | Diarrhea infectious | 428 | 588 | 160 | 470 | 59 | 59 | 0.23 | 0.92 | 0.97 | 54 | 0 | 0 | 5 | 0.92 | 1.00 | 0.96 | 0.34 | 0.92 | 1.00 |
| model_426 | Difficulty breathing | 26037 | 26037 | 0 | 20829 | 2604 | 2604 | 0.37 | 0.96 | 0.96 | 2489 | 0 | 0 | 115 | 0.96 | 1.00 | 0.98 | 0.18 | 0.96 | 1.00 |
| model_427 | Difficulty in walking | 7040 | 7040 | 0 | 5632 | 704 | 704 | 0.28 | 0.95 | 0.97 | 671 | 0 | 0 | 33 | 0.95 | 1.00 | 0.98 | 0.19 | 0.95 | 1.00 |
| model_428 | Diphtheria | 106 | 588 | 482 | 470 | 59 | 59 | 0.27 | 0.68 | 0.66 | 40 | 0 | 0 | 19 | 0.68 | 1.00 | 0.81 | 1.29 | 0.68 | 1.00 |
| model_429 | Diplopia | 6848 | 6848 | 0 | 5478 | 685 | 685 | 0.24 | 0.95 | 0.96 | 651 | 0 | 0 | 34 | 0.95 | 1.00 | 0.97 | 0.2 | 0.95 | 1.00 |
| model_430 | Disease of liver | 7092 | 7092 | 0 | 5673 | 710 | 709 | 0.25 | 0.95 | 0.94 | 676 | 0 | 0 | 34 | 0.95 | 1.00 | 0.98 | 0.19 | 0.95 | 1.00 |
| model_431 | Dislocation | 2730 | 2730 | 0 | 2184 | 273 | 273 | 0.25 | 0.97 | 0.95 | 265 | 0 | 0 | 8 | 0.97 | 1.00 | 0.99 | 0.12 | 0.97 | 1.00 |
| model_432 | Disorder Lung | 8557 | 8557 | 0 | 6845 | 856 | 856 | 0.27 | 0.96 | 0.94 | 818 | 0 | 0 | 38 | 0.96 | 1.00 | 0.98 | 0.18 | 0.96 | 1.00 |
| model_433 | Disorder Peripheral Vascular | 5165 | 5165 | 0 | 4132 | 517 | 516 | 0.26 | 0.95 | 0.94 | 490 | 0 | 0 | 27 | 0.95 | 1.00 | 0.97 | 0.21 | 0.95 | 1.00 |
| model_434 | Disorder Renal | 9086 | 9086 | 0 | 7268 | 909 | 909 | 0.28 | 0.95 | 0.93 | 861 | 0 | 0 | 48 | 0.95 | 1.00 | 0.97 | 0.21 | 0.95 | 1.00 |
| model_435 | Disorder Retinal | 2063 | 2063 | 0 | 1650 | 207 | 206 | 0.24 | 0.93 | 0.96 | 193 | 0 | 0 | 14 | 0.93 | 1.00 | 0.97 | 0.27 | 0.93 | 1.00 |
| model_436 | Disorders of lipid metabolism | 134 | 588 | 454 | 470 | 59 | 59 | 0.24 | 0.8 | 0.85 | 47 | 0 | 0 | 12 | 0.8 | 1.00 | 0.89 | 0.81 | 0.8 | 1.00 |
| model_437 | Disseminated lupus erythematosus | 3974 | 3974 | 0 | 3179 | 398 | 397 | 0.24 | 0.94 | 0.95 | 374 | 0 | 0 | 24 | 0.94 | 1.00 | 0.97 | 0.24 | 0.94 | 1.00 |
| model_438 | Distended bladder | 653 | 653 | 0 | 522 | 66 | 65 | 0.24 | 0.91 | 0.88 | 60 | 0 | 0 | 6 | 0.91 | 1.00 | 0.95 | 0.36 | 0.91 | 1.00 |
| model_439 | Diverticulitis | 6940 | 6940 | 0 | 5552 | 694 | 694 | 0.27 | 0.95 | 0.95 | 659 | 0 | 0 | 35 | 0.95 | 1.00 | 0.97 | 0.2 | 0.95 | 1.00 |
| model_440 | Diverticulosis | 2763 | 2763 | 0 | 2210 | 277 | 276 | 0.25 | 0.96 | 0.96 | 267 | 0 | 0 | 10 | 0.96 | 1.00 | 0.98 | 0.14 | 0.96 | 1.00 |
| model_441 | Dizziness | 20204 | 20204 | 0 | 16163 | 2021 | 2020 | 0.34 | 0.96 | 0.96 | 1935 | 0 | 0 | 86 | 0.96 | 1.00 | 0.98 | 0.17 | 0.96 | 1.00 |
| model_442 | Drowsiness | 15545 | 15545 | 0 | 12436 | 1555 | 1554 | 0.31 | 0.95 | 0.95 | 1476 | 0 | 0 | 79 | 0.95 | 1.00 | 0.97 | 0.2 | 0.95 | 1.00 |
| model_443 | Drug addiction | 7781 | 7781 | 0 | 6224 | 779 | 778 | 0.27 | 0.96 | 0.96 | 751 | 0 | 0 | 28 | 0.96 | 1.00 | 0.98 | 0.14 | 0.96 | 1.00 |

| | | | | | | | | | | | | | | | | | | | | |
|---|---|---|---|---|---|---|---|---|---|---|---|---|---|---|---|---|---|---|---|---|
| model_444 | Drug hypersensitivity | 9358 | 9358 | 0 | 7486 | 936 | 936 | 0.28 | 0.95 | 0.95 | 886 | 0 | 0 | 50 | 0.95 | 1.00 | 0.97 | 0.21 | 0.95 | 1.00 |
| model_445 | Drug toxicity NOS | 10952 | 10952 | 0 | 8761 | 1096 | 1095 | 0.29 | 0.95 | 0.95 | 1046 | 0 | 0 | 50 | 0.95 | 1.00 | 0.98 | 0.18 | 0.95 | 1.00 |
| model_446 | Drug withdrawal | 7956 | 7956 | 0 | 6364 | 796 | 796 | 0.27 | 0.97 | 0.95 | 769 | 0 | 0 | 27 | 0.97 | 1.00 | 0.98 | 0.14 | 0.97 | 1.00 |
| model_447 | Dry eye | 2782 | 2782 | 0 | 2225 | 279 | 278 | 0.24 | 0.95 | 0.97 | 264 | 0 | 0 | 15 | 0.95 | 1.00 | 0.97 | 0.22 | 0.95 | 1.00 |
| model_448 | Dry skin | 6124 | 6124 | 0 | 4899 | 613 | 612 | 0.26 | 0.95 | 0.96 | 585 | 0 | 0 | 28 | 0.95 | 1.00 | 0.98 | 0.18 | 0.95 | 1.00 |
| model_449 | Duodenal ulcer | 3117 | 3117 | 0 | 2493 | 312 | 312 | 0.25 | 0.92 | 0.9 | 287 | 0 | 0 | 25 | 0.92 | 1.00 | 0.96 | 0.32 | 0.92 | 1.00 |
| model_450 | Duodenal ulcer haemorrhage | 1244 | 1244 | 0 | 995 | 125 | 124 | 0.23 | 0.94 | 0.9 | 117 | 0 | 0 | 8 | 0.94 | 1.00 | 0.97 | 0.26 | 0.94 | 1.00 |
| model_451 | Duodenal ulcer perforation | 1001 | 1001 | 0 | 800 | 101 | 100 | 0.22 | 0.93 | 0.89 | 94 | 0 | 0 | 7 | 0.93 | 1.00 | 0.96 | 0.28 | 0.93 | 1.00 |
| model_452 | Duodenitis | 2964 | 2964 | 0 | 2371 | 297 | 296 | 0.24 | 0.95 | 0.96 | 281 | 0 | 0 | 16 | 0.95 | 1.00 | 0.97 | 0.22 | 0.95 | 1.00 |
| model_453 | Dwarfism | 2 | 588 | 586 | 470 | 59 | 59 | 0.28 | 0 | 0 | 0 | 0 | 0 | 59 | 0 | 0.00 | 0 | 4 | 0 | 1.00 |
| model_454 | Dysaesthesia | 1556 | 1556 | 0 | 1244 | 156 | 156 | 0.24 | 0.94 | 0.99 | 146 | 0 | 0 | 10 | 0.94 | 1.00 | 0.97 | 0.26 | 0.94 | 1.00 |
| model_455 | Dysarthria | 9869 | 9869 | 0 | 7895 | 987 | 987 | 0.32 | 0.96 | 0.96 | 952 | 0 | 0 | 35 | 0.96 | 1.00 | 0.98 | 0.14 | 0.96 | 1.00 |
| model_456 | Dyshidrosis | 553 | 588 | 35 | 470 | 59 | 59 | 0.23 | 0.86 | 0.88 | 51 | 0 | 0 | 8 | 0.86 | 1.00 | 0.93 | 0.54 | 0.86 | 1.00 |
| model_457 | Dyslexia | 179 | 588 | 409 | 470 | 59 | 59 | 0.24 | 0.9 | 0.88 | 53 | 0 | 0 | 6 | 0.9 | 1.00 | 0.95 | 0.41 | 0.9 | 1.00 |
| model_458 | Dysmenorrhea | 670 | 670 | 0 | 536 | 67 | 67 | 0.23 | 0.96 | 0.96 | 64 | 0 | 0 | 3 | 0.96 | 1.00 | 0.98 | 0.18 | 0.96 | 1.00 |
| model_459 | Dyspareunia | 598 | 598 | 0 | 478 | 60 | 60 | 0.22 | 0.95 | 0.97 | 57 | 0 | 0 | 3 | 0.95 | 1.00 | 0.97 | 0.2 | 0.95 | 1.00 |
| model_460 | Dyspepsia | 13145 | 13145 | 0 | 10516 | 1315 | 1314 | 0.3 | 0.96 | 0.96 | 1262 | 0 | 0 | 53 | 0.96 | 1.00 | 0.98 | 0.16 | 0.96 | 1.00 |
| model_461 | Dysphasia | 3096 | 3096 | 0 | 2476 | 310 | 310 | 0.24 | 0.95 | 0.94 | 293 | 0 | 0 | 17 | 0.95 | 1.00 | 0.97 | 0.22 | 0.95 | 1.00 |
| model_462 | Dysphemia | 997 | 997 | 0 | 797 | 100 | 100 | 0.23 | 0.98 | 0.97 | 98 | 0 | 0 | 2 | 0.98 | 1.00 | 0.99 | 0.08 | 0.98 | 1.00 |
| model_463 | Dysphonia | 6101 | 6101 | 0 | 4880 | 611 | 610 | 0.27 | 0.97 | 0.95 | 593 | 0 | 0 | 18 | 0.97 | 1.00 | 0.99 | 0.12 | 0.97 | 1.00 |
| model_464 | Dyspnea exertional | 9666 | 9666 | 0 | 7732 | 967 | 967 | 0.27 | 0.94 | 0.95 | 910 | 0 | 0 | 57 | 0.94 | 1.00 | 0.97 | 0.24 | 0.94 | 1.00 |
| model_465 | Dyspnoea paroxysmal nocturnal | 1528 | 1528 | 0 | 1222 | 153 | 153 | 0.23 | 0.94 | 0.93 | 144 | 0 | 0 | 9 | 0.94 | 1.00 | 0.97 | 0.24 | 0.94 | 1.00 |
| model_466 | Dysuria | 8807 | 8807 | 0 | 7045 | 881 | 881 | 0.28 | 0.95 | 0.96 | 833 | 0 | 0 | 48 | 0.95 | 1.00 | 0.97 | 0.22 | 0.95 | 1.00 |
| model_467 | EBV infection | 1216 | 1216 | 0 | 972 | 122 | 122 | 0.23 | 0.94 | 0.89 | 115 | 0 | 0 | 7 | 0.94 | 1.00 | 0.97 | 0.23 | 0.94 | 1.00 |
| model_468 | Ear ache | 5489 | 5489 | 0 | 4391 | 549 | 549 | 0.26 | 0.96 | 0.96 | 525 | 0 | 0 | 24 | 0.96 | 1.00 | 0.98 | 0.17 | 0.96 | 1.00 |
| model_469 | Ear discharge | 747 | 747 | 0 | 597 | 75 | 75 | 0.23 | 0.89 | 0.87 | 67 | 0 | 0 | 8 | 0.89 | 1.00 | 0.94 | 0.43 | 0.89 | 1.00 |
| model_470 | Ear infection | 3541 | 3541 | 0 | 2832 | 355 | 354 | 0.25 | 0.97 | 0.95 | 345 | 0 | 0 | 10 | 0.97 | 1.00 | 0.99 | 0.11 | 0.97 | 1.00 |
| model_471 | Easy bruisability | 2455 | 2455 | 0 | 1964 | 246 | 245 | 0.24 | 0.96 | 0.97 | 236 | 0 | 0 | 10 | 0.96 | 1.00 | 0.98 | 0.16 | 0.96 | 1.00 |
| model_472 | Eating disorder | 6163 | 6163 | 0 | 4930 | 617 | 616 | 0.27 | 0.96 | 0.95 | 593 | 0 | 0 | 24 | 0.96 | 1.00 | 0.98 | 0.16 | 0.96 | 1.00 |
| model_473 | Ecchymoses | 5936 | 5936 | 0 | 4748 | 594 | 594 | 0.27 | 0.94 | 0.94 | 557 | 0 | 0 | 37 | 0.94 | 1.00 | 0.97 | 0.25 | 0.94 | 1.00 |
| model_474 | Eclampsia | 50 | 588 | 538 | 470 | 59 | 59 | 0.25 | 0.63 | 0.61 | 37 | 0 | 0 | 22 | 0.63 | 1.00 | 0.77 | 1.49 | 0.63 | 1.00 |
| model_475 | Ectopic pregnancies | 277 | 588 | 311 | 470 | 59 | 59 | 0.24 | 0.95 | 0.86 | 56 | 0 | 0 | 3 | 0.95 | 1.00 | 0.97 | 0.2 | 0.95 | 1.00 |
| model_476 | Ectropion | 190 | 588 | 398 | 470 | 59 | 59 | 0.23 | 0.93 | 0.85 | 55 | 0 | 0 | 4 | 0.93 | 1.00 | 0.96 | 0.27 | 0.93 | 1.00 |
| model_477 | Eczema | 3592 | 3592 | 0 | 2873 | 360 | 359 | 0.25 | 0.97 | 0.97 | 349 | 0 | 0 | 11 | 0.97 | 1.00 | 0.98 | 0.12 | 0.97 | 1.00 |
| model_478 | Edema | 18419 | 18419 | 0 | 14735 | 1842 | 1842 | 0.32 | 0.95 | 0.96 | 1750 | 0 | 0 | 92 | 0.95 | 1.00 | 0.97 | 0.2 | 0.95 | 1.00 |
| model_479 | Edema extremities | 21981 | 21981 | 0 | 17584 | 2199 | 2198 | 0.35 | 0.96 | 0.95 | 2103 | 0 | 0 | 96 | 0.96 | 1.00 | 0.98 | 0.17 | 0.96 | 1.00 |
| model_480 | Ejaculation Premature | 700 | 700 | 0 | 560 | 70 | 70 | 0.22 | 0.97 | 1 | 68 | 0 | 0 | 2 | 0.97 | 1.00 | 0.99 | 0.11 | 0.97 | 1.00 |
| model_481 | Ekbom Syndrome | 5974 | 5974 | 0 | 4779 | 598 | 597 | 0.25 | 0.98 | 0.96 | 584 | 0 | 0 | 14 | 0.98 | 1.00 | 0.99 | 0.09 | 0.98 | 1.00 |
| model_482 | Electrolyte disorder | 5694 | 5694 | 0 | 4555 | 570 | 569 | 0.26 | 0.96 | 0.95 | 547 | 0 | 0 | 23 | 0.96 | 1.00 | 0.98 | 0.16 | 0.96 | 1.00 |
| model_483 | Elevated cholesterol | 14071 | 14071 | 0 | 11256 | 1408 | 1407 | 0.3 | 0.96 | 0.96 | 1345 | 0 | 0 | 63 | 0.96 | 1.00 | 0.98 | 0.18 | 0.96 | 1.00 |
| model_484 | Elevated erythrocyte sedimentation rate | 3787 | 3787 | 0 | 3029 | 379 | 379 | 0.25 | 0.98 | 0.98 | 370 | 0 | 0 | 9 | 0.98 | 1.00 | 0.99 | 0.09 | 0.98 | 1.00 |
| model_485 | Elevated prostate specific antigen | 1939 | 1939 | 0 | 1551 | 194 | 194 | 0.22 | 0.91 | 0.88 | 177 | 0 | 0 | 17 | 0.91 | 1.00 | 0.95 | 0.35 | 0.91 | 1.00 |
| model_486 | Elevated triglycerides | 3894 | 3894 | 0 | 3115 | 390 | 389 | 0.26 | 0.95 | 0.97 | 372 | 0 | 0 | 18 | 0.95 | 1.00 | 0.98 | 0.18 | 0.95 | 1.00 |
| model_487 | Embolism | 2651 | 2651 | 0 | 2120 | 266 | 265 | 0.25 | 0.94 | 0.91 | 249 | 0 | 0 | 17 | 0.94 | 1.00 | 0.97 | 0.26 | 0.94 | 1.00 |
| model_488 | Embolism pulmonary | 12173 | 12173 | 0 | 9738 | 1218 | 1217 | 0.28 | 0.95 | 0.94 | 1154 | 0 | 0 | 64 | 0.95 | 1.00 | 0.97 | 0.21 | 0.95 | 1.00 |
| model_489 | Emesis | 23043 | 23043 | 0 | 18434 | 2305 | 2304 | 0.36 | 0.95 | 0.95 | 2179 | 0 | 0 | 126 | 0.95 | 1.00 | 0.97 | 0.22 | 0.95 | 1.00 |
| model_490 | Emphysema | 4883 | 4883 | 0 | 3906 | 489 | 488 | 0.26 | 0.96 | 0.95 | 470 | 0 | 0 | 19 | 0.96 | 1.00 | 0.98 | 0.16 | 0.96 | 1.00 |
| model_491 | Empyema | 1215 | 1215 | 0 | 972 | 122 | 121 | 0.23 | 0.89 | 0.93 | 109 | 0 | 0 | 13 | 0.89 | 1.00 | 0.94 | 0.43 | 0.89 | 1.00 |
| model_492 | Encephalitis | 1956 | 1956 | 0 | 1564 | 196 | 196 | 0.24 | 0.9 | 0.91 | 176 | 0 | 0 | 20 | 0.9 | 1.00 | 0.95 | 0.41 | 0.9 | 1.00 |
| model_493 | Encephalitis viral | 756 | 756 | 0 | 604 | 76 | 76 | 0.22 | 0.87 | 0.88 | 66 | 0 | 0 | 10 | 0.87 | 1.00 | 0.93 | 0.53 | 0.87 | 1.00 |

| | | | | | | | | | | | | | | | | | | | | |
|---|---|---|---|---|---|---|---|---|---|---|---|---|---|---|---|---|---|---|---|---|
| model_494 | Encephalopathy | 7763 | 7763 | 0 | 6210 | 777 | 776 | 0.27 | 0.94 | 0.93 | 730 | 0 | 0 | 47 | 0.94 | 1.00 | 0.97 | 0.24 | 0.94 | 1.00 |
| model_495 | Encephalopathy hypertensive | 492 | 588 | 96 | 470 | 59 | 59 | 0.21 | 0.95 | 0.9 | 56 | 0 | 0 | 3 | 0.95 | 1.00 | 0.97 | 0.2 | 0.95 | 1.00 |
| model_496 | Encopresis | 81 | 588 | 507 | 470 | 59 | 59 | 0.24 | 0.68 | 0.59 | 40 | 0 | 0 | 19 | 0.68 | 1.00 | 0.81 | 1.29 | 0.68 | 1.00 |
| model_497 | Endocarditis | 2798 | 2798 | 0 | 2238 | 280 | 280 | 0.24 | 0.94 | 0.92 | 262 | 0 | 0 | 18 | 0.94 | 1.00 | 0.97 | 0.26 | 0.94 | 1.00 |
| model_498 | Endocrine disorder | 3225 | 3225 | 0 | 2580 | 323 | 322 | 0.23 | 0.95 | 0.98 | 306 | 0 | 0 | 17 | 0.95 | 1.00 | 0.97 | 0.21 | 0.95 | 1.00 |
| model_499 | Endometrial Atrophy | 150 | 588 | 438 | 470 | 59 | 59 | 0.24 | 0.78 | 0.76 | 46 | 0 | 0 | 13 | 0.78 | 1.00 | 0.88 | 0.88 | 0.78 | 1.00 |
| model_500 | Endometrial cancer | 491 | 588 | 97 | 470 | 59 | 59 | 0.23 | 0.93 | 0.88 | 55 | 0 | 0 | 4 | 0.93 | 1.00 | 0.96 | 0.27 | 0.93 | 1.00 |
| model_501 | Endometrial hyperplasia | 421 | 588 | 167 | 470 | 59 | 59 | 0.23 | 0.97 | 0.93 | 57 | 0 | 0 | 2 | 0.97 | 1.00 | 0.98 | 0.14 | 0.97 | 1.00 |
| model_502 | Endometriosis | 689 | 689 | 0 | 551 | 69 | 69 | 0.23 | 0.94 | 0.93 | 65 | 0 | 0 | 4 | 0.94 | 1.00 | 0.97 | 0.23 | 0.94 | 1.00 |
| model_503 | Endometritis | 145 | 588 | 443 | 470 | 59 | 59 | 0.25 | 0.88 | 0.83 | 52 | 0 | 0 | 7 | 0.88 | 1.00 | 0.94 | 0.47 | 0.88 | 1.00 |
| model_504 | Enlarged liver | 7035 | 7035 | 0 | 5628 | 704 | 703 | 0.27 | 0.93 | 0.94 | 658 | 0 | 0 | 46 | 0.93 | 1.00 | 0.97 | 0.26 | 0.93 | 1.00 |
| model_505 | Enlarged spleen | 6246 | 6246 | 0 | 4996 | 625 | 625 | 0.27 | 0.94 | 0.95 | 586 | 0 | 0 | 39 | 0.94 | 1.00 | 0.97 | 0.25 | 0.94 | 1.00 |
| model_506 | Enteritis | 1966 | 1966 | 0 | 1572 | 197 | 197 | 0.23 | 0.93 | 0.89 | 184 | 0 | 0 | 13 | 0.93 | 1.00 | 0.97 | 0.26 | 0.93 | 1.00 |
| model_507 | Enterocolitis | 1663 | 1663 | 0 | 1330 | 167 | 166 | 0.24 | 0.93 | 0.95 | 156 | 0 | 0 | 11 | 0.93 | 1.00 | 0.97 | 0.26 | 0.93 | 1.00 |
| model_508 | Entropion | 21 | 588 | 567 | 470 | 59 | 59 | 0.24 | 0.29 | 0.27 | 17 | 0 | 0 | 42 | 0.29 | 1.00 | 0.45 | 2.85 | 0.29 | 1.00 |
| model_509 | Enuresis | 1024 | 1024 | 0 | 819 | 103 | 102 | 0.23 | 0.92 | 0.88 | 95 | 0 | 0 | 8 | 0.92 | 1.00 | 0.96 | 0.31 | 0.92 | 1.00 |
| model_510 | Eosinophil Count Increased | 4227 | 4227 | 0 | 3381 | 423 | 423 | 0.24 | 0.94 | 0.93 | 396 | 0 | 0 | 27 | 0.94 | 1.00 | 0.97 | 0.26 | 0.94 | 1.00 |
| model_511 | Eosinophilic pneumonia | 519 | 588 | 69 | 470 | 59 | 59 | 0.23 | 0.9 | 0.92 | 53 | 0 | 0 | 6 | 0.9 | 1.00 | 0.95 | 0.41 | 0.9 | 1.00 |
| model_512 | Eosinophilic pneumonia acute | 51 | 588 | 537 | 470 | 59 | 59 | 0.26 | 0.68 | 0.71 | 40 | 0 | 0 | 19 | 0.68 | 1.00 | 0.81 | 1.29 | 0.68 | 1.00 |
| model_513 | Ephelides | 108 | 588 | 480 | 470 | 59 | 59 | 0.25 | 0.78 | 0.81 | 46 | 0 | 0 | 13 | 0.78 | 1.00 | 0.88 | 0.88 | 0.78 | 1.00 |
| model_514 | Epicondylitis | 1056 | 1056 | 0 | 844 | 106 | 106 | 0.22 | 0.94 | 0.96 | 100 | 0 | 0 | 6 | 0.94 | 1.00 | 0.97 | 0.23 | 0.94 | 1.00 |
| model_515 | Epididymitis | 791 | 791 | 0 | 632 | 80 | 79 | 0.23 | 0.95 | 0.94 | 76 | 0 | 0 | 4 | 0.95 | 1.00 | 0.97 | 0.2 | 0.95 | 1.00 |
| model_516 | Epidural abscess | 840 | 840 | 0 | 672 | 84 | 84 | 0.22 | 0.9 | 0.96 | 76 | 0 | 0 | 8 | 0.9 | 1.00 | 0.95 | 0.38 | 0.9 | 1.00 |
| model_517 | Epidural hematoma | 530 | 588 | 58 | 470 | 59 | 59 | 0.22 | 0.9 | 0.95 | 53 | 0 | 0 | 6 | 0.9 | 1.00 | 0.95 | 0.41 | 0.9 | 1.00 |
| model_518 | Epiglottitis | 206 | 588 | 382 | 470 | 59 | 59 | 0.24 | 0.95 | 0.95 | 56 | 0 | 0 | 3 | 0.95 | 1.00 | 0.97 | 0.2 | 0.95 | 1.00 |
| model_519 | Epilepsy | 2896 | 2896 | 0 | 2316 | 290 | 290 | 0.24 | 0.94 | 0.93 | 274 | 0 | 0 | 16 | 0.94 | 1.00 | 0.97 | 0.22 | 0.94 | 1.00 |
| model_520 | Episcleritis | 244 | 588 | 344 | 470 | 59 | 59 | 0.24 | 0.97 | 0.97 | 57 | 0 | 0 | 2 | 0.97 | 1.00 | 0.98 | 0.14 | 0.97 | 1.00 |
| model_521 | Epistaxis | 8160 | 8160 | 0 | 6528 | 816 | 816 | 0.27 | 0.95 | 0.96 | 779 | 0 | 0 | 37 | 0.95 | 1.00 | 0.98 | 0.18 | 0.95 | 1.00 |
| model_522 | Erosive gastritis | 3363 | 3363 | 0 | 2690 | 337 | 336 | 0.24 | 0.94 | 0.94 | 318 | 0 | 0 | 19 | 0.94 | 1.00 | 0.97 | 0.23 | 0.94 | 1.00 |
| model_523 | Eruption | 15526 | 15526 | 0 | 12420 | 1553 | 1553 | 0.31 | 0.95 | 0.96 | 1483 | 0 | 0 | 70 | 0.95 | 1.00 | 0.98 | 0.18 | 0.95 | 1.00 |
| model_524 | Erysipelas | 1072 | 1072 | 0 | 857 | 108 | 107 | 0.23 | 0.92 | 0.94 | 99 | 0 | 0 | 9 | 0.92 | 1.00 | 0.96 | 0.33 | 0.92 | 1.00 |
| model_525 | Erythema | 13598 | 13598 | 0 | 10878 | 1360 | 1360 | 0.28 | 0.95 | 0.96 | 1295 | 0 | 0 | 65 | 0.95 | 1.00 | 0.98 | 0.19 | 0.95 | 1.00 |
| model_526 | Erythema multiforme | 2764 | 2764 | 0 | 2211 | 277 | 276 | 0.24 | 0.93 | 0.92 | 258 | 0 | 0 | 19 | 0.93 | 1.00 | 0.96 | 0.27 | 0.93 | 1.00 |
| model_527 | Erythema nodosum | 483 | 588 | 105 | 470 | 59 | 59 | 0.22 | 1 | 0.95 | 59 | 0 | 0 | 0 | 1 | 1.00 | 1 | 0 | 1 | 1.00 |
| model_528 | Erythremia | 66 | 588 | 522 | 470 | 59 | 59 | 0.25 | 0.73 | 0.8 | 43 | 0 | 0 | 16 | 0.73 | 1.00 | 0.84 | 1.08 | 0.73 | 1.00 |
| model_529 | Esophageal cancer | 1123 | 1123 | 0 | 898 | 113 | 112 | 0.23 | 0.88 | 0.94 | 99 | 0 | 0 | 14 | 0.88 | 1.00 | 0.93 | 0.5 | 0.88 | 1.00 |
| model_530 | Esophageal perforation | 157 | 588 | 431 | 470 | 59 | 59 | 0.24 | 0.92 | 0.81 | 54 | 0 | 0 | 5 | 0.92 | 1.00 | 0.96 | 0.34 | 0.92 | 1.00 |
| model_531 | Esophageal rupture | 457 | 588 | 131 | 470 | 59 | 59 | 0.23 | 0.81 | 0.93 | 48 | 0 | 0 | 11 | 0.81 | 1.00 | 0.9 | 0.75 | 0.81 | 1.00 |
| model_532 | Esophageal spasm | 1967 | 1967 | 0 | 1573 | 197 | 197 | 0.25 | 0.95 | 0.98 | 187 | 0 | 0 | 10 | 0.95 | 1.00 | 0.97 | 0.2 | 0.95 | 1.00 |
| model_533 | Esophageal stenosis | 1503 | 1503 | 0 | 1202 | 151 | 150 | 0.23 | 0.95 | 0.95 | 144 | 0 | 0 | 7 | 0.95 | 1.00 | 0.98 | 0.19 | 0.95 | 1.00 |
| model_534 | Esophageal ulcer | 2466 | 2466 | 0 | 1972 | 247 | 247 | 0.25 | 0.95 | 0.96 | 235 | 0 | 0 | 12 | 0.95 | 1.00 | 0.98 | 0.19 | 0.95 | 1.00 |
| model_535 | Esophagitis | 9148 | 9148 | 0 | 7318 | 915 | 915 | 0.27 | 0.96 | 0.95 | 880 | 0 | 0 | 35 | 0.96 | 1.00 | 0.98 | 0.15 | 0.96 | 1.00 |
| model_536 | Essential Thrombocythemia | 133 | 588 | 455 | 470 | 59 | 59 | 0.25 | 0.9 | 0.92 | 53 | 0 | 0 | 6 | 0.9 | 1.00 | 0.95 | 0.41 | 0.9 | 1.00 |
| model_537 | Essential hypertension | 2196 | 2196 | 0 | 1756 | 220 | 220 | 0.23 | 0.97 | 0.93 | 214 | 0 | 0 | 6 | 0.97 | 1.00 | 0.99 | 0.11 | 0.97 | 1.00 |
| model_538 | Estrogen replacement | 2 | 588 | 586 | 470 | 59 | 59 | 0.23 | 0 | 0 | 0 | 0 | 0 | 59 | 0 | 0.00 | 0 | 4 | 0 | 1.00 |
| model_539 | Eustachian tube disorder | 803 | 803 | 0 | 642 | 81 | 80 | 0.23 | 0.91 | 0.93 | 74 | 0 | 0 | 7 | 0.91 | 1.00 | 0.95 | 0.35 | 0.91 | 1.00 |
| model_540 | Eustachian tube obstruction | 39 | 588 | 549 | 470 | 59 | 59 | 0.26 | 0.39 | 0.36 | 23 | 0 | 0 | 36 | 0.39 | 1.00 | 0.56 | 2.44 | 0.39 | 1.00 |
| model_541 | Excess potassium | 8902 | 8902 | 0 | 7121 | 891 | 890 | 0.27 | 0.94 | 0.94 | 840 | 0 | 0 | 51 | 0.94 | 1.00 | 0.97 | 0.23 | 0.94 | 1.00 |
| model_542 | Excessive salivation | 2205 | 2205 | 0 | 1764 | 221 | 220 | 0.24 | 0.92 | 0.95 | 203 | 0 | 0 | 18 | 0.92 | 1.00 | 0.96 | 0.33 | 0.92 | 1.00 |
| model_543 | Excessive sleepiness | 5351 | 5351 | 0 | 4280 | 536 | 535 | 0.25 | 0.95 | 0.97 | 511 | 0 | 0 | 25 | 0.95 | 1.00 | 0.98 | 0.19 | 0.95 | 1.00 |

| | | | | | | | | | | | | | | | | | | | | |
|---|---|---|---|---|---|---|---|---|---|---|---|---|---|---|---|---|---|---|---|---|
| model_544 | Excessive thirst | 5934 | 5934 | 0 | 4747 | 594 | 593 | 0.26 | 0.96 | 0.95 | 570 | 0 | 0 | 24 | 0.96 | 1.00 | 0.98 | 0.16 | 0.96 | 1.00 |
| model_545 | Excoriation | 4410 | 4410 | 0 | 3528 | 441 | 441 | 0.26 | 0.95 | 0.97 | 420 | 0 | 0 | 21 | 0.95 | 1.00 | 0.98 | 0.19 | 0.95 | 1.00 |
| model_546 | Exophthalmos | 551 | 588 | 37 | 470 | 59 | 59 | 0.23 | 0.93 | 0.93 | 55 | 0 | 0 | 4 | 0.93 | 1.00 | 0.96 | 0.27 | 0.93 | 1.00 |
| model_547 | Exploratory laparotomy | 128 | 588 | 460 | 470 | 59 | 59 | 0.25 | 0.69 | 0.81 | 41 | 0 | 0 | 18 | 0.69 | 1.00 | 0.82 | 1.22 | 0.69 | 1.00 |
| model_548 | External ear infection | 1840 | 1840 | 0 | 1472 | 184 | 184 | 0.23 | 0.92 | 0.97 | 170 | 0 | 0 | 14 | 0.92 | 1.00 | 0.96 | 0.3 | 0.92 | 1.00 |
| model_549 | Extrasystoles Ventricular | 6627 | 6627 | 0 | 5301 | 663 | 663 | 0.25 | 0.95 | 0.95 | 633 | 0 | 0 | 30 | 0.95 | 1.00 | 0.98 | 0.18 | 0.95 | 1.00 |
| model_550 | Extremity pain | 17808 | 17808 | 0 | 14246 | 1781 | 1781 | 0.32 | 0.96 | 0.95 | 1714 | 0 | 0 | 67 | 0.96 | 1.00 | 0.98 | 0.15 | 0.96 | 1.00 |
| model_551 | Eye infection | 2539 | 2539 | 0 | 2031 | 254 | 254 | 0.24 | 0.98 | 0.96 | 248 | 0 | 0 | 6 | 0.98 | 1.00 | 0.99 | 0.09 | 0.98 | 1.00 |
| model_552 | Eye injury | 1147 | 1147 | 0 | 917 | 115 | 115 | 0.23 | 0.99 | 0.89 | 114 | 0 | 0 | 1 | 0.99 | 1.00 | 1 | 0.03 | 0.99 | 1.00 |
| model_553 | Eye pain | 4673 | 4673 | 0 | 3738 | 468 | 467 | 0.26 | 0.96 | 0.96 | 447 | 0 | 0 | 21 | 0.96 | 1.00 | 0.98 | 0.18 | 0.96 | 1.00 |
| model_554 | Eye redness | 437 | 588 | 151 | 470 | 59 | 59 | 0.23 | 0.97 | 0.97 | 57 | 0 | 0 | 2 | 0.97 | 1.00 | 0.98 | 0.14 | 0.97 | 1.00 |
| model_555 | Eye swelling | 3792 | 3792 | 0 | 3033 | 380 | 379 | 0.25 | 0.94 | 0.94 | 358 | 0 | 0 | 22 | 0.94 | 1.00 | 0.97 | 0.23 | 0.94 | 1.00 |
| model_556 | Eyelid diseases | 1226 | 1226 | 0 | 980 | 123 | 123 | 0.23 | 0.94 | 0.94 | 116 | 0 | 0 | 7 | 0.94 | 1.00 | 0.97 | 0.23 | 0.94 | 1.00 |
| model_557 | Face pain | 3528 | 3528 | 0 | 2822 | 353 | 353 | 0.25 | 0.97 | 0.93 | 342 | 0 | 0 | 11 | 0.97 | 1.00 | 0.98 | 0.12 | 0.97 | 1.00 |
| model_558 | Facial flushing | 10025 | 10025 | 0 | 8020 | 1003 | 1002 | 0.29 | 0.96 | 0.97 | 961 | 0 | 0 | 42 | 0.96 | 1.00 | 0.98 | 0.17 | 0.96 | 1.00 |
| model_559 | Facial palsy | 4409 | 4409 | 0 | 3527 | 441 | 441 | 0.25 | 0.95 | 0.93 | 417 | 0 | 0 | 24 | 0.95 | 1.00 | 0.97 | 0.22 | 0.95 | 1.00 |
| model_560 | Faecal impaction | 42 | 588 | 546 | 470 | 59 | 59 | 0.25 | 0.63 | 0.46 | 37 | 0 | 0 | 22 | 0.63 | 1.00 | 0.77 | 1.49 | 0.63 | 1.00 |
| model_561 | Faecal incontinence | 5050 | 5050 | 0 | 4040 | 505 | 505 | 0.26 | 0.94 | 0.95 | 473 | 0 | 0 | 32 | 0.94 | 1.00 | 0.97 | 0.25 | 0.94 | 1.00 |
| model_562 | Failure to thrive | 3342 | 3342 | 0 | 2673 | 335 | 334 | 0.25 | 0.95 | 0.93 | 317 | 0 | 0 | 18 | 0.95 | 1.00 | 0.97 | 0.21 | 0.95 | 1.00 |
| model_563 | Fainting | 13728 | 13728 | 0 | 10982 | 1373 | 1373 | 0.3 | 0.96 | 0.96 | 1319 | 0 | 0 | 54 | 0.96 | 1.00 | 0.98 | 0.16 | 0.96 | 1.00 |
| model_564 | Familial adenomatous polyposis | 221 | 588 | 367 | 470 | 59 | 59 | 0.24 | 0.9 | 0.9 | 53 | 0 | 0 | 6 | 0.9 | 1.00 | 0.95 | 0.41 | 0.9 | 1.00 |
| model_565 | Familial hypercholesterolemia | 81 | 588 | 507 | 470 | 59 | 59 | 0.26 | 0.51 | 0.54 | 30 | 0 | 0 | 29 | 0.51 | 1.00 | 0.67 | 1.97 | 0.51 | 1.00 |
| model_566 | Fasciitis | 526 | 588 | 62 | 470 | 59 | 59 | 0.23 | 0.88 | 0.95 | 52 | 0 | 0 | 7 | 0.88 | 1.00 | 0.94 | 0.47 | 0.88 | 1.00 |
| model_567 | Fascitis plantar | 2134 | 2134 | 0 | 1707 | 214 | 213 | 0.23 | 0.94 | 0.97 | 202 | 0 | 0 | 12 | 0.94 | 1.00 | 0.97 | 0.22 | 0.94 | 1.00 |
| model_568 | Fat embolism | 392 | 588 | 196 | 470 | 59 | 59 | 0.23 | 0.95 | 0.83 | 56 | 0 | 0 | 3 | 0.95 | 1.00 | 0.97 | 0.2 | 0.95 | 1.00 |
| model_569 | Fatigue | 24260 | 24260 | 0 | 19408 | 2426 | 2426 | 0.36 | 0.96 | 0.96 | 2332 | 0 | 0 | 94 | 0.96 | 1.00 | 0.98 | 0.15 | 0.96 | 1.00 |
| model_570 | Fatty liver | 8827 | 8827 | 0 | 7061 | 883 | 883 | 0.28 | 0.95 | 0.95 | 842 | 0 | 0 | 41 | 0.95 | 1.00 | 0.98 | 0.19 | 0.95 | 1.00 |
| model_571 | Fatty stools | 273 | 588 | 315 | 470 | 59 | 59 | 0.24 | 0.93 | 0.93 | 55 | 0 | 0 | 4 | 0.93 | 1.00 | 0.96 | 0.27 | 0.93 | 1.00 |
| model_572 | Febrile convulsion | 198 | 588 | 390 | 470 | 59 | 59 | 0.25 | 0.9 | 0.86 | 53 | 0 | 0 | 6 | 0.9 | 1.00 | 0.95 | 0.41 | 0.9 | 1.00 |
| model_573 | Fecal occult blood | 2142 | 2142 | 0 | 1713 | 215 | 214 | 0.26 | 0.93 | 0.93 | 201 | 0 | 0 | 14 | 0.93 | 1.00 | 0.97 | 0.26 | 0.93 | 1.00 |
| model_574 | Feeling unwell | 14593 | 14593 | 0 | 11674 | 1460 | 1459 | 0.33 | 0.96 | 0.94 | 1402 | 0 | 0 | 58 | 0.96 | 1.00 | 0.98 | 0.16 | 0.96 | 1.00 |
| model_575 | Femoral neck fracture | 1359 | 1359 | 0 | 1087 | 136 | 136 | 0.25 | 0.96 | 0.94 | 131 | 0 | 0 | 5 | 0.96 | 1.00 | 0.98 | 0.15 | 0.96 | 1.00 |
| model_576 | Femur fracture | 2783 | 2783 | 0 | 2226 | 279 | 278 | 0.24 | 0.96 | 0.96 | 268 | 0 | 0 | 11 | 0.96 | 1.00 | 0.98 | 0.16 | 0.96 | 1.00 |
| model_577 | Fib | 3292 | 3292 | 0 | 2633 | 330 | 329 | 0.26 | 0.95 | 0.96 | 315 | 0 | 0 | 15 | 0.95 | 1.00 | 0.98 | 0.18 | 0.95 | 1.00 |
| model_578 | Fibroadenoma of breast | 231 | 588 | 357 | 470 | 59 | 59 | 0.24 | 0.83 | 0.85 | 49 | 0 | 0 | 10 | 0.83 | 1.00 | 0.91 | 0.68 | 0.83 | 1.00 |
| model_579 | Fibroids | 3209 | 3209 | 0 | 2567 | 321 | 321 | 0.25 | 0.95 | 0.95 | 304 | 0 | 0 | 17 | 0.95 | 1.00 | 0.97 | 0.21 | 0.95 | 1.00 |
| model_580 | Fibromyalgia | 7759 | 7759 | 0 | 6207 | 776 | 776 | 0.26 | 0.97 | 0.97 | 752 | 0 | 0 | 24 | 0.97 | 1.00 | 0.98 | 0.12 | 0.97 | 1.00 |
| model_581 | Fibrosing alveolitis | 646 | 646 | 0 | 516 | 65 | 65 | 0.24 | 0.95 | 0.97 | 62 | 0 | 0 | 3 | 0.95 | 1.00 | 0.98 | 0.18 | 0.95 | 1.00 |
| model_582 | Fistula | 3893 | 3893 | 0 | 3114 | 390 | 389 | 0.27 | 0.94 | 0.94 | 368 | 0 | 0 | 22 | 0.94 | 1.00 | 0.97 | 0.23 | 0.94 | 1.00 |
| model_583 | Flank pain | 4663 | 4663 | 0 | 3730 | 467 | 466 | 0.26 | 0.94 | 0.97 | 441 | 0 | 0 | 26 | 0.94 | 1.00 | 0.97 | 0.22 | 0.94 | 1.00 |
| model_584 | Flashbacks | 351 | 588 | 237 | 470 | 59 | 59 | 0.23 | 0.9 | 0.93 | 53 | 0 | 0 | 6 | 0.9 | 1.00 | 0.95 | 0.41 | 0.9 | 1.00 |
| model_585 | Flashing lights | 1581 | 1581 | 0 | 1264 | 159 | 158 | 0.24 | 0.99 | 0.97 | 157 | 0 | 0 | 2 | 0.99 | 1.00 | 0.99 | 0.05 | 0.99 | 1.00 |
| model_586 | Flatulence | 7915 | 7915 | 0 | 6332 | 792 | 791 | 0.28 | 0.96 | 0.95 | 760 | 0 | 0 | 32 | 0.96 | 1.00 | 0.98 | 0.16 | 0.96 | 1.00 |
| model_587 | Floaters | 1964 | 1964 | 0 | 1571 | 197 | 196 | 0.24 | 0.96 | 0.96 | 190 | 0 | 0 | 7 | 0.96 | 1.00 | 0.98 | 0.14 | 0.96 | 1.00 |
| model_588 | Flu | 6598 | 6598 | 0 | 5278 | 660 | 660 | 0.26 | 0.95 | 0.96 | 627 | 0 | 0 | 33 | 0.95 | 1.00 | 0.97 | 0.2 | 0.95 | 1.00 |
| model_589 | Folate deficiency | 602 | 602 | 0 | 481 | 61 | 60 | 0.22 | 0.93 | 0.95 | 57 | 0 | 0 | 4 | 0.93 | 1.00 | 0.97 | 0.26 | 0.93 | 1.00 |
| model_590 | Folliculitis | 1186 | 1186 | 0 | 948 | 119 | 119 | 0.24 | 0.96 | 0.92 | 114 | 0 | 0 | 5 | 0.96 | 1.00 | 0.98 | 0.17 | 0.96 | 1.00 |
| model_591 | Food allergy | 462 | 588 | 126 | 470 | 59 | 59 | 0.22 | 0.95 | 0.98 | 56 | 0 | 0 | 3 | 0.95 | 1.00 | 0.97 | 0.2 | 0.95 | 1.00 |
| model_592 | Food intolerance | 1076 | 1076 | 0 | 860 | 108 | 108 | 0.22 | 0.95 | 0.93 | 103 | 0 | 0 | 5 | 0.95 | 1.00 | 0.98 | 0.19 | 0.95 | 1.00 |
| model_593 | Food poisoning | 650 | 650 | 0 | 520 | 65 | 65 | 0.23 | 0.95 | 0.94 | 62 | 0 | 0 | 3 | 0.95 | 1.00 | 0.98 | 0.18 | 0.95 | 1.00 |

| | | | | | | | | | | | | | | | | | | | | |
|---|---|---|---|---|---|---|---|---|---|---|---|---|---|---|---|---|---|---|---|---|
| model_594 | Foreign body in eye | 306 | 588 | 282 | 470 | 59 | 59 | 0.23 | 0.97 | 0.93 | 57 | 0 | 0 | 2 | 0.97 | 1.00 | 0.98 | 0.14 | 0.97 | 1.00 |
| model_595 | Fracture nonunion | 600 | 600 | 0 | 480 | 60 | 60 | 0.23 | 0.97 | 0.93 | 58 | 0 | 0 | 2 | 0.97 | 1.00 | 0.98 | 0.13 | 0.97 | 1.00 |
| model_596 | Fractured pelvis NOS | 2063 | 2063 | 0 | 1650 | 207 | 206 | 0.24 | 0.94 | 0.96 | 195 | 0 | 0 | 12 | 0.94 | 1.00 | 0.97 | 0.23 | 0.94 | 1.00 |
| model_597 | Fungal disease | 8455 | 8455 | 0 | 6764 | 846 | 845 | 0.28 | 0.94 | 0.95 | 795 | 0 | 0 | 51 | 0.94 | 1.00 | 0.97 | 0.24 | 0.94 | 1.00 |
| model_598 | Furuncle | 1877 | 1877 | 0 | 1501 | 188 | 188 | 0.24 | 0.95 | 0.93 | 179 | 0 | 0 | 9 | 0.95 | 1.00 | 0.98 | 0.19 | 0.95 | 1.00 |
| model_599 | Galactorrhea | 801 | 801 | 0 | 640 | 81 | 80 | 0.24 | 0.94 | 0.94 | 76 | 0 | 0 | 5 | 0.94 | 1.00 | 0.97 | 0.25 | 0.94 | 1.00 |
| model_600 | Gall bladder | 6859 | 6859 | 0 | 5487 | 686 | 686 | 0.27 | 0.96 | 0.95 | 657 | 0 | 0 | 29 | 0.96 | 1.00 | 0.98 | 0.17 | 0.96 | 1.00 |
| model_601 | Gallbladder cancer | 247 | 588 | 341 | 470 | 59 | 59 | 0.24 | 0.85 | 0.88 | 50 | 0 | 0 | 9 | 0.85 | 1.00 | 0.92 | 0.61 | 0.85 | 1.00 |
| model_602 | Gallbladder perforation | 252 | 588 | 336 | 470 | 59 | 59 | 0.23 | 0.86 | 0.92 | 51 | 0 | 0 | 8 | 0.86 | 1.00 | 0.93 | 0.54 | 0.86 | 1.00 |
| model_603 | Gallbladder polyp | 415 | 588 | 173 | 470 | 59 | 59 | 0.23 | 0.97 | 0.9 | 57 | 0 | 0 | 2 | 0.97 | 1.00 | 0.98 | 0.14 | 0.97 | 1.00 |
| model_604 | Ganglion | 694 | 694 | 0 | 555 | 70 | 69 | 0.23 | 0.91 | 0.91 | 64 | 0 | 0 | 6 | 0.91 | 1.00 | 0.96 | 0.34 | 0.91 | 1.00 |
| model_605 | Gangrene | 2715 | 2715 | 0 | 2172 | 272 | 271 | 0.23 | 0.93 | 0.97 | 254 | 0 | 0 | 18 | 0.93 | 1.00 | 0.97 | 0.26 | 0.93 | 1.00 |
| model_606 | Gastric Cancer | 1224 | 1224 | 0 | 979 | 123 | 122 | 0.24 | 0.93 | 0.96 | 114 | 0 | 0 | 9 | 0.93 | 1.00 | 0.96 | 0.29 | 0.93 | 1.00 |
| model_607 | Gastric dilatation | 352 | 588 | 236 | 470 | 59 | 59 | 0.24 | 0.92 | 0.92 | 54 | 0 | 0 | 5 | 0.92 | 1.00 | 0.96 | 0.34 | 0.92 | 1.00 |
| model_608 | Gastric inflammation | 11070 | 11070 | 0 | 8856 | 1107 | 1107 | 0.29 | 0.95 | 0.96 | 1049 | 0 | 0 | 58 | 0.95 | 1.00 | 0.97 | 0.21 | 0.95 | 1.00 |
| model_609 | Gastric polyps | 1691 | 1691 | 0 | 1352 | 170 | 169 | 0.24 | 0.96 | 0.97 | 164 | 0 | 0 | 6 | 0.96 | 1.00 | 0.98 | 0.14 | 0.96 | 1.00 |
| model_610 | Gastric stasis | 89 | 588 | 499 | 470 | 59 | 59 | 0.25 | 0.76 | 0.75 | 45 | 0 | 0 | 14 | 0.76 | 1.00 | 0.87 | 0.95 | 0.76 | 1.00 |
| model_611 | Gastric ulcer | 6345 | 6345 | 0 | 5076 | 635 | 634 | 0.27 | 0.96 | 0.95 | 608 | 0 | 0 | 27 | 0.96 | 1.00 | 0.98 | 0.17 | 0.96 | 1.00 |
| model_612 | Gastric ulcer perforated | 715 | 715 | 0 | 572 | 72 | 71 | 0.22 | 0.99 | 0.93 | 71 | 0 | 0 | 1 | 0.99 | 1.00 | 0.99 | 0.06 | 0.99 | 1.00 |
| model_613 | Gastroduodenal ulcer | 4048 | 4048 | 0 | 3238 | 405 | 405 | 0.25 | 0.94 | 0.97 | 382 | 0 | 0 | 23 | 0.94 | 1.00 | 0.97 | 0.23 | 0.94 | 1.00 |
| model_614 | Gastroenteritis | 7307 | 7307 | 0 | 5845 | 731 | 731 | 0.28 | 0.94 | 0.95 | 690 | 0 | 0 | 41 | 0.94 | 1.00 | 0.97 | 0.22 | 0.94 | 1.00 |
| model_615 | Gastroenteritis viral | 3816 | 3816 | 0 | 3052 | 382 | 382 | 0.25 | 0.92 | 0.95 | 353 | 0 | 0 | 29 | 0.92 | 1.00 | 0.96 | 0.3 | 0.92 | 1.00 |
| model_616 | Gastrointestinal Obstruction | 460 | 588 | 128 | 470 | 59 | 59 | 0.22 | 0.92 | 0.98 | 54 | 0 | 0 | 5 | 0.92 | 1.00 | 0.96 | 0.34 | 0.92 | 1.00 |
| model_617 | Gastrointestinal bleed | 11775 | 11775 | 0 | 9420 | 1178 | 1177 | 0.29 | 0.93 | 0.95 | 1098 | 0 | 0 | 80 | 0.93 | 1.00 | 0.96 | 0.27 | 0.93 | 1.00 |
| model_618 | Genital herpes | 108 | 588 | 480 | 470 | 59 | 59 | 0.25 | 0.78 | 0.78 | 46 | 0 | 0 | 13 | 0.78 | 1.00 | 0.88 | 0.88 | 0.78 | 1.00 |
| model_619 | Gestational diabetes | 420 | 588 | 168 | 470 | 59 | 59 | 0.22 | 0.98 | 0.97 | 58 | 0 | 0 | 1 | 0.98 | 1.00 | 0.99 | 0.07 | 0.98 | 1.00 |
| model_620 | Giardiasis | 325 | 588 | 263 | 470 | 59 | 59 | 0.23 | 0.93 | 0.9 | 55 | 0 | 0 | 4 | 0.93 | 1.00 | 0.96 | 0.27 | 0.93 | 1.00 |
| model_621 | Gingival pain | 2835 | 2835 | 0 | 2268 | 284 | 283 | 0.23 | 0.95 | 0.94 | 270 | 0 | 0 | 14 | 0.95 | 1.00 | 0.97 | 0.2 | 0.95 | 1.00 |
| model_622 | Gingivitis | 5660 | 5660 | 0 | 4528 | 566 | 566 | 0.26 | 0.95 | 0.94 | 538 | 0 | 0 | 28 | 0.95 | 1.00 | 0.97 | 0.2 | 0.95 | 1.00 |
| model_623 | Glaucoma | 5736 | 5736 | 0 | 4588 | 574 | 574 | 0.24 | 0.95 | 0.97 | 546 | 0 | 0 | 28 | 0.95 | 1.00 | 0.98 | 0.2 | 0.95 | 1.00 |
| model_624 | Glioblastoma multiforme | 633 | 633 | 0 | 506 | 64 | 63 | 0.25 | 0.92 | 0.84 | 59 | 0 | 0 | 5 | 0.92 | 1.00 | 0.96 | 0.31 | 0.92 | 1.00 |
| model_625 | Glioma | 249 | 588 | 339 | 470 | 59 | 59 | 0.23 | 0.9 | 0.88 | 53 | 0 | 0 | 6 | 0.9 | 1.00 | 0.95 | 0.41 | 0.9 | 1.00 |
| model_626 | Global amnesia | 293 | 588 | 295 | 470 | 59 | 59 | 0.23 | 1 | 0.93 | 59 | 0 | 0 | 0 | 1 | 1.00 | 1 | 0 | 1 | 1.00 |
| model_627 | Glomerulonephritis | 1310 | 1310 | 0 | 1048 | 131 | 131 | 0.22 | 0.91 | 0.93 | 119 | 0 | 0 | 12 | 0.91 | 1.00 | 0.95 | 0.37 | 0.91 | 1.00 |
| model_628 | Glossitis | 1555 | 1555 | 0 | 1244 | 156 | 155 | 0.24 | 0.96 | 0.93 | 150 | 0 | 0 | 6 | 0.96 | 1.00 | 0.98 | 0.15 | 0.96 | 1.00 |
| model_629 | Glossodynia | 4102 | 4102 | 0 | 3281 | 411 | 410 | 0.25 | 0.98 | 0.97 | 401 | 0 | 0 | 10 | 0.98 | 1.00 | 0.99 | 0.1 | 0.98 | 1.00 |
| model_630 | Glucose intolerance | 2107 | 2107 | 0 | 1685 | 211 | 211 | 0.23 | 0.97 | 0.93 | 204 | 0 | 0 | 7 | 0.97 | 1.00 | 0.98 | 0.13 | 0.97 | 1.00 |
| model_631 | Glucosuria | 2894 | 2894 | 0 | 2315 | 290 | 289 | 0.24 | 0.96 | 0.93 | 279 | 0 | 0 | 11 | 0.96 | 1.00 | 0.98 | 0.15 | 0.96 | 1.00 |
| model_632 | Goiter | 3265 | 3265 | 0 | 2612 | 327 | 326 | 0.25 | 0.94 | 0.97 | 306 | 0 | 0 | 21 | 0.94 | 1.00 | 0.97 | 0.26 | 0.94 | 1.00 |
| model_633 | Gonorrhea | 49 | 588 | 539 | 470 | 59 | 59 | 0.26 | 0.27 | 0.27 | 16 | 0 | 0 | 43 | 0.27 | 1.00 | 0.43 | 2.92 | 0.27 | 1.00 |
| model_634 | Gout | 4699 | 4699 | 0 | 3759 | 470 | 470 | 0.26 | 0.94 | 0.96 | 441 | 0 | 0 | 29 | 0.94 | 1.00 | 0.97 | 0.25 | 0.94 | 1.00 |
| model_635 | Gouty arthritis | 970 | 970 | 0 | 776 | 97 | 97 | 0.24 | 0.9 | 0.98 | 87 | 0 | 0 | 10 | 0.9 | 1.00 | 0.95 | 0.41 | 0.9 | 1.00 |
| model_636 | Granuloma | 3787 | 3787 | 0 | 3029 | 379 | 379 | 0.24 | 0.97 | 0.96 | 366 | 0 | 0 | 13 | 0.97 | 1.00 | 0.98 | 0.14 | 0.97 | 1.00 |
| model_637 | Gynaecomastia | 2227 | 2227 | 0 | 1781 | 223 | 223 | 0.24 | 0.95 | 0.96 | 212 | 0 | 0 | 11 | 0.95 | 1.00 | 0.97 | 0.2 | 0.95 | 1.00 |
| model_638 | HIV disease | 593 | 593 | 0 | 474 | 60 | 59 | 0.22 | 0.93 | 0.88 | 56 | 0 | 0 | 4 | 0.93 | 1.00 | 0.97 | 0.27 | 0.93 | 1.00 |
| model_639 | HIV positive | 143 | 588 | 445 | 470 | 59 | 59 | 0.24 | 0.8 | 0.88 | 47 | 0 | 0 | 12 | 0.8 | 1.00 | 0.89 | 0.81 | 0.8 | 1.00 |
| model_640 | Haemangioma | 1536 | 1536 | 0 | 1228 | 154 | 154 | 0.23 | 0.94 | 0.95 | 145 | 0 | 0 | 9 | 0.94 | 1.00 | 0.97 | 0.23 | 0.94 | 1.00 |
| model_641 | Haemarthrosis | 722 | 722 | 0 | 577 | 73 | 72 | 0.24 | 0.95 | 0.93 | 69 | 0 | 0 | 4 | 0.95 | 1.00 | 0.97 | 0.22 | 0.95 | 1.00 |
| model_642 | Haematemesis | 6795 | 6795 | 0 | 5436 | 680 | 679 | 0.28 | 0.95 | 0.94 | 649 | 0 | 0 | 31 | 0.95 | 1.00 | 0.98 | 0.18 | 0.95 | 1.00 |
| model_643 | Haematochezia | 8355 | 8355 | 0 | 6684 | 836 | 835 | 0.26 | 0.95 | 0.95 | 794 | 0 | 0 | 42 | 0.95 | 1.00 | 0.97 | 0.2 | 0.95 | 1.00 |

| | | | | | | | | | | | | | | | | | | | | |
|---|---|---|---|---|---|---|---|---|---|---|---|---|---|---|---|---|---|---|---|---|
| model_644 | Haematoma | 6526 | 6526 | 0 | 5220 | 653 | 653 | 0.27 | 0.94 | 0.95 | 611 | 0 | 0 | 42 | 0.94 | 1.00 | 0.97 | 0.26 | 0.94 | 1.00 |
| model_645 | Haemolytic anaemia | 2725 | 2725 | 0 | 2180 | 273 | 272 | 0.25 | 0.93 | 0.92 | 254 | 0 | 0 | 19 | 0.93 | 1.00 | 0.96 | 0.28 | 0.93 | 1.00 |
| model_646 | Haemophilia | 17 | 588 | 571 | 470 | 59 | 59 | 0.24 | 0 | 0 | 0 | 0 | 0 | 59 | 0 | 0.00 | 0 | 4 | 0 | 1.00 |
| model_647 | Haemorrhage intracranial | 2730 | 2730 | 0 | 2184 | 273 | 273 | 0.25 | 0.92 | 0.95 | 252 | 0 | 0 | 21 | 0.92 | 1.00 | 0.96 | 0.31 | 0.92 | 1.00 |
| model_648 | Haemorrhage rectum | 9212 | 9212 | 0 | 7369 | 922 | 921 | 0.28 | 0.95 | 0.95 | 880 | 0 | 0 | 42 | 0.95 | 1.00 | 0.98 | 0.18 | 0.95 | 1.00 |
| model_649 | Haemorrhoids | 7690 | 7690 | 0 | 6152 | 769 | 769 | 0.28 | 0.96 | 0.97 | 742 | 0 | 0 | 27 | 0.96 | 1.00 | 0.98 | 0.14 | 0.96 | 1.00 |
| model_650 | Hair disease | 810 | 810 | 0 | 648 | 81 | 81 | 0.23 | 0.99 | 0.96 | 80 | 0 | 0 | 1 | 0.99 | 1.00 | 0.99 | 0.05 | 0.99 | 1.00 |
| model_651 | Hallucination | 12518 | 12518 | 0 | 10014 | 1252 | 1252 | 0.29 | 0.96 | 0.96 | 1205 | 0 | 0 | 47 | 0.96 | 1.00 | 0.98 | 0.15 | 0.96 | 1.00 |
| model_652 | Hay fever | 1473 | 1473 | 0 | 1178 | 148 | 147 | 0.24 | 0.98 | 0.98 | 145 | 0 | 0 | 3 | 0.98 | 1.00 | 0.99 | 0.08 | 0.98 | 1.00 |
| model_653 | Head ache | 19803 | 19803 | 0 | 15842 | 1981 | 1980 | 0.33 | 0.97 | 0.95 | 1914 | 0 | 0 | 67 | 0.97 | 1.00 | 0.98 | 0.14 | 0.97 | 1.00 |
| model_654 | Head injury | 6660 | 6660 | 0 | 5328 | 666 | 666 | 0.27 | 0.96 | 0.95 | 641 | 0 | 0 | 25 | 0.96 | 1.00 | 0.98 | 0.15 | 0.96 | 1.00 |
| model_655 | Heart attack | 16371 | 16371 | 0 | 13096 | 1638 | 1637 | 0.33 | 0.97 | 0.96 | 1583 | 0 | 0 | 55 | 0.97 | 1.00 | 0.98 | 0.13 | 0.97 | 1.00 |
| model_656 | Heart rate increased | 17567 | 17567 | 0 | 14053 | 1757 | 1757 | 0.32 | 0.96 | 0.94 | 1682 | 0 | 0 | 75 | 0.96 | 1.00 | 0.98 | 0.17 | 0.96 | 1.00 |
| model_657 | Heat rash | 338 | 588 | 250 | 470 | 59 | 59 | 0.23 | 0.97 | 0.95 | 57 | 0 | 0 | 2 | 0.97 | 1.00 | 0.98 | 0.14 | 0.97 | 1.00 |
| model_658 | Heat stroke | 476 | 588 | 112 | 470 | 59 | 59 | 0.23 | 0.95 | 0.88 | 56 | 0 | 0 | 3 | 0.95 | 1.00 | 0.97 | 0.2 | 0.95 | 1.00 |
| model_659 | Heavy menstrual period | 2246 | 2246 | 0 | 1796 | 225 | 225 | 0.24 | 0.92 | 0.96 | 208 | 0 | 0 | 17 | 0.92 | 1.00 | 0.96 | 0.3 | 0.92 | 1.00 |
| model_660 | Hematoma subdural | 3156 | 3156 | 0 | 2524 | 316 | 316 | 0.25 | 0.96 | 0.94 | 304 | 0 | 0 | 12 | 0.96 | 1.00 | 0.98 | 0.15 | 0.96 | 1.00 |
| model_661 | Hematospermia | 155 | 588 | 433 | 470 | 59 | 59 | 0.26 | 0.92 | 0.88 | 54 | 0 | 0 | 5 | 0.92 | 1.00 | 0.96 | 0.34 | 0.92 | 1.00 |
| model_662 | Hemiparesis | 6609 | 6609 | 0 | 5287 | 661 | 661 | 0.27 | 0.95 | 0.95 | 625 | 0 | 0 | 36 | 0.95 | 1.00 | 0.97 | 0.22 | 0.95 | 1.00 |
| model_663 | Hemiplegia | 3268 | 3268 | 0 | 2614 | 327 | 327 | 0.25 | 0.93 | 0.95 | 304 | 0 | 0 | 23 | 0.93 | 1.00 | 0.96 | 0.28 | 0.93 | 1.00 |
| model_664 | Hemoglobinuria | 103 | 588 | 485 | 470 | 59 | 59 | 0.23 | 0.78 | 0.85 | 46 | 0 | 0 | 13 | 0.78 | 1.00 | 0.88 | 0.88 | 0.78 | 1.00 |
| model_665 | Hemophilia B | 21 | 588 | 567 | 470 | 59 | 59 | 0.26 | 0.02 | 0.03 | 1 | 0 | 0 | 58 | 0.02 | 1.00 | 0.03 | 3.93 | 0.02 | 1.00 |
| model_666 | Hemothorax | 272 | 588 | 316 | 470 | 59 | 59 | 0.24 | 0.9 | 0.88 | 53 | 0 | 0 | 6 | 0.9 | 1.00 | 0.95 | 0.41 | 0.9 | 1.00 |
| model_667 | Hepatic encephalopathy | 2992 | 2992 | 0 | 2393 | 300 | 299 | 0.25 | 0.95 | 0.95 | 286 | 0 | 0 | 14 | 0.95 | 1.00 | 0.98 | 0.19 | 0.95 | 1.00 |
| model_668 | Hepatic failure | 8479 | 8479 | 0 | 6783 | 848 | 848 | 0.27 | 0.95 | 0.94 | 803 | 0 | 0 | 45 | 0.95 | 1.00 | 0.97 | 0.21 | 0.95 | 1.00 |
| model_669 | Hepatic necrosis | 2662 | 2662 | 0 | 2129 | 267 | 266 | 0.23 | 0.96 | 0.94 | 255 | 0 | 0 | 12 | 0.96 | 1.00 | 0.98 | 0.18 | 0.96 | 1.00 |
| model_670 | Hepatic neoplasia | 1217 | 1217 | 0 | 973 | 122 | 122 | 0.22 | 0.93 | 0.93 | 113 | 0 | 0 | 9 | 0.93 | 1.00 | 0.96 | 0.3 | 0.93 | 1.00 |
| model_671 | Hepatitis | 7251 | 7251 | 0 | 5800 | 726 | 725 | 0.27 | 0.94 | 0.94 | 684 | 0 | 0 | 42 | 0.94 | 1.00 | 0.97 | 0.23 | 0.94 | 1.00 |
| model_672 | Hepatitis A | 612 | 612 | 0 | 489 | 62 | 61 | 0.22 | 0.95 | 0.97 | 59 | 0 | 0 | 3 | 0.95 | 1.00 | 0.98 | 0.19 | 0.95 | 1.00 |
| model_673 | Hepatitis B | 1191 | 1191 | 0 | 952 | 120 | 119 | 0.23 | 0.93 | 0.92 | 112 | 0 | 0 | 8 | 0.93 | 1.00 | 0.97 | 0.27 | 0.93 | 1.00 |
| model_674 | Hepatitis B surface antigen positive | 297 | 588 | 291 | 470 | 59 | 59 | 0.23 | 0.88 | 0.81 | 52 | 0 | 0 | 7 | 0.88 | 1.00 | 0.94 | 0.47 | 0.88 | 1.00 |
| model_675 | Hepatitis C | 3208 | 3208 | 0 | 2566 | 321 | 321 | 0.24 | 0.94 | 0.96 | 303 | 0 | 0 | 18 | 0.94 | 1.00 | 0.97 | 0.22 | 0.94 | 1.00 |
| model_676 | Hepatitis C antibody positive | 210 | 588 | 378 | 470 | 59 | 59 | 0.23 | 0.88 | 0.92 | 52 | 0 | 0 | 7 | 0.88 | 1.00 | 0.94 | 0.47 | 0.88 | 1.00 |
| model_677 | Hepatitis toxic | 2201 | 2201 | 0 | 1760 | 221 | 220 | 0.23 | 0.91 | 0.96 | 201 | 0 | 0 | 20 | 0.91 | 1.00 | 0.95 | 0.36 | 0.91 | 1.00 |
| model_678 | Hepatorenal syndrome | 1507 | 1507 | 0 | 1205 | 151 | 151 | 0.23 | 0.91 | 0.96 | 137 | 0 | 0 | 14 | 0.91 | 1.00 | 0.95 | 0.37 | 0.91 | 1.00 |
| model_679 | Hernia | 3619 | 3619 | 0 | 2895 | 362 | 362 | 0.25 | 0.96 | 0.97 | 347 | 0 | 0 | 15 | 0.96 | 1.00 | 0.98 | 0.17 | 0.96 | 1.00 |
| model_680 | Hernia Inguinal | 1330 | 1330 | 0 | 1064 | 133 | 133 | 0.22 | 0.93 | 0.99 | 124 | 0 | 0 | 9 | 0.93 | 1.00 | 0.96 | 0.27 | 0.93 | 1.00 |
| model_681 | Hernia hiatal | 8257 | 8257 | 0 | 6605 | 826 | 826 | 0.26 | 0.95 | 0.95 | 788 | 0 | 0 | 38 | 0.95 | 1.00 | 0.98 | 0.18 | 0.95 | 1.00 |
| model_682 | Herpes ophthalmic | 488 | 588 | 100 | 470 | 59 | 59 | 0.22 | 0.93 | 0.95 | 55 | 0 | 0 | 4 | 0.93 | 1.00 | 0.96 | 0.27 | 0.93 | 1.00 |
| model_683 | Herpes simplex | 4340 | 4340 | 0 | 3472 | 434 | 434 | 0.24 | 0.95 | 0.96 | 413 | 0 | 0 | 21 | 0.95 | 1.00 | 0.98 | 0.19 | 0.95 | 1.00 |
| model_684 | Herpes zoster | 7359 | 7359 | 0 | 5887 | 736 | 736 | 0.28 | 0.94 | 0.96 | 694 | 0 | 0 | 42 | 0.94 | 1.00 | 0.97 | 0.23 | 0.94 | 1.00 |
| model_685 | Herxheimer reaction | 36 | 588 | 552 | 470 | 59 | 59 | 0.25 | 0.44 | 0.46 | 26 | 0 | 0 | 33 | 0.44 | 1.00 | 0.61 | 2.24 | 0.44 | 1.00 |
| model_686 | Hesitancy | 2262 | 2262 | 0 | 1809 | 227 | 226 | 0.23 | 0.96 | 0.99 | 217 | 0 | 0 | 10 | 0.96 | 1.00 | 0.98 | 0.18 | 0.96 | 1.00 |
| model_687 | Hiccough | 2166 | 2166 | 0 | 1732 | 217 | 217 | 0.24 | 0.93 | 0.94 | 201 | 0 | 0 | 16 | 0.93 | 1.00 | 0.96 | 0.29 | 0.93 | 1.00 |
| model_688 | High blood pressure | 19376 | 19376 | 0 | 15500 | 1938 | 1938 | 0.34 | 0.95 | 0.95 | 1838 | 0 | 0 | 100 | 0.95 | 1.00 | 0.97 | 0.21 | 0.95 | 1.00 |
| model_689 | High risk pregnancy | 102 | 588 | 486 | 470 | 59 | 59 | 0.26 | 0.76 | 0.78 | 45 | 0 | 0 | 14 | 0.76 | 1.00 | 0.87 | 0.95 | 0.76 | 1.00 |
| model_690 | Hip fracture | 3928 | 3928 | 0 | 3142 | 393 | 393 | 0.24 | 0.95 | 0.94 | 373 | 0 | 0 | 20 | 0.95 | 1.00 | 0.97 | 0.2 | 0.95 | 1.00 |
| model_691 | Hirsuitism | 565 | 588 | 23 | 470 | 59 | 59 | 0.23 | 0.98 | 0.95 | 58 | 0 | 0 | 1 | 0.98 | 1.00 | 0.99 | 0.07 | 0.98 | 1.00 |
| model_692 | Histoplasmosis | 989 | 989 | 0 | 791 | 99 | 99 | 0.24 | 0.95 | 0.92 | 94 | 0 | 0 | 5 | 0.95 | 1.00 | 0.97 | 0.2 | 0.95 | 1.00 |
| model_693 | Hive | 7720 | 7720 | 0 | 6176 | 772 | 772 | 0.27 | 0.96 | 0.96 | 741 | 0 | 0 | 31 | 0.96 | 1.00 | 0.98 | 0.16 | 0.96 | 1.00 |

| | | | | | | | | | | | | | | | | | | | | |
|---|---|---|---|---|---|---|---|---|---|---|---|---|---|---|---|---|---|---|---|---|
| model_694 | Hoarseness | 1646 | 1646 | 0 | 1316 | 165 | 165 | 0.22 | 0.96 | 0.94 | 158 | 0 | 0 | 7 | 0.96 | 1.00 | 0.98 | 0.17 | 0.96 | 1.00 |
| model_695 | Hordeolum | 667 | 667 | 0 | 533 | 67 | 67 | 0.22 | 0.97 | 0.97 | 65 | 0 | 0 | 2 | 0.97 | 1.00 | 0.98 | 0.12 | 0.97 | 1.00 |
| model_696 | Hot flash | 6664 | 6664 | 0 | 5331 | 667 | 666 | 0.26 | 0.94 | 0.97 | 627 | 0 | 0 | 40 | 0.94 | 1.00 | 0.97 | 0.24 | 0.94 | 1.00 |
| model_697 | Humerus fracture | 1986 | 1986 | 0 | 1588 | 199 | 199 | 0.23 | 0.95 | 0.93 | 190 | 0 | 0 | 9 | 0.95 | 1.00 | 0.98 | 0.18 | 0.95 | 1.00 |
| model_698 | Hydrocele | 446 | 588 | 142 | 470 | 59 | 59 | 0.22 | 0.95 | 0.93 | 56 | 0 | 0 | 3 | 0.95 | 1.00 | 0.97 | 0.2 | 0.95 | 1.00 |
| model_699 | Hydrocephalus | 2611 | 2611 | 0 | 2088 | 262 | 261 | 0.24 | 0.94 | 0.95 | 246 | 0 | 0 | 16 | 0.94 | 1.00 | 0.97 | 0.24 | 0.94 | 1.00 |
| model_700 | Hydronephrosis | 3881 | 3881 | 0 | 3104 | 389 | 388 | 0.25 | 0.95 | 0.93 | 369 | 0 | 0 | 20 | 0.95 | 1.00 | 0.97 | 0.21 | 0.95 | 1.00 |
| model_701 | Hydroureter | 760 | 760 | 0 | 608 | 76 | 76 | 0.23 | 0.93 | 0.93 | 71 | 0 | 0 | 5 | 0.93 | 1.00 | 0.97 | 0.26 | 0.93 | 1.00 |
| model_702 | Hyperacusia | 865 | 865 | 0 | 692 | 87 | 86 | 0.22 | 0.95 | 0.98 | 83 | 0 | 0 | 4 | 0.95 | 1.00 | 0.98 | 0.18 | 0.95 | 1.00 |
| model_703 | Hyperaesthesia | 1965 | 1965 | 0 | 1572 | 197 | 196 | 0.25 | 0.94 | 0.94 | 185 | 0 | 0 | 12 | 0.94 | 1.00 | 0.97 | 0.24 | 0.94 | 1.00 |
| model_704 | Hyperalimentation | 4816 | 4816 | 0 | 3852 | 482 | 482 | 0.25 | 0.95 | 0.96 | 456 | 0 | 0 | 26 | 0.95 | 1.00 | 0.97 | 0.22 | 0.95 | 1.00 |
| model_705 | Hyperamylasaemia | 259 | 588 | 329 | 470 | 59 | 59 | 0.24 | 0.95 | 0.83 | 56 | 0 | 0 | 3 | 0.95 | 1.00 | 0.97 | 0.2 | 0.95 | 1.00 |
| model_706 | Hypercalcinuria | 87 | 588 | 501 | 470 | 59 | 59 | 0.25 | 0.83 | 0.73 | 49 | 0 | 0 | 10 | 0.83 | 1.00 | 0.91 | 0.68 | 0.83 | 1.00 |
| model_707 | Hypercapnia | 1831 | 1831 | 0 | 1464 | 184 | 183 | 0.25 | 0.94 | 0.92 | 173 | 0 | 0 | 11 | 0.94 | 1.00 | 0.97 | 0.24 | 0.94 | 1.00 |
| model_708 | Hyperchloremia | 265 | 588 | 323 | 470 | 59 | 59 | 0.23 | 0.92 | 0.92 | 54 | 0 | 0 | 5 | 0.92 | 1.00 | 0.96 | 0.34 | 0.92 | 1.00 |
| model_709 | Hyperglycaemia | 18779 | 18779 | 0 | 15023 | 1878 | 1878 | 0.33 | 0.96 | 0.95 | 1798 | 0 | 0 | 80 | 0.96 | 1.00 | 0.98 | 0.17 | 0.96 | 1.00 |
| model_710 | Hyperhidrosis | 13438 | 13438 | 0 | 10750 | 1344 | 1344 | 0.29 | 0.95 | 0.96 | 1276 | 0 | 0 | 68 | 0.95 | 1.00 | 0.97 | 0.2 | 0.95 | 1.00 |
| model_711 | Hyperkeratosis | 2862 | 2862 | 0 | 2289 | 287 | 286 | 0.23 | 0.94 | 0.97 | 271 | 0 | 0 | 16 | 0.94 | 1.00 | 0.97 | 0.22 | 0.94 | 1.00 |
| model_712 | Hyperlipaemia | 9173 | 9173 | 0 | 7338 | 918 | 917 | 0.27 | 0.95 | 0.96 | 870 | 0 | 0 | 48 | 0.95 | 1.00 | 0.97 | 0.21 | 0.95 | 1.00 |
| model_713 | Hypermagnesemia | 387 | 588 | 201 | 470 | 59 | 59 | 0.23 | 0.93 | 0.93 | 55 | 0 | 0 | 4 | 0.93 | 1.00 | 0.96 | 0.27 | 0.93 | 1.00 |
| model_714 | Hypermetropia | 546 | 588 | 42 | 470 | 59 | 59 | 0.22 | 0.95 | 0.95 | 56 | 0 | 0 | 3 | 0.95 | 1.00 | 0.97 | 0.2 | 0.95 | 1.00 |
| model_715 | Hypernatraemia | 4485 | 4485 | 0 | 3588 | 449 | 448 | 0.26 | 0.94 | 0.91 | 421 | 0 | 0 | 28 | 0.94 | 1.00 | 0.97 | 0.25 | 0.94 | 1.00 |
| model_716 | Hyperparathyroidism | 1711 | 1711 | 0 | 1368 | 172 | 171 | 0.24 | 0.94 | 0.95 | 162 | 0 | 0 | 10 | 0.94 | 1.00 | 0.97 | 0.23 | 0.94 | 1.00 |
| model_717 | Hyperphosphatemia | 1280 | 1280 | 0 | 1024 | 128 | 128 | 0.24 | 0.95 | 0.92 | 122 | 0 | 0 | 6 | 0.95 | 1.00 | 0.98 | 0.19 | 0.95 | 1.00 |
| model_718 | Hyperpigmentation | 1525 | 1525 | 0 | 1220 | 153 | 152 | 0.23 | 0.91 | 0.92 | 139 | 0 | 0 | 14 | 0.91 | 1.00 | 0.95 | 0.37 | 0.91 | 1.00 |
| model_719 | Hyperprolactinaemia | 665 | 665 | 0 | 532 | 67 | 66 | 0.22 | 0.96 | 0.95 | 64 | 0 | 0 | 3 | 0.96 | 1.00 | 0.98 | 0.18 | 0.96 | 1.00 |
| model_720 | Hypersplenism | 274 | 588 | 314 | 470 | 59 | 59 | 0.24 | 0.88 | 0.88 | 52 | 0 | 0 | 7 | 0.88 | 1.00 | 0.94 | 0.47 | 0.88 | 1.00 |
| model_721 | Hyperthyroidism | 3463 | 3463 | 0 | 2770 | 347 | 346 | 0.26 | 0.96 | 0.95 | 333 | 0 | 0 | 14 | 0.96 | 1.00 | 0.98 | 0.16 | 0.96 | 1.00 |
| model_722 | Hypertrichosis | 306 | 588 | 282 | 470 | 59 | 59 | 0.24 | 0.95 | 0.92 | 56 | 0 | 0 | 3 | 0.95 | 1.00 | 0.97 | 0.2 | 0.95 | 1.00 |
| model_723 | Hyperuricaemia | 3413 | 3413 | 0 | 2730 | 342 | 341 | 0.26 | 0.96 | 0.94 | 329 | 0 | 0 | 13 | 0.96 | 1.00 | 0.98 | 0.15 | 0.96 | 1.00 |
| model_724 | Hypervitaminosis | 72 | 588 | 516 | 470 | 59 | 59 | 0.26 | 0.58 | 0.58 | 34 | 0 | 0 | 25 | 0.58 | 1.00 | 0.73 | 1.69 | 0.58 | 1.00 |
| model_725 | Hypoaldosteronism | 184 | 588 | 404 | 470 | 59 | 59 | 0.25 | 0.93 | 0.88 | 55 | 0 | 0 | 4 | 0.93 | 1.00 | 0.96 | 0.27 | 0.93 | 1.00 |
| model_726 | Hypochloraemia | 6428 | 6428 | 0 | 5142 | 643 | 643 | 0.27 | 0.95 | 0.95 | 612 | 0 | 0 | 31 | 0.95 | 1.00 | 0.98 | 0.19 | 0.95 | 1.00 |
| model_727 | Hypochondriasis | 64 | 588 | 524 | 470 | 59 | 59 | 0.25 | 0.78 | 0.69 | 46 | 0 | 0 | 13 | 0.78 | 1.00 | 0.88 | 0.88 | 0.78 | 1.00 |
| model_728 | Hypogammaglobulinaemia | 931 | 931 | 0 | 744 | 94 | 93 | 0.23 | 0.96 | 0.9 | 90 | 0 | 0 | 4 | 0.96 | 1.00 | 0.98 | 0.17 | 0.96 | 1.00 |
| model_729 | Hypoglycaemia | 12443 | 12443 | 0 | 9954 | 1245 | 1244 | 0.29 | 0.97 | 0.96 | 1206 | 0 | 0 | 39 | 0.97 | 1.00 | 0.98 | 0.13 | 0.97 | 1.00 |
| model_730 | Hypoglycaemia neonatal | 1206 | 1206 | 0 | 964 | 121 | 121 | 0.23 | 0.88 | 0.97 | 106 | 0 | 0 | 15 | 0.88 | 1.00 | 0.93 | 0.5 | 0.88 | 1.00 |
| model_731 | Hypogonadism | 722 | 722 | 0 | 577 | 73 | 72 | 0.27 | 0.88 | 0.92 | 64 | 0 | 0 | 9 | 0.88 | 1.00 | 0.93 | 0.49 | 0.88 | 1.00 |
| model_732 | Hypogonadotropic hypogonadism | 2 | 588 | 586 | 470 | 59 | 59 | 0.22 | 0 | 0 | 0 | 0 | 0 | 59 | 0 | 0.00 | 0 | 4 | 0 | 1.00 |
| model_733 | Hypokalaemia | 12715 | 12715 | 0 | 10172 | 1272 | 1271 | 0.31 | 0.95 | 0.96 | 1203 | 0 | 0 | 69 | 0.95 | 1.00 | 0.97 | 0.22 | 0.95 | 1.00 |
| model_734 | Hypomagnesaemia | 5408 | 5408 | 0 | 4326 | 541 | 541 | 0.26 | 0.95 | 0.94 | 512 | 0 | 0 | 29 | 0.95 | 1.00 | 0.97 | 0.21 | 0.95 | 1.00 |
| model_735 | Hypomenorrhea | 53 | 588 | 535 | 470 | 59 | 59 | 0.25 | 0.47 | 0.51 | 28 | 0 | 0 | 31 | 0.47 | 1.00 | 0.64 | 2.1 | 0.47 | 1.00 |
| model_736 | Hypoparathyroidism | 156 | 588 | 432 | 470 | 59 | 59 | 0.25 | 0.85 | 0.86 | 50 | 0 | 0 | 9 | 0.85 | 1.00 | 0.92 | 0.61 | 0.85 | 1.00 |
| model_737 | Hypophosphataemia | 2824 | 2824 | 0 | 2259 | 283 | 282 | 0.24 | 0.93 | 0.93 | 264 | 0 | 0 | 19 | 0.93 | 1.00 | 0.97 | 0.27 | 0.93 | 1.00 |
| model_738 | Hypopituitarism | 349 | 588 | 239 | 470 | 59 | 59 | 0.23 | 0.95 | 0.88 | 56 | 0 | 0 | 3 | 0.95 | 1.00 | 0.97 | 0.2 | 0.95 | 1.00 |
| model_739 | Hypospadias | 163 | 588 | 425 | 470 | 59 | 59 | 0.25 | 0.93 | 0.9 | 55 | 0 | 0 | 4 | 0.93 | 1.00 | 0.96 | 0.27 | 0.93 | 1.00 |
| model_740 | Hypotension Orthostatic | 7553 | 7553 | 0 | 6042 | 756 | 755 | 0.27 | 0.95 | 0.94 | 715 | 0 | 0 | 41 | 0.95 | 1.00 | 0.97 | 0.22 | 0.95 | 1.00 |
| model_741 | Hypothyroid | 7954 | 7954 | 0 | 6363 | 796 | 795 | 0.27 | 0.96 | 0.97 | 764 | 0 | 0 | 32 | 0.96 | 1.00 | 0.98 | 0.16 | 0.96 | 1.00 |
| model_742 | Hypoventilation | 21322 | 21322 | 0 | 17057 | 2133 | 2132 | 0.35 | 0.95 | 0.95 | 2035 | 0 | 0 | 98 | 0.95 | 1.00 | 0.98 | 0.18 | 0.95 | 1.00 |
| model_743 | Hypovitaminosis | 556 | 588 | 32 | 470 | 59 | 59 | 0.24 | 0.9 | 0.93 | 53 | 0 | 0 | 6 | 0.9 | 1.00 | 0.95 | 0.41 | 0.9 | 1.00 |

| | | | | | | | | | | | | | | | | | | | | |
|---|---|---|---|---|---|---|---|---|---|---|---|---|---|---|---|---|---|---|---|---|
| model_744 | Hypoxia | 11805 | 11805 | 0 | 9444 | 1181 | 1180 | 0.29 | 0.94 | 0.95 | 1110 | 0 | 0 | 71 | 0.94 | 1.00 | 0.97 | 0.24 | 0.94 | 1.00 |
| model_745 | Ichthyosis | 236 | 588 | 352 | 470 | 59 | 59 | 0.24 | 0.9 | 0.9 | 53 | 0 | 0 | 6 | 0.9 | 1.00 | 0.95 | 0.41 | 0.9 | 1.00 |
| model_746 | Icterus | 7944 | 7944 | 0 | 6355 | 795 | 794 | 0.28 | 0.94 | 0.94 | 744 | 0 | 0 | 51 | 0.94 | 1.00 | 0.97 | 0.26 | 0.94 | 1.00 |
| model_747 | Idiopathic thrombocytopenic purpura | 1685 | 1685 | 0 | 1348 | 169 | 168 | 0.25 | 0.88 | 0.93 | 148 | 0 | 0 | 21 | 0.88 | 1.00 | 0.93 | 0.5 | 0.88 | 1.00 |
| model_748 | IgA nephropathy | 46 | 588 | 542 | 470 | 59 | 59 | 0.27 | 0.42 | 0.61 | 25 | 0 | 0 | 34 | 0.42 | 1.00 | 0.6 | 2.31 | 0.42 | 1.00 |
| model_749 | Ild | 6573 | 6573 | 0 | 5258 | 658 | 657 | 0.26 | 0.96 | 0.94 | 629 | 0 | 0 | 29 | 0.96 | 1.00 | 0.98 | 0.18 | 0.96 | 1.00 |
| model_750 | Impetigo | 507 | 588 | 81 | 470 | 59 | 59 | 0.22 | 0.92 | 0.76 | 54 | 0 | 0 | 5 | 0.92 | 1.00 | 0.96 | 0.34 | 0.92 | 1.00 |
| model_751 | Incisional hernia | 1034 | 1034 | 0 | 827 | 104 | 103 | 0.24 | 0.92 | 0.88 | 96 | 0 | 0 | 8 | 0.92 | 1.00 | 0.96 | 0.31 | 0.92 | 1.00 |
| model_752 | Incontinence | 10616 | 10616 | 0 | 8492 | 1062 | 1062 | 0.29 | 0.97 | 0.97 | 1027 | 0 | 0 | 35 | 0.97 | 1.00 | 0.98 | 0.13 | 0.97 | 1.00 |
| model_753 | Increased platelet count | 7077 | 7077 | 0 | 5661 | 708 | 708 | 0.28 | 0.94 | 0.96 | 664 | 0 | 0 | 44 | 0.94 | 1.00 | 0.97 | 0.25 | 0.94 | 1.00 |
| model_754 | Increased white blood cell count | 11687 | 11687 | 0 | 9349 | 1169 | 1169 | 0.3 | 0.95 | 0.94 | 1109 | 0 | 0 | 60 | 0.95 | 1.00 | 0.97 | 0.21 | 0.95 | 1.00 |
| model_755 | Infection | 13172 | 13172 | 0 | 10537 | 1318 | 1317 | 0.31 | 0.95 | 0.95 | 1248 | 0 | 0 | 70 | 0.95 | 1.00 | 0.97 | 0.21 | 0.95 | 1.00 |
| model_756 | Infection Upper Respiratory | 9356 | 9356 | 0 | 7484 | 936 | 936 | 0.28 | 0.97 | 0.97 | 907 | 0 | 0 | 29 | 0.97 | 1.00 | 0.98 | 0.12 | 0.97 | 1.00 |
| model_757 | Infection Urinary Tract | 17658 | 17658 | 0 | 14126 | 1766 | 1766 | 0.33 | 0.96 | 0.95 | 1688 | 0 | 0 | 78 | 0.96 | 1.00 | 0.98 | 0.18 | 0.96 | 1.00 |
| model_758 | Infection Viral | 6589 | 6589 | 0 | 5271 | 659 | 659 | 0.27 | 0.95 | 0.95 | 623 | 0 | 0 | 36 | 0.95 | 1.00 | 0.97 | 0.22 | 0.95 | 1.00 |
| model_759 | Infectious mononucleosis | 745 | 745 | 0 | 596 | 75 | 74 | 0.22 | 0.92 | 0.91 | 69 | 0 | 0 | 6 | 0.92 | 1.00 | 0.96 | 0.32 | 0.92 | 1.00 |
| model_760 | Infertility | 47 | 588 | 541 | 470 | 59 | 59 | 0.25 | 0.56 | 0.54 | 33 | 0 | 0 | 26 | 0.56 | 1.00 | 0.72 | 1.76 | 0.56 | 1.00 |
| model_761 | Infestation | 136 | 588 | 452 | 470 | 59 | 59 | 0.25 | 0.71 | 0.66 | 42 | 0 | 0 | 17 | 0.71 | 1.00 | 0.83 | 1.15 | 0.71 | 1.00 |
| model_762 | Inflammatory bowel disease | 1475 | 1475 | 0 | 1180 | 148 | 147 | 0.24 | 0.92 | 0.95 | 136 | 0 | 0 | 12 | 0.92 | 1.00 | 0.96 | 0.32 | 0.92 | 1.00 |
| model_763 | Ingrowing nail | 1012 | 1012 | 0 | 809 | 102 | 101 | 0.22 | 0.97 | 0.93 | 99 | 0 | 0 | 3 | 0.97 | 1.00 | 0.99 | 0.12 | 0.97 | 1.00 |
| model_764 | Injury of neck | 2577 | 2577 | 0 | 2061 | 258 | 258 | 0.24 | 0.96 | 0.93 | 248 | 0 | 0 | 10 | 0.96 | 1.00 | 0.98 | 0.16 | 0.96 | 1.00 |
| model_765 | Insomnia | 15450 | 15450 | 0 | 12360 | 1545 | 1545 | 0.31 | 0.96 | 0.96 | 1477 | 0 | 0 | 68 | 0.96 | 1.00 | 0.98 | 0.18 | 0.96 | 1.00 |
| model_766 | Insulin dependent diabetes mellitus | 3335 | 3335 | 0 | 2668 | 334 | 333 | 0.24 | 0.94 | 0.96 | 313 | 0 | 0 | 21 | 0.94 | 1.00 | 0.97 | 0.25 | 0.94 | 1.00 |
| model_767 | Interstitial nephritis | 3319 | 3319 | 0 | 2655 | 332 | 332 | 0.24 | 0.94 | 0.93 | 311 | 0 | 0 | 21 | 0.94 | 1.00 | 0.97 | 0.25 | 0.94 | 1.00 |
| model_768 | Intertrigo | 206 | 588 | 382 | 470 | 59 | 59 | 0.24 | 0.9 | 0.95 | 53 | 0 | 0 | 6 | 0.9 | 1.00 | 0.95 | 0.41 | 0.9 | 1.00 |
| model_769 | Intervertebral Disc Herniation | 7154 | 7154 | 0 | 5723 | 716 | 715 | 0.26 | 0.97 | 0.97 | 693 | 0 | 0 | 23 | 0.97 | 1.00 | 0.98 | 0.13 | 0.97 | 1.00 |
| model_770 | Intervertebral disc disorder | 5455 | 5455 | 0 | 4364 | 546 | 545 | 0.25 | 0.96 | 0.97 | 522 | 0 | 0 | 24 | 0.96 | 1.00 | 0.98 | 0.18 | 0.96 | 1.00 |
| model_771 | Intestinal functional disorder | 1549 | 1549 | 0 | 1239 | 155 | 155 | 0.23 | 0.97 | 0.95 | 150 | 0 | 0 | 5 | 0.97 | 1.00 | 0.98 | 0.13 | 0.97 | 1.00 |
| model_772 | Intestinal perforation | 3405 | 3405 | 0 | 2724 | 341 | 340 | 0.24 | 0.91 | 0.93 | 311 | 0 | 0 | 30 | 0.91 | 1.00 | 0.95 | 0.35 | 0.91 | 1.00 |
| model_773 | Intestinal ulcer | 775 | 775 | 0 | 620 | 78 | 77 | 0.22 | 0.95 | 0.94 | 74 | 0 | 0 | 4 | 0.95 | 1.00 | 0.97 | 0.21 | 0.95 | 1.00 |
| model_774 | Intraocular inflammation | 858 | 858 | 0 | 686 | 86 | 86 | 0.23 | 0.94 | 0.99 | 81 | 0 | 0 | 5 | 0.94 | 1.00 | 0.97 | 0.23 | 0.94 | 1.00 |
| model_775 | Intussusception | 139 | 588 | 449 | 470 | 59 | 59 | 0.25 | 0.83 | 0.88 | 49 | 0 | 0 | 10 | 0.83 | 1.00 | 0.91 | 0.68 | 0.83 | 1.00 |
| model_776 | Iridocyclitis | 241 | 588 | 347 | 470 | 59 | 59 | 0.24 | 0.92 | 0.86 | 54 | 0 | 0 | 5 | 0.92 | 1.00 | 0.96 | 0.34 | 0.92 | 1.00 |
| model_777 | Iritis | 386 | 588 | 202 | 470 | 59 | 59 | 0.23 | 0.97 | 0.93 | 57 | 0 | 0 | 2 | 0.97 | 1.00 | 0.98 | 0.14 | 0.97 | 1.00 |
| model_778 | Iron deficiency anaemia | 5053 | 5053 | 0 | 4042 | 506 | 505 | 0.25 | 0.94 | 0.96 | 478 | 0 | 0 | 28 | 0.94 | 1.00 | 0.97 | 0.22 | 0.94 | 1.00 |
| model_779 | Irregular menstrual cycle | 1023 | 1023 | 0 | 818 | 103 | 102 | 0.24 | 0.95 | 0.99 | 98 | 0 | 0 | 5 | 0.95 | 1.00 | 0.98 | 0.19 | 0.95 | 1.00 |
| model_780 | Irritation Skin | 1220 | 1220 | 0 | 976 | 122 | 122 | 0.24 | 0.93 | 0.98 | 114 | 0 | 0 | 8 | 0.93 | 1.00 | 0.97 | 0.26 | 0.93 | 1.00 |
| model_781 | Ischaemia | 4408 | 4408 | 0 | 3526 | 441 | 441 | 0.25 | 0.92 | 0.95 | 407 | 0 | 0 | 34 | 0.92 | 1.00 | 0.96 | 0.31 | 0.92 | 1.00 |
| model_782 | Ischias | 4259 | 4259 | 0 | 3407 | 426 | 426 | 0.25 | 0.96 | 0.95 | 408 | 0 | 0 | 18 | 0.96 | 1.00 | 0.98 | 0.17 | 0.96 | 1.00 |
| model_783 | Itch | 13364 | 13364 | 0 | 10691 | 1337 | 1336 | 0.3 | 0.96 | 0.96 | 1281 | 0 | 0 | 56 | 0.96 | 1.00 | 0.98 | 0.17 | 0.96 | 1.00 |
| model_784 | JRA | 321 | 588 | 267 | 470 | 59 | 59 | 0.22 | 0.86 | 0.86 | 51 | 0 | 0 | 8 | 0.86 | 1.00 | 0.93 | 0.54 | 0.86 | 1.00 |
| model_785 | Jaundice neonatal | 202 | 588 | 386 | 470 | 59 | 59 | 0.24 | 0.95 | 0.98 | 56 | 0 | 0 | 3 | 0.95 | 1.00 | 0.97 | 0.2 | 0.95 | 1.00 |
| model_786 | Joint effusion | 2485 | 2485 | 0 | 1988 | 249 | 248 | 0.25 | 0.97 | 0.96 | 242 | 0 | 0 | 7 | 0.97 | 1.00 | 0.99 | 0.11 | 0.97 | 1.00 |
| model_787 | Joint stiffness | 3544 | 3544 | 0 | 2835 | 355 | 354 | 0.24 | 0.95 | 0.94 | 337 | 0 | 0 | 18 | 0.95 | 1.00 | 0.97 | 0.2 | 0.95 | 1.00 |
| model_788 | Joint swelling | 9187 | 9187 | 0 | 7349 | 919 | 919 | 0.29 | 0.96 | 0.96 | 885 | 0 | 0 | 34 | 0.96 | 1.00 | 0.98 | 0.15 | 0.96 | 1.00 |
| model_789 | Keloid scar | 117 | 588 | 471 | 470 | 59 | 59 | 0.24 | 0.81 | 0.81 | 48 | 0 | 0 | 11 | 0.81 | 1.00 | 0.9 | 0.75 | 0.81 | 1.00 |
| model_790 | Keratitis | 931 | 931 | 0 | 744 | 94 | 93 | 0.22 | 0.99 | 0.96 | 93 | 0 | 0 | 1 | 0.99 | 1.00 | 0.99 | 0.04 | 0.99 | 1.00 |
| model_791 | Keratitis sicca | 1097 | 1097 | 0 | 877 | 110 | 110 | 0.23 | 0.98 | 0.96 | 108 | 0 | 0 | 2 | 0.98 | 1.00 | 0.99 | 0.07 | 0.98 | 1.00 |
| model_792 | Keratoacanthomas | 471 | 588 | 117 | 470 | 59 | 59 | 0.23 | 1 | 0.9 | 59 | 0 | 0 | 0 | 1 | 1.00 | 1 | 0 | 1 | 1.00 |
| model_793 | Ketoacidosis | 1362 | 1362 | 0 | 1089 | 137 | 136 | 0.22 | 0.95 | 0.99 | 130 | 0 | 0 | 7 | 0.95 | 1.00 | 0.97 | 0.2 | 0.95 | 1.00 |

| | | | | | | | | | | | | | | | | | | | | |
|---|---|---|---|---|---|---|---|---|---|---|---|---|---|---|---|---|---|---|---|---|
| model_794 | Kidney failure | 18892 | 18892 | 0 | 15113 | 1890 | 1889 | 0.32 | 0.94 | 0.94 | 1784 | 0 | 0 | 106 | 0.94 | 1.00 | 0.97 | 0.22 | 0.94 | 1.00 |
| model_795 | Kidney neoplasm | 413 | 588 | 175 | 470 | 59 | 59 | 0.23 | 0.85 | 0.95 | 50 | 0 | 0 | 9 | 0.85 | 1.00 | 0.92 | 0.61 | 0.85 | 1.00 |
| model_796 | Kidney pain | 2523 | 2523 | 0 | 2018 | 253 | 252 | 0.24 | 0.97 | 0.97 | 246 | 0 | 0 | 7 | 0.97 | 1.00 | 0.99 | 0.11 | 0.97 | 1.00 |
| model_797 | Kidney transplant | 1364 | 1364 | 0 | 1091 | 137 | 136 | 0.23 | 0.85 | 0.89 | 117 | 0 | 0 | 20 | 0.85 | 1.00 | 0.92 | 0.58 | 0.85 | 1.00 |
| model_798 | Labyrinthitis | 1593 | 1593 | 0 | 1274 | 160 | 159 | 0.23 | 0.98 | 0.93 | 156 | 0 | 0 | 4 | 0.98 | 1.00 | 0.99 | 0.1 | 0.98 | 1.00 |
| model_799 | Laceration | 3983 | 3983 | 0 | 3186 | 399 | 398 | 0.25 | 0.95 | 0.96 | 381 | 0 | 0 | 18 | 0.95 | 1.00 | 0.98 | 0.18 | 0.95 | 1.00 |
| model_800 | Lacrimal Disorder | 211 | 588 | 377 | 470 | 59 | 59 | 0.23 | 0.86 | 0.85 | 51 | 0 | 0 | 8 | 0.86 | 1.00 | 0.93 | 0.54 | 0.86 | 1.00 |
| model_801 | Lactose intolerance | 423 | 588 | 165 | 470 | 59 | 59 | 0.22 | 0.86 | 0.88 | 51 | 0 | 0 | 8 | 0.86 | 1.00 | 0.93 | 0.54 | 0.86 | 1.00 |
| model_802 | Lagophthalmos | 38 | 588 | 550 | 470 | 59 | 59 | 0.26 | 0.12 | 0.17 | 7 | 0 | 0 | 52 | 0.12 | 1.00 | 0.21 | 3.53 | 0.12 | 1.00 |
| model_803 | Laryngeal neoplasia | 33 | 588 | 555 | 470 | 59 | 59 | 0.26 | 0.03 | 0.03 | 2 | 0 | 0 | 57 | 0.03 | 1.00 | 0.07 | 3.86 | 0.03 | 1.00 |
| model_804 | Laryngitis | 3168 | 3168 | 0 | 2534 | 317 | 317 | 0.25 | 0.96 | 0.97 | 305 | 0 | 0 | 12 | 0.96 | 1.00 | 0.98 | 0.15 | 0.96 | 1.00 |
| model_805 | Left ventricular hypertrophy | 2019 | 2019 | 0 | 1615 | 202 | 202 | 0.24 | 0.95 | 0.94 | 191 | 0 | 0 | 11 | 0.95 | 1.00 | 0.97 | 0.22 | 0.95 | 1.00 |
| model_806 | Legionella | 492 | 588 | 96 | 470 | 59 | 59 | 0.23 | 0.98 | 0.93 | 58 | 0 | 0 | 1 | 0.98 | 1.00 | 0.99 | 0.07 | 0.98 | 1.00 |
| model_807 | Leriche syndrome | 98 | 588 | 490 | 470 | 59 | 59 | 0.26 | 0.8 | 0.75 | 47 | 0 | 0 | 12 | 0.8 | 1.00 | 0.89 | 0.81 | 0.8 | 1.00 |
| model_808 | Leucocytosis | 7669 | 7669 | 0 | 6135 | 767 | 767 | 0.26 | 0.95 | 0.95 | 728 | 0 | 0 | 39 | 0.95 | 1.00 | 0.97 | 0.2 | 0.95 | 1.00 |
| model_809 | Leucopenia | 8313 | 8313 | 0 | 6650 | 832 | 831 | 0.27 | 0.94 | 0.94 | 778 | 0 | 0 | 54 | 0.94 | 1.00 | 0.97 | 0.26 | 0.94 | 1.00 |
| model_810 | Leukaemia | 4266 | 4266 | 0 | 3412 | 427 | 427 | 0.26 | 0.95 | 0.94 | 404 | 0 | 0 | 23 | 0.95 | 1.00 | 0.97 | 0.22 | 0.95 | 1.00 |
| model_811 | Leukoplakia | 303 | 588 | 285 | 470 | 59 | 59 | 0.23 | 0.92 | 0.92 | 54 | 0 | 0 | 5 | 0.92 | 1.00 | 0.96 | 0.34 | 0.92 | 1.00 |
| model_812 | Leukoplakia of mouth | 293 | 588 | 295 | 470 | 59 | 59 | 0.23 | 0.98 | 0.92 | 58 | 0 | 0 | 1 | 0.98 | 1.00 | 0.99 | 0.07 | 0.98 | 1.00 |
| model_813 | Lice Infestation | 235 | 588 | 353 | 470 | 59 | 59 | 0.24 | 0.86 | 0.83 | 51 | 0 | 0 | 8 | 0.86 | 1.00 | 0.93 | 0.54 | 0.86 | 1.00 |
| model_814 | Lichen planus | 941 | 941 | 0 | 752 | 95 | 94 | 0.22 | 0.99 | 0.96 | 94 | 0 | 0 | 1 | 0.99 | 1.00 | 0.99 | 0.04 | 0.99 | 1.00 |
| model_815 | Light sensitivity | 3768 | 3768 | 0 | 3014 | 377 | 377 | 0.26 | 0.96 | 0.96 | 361 | 0 | 0 | 16 | 0.96 | 1.00 | 0.98 | 0.17 | 0.96 | 1.00 |
| model_816 | Limb malformation | 261 | 588 | 327 | 470 | 59 | 59 | 0.23 | 0.88 | 0.93 | 52 | 0 | 0 | 7 | 0.88 | 1.00 | 0.94 | 0.47 | 0.88 | 1.00 |
| model_817 | Lipoma | 9754 | 9754 | 0 | 7803 | 976 | 975 | 0.27 | 0.96 | 0.96 | 936 | 0 | 0 | 40 | 0.96 | 1.00 | 0.98 | 0.16 | 0.96 | 1.00 |
| model_818 | Liver abscess | 1025 | 1025 | 0 | 820 | 103 | 102 | 0.23 | 0.88 | 0.82 | 91 | 0 | 0 | 12 | 0.88 | 1.00 | 0.94 | 0.47 | 0.88 | 1.00 |
| model_819 | Lobar pneumonia | 9583 | 9583 | 0 | 7666 | 959 | 958 | 0.28 | 0.94 | 0.95 | 904 | 0 | 0 | 55 | 0.94 | 1.00 | 0.97 | 0.23 | 0.94 | 1.00 |
| model_820 | Loss of consciousness | 14354 | 14354 | 0 | 11483 | 1436 | 1435 | 0.3 | 0.96 | 0.95 | 1374 | 0 | 0 | 62 | 0.96 | 1.00 | 0.98 | 0.17 | 0.96 | 1.00 |
| model_821 | Loss of weight | 18785 | 18785 | 0 | 15028 | 1879 | 1878 | 0.32 | 0.96 | 0.97 | 1800 | 0 | 0 | 79 | 0.96 | 1.00 | 0.98 | 0.17 | 0.96 | 1.00 |
| model_822 | Lower GI bleeding | 2982 | 2982 | 0 | 2385 | 299 | 298 | 0.24 | 0.93 | 0.93 | 278 | 0 | 0 | 21 | 0.93 | 1.00 | 0.96 | 0.28 | 0.93 | 1.00 |
| model_823 | Lung abscess | 712 | 712 | 0 | 569 | 72 | 71 | 0.23 | 0.93 | 0.99 | 67 | 0 | 0 | 5 | 0.93 | 1.00 | 0.96 | 0.28 | 0.93 | 1.00 |
| model_824 | Lung adenocarcinoma | 739 | 739 | 0 | 591 | 74 | 74 | 0.24 | 0.97 | 0.93 | 72 | 0 | 0 | 2 | 0.97 | 1.00 | 0.99 | 0.11 | 0.97 | 1.00 |
| model_825 | Lung edema | 12309 | 12309 | 0 | 9847 | 1231 | 1231 | 0.29 | 0.94 | 0.95 | 1162 | 0 | 0 | 69 | 0.94 | 1.00 | 0.97 | 0.22 | 0.94 | 1.00 |
| model_826 | Lung fibrosis | 4802 | 4802 | 0 | 3841 | 481 | 480 | 0.25 | 0.95 | 0.94 | 457 | 0 | 0 | 24 | 0.95 | 1.00 | 0.97 | 0.2 | 0.95 | 1.00 |
| model_827 | Lung infiltration | 8465 | 8465 | 0 | 6772 | 847 | 846 | 0.28 | 0.94 | 0.93 | 792 | 0 | 0 | 55 | 0.94 | 1.00 | 0.97 | 0.26 | 0.94 | 1.00 |
| model_828 | Lung neoplasm malignant | 4121 | 4121 | 0 | 3296 | 413 | 412 | 0.24 | 0.95 | 0.96 | 394 | 0 | 0 | 19 | 0.95 | 1.00 | 0.98 | 0.18 | 0.95 | 1.00 |
| model_829 | Lung neoplasms | 6472 | 6472 | 0 | 5177 | 648 | 647 | 0.26 | 0.94 | 0.94 | 612 | 0 | 0 | 36 | 0.94 | 1.00 | 0.97 | 0.22 | 0.94 | 1.00 |
| model_830 | Lyell | 4760 | 4760 | 0 | 3808 | 476 | 476 | 0.25 | 0.93 | 0.9 | 441 | 0 | 0 | 35 | 0.93 | 1.00 | 0.96 | 0.29 | 0.93 | 1.00 |
| model_831 | Lyme Disease | 651 | 651 | 0 | 520 | 66 | 65 | 0.23 | 1 | 0.95 | 66 | 0 | 0 | 0 | 1 | 1.00 | 1 | 0 | 1 | 1.00 |
| model_832 | Lymphangitis | 421 | 588 | 167 | 470 | 59 | 59 | 0.23 | 0.92 | 0.86 | 54 | 0 | 0 | 5 | 0.92 | 1.00 | 0.96 | 0.34 | 0.92 | 1.00 |
| model_833 | Lymphatic Disorders | 194 | 588 | 394 | 470 | 59 | 59 | 0.22 | 0.9 | 0.93 | 53 | 0 | 0 | 6 | 0.9 | 1.00 | 0.95 | 0.41 | 0.9 | 1.00 |
| model_834 | Lymphedema | 2278 | 2278 | 0 | 1822 | 228 | 228 | 0.24 | 0.96 | 0.9 | 219 | 0 | 0 | 9 | 0.96 | 1.00 | 0.98 | 0.16 | 0.96 | 1.00 |
| model_835 | Lymphocytes decreased | 2327 | 2327 | 0 | 1861 | 233 | 233 | 0.24 | 0.92 | 0.95 | 215 | 0 | 0 | 18 | 0.92 | 1.00 | 0.96 | 0.31 | 0.92 | 1.00 |
| model_836 | Lymphocytosis | 685 | 685 | 0 | 548 | 69 | 68 | 0.24 | 0.97 | 0.99 | 67 | 0 | 0 | 2 | 0.97 | 1.00 | 0.99 | 0.12 | 0.97 | 1.00 |
| model_837 | Lymphoma | 3418 | 3418 | 0 | 2734 | 342 | 342 | 0.24 | 0.96 | 0.93 | 329 | 0 | 0 | 13 | 0.96 | 1.00 | 0.98 | 0.15 | 0.96 | 1.00 |
| model_838 | MPD | 514 | 588 | 74 | 470 | 59 | 59 | 0.22 | 0.93 | 0.93 | 55 | 0 | 0 | 4 | 0.93 | 1.00 | 0.96 | 0.27 | 0.93 | 1.00 |
| model_839 | Macrocytosis | 917 | 917 | 0 | 733 | 92 | 92 | 0.24 | 0.95 | 0.9 | 87 | 0 | 0 | 5 | 0.95 | 1.00 | 0.97 | 0.22 | 0.95 | 1.00 |
| model_840 | Macroglossia | 214 | 588 | 374 | 470 | 59 | 59 | 0.23 | 0.95 | 0.85 | 56 | 0 | 0 | 3 | 0.95 | 1.00 | 0.97 | 0.2 | 0.95 | 1.00 |
| model_841 | Macula lutea degeneration | 1905 | 1905 | 0 | 1524 | 191 | 190 | 0.24 | 0.95 | 0.95 | 181 | 0 | 0 | 10 | 0.95 | 1.00 | 0.97 | 0.21 | 0.95 | 1.00 |
| model_842 | Major depression | 4636 | 4636 | 0 | 3708 | 464 | 464 | 0.25 | 0.98 | 0.96 | 453 | 0 | 0 | 11 | 0.98 | 1.00 | 0.99 | 0.09 | 0.98 | 1.00 |
| model_843 | Malabsorption | 1258 | 1258 | 0 | 1006 | 126 | 126 | 0.23 | 0.94 | 0.95 | 119 | 0 | 0 | 7 | 0.94 | 1.00 | 0.97 | 0.22 | 0.94 | 1.00 |

| | | | | | | | | | | | | | | | | | | | | |
|---|---|---|---|---|---|---|---|---|---|---|---|---|---|---|---|---|---|---|---|---|
| model_844 | Malignant hypertension | 1355 | 1355 | 0 | 1084 | 136 | 135 | 0.22 | 0.99 | 0.98 | 135 | 0 | 0 | 1 | 0.99 | 1.00 | 1 | 0.03 | 0.99 | 1.00 |
| model_845 | Malignant melanoma | 2171 | 2171 | 0 | 1736 | 218 | 217 | 0.25 | 0.93 | 0.95 | 203 | 0 | 0 | 15 | 0.93 | 1.00 | 0.96 | 0.28 | 0.93 | 1.00 |
| model_846 | Malnourished | 7308 | 7308 | 0 | 5846 | 731 | 731 | 0.25 | 0.95 | 0.94 | 694 | 0 | 0 | 37 | 0.95 | 1.00 | 0.97 | 0.2 | 0.95 | 1.00 |
| model_847 | Masculinization | 28 | 588 | 560 | 470 | 59 | 59 | 0.25 | 0.25 | 0.31 | 15 | 0 | 0 | 44 | 0.25 | 1.00 | 0.41 | 2.98 | 0.25 | 1.00 |
| model_848 | Mast cell disease | 93 | 588 | 495 | 470 | 59 | 59 | 0.24 | 0.83 | 0.78 | 49 | 0 | 0 | 10 | 0.83 | 1.00 | 0.91 | 0.68 | 0.83 | 1.00 |
| model_849 | Mediastinal disorder | 832 | 832 | 0 | 665 | 84 | 83 | 0.23 | 0.96 | 0.88 | 81 | 0 | 0 | 3 | 0.96 | 1.00 | 0.98 | 0.14 | 0.96 | 1.00 |
| model_850 | Mediastinal tumor | 61 | 588 | 527 | 470 | 59 | 59 | 0.26 | 0.51 | 0.73 | 30 | 0 | 0 | 29 | 0.51 | 1.00 | 0.67 | 1.97 | 0.51 | 1.00 |
| model_851 | Megacolon | 675 | 675 | 0 | 540 | 68 | 67 | 0.23 | 0.97 | 0.93 | 66 | 0 | 0 | 2 | 0.97 | 1.00 | 0.99 | 0.12 | 0.97 | 1.00 |
| model_852 | Meibomianitis | 23 | 588 | 565 | 470 | 59 | 59 | 0.27 | 0 | 0 | 0 | 0 | 0 | 59 | 0 | 0.00 | 0 | 4 | 0 | 1.00 |
| model_853 | Melanocytic naevus | 2209 | 2209 | 0 | 1767 | 221 | 221 | 0.24 | 0.96 | 0.97 | 213 | 0 | 0 | 8 | 0.96 | 1.00 | 0.98 | 0.14 | 0.96 | 1.00 |
| model_854 | Melanosis | 29 | 588 | 559 | 470 | 59 | 59 | 0.25 | 0.76 | 0.64 | 45 | 0 | 0 | 14 | 0.76 | 1.00 | 0.87 | 0.95 | 0.76 | 1.00 |
| model_855 | Meningeal irritation | 247 | 588 | 341 | 470 | 59 | 59 | 0.22 | 0.9 | 0.86 | 53 | 0 | 0 | 6 | 0.9 | 1.00 | 0.95 | 0.41 | 0.9 | 1.00 |
| model_856 | Meningioma | 1477 | 1477 | 0 | 1181 | 148 | 148 | 0.24 | 0.97 | 0.94 | 143 | 0 | 0 | 5 | 0.97 | 1.00 | 0.98 | 0.14 | 0.97 | 1.00 |
| model_857 | Meningitides | 2281 | 2281 | 0 | 1824 | 229 | 228 | 0.24 | 0.96 | 0.9 | 219 | 0 | 0 | 10 | 0.96 | 1.00 | 0.98 | 0.17 | 0.96 | 1.00 |
| model_858 | Meningitis Viral | 870 | 870 | 0 | 696 | 87 | 87 | 0.23 | 0.92 | 0.93 | 80 | 0 | 0 | 7 | 0.92 | 1.00 | 0.96 | 0.32 | 0.92 | 1.00 |
| model_859 | Meningocele | 142 | 588 | 446 | 470 | 59 | 59 | 0.23 | 0.88 | 0.9 | 52 | 0 | 0 | 7 | 0.88 | 1.00 | 0.94 | 0.47 | 0.88 | 1.00 |
| model_860 | Menometrorrhagia | 455 | 588 | 133 | 470 | 59 | 59 | 0.23 | 0.81 | 0.8 | 48 | 0 | 0 | 11 | 0.81 | 1.00 | 0.9 | 0.75 | 0.81 | 1.00 |
| model_861 | Menstrual Disorder | 989 | 989 | 0 | 791 | 99 | 99 | 0.22 | 0.97 | 0.98 | 96 | 0 | 0 | 3 | 0.97 | 1.00 | 0.98 | 0.12 | 0.97 | 1.00 |
| model_862 | Mental retardation | 12 | 588 | 576 | 470 | 59 | 59 | 0.23 | 0.05 | 0.03 | 3 | 0 | 0 | 56 | 0.05 | 1.00 | 0.1 | 3.8 | 0.05 | 1.00 |
| model_863 | Metabolic acidosis | 8259 | 8259 | 0 | 6607 | 826 | 826 | 0.27 | 0.95 | 0.92 | 785 | 0 | 0 | 41 | 0.95 | 1.00 | 0.97 | 0.2 | 0.95 | 1.00 |
| model_864 | Metabolic alkalosis | 3331 | 3331 | 0 | 2664 | 334 | 333 | 0.23 | 0.95 | 0.94 | 316 | 0 | 0 | 18 | 0.95 | 1.00 | 0.97 | 0.22 | 0.95 | 1.00 |
| model_865 | Metabolic encephalopathy | 2742 | 2742 | 0 | 2193 | 275 | 274 | 0.24 | 0.92 | 0.92 | 253 | 0 | 0 | 22 | 0.92 | 1.00 | 0.96 | 0.32 | 0.92 | 1.00 |
| model_866 | Methaemoglobinaemia | 516 | 588 | 72 | 470 | 59 | 59 | 0.24 | 0.88 | 0.88 | 52 | 0 | 0 | 7 | 0.88 | 1.00 | 0.94 | 0.47 | 0.88 | 1.00 |
| model_867 | Micturition urgency | 3524 | 3524 | 0 | 2819 | 353 | 352 | 0.26 | 0.98 | 0.97 | 345 | 0 | 0 | 8 | 0.98 | 1.00 | 0.99 | 0.09 | 0.98 | 1.00 |
| model_868 | Migraine | 9621 | 9621 | 0 | 7696 | 963 | 962 | 0.28 | 0.96 | 0.95 | 922 | 0 | 0 | 41 | 0.96 | 1.00 | 0.98 | 0.17 | 0.96 | 1.00 |
| model_869 | Milia | 403 | 588 | 185 | 470 | 59 | 59 | 0.23 | 0.93 | 0.93 | 55 | 0 | 0 | 4 | 0.93 | 1.00 | 0.96 | 0.27 | 0.93 | 1.00 |
| model_870 | Mitral valve disease NOS | 2174 | 2174 | 0 | 1739 | 218 | 217 | 0.23 | 0.95 | 0.94 | 207 | 0 | 0 | 11 | 0.95 | 1.00 | 0.97 | 0.2 | 0.95 | 1.00 |
| model_871 | Mitral valve prolapse | 2677 | 2677 | 0 | 2141 | 268 | 268 | 0.25 | 0.97 | 0.97 | 260 | 0 | 0 | 8 | 0.97 | 1.00 | 0.98 | 0.12 | 0.97 | 1.00 |
| model_872 | Mixed connective tissue disease | 192 | 588 | 396 | 470 | 59 | 59 | 0.25 | 0.92 | 0.92 | 54 | 0 | 0 | 5 | 0.92 | 1.00 | 0.96 | 0.34 | 0.92 | 1.00 |
| model_873 | Mod | 10531 | 10531 | 0 | 8424 | 1054 | 1053 | 0.29 | 0.96 | 0.95 | 1015 | 0 | 0 | 39 | 0.96 | 1.00 | 0.98 | 0.15 | 0.96 | 1.00 |
| model_874 | Molluscum contagiosum | 108 | 588 | 480 | 470 | 59 | 59 | 0.26 | 0.71 | 0.75 | 42 | 0 | 0 | 17 | 0.71 | 1.00 | 0.83 | 1.15 | 0.71 | 1.00 |
| model_875 | Monoclonal gammopathy | 1182 | 1182 | 0 | 945 | 119 | 118 | 0.22 | 0.94 | 0.93 | 112 | 0 | 0 | 7 | 0.94 | 1.00 | 0.97 | 0.24 | 0.94 | 1.00 |
| model_876 | Morphea | 104 | 588 | 484 | 470 | 59 | 59 | 0.25 | 0.92 | 0.8 | 54 | 0 | 0 | 5 | 0.92 | 1.00 | 0.96 | 0.34 | 0.92 | 1.00 |
| model_877 | Motion sickness | 322 | 588 | 266 | 470 | 59 | 59 | 0.24 | 0.97 | 0.95 | 57 | 0 | 0 | 2 | 0.97 | 1.00 | 0.98 | 0.14 | 0.97 | 1.00 |
| model_878 | Motor retardation | 960 | 960 | 0 | 768 | 96 | 96 | 0.22 | 0.93 | 0.89 | 89 | 0 | 0 | 7 | 0.93 | 1.00 | 0.96 | 0.29 | 0.93 | 1.00 |
| model_879 | Mouth bleeding | 2232 | 2232 | 0 | 1785 | 224 | 223 | 0.23 | 0.94 | 0.92 | 210 | 0 | 0 | 14 | 0.94 | 1.00 | 0.97 | 0.25 | 0.94 | 1.00 |
| model_880 | Mouth pain | 5654 | 5654 | 0 | 4523 | 566 | 565 | 0.27 | 0.96 | 0.93 | 545 | 0 | 0 | 21 | 0.96 | 1.00 | 0.98 | 0.15 | 0.96 | 1.00 |
| model_881 | Mouth ulcer | 5416 | 5416 | 0 | 4332 | 542 | 542 | 0.28 | 0.93 | 0.94 | 502 | 0 | 0 | 40 | 0.93 | 1.00 | 0.96 | 0.3 | 0.93 | 1.00 |
| model_882 | Movement disorder | 4521 | 4521 | 0 | 3616 | 453 | 452 | 0.26 | 0.96 | 0.97 | 437 | 0 | 0 | 16 | 0.96 | 1.00 | 0.98 | 0.14 | 0.96 | 1.00 |
| model_883 | Mucositis oral | 7554 | 7554 | 0 | 6043 | 756 | 755 | 0.27 | 0.95 | 0.96 | 719 | 0 | 0 | 37 | 0.95 | 1.00 | 0.97 | 0.2 | 0.95 | 1.00 |
| model_884 | Multifocal leukoencephalopathy | 1089 | 1089 | 0 | 871 | 109 | 109 | 0.25 | 0.94 | 0.94 | 103 | 0 | 0 | 6 | 0.94 | 1.00 | 0.97 | 0.22 | 0.94 | 1.00 |
| model_885 | Multiple Myeloma | 4427 | 4427 | 0 | 3541 | 443 | 443 | 0.25 | 0.95 | 0.96 | 422 | 0 | 0 | 21 | 0.95 | 1.00 | 0.98 | 0.19 | 0.95 | 1.00 |
| model_886 | Multiple sclerosis | 5272 | 5272 | 0 | 4217 | 528 | 527 | 0.26 | 0.96 | 0.96 | 506 | 0 | 0 | 22 | 0.96 | 1.00 | 0.98 | 0.17 | 0.96 | 1.00 |
| model_887 | Mumps | 600 | 600 | 0 | 480 | 60 | 60 | 0.22 | 0.9 | 0.95 | 54 | 0 | 0 | 6 | 0.9 | 1.00 | 0.95 | 0.4 | 0.9 | 1.00 |
| model_888 | Muscle Rupture | 333 | 588 | 255 | 470 | 59 | 59 | 0.23 | 0.97 | 0.9 | 57 | 0 | 0 | 2 | 0.97 | 1.00 | 0.98 | 0.14 | 0.97 | 1.00 |
| model_889 | Muscle disorder | 5484 | 5484 | 0 | 4387 | 549 | 548 | 0.26 | 0.95 | 0.95 | 524 | 0 | 0 | 25 | 0.95 | 1.00 | 0.98 | 0.18 | 0.95 | 1.00 |
| model_890 | Muscle inflammation | 3461 | 3461 | 0 | 2768 | 347 | 346 | 0.24 | 0.96 | 0.95 | 333 | 0 | 0 | 14 | 0.96 | 1.00 | 0.98 | 0.16 | 0.96 | 1.00 |
| model_891 | Muscle paresis | 2946 | 2946 | 0 | 2356 | 295 | 295 | 0.24 | 0.95 | 0.95 | 279 | 0 | 0 | 16 | 0.95 | 1.00 | 0.97 | 0.22 | 0.95 | 1.00 |
| model_892 | Muscle spasm | 15332 | 15332 | 0 | 12265 | 1534 | 1533 | 0.3 | 0.96 | 0.96 | 1468 | 0 | 0 | 66 | 0.96 | 1.00 | 0.98 | 0.17 | 0.96 | 1.00 |
| model_893 | Muscle strain | 3009 | 3009 | 0 | 2407 | 301 | 301 | 0.23 | 0.98 | 0.97 | 295 | 0 | 0 | 6 | 0.98 | 1.00 | 0.99 | 0.08 | 0.98 | 1.00 |

| | | | | | | | | | | | | | | | | | | | | |
|---|---|---|---|---|---|---|---|---|---|---|---|---|---|---|---|---|---|---|---|---|
| model_894 | Muscle weakness | 11410 | 11410 | 0 | 9128 | 1141 | 1141 | 0.28 | 0.96 | 0.95 | 1093 | 0 | 0 | 48 | 0.96 | 1.00 | 0.98 | 0.17 | 0.96 | 1.00 |
| model_895 | Musculoskeletal deformity | 38 | 588 | 550 | 470 | 59 | 59 | 0.27 | 0.31 | 0.34 | 18 | 0 | 0 | 41 | 0.31 | 1.00 | 0.47 | 2.78 | 0.31 | 1.00 |
| model_896 | Musculoskeletal pain | 6766 | 6766 | 0 | 5412 | 677 | 677 | 0.26 | 0.96 | 0.95 | 652 | 0 | 0 | 25 | 0.96 | 1.00 | 0.98 | 0.15 | 0.96 | 1.00 |
| model_897 | Myasthenia gravis | 1172 | 1172 | 0 | 937 | 118 | 117 | 0.24 | 0.97 | 0.98 | 115 | 0 | 0 | 3 | 0.97 | 1.00 | 0.99 | 0.1 | 0.97 | 1.00 |
| model_898 | Mycobacterium tuberculosis infection | 2038 | 2038 | 0 | 1630 | 204 | 204 | 0.24 | 0.96 | 0.98 | 196 | 0 | 0 | 8 | 0.96 | 1.00 | 0.98 | 0.16 | 0.96 | 1.00 |
| model_899 | Mycosis fungoides | 608 | 608 | 0 | 486 | 61 | 61 | 0.22 | 0.95 | 0.97 | 58 | 0 | 0 | 3 | 0.95 | 1.00 | 0.97 | 0.2 | 0.95 | 1.00 |
| model_900 | Myelitis | 847 | 847 | 0 | 677 | 85 | 85 | 0.22 | 0.92 | 0.94 | 78 | 0 | 0 | 7 | 0.92 | 1.00 | 0.96 | 0.33 | 0.92 | 1.00 |
| model_901 | Myelodysplasia | 3172 | 3172 | 0 | 2537 | 318 | 317 | 0.26 | 0.93 | 0.91 | 295 | 0 | 0 | 23 | 0.93 | 1.00 | 0.96 | 0.29 | 0.93 | 1.00 |
| model_902 | Myelopathy | 1715 | 1715 | 0 | 1372 | 172 | 171 | 0.23 | 0.92 | 0.95 | 159 | 0 | 0 | 13 | 0.92 | 1.00 | 0.96 | 0.3 | 0.92 | 1.00 |
| model_903 | Myocarditis | 1901 | 1901 | 0 | 1520 | 191 | 190 | 0.24 | 0.93 | 0.93 | 177 | 0 | 0 | 14 | 0.93 | 1.00 | 0.96 | 0.29 | 0.93 | 1.00 |
| model_904 | Myoclonic epilepsy | 397 | 588 | 191 | 470 | 59 | 59 | 0.23 | 0.95 | 0.95 | 56 | 0 | 0 | 3 | 0.95 | 1.00 | 0.97 | 0.2 | 0.95 | 1.00 |
| model_905 | Myoglobinuria | 4257 | 4257 | 0 | 3405 | 426 | 426 | 0.25 | 0.92 | 0.93 | 392 | 0 | 0 | 34 | 0.92 | 1.00 | 0.96 | 0.32 | 0.92 | 1.00 |
| model_906 | Myoma | 13 | 588 | 575 | 470 | 59 | 59 | 0.25 | 0 | 0 | 0 | 0 | 0 | 59 | 0 | 0.00 | 0 | 4 | 0 | 1.00 |
| model_907 | Myopia | 548 | 588 | 40 | 470 | 59 | 59 | 0.24 | 0.93 | 0.95 | 55 | 0 | 0 | 4 | 0.93 | 1.00 | 0.96 | 0.27 | 0.93 | 1.00 |
| model_908 | Myringitis | 23 | 588 | 565 | 470 | 59 | 59 | 0.27 | 0 | 0 | 0 | 0 | 0 | 59 | 0 | 0.00 | 0 | 4 | 0 | 1.00 |
| model_909 | Nail disorder | 1955 | 1955 | 0 | 1564 | 196 | 195 | 0.24 | 0.94 | 0.98 | 185 | 0 | 0 | 11 | 0.94 | 1.00 | 0.97 | 0.22 | 0.94 | 1.00 |
| model_910 | Narcolepsy | 14367 | 14367 | 0 | 11493 | 1437 | 1437 | 0.33 | 0.95 | 0.95 | 1364 | 0 | 0 | 73 | 0.95 | 1.00 | 0.97 | 0.2 | 0.95 | 1.00 |
| model_911 | Nasal congestion | 6809 | 6809 | 0 | 5447 | 681 | 681 | 0.27 | 0.95 | 0.94 | 648 | 0 | 0 | 33 | 0.95 | 1.00 | 0.98 | 0.19 | 0.95 | 1.00 |
| model_912 | Nasal polyp | 563 | 588 | 25 | 470 | 59 | 59 | 0.23 | 0.88 | 0.95 | 52 | 0 | 0 | 7 | 0.88 | 1.00 | 0.94 | 0.47 | 0.88 | 1.00 |
| model_913 | Nasal septal perforation | 450 | 588 | 138 | 470 | 59 | 59 | 0.22 | 0.81 | 0.9 | 48 | 0 | 0 | 11 | 0.81 | 1.00 | 0.9 | 0.75 | 0.81 | 1.00 |
| model_914 | Nasal septum disorder | 288 | 588 | 300 | 470 | 59 | 59 | 0.23 | 0.92 | 0.86 | 54 | 0 | 0 | 5 | 0.92 | 1.00 | 0.96 | 0.34 | 0.92 | 1.00 |
| model_915 | Nasal sinus congestion | 3048 | 3048 | 0 | 2438 | 305 | 305 | 0.26 | 0.95 | 0.96 | 289 | 0 | 0 | 16 | 0.95 | 1.00 | 0.97 | 0.21 | 0.95 | 1.00 |
| model_916 | Nasopharyngitis | 6226 | 6226 | 0 | 4980 | 623 | 623 | 0.26 | 0.96 | 0.96 | 596 | 0 | 0 | 27 | 0.96 | 1.00 | 0.98 | 0.17 | 0.96 | 1.00 |
| model_917 | Nausea | 25190 | 25190 | 0 | 20152 | 2519 | 2519 | 0.37 | 0.97 | 0.96 | 2438 | 0 | 0 | 81 | 0.97 | 1.00 | 0.98 | 0.13 | 0.97 | 1.00 |
| model_918 | Near Syncope | 2828 | 2828 | 0 | 2262 | 283 | 283 | 0.25 | 0.95 | 0.95 | 268 | 0 | 0 | 15 | 0.95 | 1.00 | 0.97 | 0.21 | 0.95 | 1.00 |
| model_919 | Neck mass | 981 | 981 | 0 | 784 | 99 | 98 | 0.23 | 0.99 | 0.95 | 98 | 0 | 0 | 1 | 0.99 | 1.00 | 0.99 | 0.04 | 0.99 | 1.00 |
| model_920 | Neonatal respiratory distress syndrome | 507 | 588 | 81 | 470 | 59 | 59 | 0.22 | 0.9 | 0.85 | 53 | 0 | 0 | 6 | 0.9 | 1.00 | 0.95 | 0.41 | 0.9 | 1.00 |
| model_921 | Neoplasia | 2236 | 2236 | 0 | 1788 | 224 | 224 | 0.23 | 0.93 | 0.96 | 209 | 0 | 0 | 15 | 0.93 | 1.00 | 0.97 | 0.27 | 0.93 | 1.00 |
| model_922 | Neoplasm prostate | 165 | 588 | 423 | 470 | 59 | 59 | 0.24 | 0.83 | 0.88 | 49 | 0 | 0 | 10 | 0.83 | 1.00 | 0.91 | 0.68 | 0.83 | 1.00 |
| model_923 | Nephrocalcinosis | 2652 | 2652 | 0 | 2121 | 266 | 265 | 0.23 | 0.95 | 0.94 | 253 | 0 | 0 | 13 | 0.95 | 1.00 | 0.97 | 0.2 | 0.95 | 1.00 |
| model_924 | Nephrogenic diabetes insipidus | 1219 | 1219 | 0 | 975 | 122 | 122 | 0.23 | 0.99 | 0.98 | 121 | 0 | 0 | 1 | 0.99 | 1.00 | 1 | 0.03 | 0.99 | 1.00 |
| model_925 | Nephrolithiasis | 7249 | 7249 | 0 | 5799 | 725 | 725 | 0.27 | 0.97 | 0.97 | 701 | 0 | 0 | 24 | 0.97 | 1.00 | 0.98 | 0.13 | 0.97 | 1.00 |
| model_926 | Nephrosclerosis | 5265 | 5265 | 0 | 4212 | 527 | 526 | 0.25 | 0.96 | 0.96 | 507 | 0 | 0 | 20 | 0.96 | 1.00 | 0.98 | 0.15 | 0.96 | 1.00 |
| model_927 | Nephrotic syndrome | 2075 | 2075 | 0 | 1660 | 208 | 207 | 0.23 | 0.95 | 0.95 | 197 | 0 | 0 | 11 | 0.95 | 1.00 | 0.97 | 0.21 | 0.95 | 1.00 |
| model_928 | Nervous tension | 8260 | 8260 | 0 | 6608 | 826 | 826 | 0.26 | 0.97 | 0.96 | 803 | 0 | 0 | 23 | 0.97 | 1.00 | 0.99 | 0.11 | 0.97 | 1.00 |
| model_929 | Neumonia | 24430 | 24430 | 0 | 19544 | 2443 | 2443 | 0.37 | 0.95 | 0.94 | 2309 | 0 | 0 | 134 | 0.95 | 1.00 | 0.97 | 0.22 | 0.95 | 1.00 |
| model_930 | Neuralgia | 5374 | 5374 | 0 | 4299 | 538 | 537 | 0.25 | 0.95 | 0.96 | 512 | 0 | 0 | 26 | 0.95 | 1.00 | 0.98 | 0.19 | 0.95 | 1.00 |
| model_931 | Neuritis | 2578 | 2578 | 0 | 2062 | 258 | 258 | 0.25 | 0.94 | 0.95 | 242 | 0 | 0 | 16 | 0.94 | 1.00 | 0.97 | 0.25 | 0.94 | 1.00 |
| model_932 | Neuroblastoma | 76 | 588 | 512 | 470 | 59 | 59 | 0.24 | 0.73 | 0.53 | 43 | 0 | 0 | 16 | 0.73 | 1.00 | 0.84 | 1.08 | 0.73 | 1.00 |
| model_933 | Neurodermatitis | 720 | 720 | 0 | 576 | 72 | 72 | 0.23 | 0.89 | 0.96 | 64 | 0 | 0 | 8 | 0.89 | 1.00 | 0.94 | 0.44 | 0.89 | 1.00 |
| model_934 | Neurofibromatosis | 2 | 588 | 586 | 470 | 59 | 59 | 0.21 | 0 | 0 | 0 | 0 | 0 | 59 | 0 | 0.00 | 0 | 4 | 0 | 1.00 |
| model_935 | Neurogenic bladder | 1837 | 1837 | 0 | 1469 | 184 | 184 | 0.24 | 0.92 | 0.92 | 169 | 0 | 0 | 15 | 0.92 | 1.00 | 0.96 | 0.33 | 0.92 | 1.00 |
| model_936 | Neuroma | 433 | 588 | 155 | 470 | 59 | 59 | 0.22 | 0.92 | 0.95 | 54 | 0 | 0 | 5 | 0.92 | 1.00 | 0.96 | 0.34 | 0.92 | 1.00 |
| model_937 | Neuromuscular Disorder | 340 | 588 | 248 | 470 | 59 | 59 | 0.23 | 1 | 0.88 | 59 | 0 | 0 | 0 | 1 | 1.00 | 1 | 0 | 1 | 1.00 |
| model_938 | Neuropathy | 17910 | 17910 | 0 | 14328 | 1791 | 1791 | 0.31 | 0.96 | 0.95 | 1717 | 0 | 0 | 74 | 0.96 | 1.00 | 0.98 | 0.17 | 0.96 | 1.00 |
| model_939 | Neuropathy peripheral | 11316 | 11316 | 0 | 9052 | 1132 | 1132 | 0.29 | 0.95 | 0.96 | 1081 | 0 | 0 | 51 | 0.95 | 1.00 | 0.98 | 0.18 | 0.95 | 1.00 |
| model_940 | Neurosis | 482 | 588 | 106 | 470 | 59 | 59 | 0.23 | 0.93 | 0.9 | 55 | 0 | 0 | 4 | 0.93 | 1.00 | 0.96 | 0.27 | 0.93 | 1.00 |
| model_941 | Neutropenia | 15171 | 15171 | 0 | 12136 | 1518 | 1517 | 0.3 | 0.94 | 0.94 | 1427 | 0 | 0 | 91 | 0.94 | 1.00 | 0.97 | 0.24 | 0.94 | 1.00 |
| model_942 | Night blindness | 584 | 588 | 4 | 470 | 59 | 59 | 0.21 | 0.97 | 0.92 | 57 | 0 | 0 | 2 | 0.97 | 1.00 | 0.98 | 0.14 | 0.97 | 1.00 |
| model_943 | Night cramps | 744 | 744 | 0 | 595 | 75 | 74 | 0.23 | 0.97 | 0.97 | 73 | 0 | 0 | 2 | 0.97 | 1.00 | 0.99 | 0.11 | 0.97 | 1.00 |

| | | | | | | | | | | | | | | | | | | | | |
|---|---|---|---|---|---|---|---|---|---|---|---|---|---|---|---|---|---|---|---|---|
| model_944 | Night sweat | 7246 | 7246 | 0 | 5796 | 725 | 725 | 0.28 | 0.96 | 0.95 | 697 | 0 | 0 | 28 | 0.96 | 1.00 | 0.98 | 0.15 | 0.96 | 1.00 |
| model_945 | Nightmare | 4591 | 4591 | 0 | 3672 | 460 | 459 | 0.25 | 0.97 | 0.99 | 448 | 0 | 0 | 12 | 0.97 | 1.00 | 0.99 | 0.1 | 0.97 | 1.00 |
| model_946 | Nocturia | 4247 | 4247 | 0 | 3397 | 425 | 425 | 0.25 | 0.95 | 0.94 | 404 | 0 | 0 | 21 | 0.95 | 1.00 | 0.97 | 0.2 | 0.95 | 1.00 |
| model_947 | Nodule | 3349 | 3349 | 0 | 2679 | 335 | 335 | 0.25 | 0.96 | 0.95 | 321 | 0 | 0 | 14 | 0.96 | 1.00 | 0.98 | 0.17 | 0.96 | 1.00 |
| model_948 | Nodule Skin | 2254 | 2254 | 0 | 1803 | 226 | 225 | 0.24 | 0.93 | 0.95 | 210 | 0 | 0 | 16 | 0.93 | 1.00 | 0.96 | 0.28 | 0.93 | 1.00 |
| model_949 | Normal pressure hydrocephalus | 515 | 588 | 73 | 470 | 59 | 59 | 0.28 | 0.81 | 0.83 | 48 | 0 | 0 | 11 | 0.81 | 1.00 | 0.9 | 0.75 | 0.81 | 1.00 |
| model_950 | Nystagmus | 2319 | 2319 | 0 | 1855 | 232 | 232 | 0.24 | 0.94 | 0.93 | 217 | 0 | 0 | 15 | 0.94 | 1.00 | 0.97 | 0.26 | 0.94 | 1.00 |
| model_951 | OPTIC NEURITIS | 1719 | 1719 | 0 | 1375 | 172 | 172 | 0.23 | 0.95 | 0.94 | 163 | 0 | 0 | 9 | 0.95 | 1.00 | 0.97 | 0.21 | 0.95 | 1.00 |
| model_952 | Obesity | 6715 | 6715 | 0 | 5372 | 672 | 671 | 0.27 | 0.96 | 0.97 | 644 | 0 | 0 | 28 | 0.96 | 1.00 | 0.98 | 0.17 | 0.96 | 1.00 |
| model_953 | Obstructive uropathy | 1375 | 1375 | 0 | 1100 | 138 | 137 | 0.24 | 0.96 | 0.89 | 133 | 0 | 0 | 5 | 0.96 | 1.00 | 0.98 | 0.14 | 0.96 | 1.00 |
| model_954 | Ocular hypertension | 333 | 588 | 255 | 470 | 59 | 59 | 0.24 | 0.81 | 0.93 | 48 | 0 | 0 | 11 | 0.81 | 1.00 | 0.9 | 0.75 | 0.81 | 1.00 |
| model_955 | Odynophagia | 2993 | 2993 | 0 | 2394 | 300 | 299 | 0.24 | 0.93 | 0.94 | 280 | 0 | 0 | 20 | 0.93 | 1.00 | 0.97 | 0.27 | 0.93 | 1.00 |
| model_956 | Oily Skin | 36 | 588 | 552 | 470 | 59 | 59 | 0.26 | 0.39 | 0.42 | 23 | 0 | 0 | 36 | 0.39 | 1.00 | 0.56 | 2.44 | 0.39 | 1.00 |
| model_957 | Oligomenorrhea | 272 | 588 | 316 | 470 | 59 | 59 | 0.23 | 0.86 | 0.93 | 51 | 0 | 0 | 8 | 0.86 | 1.00 | 0.93 | 0.54 | 0.86 | 1.00 |
| model_958 | Onycholysis | 491 | 588 | 97 | 470 | 59 | 59 | 0.21 | 0.92 | 0.95 | 54 | 0 | 0 | 5 | 0.92 | 1.00 | 0.96 | 0.34 | 0.92 | 1.00 |
| model_959 | Onychomycosis | 2080 | 2080 | 0 | 1664 | 208 | 208 | 0.23 | 0.96 | 0.96 | 199 | 0 | 0 | 9 | 0.96 | 1.00 | 0.98 | 0.17 | 0.96 | 1.00 |
| model_960 | Oophorectomy | 533 | 588 | 55 | 470 | 59 | 59 | 0.24 | 0.9 | 0.92 | 53 | 0 | 0 | 6 | 0.9 | 1.00 | 0.95 | 0.41 | 0.9 | 1.00 |
| model_961 | Ophthalmoplegia | 425 | 588 | 163 | 470 | 59 | 59 | 0.22 | 0.92 | 0.88 | 54 | 0 | 0 | 5 | 0.92 | 1.00 | 0.96 | 0.34 | 0.92 | 1.00 |
| model_962 | Optic atrophy | 796 | 796 | 0 | 636 | 80 | 80 | 0.23 | 0.95 | 0.99 | 76 | 0 | 0 | 4 | 0.95 | 1.00 | 0.97 | 0.2 | 0.95 | 1.00 |
| model_963 | Optic disc edema | 1536 | 1536 | 0 | 1228 | 154 | 154 | 0.23 | 0.95 | 0.95 | 146 | 0 | 0 | 8 | 0.95 | 1.00 | 0.97 | 0.21 | 0.95 | 1.00 |
| model_964 | Oral neoplasm | 70 | 588 | 518 | 470 | 59 | 59 | 0.25 | 0.63 | 0.81 | 37 | 0 | 0 | 22 | 0.63 | 1.00 | 0.77 | 1.49 | 0.63 | 1.00 |
| model_965 | Orchiectomy | 62 | 588 | 526 | 470 | 59 | 59 | 0.25 | 0.59 | 0.56 | 35 | 0 | 0 | 24 | 0.59 | 1.00 | 0.74 | 1.63 | 0.59 | 1.00 |
| model_966 | Orchitis | 293 | 588 | 295 | 470 | 59 | 59 | 0.23 | 0.88 | 0.97 | 52 | 0 | 0 | 7 | 0.88 | 1.00 | 0.94 | 0.47 | 0.88 | 1.00 |
| model_967 | Orthopnea | 3230 | 3230 | 0 | 2584 | 323 | 323 | 0.24 | 0.95 | 0.95 | 306 | 0 | 0 | 17 | 0.95 | 1.00 | 0.97 | 0.21 | 0.95 | 1.00 |
| model_968 | Osteoarthritis | 10352 | 10352 | 0 | 8281 | 1036 | 1035 | 0.29 | 0.95 | 0.95 | 988 | 0 | 0 | 48 | 0.95 | 1.00 | 0.98 | 0.19 | 0.95 | 1.00 |
| model_969 | Osteochondrosis | 280 | 588 | 308 | 470 | 59 | 59 | 0.25 | 0.93 | 0.98 | 55 | 0 | 0 | 4 | 0.93 | 1.00 | 0.96 | 0.27 | 0.93 | 1.00 |
| model_970 | Osteogenesis Imperfecta | 99 | 588 | 489 | 470 | 59 | 59 | 0.26 | 0.58 | 0.68 | 34 | 0 | 0 | 25 | 0.58 | 1.00 | 0.73 | 1.69 | 0.58 | 1.00 |
| model_971 | Osteoma | 112 | 588 | 476 | 470 | 59 | 59 | 0.25 | 0.81 | 0.81 | 48 | 0 | 0 | 11 | 0.81 | 1.00 | 0.9 | 0.75 | 0.81 | 1.00 |
| model_972 | Osteomalacia | 3448 | 3448 | 0 | 2758 | 345 | 345 | 0.25 | 0.93 | 0.97 | 321 | 0 | 0 | 24 | 0.93 | 1.00 | 0.96 | 0.28 | 0.93 | 1.00 |
| model_973 | Osteomyelitis | 8054 | 8054 | 0 | 6443 | 806 | 805 | 0.28 | 0.94 | 0.95 | 757 | 0 | 0 | 49 | 0.94 | 1.00 | 0.97 | 0.24 | 0.94 | 1.00 |
| model_974 | Osteopetrosis | 116 | 588 | 472 | 470 | 59 | 59 | 0.26 | 0.71 | 0.66 | 42 | 0 | 0 | 17 | 0.71 | 1.00 | 0.83 | 1.15 | 0.71 | 1.00 |
| model_975 | Osteoporosis | 6854 | 6854 | 0 | 5483 | 686 | 685 | 0.27 | 0.95 | 0.96 | 653 | 0 | 0 | 33 | 0.95 | 1.00 | 0.98 | 0.19 | 0.95 | 1.00 |
| model_976 | Otitis media | 3016 | 3016 | 0 | 2412 | 302 | 302 | 0.24 | 0.93 | 0.96 | 282 | 0 | 0 | 20 | 0.93 | 1.00 | 0.97 | 0.26 | 0.93 | 1.00 |
| model_977 | Otosclerosis | 29 | 588 | 559 | 470 | 59 | 59 | 0.26 | 0.22 | 0.22 | 13 | 0 | 0 | 46 | 0.22 | 1.00 | 0.36 | 3.12 | 0.22 | 1.00 |
| model_978 | Ovarian Cyst | 3299 | 3299 | 0 | 2639 | 330 | 330 | 0.24 | 0.94 | 0.95 | 310 | 0 | 0 | 20 | 0.94 | 1.00 | 0.97 | 0.24 | 0.94 | 1.00 |
| model_979 | Ovarian cancer | 1217 | 1217 | 0 | 973 | 122 | 122 | 0.23 | 0.96 | 1 | 117 | 0 | 0 | 5 | 0.96 | 1.00 | 0.98 | 0.16 | 0.96 | 1.00 |
| model_980 | Ovulation pain | 36 | 588 | 552 | 470 | 59 | 59 | 0.25 | 0.36 | 0.49 | 21 | 0 | 0 | 38 | 0.36 | 1.00 | 0.53 | 2.58 | 0.36 | 1.00 |
| model_981 | PMs | 456 | 588 | 132 | 470 | 59 | 59 | 0.24 | 0.95 | 0.98 | 56 | 0 | 0 | 3 | 0.95 | 1.00 | 0.97 | 0.2 | 0.95 | 1.00 |
| model_982 | Pain | 23894 | 23894 | 0 | 19115 | 2390 | 2389 | 0.36 | 0.95 | 0.95 | 2279 | 0 | 0 | 111 | 0.95 | 1.00 | 0.98 | 0.19 | 0.95 | 1.00 |
| model_983 | Pain in throat | 5708 | 5708 | 0 | 4566 | 571 | 571 | 0.25 | 0.95 | 0.96 | 542 | 0 | 0 | 29 | 0.95 | 1.00 | 0.97 | 0.2 | 0.95 | 1.00 |
| model_984 | Pain pelvic | 3399 | 3399 | 0 | 2719 | 340 | 340 | 0.23 | 0.94 | 0.94 | 321 | 0 | 0 | 19 | 0.94 | 1.00 | 0.97 | 0.22 | 0.94 | 1.00 |
| model_985 | Palpitation | 11445 | 11445 | 0 | 9156 | 1145 | 1144 | 0.28 | 0.97 | 0.96 | 1114 | 0 | 0 | 31 | 0.97 | 1.00 | 0.99 | 0.11 | 0.97 | 1.00 |
| model_986 | Palsies | 4002 | 4002 | 0 | 3201 | 401 | 400 | 0.24 | 0.95 | 0.93 | 379 | 0 | 0 | 22 | 0.95 | 1.00 | 0.97 | 0.22 | 0.95 | 1.00 |
| model_987 | Pancreatic cancer | 1328 | 1328 | 0 | 1062 | 133 | 133 | 0.22 | 0.91 | 0.98 | 121 | 0 | 0 | 12 | 0.91 | 1.00 | 0.95 | 0.36 | 0.91 | 1.00 |
| model_988 | Pancreatic pseudocyst | 961 | 961 | 0 | 768 | 97 | 96 | 0.23 | 0.92 | 0.94 | 89 | 0 | 0 | 8 | 0.92 | 1.00 | 0.96 | 0.33 | 0.92 | 1.00 |
| model_989 | Pancreatitis | 10749 | 10749 | 0 | 8599 | 1075 | 1075 | 0.28 | 0.96 | 0.95 | 1030 | 0 | 0 | 45 | 0.96 | 1.00 | 0.98 | 0.17 | 0.96 | 1.00 |
| model_990 | Pancreatitis relapsing | 4180 | 4180 | 0 | 3344 | 418 | 418 | 0.24 | 0.96 | 0.95 | 402 | 0 | 0 | 16 | 0.96 | 1.00 | 0.98 | 0.15 | 0.96 | 1.00 |
| model_991 | Panic attack | 6670 | 6670 | 0 | 5336 | 667 | 667 | 0.28 | 0.96 | 0.97 | 641 | 0 | 0 | 26 | 0.96 | 1.00 | 0.98 | 0.16 | 0.96 | 1.00 |
| model_992 | Panic disorder | 3243 | 3243 | 0 | 2594 | 325 | 324 | 0.24 | 0.97 | 0.97 | 314 | 0 | 0 | 11 | 0.97 | 1.00 | 0.98 | 0.14 | 0.97 | 1.00 |
| model_993 | Panniculitis | 589 | 589 | 0 | 471 | 59 | 59 | 0.22 | 0.98 | 0.95 | 58 | 0 | 0 | 1 | 0.98 | 1.00 | 0.99 | 0.07 | 0.98 | 1.00 |

| | | | | | | | | | | | | | | | | | | | | |
|---|---|---|---|---|---|---|---|---|---|---|---|---|---|---|---|---|---|---|---|---|
| model_994 | Papillary necrosis | 121 | 588 | 467 | 470 | 59 | 59 | 0.25 | 0.83 | 0.86 | 49 | 0 | 0 | 10 | 0.83 | 1.00 | 0.91 | 0.68 | 0.83 | 1.00 |
| model_995 | Papilloma | 432 | 588 | 156 | 470 | 59 | 59 | 0.24 | 0.98 | 0.95 | 58 | 0 | 0 | 1 | 0.98 | 1.00 | 0.99 | 0.07 | 0.98 | 1.00 |
| model_996 | Paraesthesia | 13144 | 13144 | 0 | 10515 | 1315 | 1314 | 0.33 | 0.95 | 0.95 | 1251 | 0 | 0 | 64 | 0.95 | 1.00 | 0.98 | 0.19 | 0.95 | 1.00 |
| model_997 | Paranoia | 5135 | 5135 | 0 | 4108 | 514 | 513 | 0.25 | 0.97 | 0.97 | 498 | 0 | 0 | 16 | 0.97 | 1.00 | 0.98 | 0.12 | 0.97 | 1.00 |
| model_998 | Paraparesis | 729 | 729 | 0 | 583 | 73 | 73 | 0.23 | 0.93 | 0.9 | 68 | 0 | 0 | 5 | 0.93 | 1.00 | 0.96 | 0.27 | 0.93 | 1.00 |
| model_999 | Paraplegia | 1596 | 1596 | 0 | 1276 | 160 | 160 | 0.24 | 0.93 | 0.9 | 148 | 0 | 0 | 12 | 0.93 | 1.00 | 0.96 | 0.3 | 0.93 | 1.00 |
| model_1000 | Parapsoriasis | 72 | 588 | 516 | 470 | 59 | 59 | 0.25 | 0.85 | 0.83 | 50 | 0 | 0 | 9 | 0.85 | 1.00 | 0.92 | 0.61 | 0.85 | 1.00 |
| model_1001 | Parasitic infection intestinal | 10 | 588 | 578 | 470 | 59 | 59 | 0.23 | 0 | 0 | 0 | 0 | 0 | 59 | 0 | 0.00 | 0 | 4 | 0 | 1.00 |
| model_1002 | Parkinson | 1668 | 1668 | 0 | 1334 | 167 | 167 | 0.24 | 0.93 | 0.92 | 156 | 0 | 0 | 11 | 0.93 | 1.00 | 0.97 | 0.26 | 0.93 | 1.00 |
| model_1003 | Paronychia | 972 | 972 | 0 | 777 | 98 | 97 | 0.24 | 0.99 | 0.95 | 97 | 0 | 0 | 1 | 0.99 | 1.00 | 0.99 | 0.04 | 0.99 | 1.00 |
| model_1004 | Parotid gland enlargement | 799 | 799 | 0 | 639 | 80 | 80 | 0.22 | 0.94 | 0.95 | 75 | 0 | 0 | 5 | 0.94 | 1.00 | 0.97 | 0.25 | 0.94 | 1.00 |
| model_1005 | Parotitis | 848 | 848 | 0 | 678 | 85 | 85 | 0.22 | 0.94 | 0.96 | 80 | 0 | 0 | 5 | 0.94 | 1.00 | 0.97 | 0.24 | 0.94 | 1.00 |
| model_1006 | Patent ductus arteriosis | 790 | 790 | 0 | 632 | 79 | 79 | 0.23 | 0.96 | 0.94 | 76 | 0 | 0 | 3 | 0.96 | 1.00 | 0.98 | 0.15 | 0.96 | 1.00 |
| model_1007 | Peliosis | 3479 | 3479 | 0 | 2783 | 348 | 348 | 0.24 | 0.94 | 0.92 | 326 | 0 | 0 | 22 | 0.94 | 1.00 | 0.97 | 0.25 | 0.94 | 1.00 |
| model_1008 | Pelvic Inflammatory Disease | 932 | 932 | 0 | 745 | 94 | 93 | 0.23 | 0.97 | 0.96 | 91 | 0 | 0 | 3 | 0.97 | 1.00 | 0.98 | 0.13 | 0.97 | 1.00 |
| model_1009 | Pelvic infection | 219 | 588 | 369 | 470 | 59 | 59 | 0.23 | 0.93 | 0.83 | 55 | 0 | 0 | 4 | 0.93 | 1.00 | 0.96 | 0.27 | 0.93 | 1.00 |
| model_1010 | Pemphigus | 201 | 588 | 387 | 470 | 59 | 59 | 0.23 | 0.86 | 0.81 | 51 | 0 | 0 | 8 | 0.86 | 1.00 | 0.93 | 0.54 | 0.86 | 1.00 |
| model_1011 | Penile discharge | 120 | 588 | 468 | 470 | 59 | 59 | 0.24 | 0.86 | 0.93 | 51 | 0 | 0 | 8 | 0.86 | 1.00 | 0.93 | 0.54 | 0.86 | 1.00 |
| model_1012 | Penile pain | 417 | 588 | 171 | 470 | 59 | 59 | 0.23 | 0.93 | 0.88 | 55 | 0 | 0 | 4 | 0.93 | 1.00 | 0.96 | 0.27 | 0.93 | 1.00 |
| model_1013 | Penile swelling | 160 | 588 | 428 | 470 | 59 | 59 | 0.25 | 0.86 | 0.93 | 51 | 0 | 0 | 8 | 0.86 | 1.00 | 0.93 | 0.54 | 0.86 | 1.00 |
| model_1014 | Periarteritis nodosa | 97 | 588 | 491 | 470 | 59 | 59 | 0.25 | 0.85 | 0.83 | 50 | 0 | 0 | 9 | 0.85 | 1.00 | 0.92 | 0.61 | 0.85 | 1.00 |
| model_1015 | Pericardial effusion | 8775 | 8775 | 0 | 7020 | 878 | 877 | 0.27 | 0.95 | 0.94 | 830 | 0 | 0 | 48 | 0.95 | 1.00 | 0.97 | 0.22 | 0.95 | 1.00 |
| model_1016 | Pericarditis | 2917 | 2917 | 0 | 2333 | 292 | 292 | 0.24 | 0.95 | 0.96 | 278 | 0 | 0 | 14 | 0.95 | 1.00 | 0.98 | 0.19 | 0.95 | 1.00 |
| model_1017 | Periodontal disease | 2478 | 2478 | 0 | 1982 | 248 | 248 | 0.25 | 0.94 | 0.95 | 233 | 0 | 0 | 15 | 0.94 | 1.00 | 0.97 | 0.24 | 0.94 | 1.00 |
| model_1018 | Periodontitis | 1552 | 1552 | 0 | 1241 | 156 | 155 | 0.23 | 0.97 | 0.93 | 151 | 0 | 0 | 5 | 0.97 | 1.00 | 0.98 | 0.13 | 0.97 | 1.00 |
| model_1019 | Periostitis | 257 | 588 | 331 | 470 | 59 | 59 | 0.24 | 0.92 | 0.9 | 54 | 0 | 0 | 5 | 0.92 | 1.00 | 0.96 | 0.34 | 0.92 | 1.00 |
| model_1020 | Peripheral nerve injury | 89 | 588 | 499 | 470 | 59 | 59 | 0.25 | 0.92 | 0.8 | 54 | 0 | 0 | 5 | 0.92 | 1.00 | 0.96 | 0.34 | 0.92 | 1.00 |
| model_1021 | Perirectal abscess | 1119 | 1119 | 0 | 895 | 112 | 112 | 0.24 | 0.94 | 0.93 | 105 | 0 | 0 | 7 | 0.94 | 1.00 | 0.97 | 0.25 | 0.94 | 1.00 |
| model_1022 | Peritoneal adhesions | 428 | 588 | 160 | 470 | 59 | 59 | 0.23 | 0.97 | 0.93 | 57 | 0 | 0 | 2 | 0.97 | 1.00 | 0.98 | 0.14 | 0.97 | 1.00 |
| model_1023 | Peritonitis | 4407 | 4407 | 0 | 3525 | 441 | 441 | 0.25 | 0.93 | 0.93 | 412 | 0 | 0 | 29 | 0.93 | 1.00 | 0.97 | 0.26 | 0.93 | 1.00 |
| model_1024 | Personality disorder | 2464 | 2464 | 0 | 1971 | 247 | 246 | 0.25 | 0.95 | 0.94 | 234 | 0 | 0 | 13 | 0.95 | 1.00 | 0.97 | 0.21 | 0.95 | 1.00 |
| model_1025 | Pertussis | 64 | 588 | 524 | 470 | 59 | 59 | 0.26 | 0.8 | 0.76 | 47 | 0 | 0 | 12 | 0.8 | 1.00 | 0.89 | 0.81 | 0.8 | 1.00 |
| model_1026 | Petechia | 4028 | 4028 | 0 | 3222 | 403 | 403 | 0.25 | 0.95 | 0.94 | 382 | 0 | 0 | 21 | 0.95 | 1.00 | 0.97 | 0.21 | 0.95 | 1.00 |
| model_1027 | Petit mal | 3164 | 3164 | 0 | 2531 | 317 | 316 | 0.25 | 0.91 | 0.93 | 290 | 0 | 0 | 27 | 0.91 | 1.00 | 0.96 | 0.34 | 0.91 | 1.00 |
| model_1028 | Peyronies Disease | 133 | 588 | 455 | 470 | 59 | 59 | 0.25 | 0.95 | 0.85 | 56 | 0 | 0 | 3 | 0.95 | 1.00 | 0.97 | 0.2 | 0.95 | 1.00 |
| model_1029 | Phaeochromocytoma | 307 | 588 | 281 | 470 | 59 | 59 | 0.24 | 0.93 | 0.97 | 55 | 0 | 0 | 4 | 0.93 | 1.00 | 0.96 | 0.27 | 0.93 | 1.00 |
| model_1030 | Pharyngeal inflammation | 5215 | 5215 | 0 | 4172 | 522 | 521 | 0.25 | 0.94 | 0.96 | 493 | 0 | 0 | 29 | 0.94 | 1.00 | 0.97 | 0.22 | 0.94 | 1.00 |
| model_1031 | Pharyngitis Streptococcal | 1872 | 1872 | 0 | 1497 | 188 | 187 | 0.24 | 0.91 | 0.96 | 172 | 0 | 0 | 16 | 0.91 | 1.00 | 0.96 | 0.34 | 0.91 | 1.00 |
| model_1032 | Phimosis | 620 | 620 | 0 | 496 | 62 | 62 | 0.23 | 0.98 | 1 | 61 | 0 | 0 | 1 | 0.98 | 1.00 | 0.99 | 0.06 | 0.98 | 1.00 |
| model_1033 | Phlebitis | 2941 | 2941 | 0 | 2352 | 295 | 294 | 0.25 | 0.94 | 0.94 | 277 | 0 | 0 | 18 | 0.94 | 1.00 | 0.97 | 0.24 | 0.94 | 1.00 |
| model_1034 | Phlebitis superficial | 583 | 588 | 5 | 470 | 59 | 59 | 0.23 | 0.95 | 0.98 | 56 | 0 | 0 | 3 | 0.95 | 1.00 | 0.97 | 0.2 | 0.95 | 1.00 |
| model_1035 | Phlebothrombosis | 9464 | 9464 | 0 | 7571 | 947 | 946 | 0.29 | 0.95 | 0.96 | 900 | 0 | 0 | 47 | 0.95 | 1.00 | 0.97 | 0.2 | 0.95 | 1.00 |
| model_1036 | Phobia | 275 | 588 | 313 | 470 | 59 | 59 | 0.23 | 0.88 | 0.93 | 52 | 0 | 0 | 7 | 0.88 | 1.00 | 0.94 | 0.47 | 0.88 | 1.00 |
| model_1037 | Pigmentary retinopathy | 131 | 588 | 457 | 470 | 59 | 59 | 0.26 | 0.86 | 0.76 | 51 | 0 | 0 | 8 | 0.86 | 1.00 | 0.93 | 0.54 | 0.86 | 1.00 |
| model_1038 | Pigmentation disorder | 991 | 991 | 0 | 792 | 100 | 99 | 0.24 | 0.98 | 0.93 | 98 | 0 | 0 | 2 | 0.98 | 1.00 | 0.99 | 0.08 | 0.98 | 1.00 |
| model_1039 | Pilonidal cyst | 233 | 588 | 355 | 470 | 59 | 59 | 0.24 | 0.86 | 0.8 | 51 | 0 | 0 | 8 | 0.86 | 1.00 | 0.93 | 0.54 | 0.86 | 1.00 |
| model_1040 | Pityriasis | 22 | 588 | 566 | 470 | 59 | 59 | 0.25 | 0.22 | 0.2 | 13 | 0 | 0 | 46 | 0.22 | 1.00 | 0.36 | 3.12 | 0.22 | 1.00 |
| model_1041 | Platelet disorder | 968 | 968 | 0 | 774 | 97 | 97 | 0.23 | 0.89 | 0.9 | 86 | 0 | 0 | 11 | 0.89 | 1.00 | 0.94 | 0.45 | 0.89 | 1.00 |
| model_1042 | Pleural Effusion | 16899 | 16899 | 0 | 13519 | 1690 | 1690 | 0.32 | 0.95 | 0.94 | 1612 | 0 | 0 | 78 | 0.95 | 1.00 | 0.98 | 0.18 | 0.95 | 1.00 |
| model_1043 | Pleural fibrosis | 1696 | 1696 | 0 | 1356 | 170 | 170 | 0.24 | 0.95 | 0.94 | 162 | 0 | 0 | 8 | 0.95 | 1.00 | 0.98 | 0.19 | 0.95 | 1.00 |

| | | | | | | | | | | | | | | | | | | | | |
|---|---|---|---|---|---|---|---|---|---|---|---|---|---|---|---|---|---|---|---|---|
| model_1044 | Pleural pain | 21781 | 21781 | 0 | 17424 | 2179 | 2178 | 0.33 | 0.97 | 0.95 | 2110 | 0 | 0 | 69 | 0.97 | 1.00 | 0.98 | 0.13 | 0.97 | 1.00 |
| model_1045 | Pleurisy | 3382 | 3382 | 0 | 2705 | 339 | 338 | 0.24 | 0.94 | 0.95 | 319 | 0 | 0 | 20 | 0.94 | 1.00 | 0.97 | 0.24 | 0.94 | 1.00 |
| model_1046 | Pneumoconiosis | 527 | 588 | 61 | 470 | 59 | 59 | 0.23 | 0.9 | 0.9 | 53 | 0 | 0 | 6 | 0.9 | 1.00 | 0.95 | 0.41 | 0.9 | 1.00 |
| model_1047 | Pneumocystis carinii infection | 302 | 588 | 286 | 470 | 59 | 59 | 0.22 | 0.86 | 0.78 | 51 | 0 | 0 | 8 | 0.86 | 1.00 | 0.93 | 0.54 | 0.86 | 1.00 |
| model_1048 | Pneumocystis carinii pneumonia | 827 | 827 | 0 | 661 | 83 | 83 | 0.23 | 0.9 | 0.87 | 75 | 0 | 0 | 8 | 0.9 | 1.00 | 0.95 | 0.39 | 0.9 | 1.00 |
| model_1049 | Pneumonectomy | 73 | 588 | 515 | 470 | 59 | 59 | 0.26 | 0.58 | 0.63 | 34 | 0 | 0 | 25 | 0.58 | 1.00 | 0.73 | 1.69 | 0.58 | 1.00 |
| model_1050 | Pneumonia Klebsiella | 1792 | 1792 | 0 | 1433 | 180 | 179 | 0.23 | 0.96 | 0.9 | 172 | 0 | 0 | 8 | 0.96 | 1.00 | 0.98 | 0.18 | 0.96 | 1.00 |
| model_1051 | Pneumonia primary atypical | 1343 | 1343 | 0 | 1074 | 135 | 134 | 0.23 | 0.93 | 0.96 | 126 | 0 | 0 | 9 | 0.93 | 1.00 | 0.97 | 0.27 | 0.93 | 1.00 |
| model_1052 | Pneumonia staphylococcal | 2609 | 2609 | 0 | 2087 | 261 | 261 | 0.24 | 0.93 | 0.92 | 244 | 0 | 0 | 17 | 0.93 | 1.00 | 0.97 | 0.26 | 0.93 | 1.00 |
| model_1053 | Pneumothorax | 5342 | 5342 | 0 | 4273 | 535 | 534 | 0.25 | 0.94 | 0.93 | 504 | 0 | 0 | 31 | 0.94 | 1.00 | 0.97 | 0.23 | 0.94 | 1.00 |
| model_1054 | Polio | 80 | 588 | 508 | 470 | 59 | 59 | 0.27 | 0.42 | 0.61 | 25 | 0 | 0 | 34 | 0.42 | 1.00 | 0.6 | 2.31 | 0.42 | 1.00 |
| model_1055 | Polyarthritis | 2005 | 2005 | 0 | 1604 | 201 | 200 | 0.24 | 0.99 | 0.97 | 199 | 0 | 0 | 2 | 0.99 | 1.00 | 1 | 0.04 | 0.99 | 1.00 |
| model_1056 | Polycystic ovary disease | 286 | 588 | 302 | 470 | 59 | 59 | 0.23 | 0.95 | 0.88 | 56 | 0 | 0 | 3 | 0.95 | 1.00 | 0.97 | 0.2 | 0.95 | 1.00 |
| model_1057 | Polymenorrhea | 172 | 588 | 416 | 470 | 59 | 59 | 0.25 | 0.86 | 0.9 | 51 | 0 | 0 | 8 | 0.86 | 1.00 | 0.93 | 0.54 | 0.86 | 1.00 |
| model_1058 | Polymyalgia rheumatica | 1220 | 1220 | 0 | 976 | 122 | 122 | 0.24 | 0.93 | 0.94 | 113 | 0 | 0 | 9 | 0.93 | 1.00 | 0.96 | 0.3 | 0.93 | 1.00 |
| model_1059 | Polymyositis | 1109 | 1109 | 0 | 887 | 111 | 111 | 0.23 | 0.95 | 0.98 | 106 | 0 | 0 | 5 | 0.95 | 1.00 | 0.98 | 0.18 | 0.95 | 1.00 |
| model_1060 | Polyneuropathy | 3450 | 3450 | 0 | 2760 | 345 | 345 | 0.23 | 0.92 | 0.93 | 319 | 0 | 0 | 26 | 0.92 | 1.00 | 0.96 | 0.3 | 0.92 | 1.00 |
| model_1061 | Polyp | 2540 | 2540 | 0 | 2032 | 254 | 254 | 0.24 | 0.97 | 0.94 | 246 | 0 | 0 | 8 | 0.97 | 1.00 | 0.98 | 0.13 | 0.97 | 1.00 |
| model_1062 | Polyuria | 3366 | 3366 | 0 | 2692 | 337 | 337 | 0.26 | 0.96 | 0.94 | 325 | 0 | 0 | 12 | 0.96 | 1.00 | 0.98 | 0.14 | 0.96 | 1.00 |
| model_1063 | Porphyria | 250 | 588 | 338 | 470 | 59 | 59 | 0.24 | 0.88 | 0.85 | 52 | 0 | 0 | 7 | 0.88 | 1.00 | 0.94 | 0.47 | 0.88 | 1.00 |
| model_1064 | Portal hypertension | 1792 | 1792 | 0 | 1433 | 180 | 179 | 0.25 | 0.91 | 0.93 | 164 | 0 | 0 | 16 | 0.91 | 1.00 | 0.95 | 0.36 | 0.91 | 1.00 |
| model_1065 | Portal vein thrombosis | 1016 | 1016 | 0 | 812 | 102 | 102 | 0.23 | 0.92 | 0.93 | 94 | 0 | 0 | 8 | 0.92 | 1.00 | 0.96 | 0.31 | 0.92 | 1.00 |
| model_1066 | Post nasal drip | 1907 | 1907 | 0 | 1525 | 191 | 191 | 0.26 | 0.96 | 0.96 | 184 | 0 | 0 | 7 | 0.96 | 1.00 | 0.98 | 0.15 | 0.96 | 1.00 |
| model_1067 | Post thrombotic syndrome | 208 | 588 | 380 | 470 | 59 | 59 | 0.25 | 0.85 | 0.86 | 50 | 0 | 0 | 9 | 0.85 | 1.00 | 0.92 | 0.61 | 0.85 | 1.00 |
| model_1068 | Post traumatic stress disorder | 2307 | 2307 | 0 | 1845 | 231 | 231 | 0.24 | 0.96 | 0.96 | 221 | 0 | 0 | 10 | 0.96 | 1.00 | 0.98 | 0.17 | 0.96 | 1.00 |
| model_1069 | Postherpetic Neuralgia | 1084 | 1084 | 0 | 867 | 109 | 108 | 0.24 | 0.96 | 0.92 | 105 | 0 | 0 | 4 | 0.96 | 1.00 | 0.98 | 0.15 | 0.96 | 1.00 |
| model_1070 | Postmenopausal bleeding | 1130 | 1130 | 0 | 904 | 113 | 113 | 0.21 | 0.92 | 0.96 | 104 | 0 | 0 | 9 | 0.92 | 1.00 | 0.96 | 0.32 | 0.92 | 1.00 |
| model_1071 | Pregnancy induced hypertension | 174 | 588 | 414 | 470 | 59 | 59 | 0.25 | 0.88 | 0.9 | 52 | 0 | 0 | 7 | 0.88 | 1.00 | 0.94 | 0.47 | 0.88 | 1.00 |
| model_1072 | Premature menopause | 1695 | 1695 | 0 | 1356 | 170 | 169 | 0.24 | 0.96 | 0.96 | 163 | 0 | 0 | 7 | 0.96 | 1.00 | 0.98 | 0.16 | 0.96 | 1.00 |
| model_1073 | Premature separation of placenta | 168 | 588 | 420 | 470 | 59 | 59 | 0.24 | 0.78 | 0.83 | 46 | 0 | 0 | 13 | 0.78 | 1.00 | 0.88 | 0.88 | 0.78 | 1.00 |
| model_1074 | Priapism | 465 | 588 | 123 | 470 | 59 | 59 | 0.22 | 0.97 | 0.98 | 57 | 0 | 0 | 2 | 0.97 | 1.00 | 0.98 | 0.14 | 0.97 | 1.00 |
| model_1075 | Primary biliary cirrhosis | 659 | 659 | 0 | 527 | 66 | 66 | 0.23 | 0.92 | 0.95 | 61 | 0 | 0 | 5 | 0.92 | 1.00 | 0.96 | 0.3 | 0.92 | 1.00 |
| model_1076 | Prinzmetal variant angina | 466 | 588 | 122 | 470 | 59 | 59 | 0.22 | 0.93 | 0.97 | 55 | 0 | 0 | 4 | 0.93 | 1.00 | 0.96 | 0.27 | 0.93 | 1.00 |
| model_1077 | Proctalgia | 2503 | 2503 | 0 | 2002 | 251 | 250 | 0.24 | 0.93 | 0.92 | 233 | 0 | 0 | 18 | 0.93 | 1.00 | 0.96 | 0.29 | 0.93 | 1.00 |
| model_1078 | Proctitis | 966 | 966 | 0 | 772 | 97 | 97 | 0.24 | 0.95 | 0.92 | 92 | 0 | 0 | 5 | 0.95 | 1.00 | 0.97 | 0.21 | 0.95 | 1.00 |
| model_1079 | Prolactinoma | 66 | 588 | 522 | 470 | 59 | 59 | 0.26 | 0.64 | 0.51 | 38 | 0 | 0 | 21 | 0.64 | 1.00 | 0.78 | 1.42 | 0.64 | 1.00 |
| model_1080 | Prostatism | 424 | 588 | 164 | 470 | 59 | 59 | 0.23 | 0.97 | 0.86 | 57 | 0 | 0 | 2 | 0.97 | 1.00 | 0.98 | 0.14 | 0.97 | 1.00 |
| model_1081 | Prostatitis | 1861 | 1861 | 0 | 1488 | 187 | 186 | 0.23 | 0.97 | 0.97 | 182 | 0 | 0 | 5 | 0.97 | 1.00 | 0.99 | 0.11 | 0.97 | 1.00 |
| model_1082 | Proteinuria | 4720 | 4720 | 0 | 3776 | 472 | 472 | 0.26 | 0.97 | 0.97 | 457 | 0 | 0 | 15 | 0.97 | 1.00 | 0.98 | 0.13 | 0.97 | 1.00 |
| model_1083 | Psoriasis | 3856 | 3856 | 0 | 3084 | 386 | 386 | 0.24 | 0.98 | 0.96 | 379 | 0 | 0 | 7 | 0.98 | 1.00 | 0.99 | 0.07 | 0.98 | 1.00 |
| model_1084 | Psoriatic arthritis | 1119 | 1119 | 0 | 895 | 112 | 112 | 0.23 | 0.89 | 0.93 | 100 | 0 | 0 | 12 | 0.89 | 1.00 | 0.94 | 0.43 | 0.89 | 1.00 |
| model_1085 | Psychoses | 5732 | 5732 | 0 | 4585 | 574 | 573 | 0.26 | 0.96 | 0.95 | 550 | 0 | 0 | 24 | 0.96 | 1.00 | 0.98 | 0.17 | 0.96 | 1.00 |
| model_1086 | Psychosexual disorder | 2760 | 2760 | 0 | 2208 | 276 | 276 | 0.24 | 0.97 | 0.96 | 269 | 0 | 0 | 7 | 0.97 | 1.00 | 0.99 | 0.1 | 0.97 | 1.00 |
| model_1087 | Psychosomatic disease | 253 | 588 | 335 | 470 | 59 | 59 | 0.23 | 0.9 | 0.83 | 53 | 0 | 0 | 6 | 0.9 | 1.00 | 0.95 | 0.41 | 0.9 | 1.00 |
| model_1088 | Pterygium | 153 | 588 | 435 | 470 | 59 | 59 | 0.23 | 0.88 | 0.95 | 52 | 0 | 0 | 7 | 0.88 | 1.00 | 0.94 | 0.47 | 0.88 | 1.00 |
| model_1089 | Pulmonary arrest | 6027 | 6027 | 0 | 4821 | 603 | 603 | 0.26 | 0.94 | 0.95 | 569 | 0 | 0 | 34 | 0.94 | 1.00 | 0.97 | 0.23 | 0.94 | 1.00 |
| model_1090 | Pulmonary hypertension | 8677 | 8677 | 0 | 6941 | 868 | 868 | 0.27 | 0.95 | 0.95 | 824 | 0 | 0 | 44 | 0.95 | 1.00 | 0.97 | 0.2 | 0.95 | 1.00 |
| model_1091 | Pulmonary mass | 2256 | 2256 | 0 | 1804 | 226 | 226 | 0.24 | 0.93 | 0.91 | 211 | 0 | 0 | 15 | 0.93 | 1.00 | 0.97 | 0.27 | 0.93 | 1.00 |
| model_1092 | Pyelonephritis | 4183 | 4183 | 0 | 3346 | 419 | 418 | 0.25 | 0.94 | 0.96 | 395 | 0 | 0 | 24 | 0.94 | 1.00 | 0.97 | 0.23 | 0.94 | 1.00 |
| model_1093 | Pyloric stenosis | 202 | 588 | 386 | 470 | 59 | 59 | 0.24 | 0.88 | 0.75 | 52 | 0 | 0 | 7 | 0.88 | 1.00 | 0.94 | 0.47 | 0.88 | 1.00 |

| | | | | | | | | | | | | | | | | | | | | |
|---|---|---|---|---|---|---|---|---|---|---|---|---|---|---|---|---|---|---|---|---|
| model_1094 | Pyoderma | 6844 | 6844 | 0 | 5475 | 685 | 684 | 0.26 | 0.94 | 0.97 | 645 | 0 | 0 | 40 | 0.94 | 1.00 | 0.97 | 0.23 | 0.94 | 1.00 |
| model_1095 | Pyoderma gangrenosum | 901 | 901 | 0 | 720 | 91 | 90 | 0.24 | 0.93 | 0.94 | 85 | 0 | 0 | 6 | 0.93 | 1.00 | 0.97 | 0.26 | 0.93 | 1.00 |
| model_1096 | Pyogenic granuloma | 103 | 588 | 485 | 470 | 59 | 59 | 0.23 | 0.78 | 0.85 | 46 | 0 | 0 | 13 | 0.78 | 1.00 | 0.88 | 0.88 | 0.78 | 1.00 |
| model_1097 | Pyuria | 1139 | 1139 | 0 | 911 | 114 | 114 | 0.24 | 0.94 | 0.96 | 107 | 0 | 0 | 7 | 0.94 | 1.00 | 0.97 | 0.25 | 0.94 | 1.00 |
| model_1098 | Quadriplegia | 696 | 696 | 0 | 556 | 70 | 70 | 0.23 | 0.94 | 0.94 | 66 | 0 | 0 | 4 | 0.94 | 1.00 | 0.97 | 0.23 | 0.94 | 1.00 |
| model_1099 | Rabies | 1 | 588 | 587 | 470 | 59 | 59 | 0.22 | 0 | 0 | 0 | 0 | 0 | 59 | 0 | 0.00 | 0 | 4 | 0 | 1.00 |
| model_1100 | Radiation pneumonitis | 489 | 588 | 99 | 470 | 59 | 59 | 0.24 | 0.9 | 0.97 | 53 | 0 | 0 | 6 | 0.9 | 1.00 | 0.95 | 0.41 | 0.9 | 1.00 |
| model_1101 | Radiculopathy | 3348 | 3348 | 0 | 2678 | 335 | 335 | 0.26 | 0.96 | 0.94 | 320 | 0 | 0 | 15 | 0.96 | 1.00 | 0.98 | 0.18 | 0.96 | 1.00 |
| model_1102 | Raynauds phenomenon | 1942 | 1942 | 0 | 1553 | 195 | 194 | 0.23 | 0.96 | 0.95 | 188 | 0 | 0 | 7 | 0.96 | 1.00 | 0.98 | 0.14 | 0.96 | 1.00 |
| model_1103 | Rectal Tenesmus | 359 | 588 | 229 | 470 | 59 | 59 | 0.23 | 0.86 | 0.98 | 51 | 0 | 0 | 8 | 0.86 | 1.00 | 0.93 | 0.54 | 0.86 | 1.00 |
| model_1104 | Rectal discharge | 240 | 588 | 348 | 470 | 59 | 59 | 0.23 | 0.88 | 1 | 52 | 0 | 0 | 7 | 0.88 | 1.00 | 0.94 | 0.47 | 0.88 | 1.00 |
| model_1105 | Rectal polyp | 1313 | 1313 | 0 | 1050 | 132 | 131 | 0.24 | 0.95 | 0.96 | 125 | 0 | 0 | 7 | 0.95 | 1.00 | 0.97 | 0.21 | 0.95 | 1.00 |
| model_1106 | Rectal prolapse | 624 | 624 | 0 | 499 | 63 | 62 | 0.21 | 0.9 | 0.89 | 57 | 0 | 0 | 6 | 0.9 | 1.00 | 0.95 | 0.38 | 0.9 | 1.00 |
| model_1107 | Reflex sympathetic dystrophy | 289 | 588 | 299 | 470 | 59 | 59 | 0.32 | 0.97 | 0.95 | 57 | 0 | 0 | 2 | 0.97 | 1.00 | 0.98 | 0.14 | 0.97 | 1.00 |
| model_1108 | Reflux esophagitis | 3343 | 3343 | 0 | 2674 | 335 | 334 | 0.25 | 0.96 | 0.97 | 321 | 0 | 0 | 14 | 0.96 | 1.00 | 0.98 | 0.17 | 0.96 | 1.00 |
| model_1109 | Refraction disorder | 65 | 588 | 523 | 470 | 59 | 59 | 0.26 | 0.63 | 0.68 | 37 | 0 | 0 | 22 | 0.63 | 1.00 | 0.77 | 1.49 | 0.63 | 1.00 |
| model_1110 | Regurgitation | 303 | 588 | 285 | 470 | 59 | 59 | 0.23 | 0.88 | 0.97 | 52 | 0 | 0 | 7 | 0.88 | 1.00 | 0.94 | 0.47 | 0.88 | 1.00 |
| model_1111 | Renal abscess | 417 | 588 | 171 | 470 | 59 | 59 | 0.22 | 0.92 | 0.97 | 54 | 0 | 0 | 5 | 0.92 | 1.00 | 0.96 | 0.34 | 0.92 | 1.00 |
| model_1112 | Renal agenesis | 3 | 588 | 585 | 470 | 59 | 59 | 0.23 | 0 | 0 | 0 | 0 | 0 | 59 | 0 | 0.00 | 0 | 4 | 0 | 1.00 |
| model_1113 | Renal cancer | 115 | 588 | 473 | 470 | 59 | 59 | 0.25 | 0.92 | 0.92 | 54 | 0 | 0 | 5 | 0.92 | 1.00 | 0.96 | 0.34 | 0.92 | 1.00 |
| model_1114 | Renal colic | 572 | 588 | 16 | 470 | 59 | 59 | 0.21 | 0.93 | 0.97 | 55 | 0 | 0 | 4 | 0.93 | 1.00 | 0.96 | 0.27 | 0.93 | 1.00 |
| model_1115 | Renal cyst | 5447 | 5447 | 0 | 4357 | 545 | 545 | 0.26 | 0.95 | 0.95 | 517 | 0 | 0 | 28 | 0.95 | 1.00 | 0.97 | 0.21 | 0.95 | 1.00 |
| model_1116 | Renal mass | 1384 | 1384 | 0 | 1107 | 139 | 138 | 0.25 | 0.91 | 0.94 | 127 | 0 | 0 | 12 | 0.91 | 1.00 | 0.95 | 0.35 | 0.91 | 1.00 |
| model_1117 | Renal tubular acidosis | 921 | 921 | 0 | 736 | 93 | 92 | 0.23 | 0.95 | 0.93 | 88 | 0 | 0 | 5 | 0.95 | 1.00 | 0.97 | 0.22 | 0.95 | 1.00 |
| model_1118 | Renovascular hypertension | 13 | 588 | 575 | 470 | 59 | 59 | 0.24 | 0 | 0 | 0 | 0 | 0 | 59 | 0 | 0.00 | 0 | 4 | 0 | 1.00 |
| model_1119 | Respiratory alkalosis | 1093 | 1093 | 0 | 874 | 110 | 109 | 0.22 | 0.95 | 0.95 | 104 | 0 | 0 | 6 | 0.95 | 1.00 | 0.97 | 0.22 | 0.95 | 1.00 |
| model_1120 | Respiratory failure | 15961 | 15961 | 0 | 12768 | 1597 | 1596 | 0.31 | 0.94 | 0.93 | 1508 | 0 | 0 | 89 | 0.94 | 1.00 | 0.97 | 0.22 | 0.94 | 1.00 |
| model_1121 | Retinal artery occlusion | 735 | 735 | 0 | 588 | 74 | 73 | 0.24 | 0.95 | 0.96 | 70 | 0 | 0 | 4 | 0.95 | 1.00 | 0.97 | 0.22 | 0.95 | 1.00 |
| model_1122 | Retinal bleeding | 2197 | 2197 | 0 | 1757 | 220 | 220 | 0.25 | 0.94 | 0.95 | 207 | 0 | 0 | 13 | 0.94 | 1.00 | 0.97 | 0.24 | 0.94 | 1.00 |
| model_1123 | Retinal degeneration | 292 | 588 | 296 | 470 | 59 | 59 | 0.24 | 0.97 | 0.88 | 57 | 0 | 0 | 2 | 0.97 | 1.00 | 0.98 | 0.14 | 0.97 | 1.00 |
| model_1124 | Retinal scar | 98 | 588 | 490 | 470 | 59 | 59 | 0.27 | 0.78 | 0.85 | 46 | 0 | 0 | 13 | 0.78 | 1.00 | 0.88 | 0.88 | 0.78 | 1.00 |
| model_1125 | Retinal vein thrombosis | 230 | 588 | 358 | 470 | 59 | 59 | 0.22 | 0.9 | 0.88 | 53 | 0 | 0 | 6 | 0.9 | 1.00 | 0.95 | 0.41 | 0.9 | 1.00 |
| model_1126 | Retinitis | 195 | 588 | 393 | 470 | 59 | 59 | 0.23 | 0.95 | 0.88 | 56 | 0 | 0 | 3 | 0.95 | 1.00 | 0.97 | 0.2 | 0.95 | 1.00 |
| model_1127 | Retinopathy hypertensive | 205 | 588 | 383 | 470 | 59 | 59 | 0.23 | 0.95 | 0.88 | 56 | 0 | 0 | 3 | 0.95 | 1.00 | 0.97 | 0.2 | 0.95 | 1.00 |
| model_1128 | Retinopathy of prematurity | 86 | 588 | 502 | 470 | 59 | 59 | 0.25 | 0.63 | 0.69 | 37 | 0 | 0 | 22 | 0.63 | 1.00 | 0.77 | 1.49 | 0.63 | 1.00 |
| model_1129 | Retrocollis | 755 | 755 | 0 | 604 | 76 | 75 | 0.23 | 0.96 | 0.96 | 73 | 0 | 0 | 3 | 0.96 | 1.00 | 0.98 | 0.16 | 0.96 | 1.00 |
| model_1130 | Retroperitoneal fibrosis | 245 | 588 | 343 | 470 | 59 | 59 | 0.24 | 0.9 | 0.9 | 53 | 0 | 0 | 6 | 0.9 | 1.00 | 0.95 | 0.41 | 0.9 | 1.00 |
| model_1131 | Rhabdomyolysis | 8053 | 8053 | 0 | 6442 | 806 | 805 | 0.27 | 0.94 | 0.94 | 757 | 0 | 0 | 49 | 0.94 | 1.00 | 0.97 | 0.24 | 0.94 | 1.00 |
| model_1132 | Rhabdomyosarcoma | 40 | 588 | 548 | 470 | 59 | 59 | 0.28 | 0.29 | 0.24 | 17 | 0 | 0 | 42 | 0.29 | 1.00 | 0.45 | 2.85 | 0.29 | 1.00 |
| model_1133 | Rhagades | 1446 | 1446 | 0 | 1156 | 145 | 145 | 0.23 | 0.92 | 0.92 | 134 | 0 | 0 | 11 | 0.92 | 1.00 | 0.96 | 0.3 | 0.92 | 1.00 |
| model_1134 | Rheumatic fever | 109 | 588 | 479 | 470 | 59 | 59 | 0.26 | 0.69 | 0.66 | 41 | 0 | 0 | 18 | 0.69 | 1.00 | 0.82 | 1.22 | 0.69 | 1.00 |
| model_1135 | Rheumatic heart disease | 134 | 588 | 454 | 470 | 59 | 59 | 0.24 | 0.8 | 0.76 | 47 | 0 | 0 | 12 | 0.8 | 1.00 | 0.89 | 0.81 | 0.8 | 1.00 |
| model_1136 | Rhinitis | 4757 | 4757 | 0 | 3805 | 476 | 476 | 0.24 | 0.97 | 0.96 | 462 | 0 | 0 | 14 | 0.97 | 1.00 | 0.99 | 0.12 | 0.97 | 1.00 |
| model_1137 | Rhinitis Seasonal | 322 | 588 | 266 | 470 | 59 | 59 | 0.23 | 0.85 | 0.86 | 50 | 0 | 0 | 9 | 0.85 | 1.00 | 0.92 | 0.61 | 0.85 | 1.00 |
| model_1138 | Rhinorrhea | 6629 | 6629 | 0 | 5303 | 663 | 663 | 0.27 | 0.95 | 0.95 | 633 | 0 | 0 | 30 | 0.95 | 1.00 | 0.98 | 0.18 | 0.95 | 1.00 |
| model_1139 | Rib fracture | 5296 | 5296 | 0 | 4236 | 530 | 530 | 0.25 | 0.95 | 0.97 | 503 | 0 | 0 | 27 | 0.95 | 1.00 | 0.97 | 0.2 | 0.95 | 1.00 |
| model_1140 | Rickets | 8 | 588 | 580 | 470 | 59 | 59 | 0.23 | 0 | 0 | 0 | 0 | 0 | 59 | 0 | 0.00 | 0 | 4 | 0 | 1.00 |
| model_1141 | Right heart failure | 2935 | 2935 | 0 | 2348 | 294 | 293 | 0.25 | 0.94 | 0.92 | 275 | 0 | 0 | 19 | 0.94 | 1.00 | 0.97 | 0.26 | 0.94 | 1.00 |
| model_1142 | Road traffic accident | 9240 | 9240 | 0 | 7392 | 924 | 924 | 0.28 | 0.96 | 0.96 | 891 | 0 | 0 | 33 | 0.96 | 1.00 | 0.98 | 0.14 | 0.96 | 1.00 |
| model_1143 | Rotator cuff syndrome | 5649 | 5649 | 0 | 4519 | 565 | 565 | 0.26 | 0.96 | 0.97 | 543 | 0 | 0 | 22 | 0.96 | 1.00 | 0.98 | 0.16 | 0.96 | 1.00 |

| | | | | | | | | | | | | | | | | | | | | |
|---|---|---|---|---|---|---|---|---|---|---|---|---|---|---|---|---|---|---|---|---|
| model_1144 | Rubella | 9 | 588 | 579 | 470 | 59 | 59 | 0.22 | 0 | 0 | 0 | 0 | 0 | 59 | 0 | 0.00 | 0 | 4 | 0 | 1.00 |
| model_1145 | Rupture of spleen | 469 | 588 | 119 | 470 | 59 | 59 | 0.22 | 0.92 | 0.92 | 54 | 0 | 0 | 5 | 0.92 | 1.00 | 0.96 | 0.34 | 0.92 | 1.00 |
| model_1146 | SCLC | 709 | 709 | 0 | 567 | 71 | 71 | 0.22 | 0.9 | 0.94 | 64 | 0 | 0 | 7 | 0.9 | 1.00 | 0.95 | 0.39 | 0.9 | 1.00 |
| model_1147 | Sacroiliitis | 826 | 826 | 0 | 660 | 83 | 83 | 0.23 | 0.99 | 0.96 | 82 | 0 | 0 | 1 | 0.99 | 1.00 | 0.99 | 0.05 | 0.99 | 1.00 |
| model_1148 | Salivary Gland Calculus | 205 | 588 | 383 | 470 | 59 | 59 | 0.25 | 0.93 | 0.81 | 55 | 0 | 0 | 4 | 0.93 | 1.00 | 0.96 | 0.27 | 0.93 | 1.00 |
| model_1149 | Salivary gland enlargement | 79 | 588 | 509 | 470 | 59 | 59 | 0.25 | 0.83 | 0.78 | 49 | 0 | 0 | 10 | 0.83 | 1.00 | 0.91 | 0.68 | 0.83 | 1.00 |
| model_1150 | Salivary gland inflammation | 1154 | 1154 | 0 | 923 | 116 | 115 | 0.22 | 0.97 | 0.98 | 113 | 0 | 0 | 3 | 0.97 | 1.00 | 0.99 | 0.1 | 0.97 | 1.00 |
| model_1151 | Salmonella | 353 | 588 | 235 | 470 | 59 | 59 | 0.24 | 0.92 | 0.93 | 54 | 0 | 0 | 5 | 0.92 | 1.00 | 0.96 | 0.34 | 0.92 | 1.00 |
| model_1152 | Salpingitis | 172 | 588 | 416 | 470 | 59 | 59 | 0.25 | 0.97 | 0.9 | 57 | 0 | 0 | 2 | 0.97 | 1.00 | 0.98 | 0.14 | 0.97 | 1.00 |
| model_1153 | Sarcoidosis | 1579 | 1579 | 0 | 1263 | 158 | 158 | 0.24 | 0.97 | 0.95 | 153 | 0 | 0 | 5 | 0.97 | 1.00 | 0.98 | 0.13 | 0.97 | 1.00 |
| model_1154 | Sarcoma | 502 | 588 | 86 | 470 | 59 | 59 | 0.23 | 0.93 | 0.95 | 55 | 0 | 0 | 4 | 0.93 | 1.00 | 0.96 | 0.27 | 0.93 | 1.00 |
| model_1155 | Scar | 6673 | 6673 | 0 | 5338 | 668 | 667 | 0.27 | 0.96 | 0.97 | 642 | 0 | 0 | 26 | 0.96 | 1.00 | 0.98 | 0.16 | 0.96 | 1.00 |
| model_1156 | Schizoaffective disorder | 1060 | 1060 | 0 | 848 | 106 | 106 | 0.24 | 0.96 | 0.95 | 102 | 0 | 0 | 4 | 0.96 | 1.00 | 0.98 | 0.15 | 0.96 | 1.00 |
| model_1157 | Schizophrenia | 1358 | 1358 | 0 | 1086 | 136 | 136 | 0.24 | 0.96 | 0.96 | 130 | 0 | 0 | 6 | 0.96 | 1.00 | 0.98 | 0.18 | 0.96 | 1.00 |
| model_1158 | Scleritis | 152 | 588 | 436 | 470 | 59 | 59 | 0.25 | 0.95 | 0.95 | 56 | 0 | 0 | 3 | 0.95 | 1.00 | 0.97 | 0.2 | 0.95 | 1.00 |
| model_1159 | Scleroderma | 1564 | 1564 | 0 | 1251 | 157 | 156 | 0.23 | 0.96 | 0.96 | 151 | 0 | 0 | 6 | 0.96 | 1.00 | 0.98 | 0.15 | 0.96 | 1.00 |
| model_1160 | Scoliosis | 2293 | 2293 | 0 | 1834 | 230 | 229 | 0.23 | 0.97 | 0.95 | 223 | 0 | 0 | 7 | 0.97 | 1.00 | 0.98 | 0.12 | 0.97 | 1.00 |
| model_1161 | Scotoma | 2983 | 2983 | 0 | 2386 | 299 | 298 | 0.24 | 0.96 | 0.94 | 286 | 0 | 0 | 13 | 0.96 | 1.00 | 0.98 | 0.17 | 0.96 | 1.00 |
| model_1162 | Seasonal Affective Disorder | 51 | 588 | 537 | 470 | 59 | 59 | 0.27 | 0.66 | 0.56 | 39 | 0 | 0 | 20 | 0.66 | 1.00 | 0.8 | 1.36 | 0.66 | 1.00 |
| model_1163 | Seborrhoeic keratosis | 1282 | 1282 | 0 | 1025 | 129 | 128 | 0.23 | 0.93 | 0.96 | 120 | 0 | 0 | 9 | 0.93 | 1.00 | 0.96 | 0.28 | 0.93 | 1.00 |
| model_1164 | Sensory disturbance | 4227 | 4227 | 0 | 3381 | 423 | 423 | 0.25 | 0.96 | 0.95 | 406 | 0 | 0 | 17 | 0.96 | 1.00 | 0.98 | 0.16 | 0.96 | 1.00 |
| model_1165 | Sepsis | 16575 | 16575 | 0 | 13260 | 1658 | 1657 | 0.32 | 0.94 | 0.93 | 1554 | 0 | 0 | 104 | 0.94 | 1.00 | 0.97 | 0.25 | 0.94 | 1.00 |
| model_1166 | Septic abortion | 3 | 588 | 585 | 470 | 59 | 59 | 0.22 | 0 | 0 | 0 | 0 | 0 | 59 | 0 | 0.00 | 0 | 4 | 0 | 1.00 |
| model_1167 | Septic shock | 9254 | 9254 | 0 | 7403 | 926 | 925 | 0.29 | 0.95 | 0.94 | 882 | 0 | 0 | 44 | 0.95 | 1.00 | 0.98 | 0.19 | 0.95 | 1.00 |
| model_1168 | Seroma | 1602 | 1602 | 0 | 1281 | 161 | 160 | 0.24 | 0.95 | 0.93 | 153 | 0 | 0 | 8 | 0.95 | 1.00 | 0.97 | 0.2 | 0.95 | 1.00 |
| model_1169 | Serum sickness | 1110 | 1110 | 0 | 888 | 111 | 111 | 0.23 | 0.94 | 0.94 | 104 | 0 | 0 | 7 | 0.94 | 1.00 | 0.97 | 0.25 | 0.94 | 1.00 |
| model_1170 | Sexually transmitted diseases | 5 | 588 | 583 | 470 | 59 | 59 | 0.23 | 0 | 0 | 0 | 0 | 0 | 59 | 0 | 0.00 | 0 | 4 | 0 | 1.00 |
| model_1171 | Shoulder pain | 4026 | 4026 | 0 | 3220 | 403 | 403 | 0.25 | 0.95 | 0.97 | 382 | 0 | 0 | 21 | 0.95 | 1.00 | 0.97 | 0.21 | 0.95 | 1.00 |
| model_1172 | Sick sinus syndrome | 2694 | 2694 | 0 | 2155 | 270 | 269 | 0.25 | 0.93 | 0.96 | 250 | 0 | 0 | 20 | 0.93 | 1.00 | 0.96 | 0.3 | 0.93 | 1.00 |
| model_1173 | Sinus arrest | 681 | 681 | 0 | 544 | 69 | 68 | 0.23 | 0.94 | 0.94 | 65 | 0 | 0 | 4 | 0.94 | 1.00 | 0.97 | 0.23 | 0.94 | 1.00 |
| model_1174 | Sinus bradycardia | 5628 | 5628 | 0 | 4502 | 563 | 563 | 0.26 | 0.95 | 0.94 | 536 | 0 | 0 | 27 | 0.95 | 1.00 | 0.98 | 0.19 | 0.95 | 1.00 |
| model_1175 | Sinus headache | 1791 | 1791 | 0 | 1432 | 180 | 179 | 0.23 | 0.97 | 0.97 | 175 | 0 | 0 | 5 | 0.97 | 1.00 | 0.99 | 0.11 | 0.97 | 1.00 |
| model_1176 | Sinus tachycardia | 10348 | 10348 | 0 | 8278 | 1035 | 1035 | 0.29 | 0.94 | 0.96 | 977 | 0 | 0 | 58 | 0.94 | 1.00 | 0.97 | 0.22 | 0.94 | 1.00 |
| model_1177 | Sinusitis | 12955 | 12955 | 0 | 10364 | 1296 | 1295 | 0.29 | 0.96 | 0.97 | 1242 | 0 | 0 | 54 | 0.96 | 1.00 | 0.98 | 0.17 | 0.96 | 1.00 |
| model_1178 | Sjogrens syndrome | 1147 | 1147 | 0 | 917 | 115 | 115 | 0.23 | 0.93 | 0.95 | 107 | 0 | 0 | 8 | 0.93 | 1.00 | 0.96 | 0.28 | 0.93 | 1.00 |
| model_1179 | Skin Striae | 733 | 733 | 0 | 586 | 74 | 73 | 0.22 | 0.91 | 0.92 | 67 | 0 | 0 | 7 | 0.91 | 1.00 | 0.95 | 0.38 | 0.91 | 1.00 |
| model_1180 | Skin abrasion | 2 | 588 | 586 | 470 | 59 | 59 | 0.22 | 0 | 0 | 0 | 0 | 0 | 59 | 0 | 0.00 | 0 | 4 | 0 | 1.00 |
| model_1181 | Skin lesion | 6764 | 6764 | 0 | 5411 | 677 | 676 | 0.26 | 0.91 | 0.94 | 615 | 0 | 0 | 62 | 0.91 | 1.00 | 0.95 | 0.37 | 0.91 | 1.00 |
| model_1182 | Sleep apnea | 9190 | 9190 | 0 | 7352 | 919 | 919 | 0.28 | 0.95 | 0.96 | 874 | 0 | 0 | 45 | 0.95 | 1.00 | 0.97 | 0.2 | 0.95 | 1.00 |
| model_1183 | Sleep disorder | 9102 | 9102 | 0 | 7281 | 911 | 910 | 0.28 | 0.97 | 0.96 | 880 | 0 | 0 | 31 | 0.97 | 1.00 | 0.98 | 0.14 | 0.97 | 1.00 |
| model_1184 | Sleep walking | 3348 | 3348 | 0 | 2678 | 335 | 335 | 0.25 | 0.96 | 0.97 | 323 | 0 | 0 | 12 | 0.96 | 1.00 | 0.98 | 0.14 | 0.96 | 1.00 |
| model_1185 | Small intestinal obstruction | 4839 | 4839 | 0 | 3871 | 484 | 484 | 0.25 | 0.95 | 0.94 | 462 | 0 | 0 | 22 | 0.95 | 1.00 | 0.98 | 0.18 | 0.95 | 1.00 |
| model_1186 | Sneezing | 3043 | 3043 | 0 | 2434 | 305 | 304 | 0.25 | 0.95 | 0.96 | 291 | 0 | 0 | 14 | 0.95 | 1.00 | 0.98 | 0.18 | 0.95 | 1.00 |
| model_1187 | Soft tissue infection | 991 | 991 | 0 | 792 | 100 | 99 | 0.22 | 0.89 | 0.97 | 89 | 0 | 0 | 11 | 0.89 | 1.00 | 0.94 | 0.44 | 0.89 | 1.00 |
| model_1188 | Soft tissue injuries | 649 | 649 | 0 | 519 | 65 | 65 | 0.23 | 0.97 | 0.95 | 63 | 0 | 0 | 2 | 0.97 | 1.00 | 0.98 | 0.12 | 0.97 | 1.00 |
| model_1189 | Spider angioma | 362 | 588 | 226 | 470 | 59 | 59 | 0.22 | 0.93 | 0.95 | 55 | 0 | 0 | 4 | 0.93 | 1.00 | 0.96 | 0.27 | 0.93 | 1.00 |
| model_1190 | Spinal Compression Fracture | 3677 | 3677 | 0 | 2941 | 368 | 368 | 0.26 | 0.95 | 0.95 | 349 | 0 | 0 | 19 | 0.95 | 1.00 | 0.97 | 0.21 | 0.95 | 1.00 |
| model_1191 | Spinal cord injury | 354 | 588 | 234 | 470 | 59 | 59 | 0.22 | 0.95 | 0.95 | 56 | 0 | 0 | 3 | 0.95 | 1.00 | 0.97 | 0.2 | 0.95 | 1.00 |
| model_1192 | Spinal fracture | 3131 | 3131 | 0 | 2504 | 314 | 313 | 0.25 | 0.95 | 0.96 | 299 | 0 | 0 | 15 | 0.95 | 1.00 | 0.98 | 0.19 | 0.95 | 1.00 |
| model_1193 | Spinning sensation | 6835 | 6835 | 0 | 5468 | 684 | 683 | 0.28 | 0.95 | 0.97 | 650 | 0 | 0 | 34 | 0.95 | 1.00 | 0.97 | 0.2 | 0.95 | 1.00 |

| | | | | | | | | | | | | | | | | | | | | |
|---|---|---|---|---|---|---|---|---|---|---|---|---|---|---|---|---|---|---|---|---|
| model_1194 | Splenectomy | 530 | 588 | 58 | 470 | 59 | 59 | 0.22 | 0.92 | 0.86 | 54 | 0 | 0 | 5 | 0.92 | 1.00 | 0.96 | 0.34 | 0.92 | 1.00 |
| model_1195 | Splenic infarction | 1300 | 1300 | 0 | 1040 | 130 | 130 | 0.23 | 0.93 | 0.89 | 121 | 0 | 0 | 9 | 0.93 | 1.00 | 0.96 | 0.28 | 0.93 | 1.00 |
| model_1196 | Splinter | 3 | 588 | 585 | 470 | 59 | 59 | 0.21 | 0 | 0 | 0 | 0 | 0 | 59 | 0 | 0.00 | 0 | 4 | 0 | 1.00 |
| model_1197 | Spondylitis | 765 | 765 | 0 | 612 | 77 | 76 | 0.23 | 0.97 | 0.97 | 75 | 0 | 0 | 2 | 0.97 | 1.00 | 0.99 | 0.1 | 0.97 | 1.00 |
| model_1198 | Spondylosis | 913 | 913 | 0 | 730 | 92 | 91 | 0.22 | 0.95 | 0.97 | 87 | 0 | 0 | 5 | 0.95 | 1.00 | 0.97 | 0.22 | 0.95 | 1.00 |
| model_1199 | Squamous cell carcinoma of skin | 1042 | 1042 | 0 | 833 | 105 | 104 | 0.23 | 0.94 | 0.95 | 99 | 0 | 0 | 6 | 0.94 | 1.00 | 0.97 | 0.23 | 0.94 | 1.00 |
| model_1200 | Status epilepticus | 1904 | 1904 | 0 | 1523 | 191 | 190 | 0.23 | 0.92 | 0.91 | 176 | 0 | 0 | 15 | 0.92 | 1.00 | 0.96 | 0.31 | 0.92 | 1.00 |
| model_1201 | Still birth | 11952 | 11952 | 0 | 9561 | 1196 | 1195 | 0.29 | 0.95 | 0.95 | 1134 | 0 | 0 | 62 | 0.95 | 1.00 | 0.97 | 0.21 | 0.95 | 1.00 |
| model_1202 | Strabismus | 1995 | 1995 | 0 | 1596 | 200 | 199 | 0.25 | 0.91 | 0.92 | 182 | 0 | 0 | 18 | 0.91 | 1.00 | 0.95 | 0.36 | 0.91 | 1.00 |
| model_1203 | Streptococcal infection | 3881 | 3881 | 0 | 3104 | 389 | 388 | 0.24 | 0.91 | 0.9 | 353 | 0 | 0 | 36 | 0.91 | 1.00 | 0.95 | 0.37 | 0.91 | 1.00 |
| model_1204 | Stress incontinence | 2619 | 2619 | 0 | 2095 | 262 | 262 | 0.24 | 0.95 | 0.97 | 249 | 0 | 0 | 13 | 0.95 | 1.00 | 0.97 | 0.2 | 0.95 | 1.00 |
| model_1205 | Stridor | 1552 | 1552 | 0 | 1241 | 156 | 155 | 0.22 | 0.93 | 0.97 | 145 | 0 | 0 | 11 | 0.93 | 1.00 | 0.96 | 0.28 | 0.93 | 1.00 |
| model_1206 | Subarachnoid haemorrhage | 2960 | 2960 | 0 | 2368 | 296 | 296 | 0.25 | 0.93 | 0.97 | 274 | 0 | 0 | 22 | 0.93 | 1.00 | 0.96 | 0.3 | 0.93 | 1.00 |
| model_1207 | Substance abuse | 644 | 644 | 0 | 515 | 65 | 64 | 0.22 | 0.98 | 0.94 | 64 | 0 | 0 | 1 | 0.98 | 1.00 | 0.99 | 0.06 | 0.98 | 1.00 |
| model_1208 | Sunburn | 3420 | 3420 | 0 | 2736 | 342 | 342 | 0.24 | 0.97 | 0.95 | 332 | 0 | 0 | 10 | 0.97 | 1.00 | 0.99 | 0.12 | 0.97 | 1.00 |
| model_1209 | Superficial thrombophlebitis | 1974 | 1974 | 0 | 1579 | 198 | 197 | 0.24 | 0.94 | 0.96 | 186 | 0 | 0 | 12 | 0.94 | 1.00 | 0.97 | 0.24 | 0.94 | 1.00 |
| model_1210 | Superior vena cava syndrome | 634 | 634 | 0 | 507 | 64 | 63 | 0.23 | 0.89 | 0.92 | 57 | 0 | 0 | 7 | 0.89 | 1.00 | 0.94 | 0.44 | 0.89 | 1.00 |
| model_1211 | Supernumerary nipple | 5 | 588 | 583 | 470 | 59 | 59 | 0.22 | 0 | 0 | 0 | 0 | 0 | 59 | 0 | 0.00 | 0 | 4 | 0 | 1.00 |
| model_1212 | Supraventricular tachycardia | 5905 | 5905 | 0 | 4724 | 591 | 590 | 0.26 | 0.94 | 0.96 | 555 | 0 | 0 | 36 | 0.94 | 1.00 | 0.97 | 0.24 | 0.94 | 1.00 |
| model_1213 | Swollen scrotum | 568 | 588 | 20 | 470 | 59 | 59 | 0.22 | 0.92 | 0.92 | 54 | 0 | 0 | 5 | 0.92 | 1.00 | 0.96 | 0.34 | 0.92 | 1.00 |
| model_1214 | Syncope vasovagal | 3147 | 3147 | 0 | 2517 | 315 | 315 | 0.25 | 0.97 | 0.95 | 305 | 0 | 0 | 10 | 0.97 | 1.00 | 0.98 | 0.13 | 0.97 | 1.00 |
| model_1215 | Synovial cyst | 1929 | 1929 | 0 | 1543 | 193 | 193 | 0.24 | 0.96 | 0.97 | 186 | 0 | 0 | 7 | 0.96 | 1.00 | 0.98 | 0.15 | 0.96 | 1.00 |
| model_1216 | Synovitis | 1838 | 1838 | 0 | 1470 | 184 | 184 | 0.25 | 0.97 | 0.97 | 179 | 0 | 0 | 5 | 0.97 | 1.00 | 0.99 | 0.11 | 0.97 | 1.00 |
| model_1217 | Syphilis | 105 | 588 | 483 | 470 | 59 | 59 | 0.25 | 0.75 | 0.71 | 44 | 0 | 0 | 15 | 0.75 | 1.00 | 0.85 | 1.02 | 0.75 | 1.00 |
| model_1218 | Tachycardia ventricular | 7098 | 7098 | 0 | 5678 | 710 | 710 | 0.26 | 0.91 | 0.94 | 646 | 0 | 0 | 64 | 0.91 | 1.00 | 0.95 | 0.36 | 0.91 | 1.00 |
| model_1219 | Tardive dyskinesia | 1997 | 1997 | 0 | 1597 | 200 | 200 | 0.23 | 0.99 | 0.96 | 198 | 0 | 0 | 2 | 0.99 | 1.00 | 0.99 | 0.04 | 0.99 | 1.00 |
| model_1220 | Telangiectases | 583 | 588 | 5 | 470 | 59 | 59 | 0.23 | 0.98 | 0.97 | 58 | 0 | 0 | 1 | 0.98 | 1.00 | 0.99 | 0.07 | 0.98 | 1.00 |
| model_1221 | Temporal arteritis | 654 | 654 | 0 | 523 | 66 | 65 | 0.22 | 0.92 | 0.88 | 61 | 0 | 0 | 5 | 0.92 | 1.00 | 0.96 | 0.3 | 0.92 | 1.00 |
| model_1222 | Tendinitis | 4595 | 4595 | 0 | 3676 | 460 | 459 | 0.25 | 0.96 | 0.95 | 442 | 0 | 0 | 18 | 0.96 | 1.00 | 0.98 | 0.16 | 0.96 | 1.00 |
| model_1223 | Tendon injury | 2491 | 2491 | 0 | 1992 | 250 | 249 | 0.23 | 0.95 | 0.93 | 237 | 0 | 0 | 13 | 0.95 | 1.00 | 0.97 | 0.21 | 0.95 | 1.00 |
| model_1224 | Tenosynovitis | 1331 | 1331 | 0 | 1064 | 134 | 133 | 0.23 | 0.94 | 0.94 | 126 | 0 | 0 | 8 | 0.94 | 1.00 | 0.97 | 0.24 | 0.94 | 1.00 |
| model_1225 | Tension headache | 1252 | 1252 | 0 | 1001 | 126 | 125 | 0.24 | 0.96 | 0.99 | 121 | 0 | 0 | 5 | 0.96 | 1.00 | 0.98 | 0.16 | 0.96 | 1.00 |
| model_1226 | Testes pain | 930 | 930 | 0 | 744 | 93 | 93 | 0.24 | 0.94 | 0.92 | 87 | 0 | 0 | 6 | 0.94 | 1.00 | 0.97 | 0.26 | 0.94 | 1.00 |
| model_1227 | Testicular atrophy | 102 | 588 | 486 | 470 | 59 | 59 | 0.25 | 0.86 | 0.76 | 51 | 0 | 0 | 8 | 0.86 | 1.00 | 0.93 | 0.54 | 0.86 | 1.00 |
| model_1228 | Testicular swelling | 714 | 714 | 0 | 571 | 72 | 71 | 0.24 | 0.97 | 0.99 | 70 | 0 | 0 | 2 | 0.97 | 1.00 | 0.99 | 0.11 | 0.97 | 1.00 |
| model_1229 | Tetanus | 16 | 588 | 572 | 470 | 59 | 59 | 0.25 | 0.17 | 0.17 | 10 | 0 | 0 | 49 | 0.17 | 1.00 | 0.29 | 3.32 | 0.17 | 1.00 |
| model_1230 | Tetany | 377 | 588 | 211 | 470 | 59 | 59 | 0.22 | 0.93 | 0.97 | 55 | 0 | 0 | 4 | 0.93 | 1.00 | 0.96 | 0.27 | 0.93 | 1.00 |
| model_1231 | Tetralogy of Fallot | 260 | 588 | 328 | 470 | 59 | 59 | 0.22 | 0.86 | 0.92 | 51 | 0 | 0 | 8 | 0.86 | 1.00 | 0.93 | 0.54 | 0.86 | 1.00 |
| model_1232 | Thrombocytopenia | 17060 | 17060 | 0 | 13648 | 1706 | 1706 | 0.33 | 0.95 | 0.93 | 1619 | 0 | 0 | 87 | 0.95 | 1.00 | 0.97 | 0.2 | 0.95 | 1.00 |
| model_1233 | Thrombophlebitis | 1865 | 1865 | 0 | 1492 | 187 | 186 | 0.23 | 0.96 | 0.85 | 179 | 0 | 0 | 8 | 0.96 | 1.00 | 0.98 | 0.17 | 0.96 | 1.00 |
| model_1234 | Thrombotic microangiopathy | 2716 | 2716 | 0 | 2172 | 272 | 272 | 0.25 | 0.94 | 0.89 | 255 | 0 | 0 | 17 | 0.94 | 1.00 | 0.97 | 0.25 | 0.94 | 1.00 |
| model_1235 | Thyroid adenoma | 77 | 588 | 511 | 470 | 59 | 59 | 0.27 | 0.68 | 0.64 | 40 | 0 | 0 | 19 | 0.68 | 1.00 | 0.81 | 1.29 | 0.68 | 1.00 |
| model_1236 | Thyroid cancer | 404 | 588 | 184 | 470 | 59 | 59 | 0.24 | 0.97 | 0.92 | 57 | 0 | 0 | 2 | 0.97 | 1.00 | 0.98 | 0.14 | 0.97 | 1.00 |
| model_1237 | Thyroid cyst | 323 | 588 | 265 | 470 | 59 | 59 | 0.24 | 0.88 | 0.95 | 52 | 0 | 0 | 7 | 0.88 | 1.00 | 0.94 | 0.47 | 0.88 | 1.00 |
| model_1238 | Thyroid disease | 3850 | 3850 | 0 | 3080 | 385 | 385 | 0.26 | 0.96 | 0.98 | 371 | 0 | 0 | 14 | 0.96 | 1.00 | 0.98 | 0.15 | 0.96 | 1.00 |
| model_1239 | Thyroid neoplasia | 1947 | 1947 | 0 | 1557 | 195 | 195 | 0.24 | 0.94 | 0.97 | 184 | 0 | 0 | 11 | 0.94 | 1.00 | 0.97 | 0.23 | 0.94 | 1.00 |
| model_1240 | Thyroidectomy | 454 | 588 | 134 | 470 | 59 | 59 | 0.24 | 0.93 | 0.92 | 55 | 0 | 0 | 4 | 0.93 | 1.00 | 0.96 | 0.27 | 0.93 | 1.00 |
| model_1241 | Thyroiditis | 424 | 588 | 164 | 470 | 59 | 59 | 0.24 | 0.83 | 0.9 | 49 | 0 | 0 | 10 | 0.83 | 1.00 | 0.91 | 0.68 | 0.83 | 1.00 |
| model_1242 | Tinea | 320 | 588 | 268 | 470 | 59 | 59 | 0.24 | 0.93 | 0.95 | 55 | 0 | 0 | 4 | 0.93 | 1.00 | 0.96 | 0.27 | 0.93 | 1.00 |
| model_1243 | Tinea Capitis | 57 | 588 | 531 | 470 | 59 | 59 | 0.28 | 0.41 | 0.61 | 24 | 0 | 0 | 35 | 0.41 | 1.00 | 0.58 | 2.37 | 0.41 | 1.00 |

| | | | | | | | | | | | | | | | | | | | | |
|---|---|---|---|---|---|---|---|---|---|---|---|---|---|---|---|---|---|---|---|---|
| model_1244 | Tinea cruris | 361 | 588 | 227 | 470 | 59 | 59 | 0.23 | 0.88 | 0.93 | 52 | 0 | 0 | 7 | 0.88 | 1.00 | 0.94 | 0.47 | 0.88 | 1.00 |
| model_1245 | Tinea pedis | 769 | 769 | 0 | 615 | 77 | 77 | 0.23 | 0.99 | 0.95 | 76 | 0 | 0 | 1 | 0.99 | 1.00 | 0.99 | 0.05 | 0.99 | 1.00 |
| model_1246 | Tinea versicolor | 237 | 588 | 351 | 470 | 59 | 59 | 0.24 | 0.85 | 0.93 | 50 | 0 | 0 | 9 | 0.85 | 1.00 | 0.92 | 0.61 | 0.85 | 1.00 |
| model_1247 | Tinnitus | 6437 | 6437 | 0 | 5149 | 644 | 644 | 0.27 | 0.97 | 0.98 | 622 | 0 | 0 | 22 | 0.97 | 1.00 | 0.98 | 0.14 | 0.97 | 1.00 |
| model_1248 | Tonsillar hypertrophy | 148 | 588 | 440 | 470 | 59 | 59 | 0.26 | 0.8 | 0.8 | 47 | 0 | 0 | 12 | 0.8 | 1.00 | 0.89 | 0.81 | 0.8 | 1.00 |
| model_1249 | Tonsillectomy | 272 | 588 | 316 | 470 | 59 | 59 | 0.25 | 0.92 | 0.92 | 54 | 0 | 0 | 5 | 0.92 | 1.00 | 0.96 | 0.34 | 0.92 | 1.00 |
| model_1250 | Tonsillitis | 783 | 783 | 0 | 626 | 79 | 78 | 0.24 | 0.86 | 0.95 | 68 | 0 | 0 | 11 | 0.86 | 1.00 | 0.93 | 0.56 | 0.86 | 1.00 |
| model_1251 | Tooth Impacted | 676 | 676 | 0 | 540 | 68 | 68 | 0.25 | 0.94 | 0.93 | 64 | 0 | 0 | 4 | 0.94 | 1.00 | 0.97 | 0.24 | 0.94 | 1.00 |
| model_1252 | Tooth disease | 5573 | 5573 | 0 | 4458 | 558 | 557 | 0.27 | 0.97 | 0.95 | 540 | 0 | 0 | 18 | 0.97 | 1.00 | 0.98 | 0.13 | 0.97 | 1.00 |
| model_1253 | Toxic shock | 751 | 751 | 0 | 600 | 76 | 75 | 0.24 | 0.88 | 0.87 | 67 | 0 | 0 | 9 | 0.88 | 1.00 | 0.94 | 0.47 | 0.88 | 1.00 |
| model_1254 | Toxoplasmosis | 248 | 588 | 340 | 470 | 59 | 59 | 0.22 | 0.92 | 0.9 | 54 | 0 | 0 | 5 | 0.92 | 1.00 | 0.96 | 0.34 | 0.92 | 1.00 |
| model_1255 | Tracheitis | 853 | 853 | 0 | 682 | 86 | 85 | 0.22 | 0.93 | 0.99 | 80 | 0 | 0 | 6 | 0.93 | 1.00 | 0.96 | 0.28 | 0.93 | 1.00 |
| model_1256 | Tracheobronchitis | 882 | 882 | 0 | 705 | 89 | 88 | 0.23 | 0.96 | 0.94 | 85 | 0 | 0 | 4 | 0.96 | 1.00 | 0.98 | 0.18 | 0.96 | 1.00 |
| model_1257 | Tracheostomy | 1020 | 1020 | 0 | 816 | 102 | 102 | 0.22 | 0.85 | 0.91 | 87 | 0 | 0 | 15 | 0.85 | 1.00 | 0.92 | 0.59 | 0.85 | 1.00 |
| model_1258 | Transfusion reaction | 1176 | 1176 | 0 | 940 | 118 | 118 | 0.23 | 0.89 | 0.84 | 105 | 0 | 0 | 13 | 0.89 | 1.00 | 0.94 | 0.44 | 0.89 | 1.00 |
| model_1259 | Transient ischaemic attack | 9538 | 9538 | 0 | 7630 | 954 | 954 | 0.29 | 0.96 | 0.95 | 916 | 0 | 0 | 38 | 0.96 | 1.00 | 0.98 | 0.16 | 0.96 | 1.00 |
| model_1260 | Transurethral resection of the prostate | 293 | 588 | 295 | 470 | 59 | 59 | 0.24 | 0.86 | 0.9 | 51 | 0 | 0 | 8 | 0.86 | 1.00 | 0.93 | 0.54 | 0.86 | 1.00 |
| model_1261 | Traumatic amputation | 12 | 588 | 576 | 470 | 59 | 59 | 0.25 | 0 | 0 | 0 | 0 | 0 | 59 | 0 | 0.00 | 0 | 4 | 0 | 1.00 |
| model_1262 | Traumatic arthropathy | 117 | 588 | 471 | 470 | 59 | 59 | 0.26 | 0.9 | 0.86 | 53 | 0 | 0 | 6 | 0.9 | 1.00 | 0.95 | 0.41 | 0.9 | 1.00 |
| model_1263 | Tremor | 14419 | 14419 | 0 | 11535 | 1442 | 1442 | 0.32 | 0.95 | 0.95 | 1364 | 0 | 0 | 78 | 0.95 | 1.00 | 0.97 | 0.22 | 0.95 | 1.00 |
| model_1264 | Trichomoniasis | 154 | 588 | 434 | 470 | 59 | 59 | 0.26 | 0.85 | 0.93 | 50 | 0 | 0 | 9 | 0.85 | 1.00 | 0.92 | 0.61 | 0.85 | 1.00 |
| model_1265 | Trifascicular block | 26 | 588 | 562 | 470 | 59 | 59 | 0.25 | 0.29 | 0.31 | 17 | 0 | 0 | 42 | 0.29 | 1.00 | 0.45 | 2.85 | 0.29 | 1.00 |
| model_1266 | Trigeminal neuralgia | 3308 | 3308 | 0 | 2646 | 331 | 331 | 0.26 | 0.96 | 0.96 | 319 | 0 | 0 | 12 | 0.96 | 1.00 | 0.98 | 0.15 | 0.96 | 1.00 |
| model_1267 | Trigger finger | 1085 | 1085 | 0 | 868 | 109 | 108 | 0.23 | 0.95 | 0.99 | 104 | 0 | 0 | 5 | 0.95 | 1.00 | 0.98 | 0.18 | 0.95 | 1.00 |
| model_1268 | Trisomy 21 | 174 | 588 | 414 | 470 | 59 | 59 | 0.23 | 0.88 | 0.92 | 52 | 0 | 0 | 7 | 0.88 | 1.00 | 0.94 | 0.47 | 0.88 | 1.00 |
| model_1269 | Tuberculin test positive | 267 | 588 | 321 | 470 | 59 | 59 | 0.23 | 0.92 | 0.76 | 54 | 0 | 0 | 5 | 0.92 | 1.00 | 0.96 | 0.34 | 0.92 | 1.00 |
| model_1270 | Tympanic membrane disorder | 149 | 588 | 439 | 470 | 59 | 59 | 0.24 | 0.88 | 0.88 | 52 | 0 | 0 | 7 | 0.88 | 1.00 | 0.94 | 0.47 | 0.88 | 1.00 |
| model_1271 | Tympanic membrane perforation | 746 | 746 | 0 | 596 | 75 | 75 | 0.24 | 0.97 | 0.96 | 73 | 0 | 0 | 2 | 0.97 | 1.00 | 0.99 | 0.11 | 0.97 | 1.00 |
| model_1272 | Typhoid | 10 | 588 | 578 | 470 | 59 | 59 | 0.23 | 0 | 0 | 0 | 0 | 0 | 59 | 0 | 0.00 | 0 | 4 | 0 | 1.00 |
| model_1273 | Ulcer | 5661 | 5661 | 0 | 4528 | 567 | 566 | 0.27 | 0.96 | 0.93 | 542 | 0 | 0 | 25 | 0.96 | 1.00 | 0.98 | 0.18 | 0.96 | 1.00 |
| model_1274 | Umbilical hernia | 1842 | 1842 | 0 | 1473 | 185 | 184 | 0.24 | 0.95 | 0.93 | 175 | 0 | 0 | 10 | 0.95 | 1.00 | 0.97 | 0.22 | 0.95 | 1.00 |
| model_1275 | Upper gastrointestinal bleeding | 4502 | 4502 | 0 | 3601 | 451 | 450 | 0.26 | 0.92 | 0.92 | 413 | 0 | 0 | 38 | 0.92 | 1.00 | 0.96 | 0.34 | 0.92 | 1.00 |
| model_1276 | Ureteral obstruction | 2553 | 2553 | 0 | 2042 | 256 | 255 | 0.24 | 0.95 | 0.93 | 243 | 0 | 0 | 13 | 0.95 | 1.00 | 0.97 | 0.2 | 0.95 | 1.00 |
| model_1277 | Urethritis | 363 | 588 | 225 | 470 | 59 | 59 | 0.25 | 0.95 | 0.9 | 56 | 0 | 0 | 3 | 0.95 | 1.00 | 0.97 | 0.2 | 0.95 | 1.00 |
| model_1278 | Urogenital abnormalities | 32 | 588 | 556 | 470 | 59 | 59 | 0.25 | 0.34 | 0.53 | 20 | 0 | 0 | 39 | 0.34 | 1.00 | 0.51 | 2.64 | 0.34 | 1.00 |
| model_1279 | Urosepsis | 3932 | 3932 | 0 | 3145 | 394 | 393 | 0.25 | 0.96 | 0.96 | 380 | 0 | 0 | 14 | 0.96 | 1.00 | 0.98 | 0.14 | 0.96 | 1.00 |
| model_1280 | Uterine bleeding | 935 | 935 | 0 | 748 | 94 | 93 | 0.23 | 0.96 | 0.96 | 90 | 0 | 0 | 4 | 0.96 | 1.00 | 0.98 | 0.17 | 0.96 | 1.00 |
| model_1281 | Uterine infection | 35 | 588 | 553 | 470 | 59 | 59 | 0.27 | 0.51 | 0.47 | 30 | 0 | 0 | 29 | 0.51 | 1.00 | 0.67 | 1.97 | 0.51 | 1.00 |
| model_1282 | Uterine polyp | 788 | 788 | 0 | 630 | 79 | 79 | 0.24 | 0.92 | 0.96 | 73 | 0 | 0 | 6 | 0.92 | 1.00 | 0.96 | 0.3 | 0.92 | 1.00 |
| model_1283 | Vaginal discharge | 1553 | 1553 | 0 | 1242 | 156 | 155 | 0.24 | 0.95 | 0.97 | 148 | 0 | 0 | 8 | 0.95 | 1.00 | 0.97 | 0.21 | 0.95 | 1.00 |
| model_1284 | Vaginal dysplasia | 107 | 588 | 481 | 470 | 59 | 59 | 0.27 | 0.53 | 0.59 | 31 | 0 | 0 | 28 | 0.53 | 1.00 | 0.69 | 1.9 | 0.53 | 1.00 |
| model_1285 | Vaginal prolapse | 205 | 588 | 383 | 470 | 59 | 59 | 0.26 | 0.81 | 0.85 | 48 | 0 | 0 | 11 | 0.81 | 1.00 | 0.9 | 0.75 | 0.81 | 1.00 |
| model_1286 | Varicocele | 89 | 588 | 499 | 470 | 59 | 59 | 0.25 | 0.68 | 0.64 | 40 | 0 | 0 | 19 | 0.68 | 1.00 | 0.81 | 1.29 | 0.68 | 1.00 |
| model_1287 | Varicose vein | 1905 | 1905 | 0 | 1524 | 191 | 190 | 0.24 | 0.95 | 0.95 | 181 | 0 | 0 | 10 | 0.95 | 1.00 | 0.97 | 0.21 | 0.95 | 1.00 |
| model_1288 | Vascular anomaly | 126 | 588 | 462 | 470 | 59 | 59 | 0.24 | 0.88 | 0.73 | 52 | 0 | 0 | 7 | 0.88 | 1.00 | 0.94 | 0.47 | 0.88 | 1.00 |
| model_1289 | Vascular headache | 37 | 588 | 551 | 470 | 59 | 59 | 0.26 | 0.24 | 0.31 | 14 | 0 | 0 | 45 | 0.24 | 1.00 | 0.38 | 3.05 | 0.24 | 1.00 |
| model_1290 | Vasomotor Rhinitis | 28 | 588 | 560 | 470 | 59 | 59 | 0.25 | 0.17 | 0.25 | 10 | 0 | 0 | 49 | 0.17 | 1.00 | 0.29 | 3.32 | 0.17 | 1.00 |
| model_1291 | Venous insufficiency | 1962 | 1962 | 0 | 1569 | 197 | 196 | 0.24 | 0.97 | 0.89 | 191 | 0 | 0 | 6 | 0.97 | 1.00 | 0.98 | 0.12 | 0.97 | 1.00 |
| model_1292 | Ventricular fibrillation | 5327 | 5327 | 0 | 4261 | 533 | 533 | 0.24 | 0.95 | 0.93 | 507 | 0 | 0 | 26 | 0.95 | 1.00 | 0.98 | 0.2 | 0.95 | 1.00 |
| model_1293 | Ventricular septal defect | 842 | 842 | 0 | 673 | 85 | 84 | 0.22 | 0.94 | 0.94 | 80 | 0 | 0 | 5 | 0.94 | 1.00 | 0.97 | 0.24 | 0.94 | 1.00 |

| | | | | | | | | | | | | | | | | | | | | |
|---|---|---|---|---|---|---|---|---|---|---|---|---|---|---|---|---|---|---|---|---|
| model_1294 | Vestibular disorder | 1489 | 1489 | 0 | 1191 | 149 | 149 | 0.24 | 0.96 | 0.97 | 143 | 0 | 0 | 6 | 0.96 | 1.00 | 0.98 | 0.16 | 0.96 | 1.00 |
| model_1295 | Viral Pharyngitis | 898 | 898 | 0 | 718 | 90 | 90 | 0.23 | 0.98 | 0.94 | 88 | 0 | 0 | 2 | 0.98 | 1.00 | 0.99 | 0.09 | 0.98 | 1.00 |
| model_1296 | Viral pneumonia | 853 | 853 | 0 | 682 | 86 | 85 | 0.24 | 0.94 | 0.98 | 81 | 0 | 0 | 5 | 0.94 | 1.00 | 0.97 | 0.23 | 0.94 | 1.00 |
| model_1297 | Viral rash NOS | 259 | 588 | 329 | 470 | 59 | 59 | 0.24 | 0.85 | 0.85 | 50 | 0 | 0 | 9 | 0.85 | 1.00 | 0.92 | 0.61 | 0.85 | 1.00 |
| model_1298 | Vitamin B 12 deficiency | 1618 | 1618 | 0 | 1294 | 162 | 162 | 0.25 | 0.96 | 0.92 | 155 | 0 | 0 | 7 | 0.96 | 1.00 | 0.98 | 0.17 | 0.96 | 1.00 |
| model_1299 | Vitamin B deficiency | 128 | 588 | 460 | 470 | 59 | 59 | 0.26 | 0.8 | 0.61 | 47 | 0 | 0 | 12 | 0.8 | 1.00 | 0.89 | 0.81 | 0.8 | 1.00 |
| model_1300 | Vitamin D Deficiency | 879 | 879 | 0 | 703 | 88 | 88 | 0.23 | 0.95 | 0.92 | 84 | 0 | 0 | 4 | 0.95 | 1.00 | 0.98 | 0.18 | 0.95 | 1.00 |
| model_1301 | Vitiligo | 170 | 588 | 418 | 470 | 59 | 59 | 0.26 | 0.9 | 0.88 | 53 | 0 | 0 | 6 | 0.9 | 1.00 | 0.95 | 0.41 | 0.9 | 1.00 |
| model_1302 | Vitreous detachment | 685 | 685 | 0 | 548 | 69 | 68 | 0.22 | 0.99 | 0.99 | 68 | 0 | 0 | 1 | 0.99 | 1.00 | 0.99 | 0.06 | 0.99 | 1.00 |
| model_1303 | Vitreous haemorrhage | 1251 | 1251 | 0 | 1000 | 126 | 125 | 0.24 | 0.94 | 0.94 | 118 | 0 | 0 | 8 | 0.94 | 1.00 | 0.97 | 0.25 | 0.94 | 1.00 |
| model_1304 | Vocal cord paralysis | 963 | 963 | 0 | 770 | 97 | 96 | 0.23 | 0.95 | 0.96 | 92 | 0 | 0 | 5 | 0.95 | 1.00 | 0.97 | 0.21 | 0.95 | 1.00 |
| model_1305 | Volvulus | 657 | 657 | 0 | 525 | 66 | 66 | 0.23 | 1 | 0.98 | 66 | 0 | 0 | 0 | 1 | 1.00 | 1 | 0 | 1 | 1.00 |
| model_1306 | Vulval abscess | 137 | 588 | 451 | 470 | 59 | 59 | 0.25 | 0.83 | 0.75 | 49 | 0 | 0 | 10 | 0.83 | 1.00 | 0.91 | 0.68 | 0.83 | 1.00 |
| model_1307 | Vulvitis | 35 | 588 | 553 | 470 | 59 | 59 | 0.25 | 0.51 | 0.44 | 30 | 0 | 0 | 29 | 0.51 | 1.00 | 0.67 | 1.97 | 0.51 | 1.00 |
| model_1308 | Vulvovaginitis | 179 | 588 | 409 | 470 | 59 | 59 | 0.23 | 0.86 | 0.93 | 51 | 0 | 0 | 8 | 0.86 | 1.00 | 0.93 | 0.54 | 0.86 | 1.00 |
| model_1309 | Wandering atrial pacemaker | 134 | 588 | 454 | 470 | 59 | 59 | 0.25 | 0.78 | 0.81 | 46 | 0 | 0 | 13 | 0.78 | 1.00 | 0.88 | 0.88 | 0.78 | 1.00 |
| model_1310 | Weight gain | 17112 | 17112 | 0 | 13689 | 1712 | 1711 | 0.32 | 0.96 | 0.96 | 1643 | 0 | 0 | 69 | 0.96 | 1.00 | 0.98 | 0.16 | 0.96 | 1.00 |
| model_1311 | Wheeze | 8441 | 8441 | 0 | 6752 | 845 | 844 | 0.28 | 0.96 | 0.95 | 815 | 0 | 0 | 30 | 0.96 | 1.00 | 0.98 | 0.14 | 0.96 | 1.00 |
| model_1312 | Whiplash Injury | 483 | 588 | 105 | 470 | 59 | 59 | 0.23 | 0.98 | 0.97 | 58 | 0 | 0 | 1 | 0.98 | 1.00 | 0.99 | 0.07 | 0.98 | 1.00 |
| model_1313 | Wound dehiscence | 2993 | 2993 | 0 | 2394 | 300 | 299 | 0.23 | 0.93 | 0.89 | 278 | 0 | 0 | 22 | 0.93 | 1.00 | 0.96 | 0.29 | 0.93 | 1.00 |
| model_1314 | Wrist fracture | 2310 | 2310 | 0 | 1848 | 231 | 231 | 0.25 | 0.95 | 0.94 | 219 | 0 | 0 | 12 | 0.95 | 1.00 | 0.97 | 0.21 | 0.95 | 1.00 |
| model_1315 | Xanthoma | 67 | 588 | 521 | 470 | 59 | 59 | 0.26 | 0.66 | 0.73 | 39 | 0 | 0 | 20 | 0.66 | 1.00 | 0.8 | 1.36 | 0.66 | 1.00 |
| model_1316 | Xerosis | 857 | 857 | 0 | 685 | 86 | 86 | 0.23 | 0.88 | 0.93 | 76 | 0 | 0 | 10 | 0.88 | 1.00 | 0.94 | 0.47 | 0.88 | 1.00 |
